# Disentangling the shared genetics of ADHD, cannabis use disorder and cannabis use and prediction of cannabis use disorder in ADHD

**DOI:** 10.1101/2024.02.22.24303124

**Authors:** Trine Tollerup Nielsen, Jinjie Duan, Daniel F. Levey, G. Bragi Walters, Emma C. Johnson, Thorgeir Thorgeirsson, VA Million Veteran Program, Thomas Werge, Preben Bo Mortensen, Hreinn Stefansson, Kari Stefansson, David M. Hougaard, Arpana Agrawal, Joel Gelernter, Jakob Grove, Anders D. Børglum, Ditte Demontis

## Abstract

Cannabis use disorder (CUD) and cannabis use (CU) are prevalent conditions cooccurring with ADHD, but not much is known about the underlying shared genetics. Here we perform cross-disorder GWAS meta-analyses of ADHD and CUD or CU to identify pleiotropic risk loci and evaluate differences in the genetics of ADHD-CUD and ADHD-CU, and subsequently we dissect the polygenic architecture of CUD comorbidity in ADHD in the iPSYCH cohort.

There was a higher genetic overlap of ADHD and CUD than observed for ADHD and CU and we found a significant direct effect of ADHD genetic risk on CUD with only a minor part (12%) mediated by the genetics of CU. We identified 36 genome-wide significant loci for ADHD-CUD and 10 loci for ADHD-CU, with concordant direction of effect on the phenotypes. Three different approaches identified *DRD2*, which encodes the dopamine 2 receptor, as a risk gene for ADHD-CUD and, overall, ADHD-CUD risk genes were associated with high expression across several brain tissues and brain developmental stages, which was not observed for ADHD-CU genes. ADHD-CUD and ADHD-CU demonstrated similar genetic correlations with substance use phenotypes, while they differed significantly with respect to substance use disorder (SUD) phenotypes. ADHD-CUD individuals had significantly increased polygenic score (PGS) for psychiatric disorders compared to ADHD without CUD and increased burden of rare deleterious variants. Stratifying individuals with ADHD by their CUD-PGS revealed an absolute risk of 22% for comorbid CUD among the 20% of cases with the highest CUD-PGS, which was strikingly higher than the absolute risk of 1.6% observed among the 20% of controls with the highest CUD-PGS. Sex-specific analyses identified substantial differences in the absolute risk of comorbid CUD between males and females with ADHD, with a ∼10% higher CUD risk among males than females in the high-risk CUD-PGS group (24% risk for males and 14% risk for females).

## Introduction

Attention deficit hyperactivity disorder (ADHD) is a common neurodevelopmental psychiatric disorder with onset in childhood. It affects around 5% of children and often persist into adulthood where around 2.5% are affected^1^. The disorder is characterized by externalizing behaviours, such as age-inappropriate levels of impulsivity, hyperactivity, and inattention^1^. The genetic risk component is high with a heritability estimate of 0.74^2^, and we have previously demonstrated that a substantial amount of risk for ADHD can be explained by common genetic variation (14 - 22 %)^3,4^.

Individuals with ADHD have a 10-time increased risk of developing substance use disorders (SUD) compared to individuals without a prior diagnosis^5^, and the prevalence of SUD is around 23% among individuals with ADHD^6^. A comorbid SUD diagnosis has a strong negative impact on life-quality and is associated with a six- to sevenfold increased mortality rate in individuals with ADHD compared to the general population^7^, stressing that comorbid SUD is a severe, detrimental condition. One of the SUD conditions most commonly cooccurring with ADHD is cannabis use disorder (CUD)^8^, and it is particularly common among young people with ADHD; e.g., among people under 18 years of age seeking SUD treatment in the Danish health care system, 90% reported cannabis as their main drug in 2022^9^.

Externalizing and impulsive behaviors associated with ADHD might increase risk for initiating cannabis use^10,11^ and subsequent CUD. The risk could be promoted by a strong impact of the drug on individuals with ADHD, e.g. by affecting dopamine levels in the brain^12^, a neurotransmitter assumed to be dysregulated in individuals with ADHD^13^. Several lines of evidence suggest that genetic risk factors are shared across ADHD and CUD; in a large genome-wide association study (GWAS) meta-analysis of CUD we identified a significant genetic correlation (r_g_) of ADHD with both CUD (r_g_=0.62) and cannabis use (CU, r_g_=0.28)^14^, polygenic score (PGS) analyses have suggested genetics to be a contributor to ADHD-CUD comorbidity^15^, and structural equation modelling has found ADHD to group on a latent genetic factor representing shared genetic variance across several psychiatric disorders including CUD^16^.

Recent GWAS have found that the polygenic architecture of diagnosed SUD only partly overlap the polygenic architecture of substance use i.e., genetic correlation analyses of alcohol use disorder with alcohol use was 0.52^17^ and the genetic correlation of CUD with CU was 0.50^14^. ADHD has a genetic overlap with both CUD and CU (as described above), but it is not known to what extent the part of the genetic architecture of CUD and CU that overlaps with ADHD differ from one another. This study aims to characterize the shared genetics of ADHD with CUD and CU respectively and unravel their potential similarities and differences. This study also aims to characterize the polygenic risk component associated with ADHD comorbid with CUD and evaluate the ability of PGSs for predicting comorbid CUD in ADHD.

Here we perform cross-disorder GWAS of ADHD and CUD (referred to as ADHD-CUD) and ADHD and CU (referred to as ADHD-CU) to identify shared genetic risk loci and evaluate the extent to which the shared genetics of ADHD-CUD differ from the shared genetics of ADHD-CU. We use Gaussian mixture modelling to estimate the total number of common variants influencing ADHD and CUD and ADHD and CU. We investigate differences in the polygenic architecture of ADHD-CUD and ADHD-CU through genetic correlation analyses and intersecting with functional genomics data and assess ADHD-CUD and ADHD-CU risk gene expression across brain developmental stages. Subsequently, we focus specifically on ADHD-CUD comorbidity and use the large Danish iPSYCH^18,19^ case-cohort to: (I) explore the role of rare deleterious variants in ADHD-CUD, (II) dissect the polygenic architecture of ADHD-CUD in PGS analyses, and (III) to predict the absolute risk for comorbid CUD among individuals with ADHD stratified by their PGS.

## RESULTS

### Genetic overlap of ADHD with CUD and CU and mediation analyses

To estimate the extent to which common genetic variants are shared between ADHD, CUD and CU we used several approaches. First we performed genetic correlation analyses using GWAS summary statistics from meta-analysis of ADHD (38,691 ADHD; 186,843 controls)^3^, CUD (42,281 CUD; 843,744 controls)^20^ and CU (162,082 individuals)^21^, showing results consistent with previous reports (ADHD vs CUD; r_g_= 0.57, standard error [s.e.] = 0.04; ADHD vs CU; r_g_= 0.20, s.e. = 0.04; CUD vs CU; r_g_= 0.44, s.e. = 0.05; Supplementary Table 1). Then, we evaluated the extent to which the three phenotypes load on a shared genetic latent factor by modelling their genetic covariance structure in a common factor model using Genomic SEM^22^. All three phenotypes were significantly associated with the underlying shared genetic latent factor, with CUD having the highest loading (∼1, s.e. = 0.1), while similar loadings were observed for ADHD (0.57, s.e. = 0.05) and CU (0.42 s.e. = 0.04) (Supplementary Figure 1; Supplementary Table 2). We also used Genomic SEM to perform genetic mediation analyses and identified a significant direct effect of ADHD genetics on CUD (effect = 0.52, P=1.58x10^-26^) while only a minor part of the total effect of ADHD on CUD was mediated by CU genetics (12%) (Supplementary Table 2.b; Supplementary Figure 2).

To quantify the total number of shared common variants we used univariate Gaussian mixture modelling implemented in MiXeR^23^ to calculate (i) the number of variants influencing each phenotype (explaining 90% of the single nucleotide polymorphism (SNP) heritability (h^2^_SNP_)) and (ii) the number of shared variants. ADHD was influenced by ∼7,300 common variants as reported previously^3^, CUD by ∼8,300 variants and CU by ∼7,700 (Supplementary Table 3). A higher number of shared variants was observed for ADHD and CUD (∼7,000 variants) than for ADHD and CU (∼5,100 variants) and a larger proportion of the shared variants showed concordant directional effects with CUD (75%) than with CU (60%) (Supplementary Table 3).

Additionally, Mendelian randomization analyses identified a bidirectional significant causal relationship between ADHD and CUD (ADHD -> CUD Inverse Variance Weighted (IVW) beta = 0.31, s.e. = 0.06, P = 5.22×10^−7^; CUD -> ADHD IVW beta = 0.52, s.e. = 0.07, P = 2.90×10^−5^; Supplementary Table 4 and 5), which were supported by sensitivity analyses, especially GSMR with Heidi-outlier removal of pleiotropic variants, indicating that CUD has a larger causal effect on ADHD than the reverse.

### Cross-disorder GWAS meta-analyses of ADHD, CUD and CU

We performed two cross-disorder GWAS meta-analyses of (**I)** ADHD and CUD (referred to as ADHD-CUD) and **(II)** ADHD and CU (referred to as ADHD-CU) to identify shared risk variants across the two pairs of phenotypes, using the to-date largest GWAS meta-analyses of ADHD^3^, CUD^20^ and CU^21^ described above. We used ‘Association analysis based on SubSETs’ (ASSET)^24^, to identify variants with concordant and discordant allelic direction of effects on the phenotypes. We identified 36 genome-wide significant ADHD-CUD risk loci with concordant direction of effect (Supplementary Table 6, Supplementary Figure 3.a), and two loci with discordant direction of effect (Supplementary Table 7, Supplementary Figure 4.a). Two concordant loci on chromosomes 3 and 11 had independent secondary lead variants (*r*^2^ < 0.1 between the index variant and the secondary lead variant within a region of 0.5 Mb) that remained significant after conditional analysis (two secondary lead variants on chromosome 11 and one secondary independent lead variant on chromosome 3, Supplementary Table 8). All concordant index variants and independent secondary lead variants demonstrated a high posterior probability (m-value) for both phenotypes contributing to the observed signal (m > 0.85; Supplementary Table 6). The locus most strongly associated in ADHD-CUD was located on chromosome 11 (downstream *METTL15*, rs10835372; OR=1.06; P= 1.45×10^-18^) and is also genome-wide significantly associated with ADHD^3^ and CUD^20^ (Supplementary Table 6). The second most strongly associated locus was located on chromosome 7 (in *FOXP2;* rs1989903, OR=1.05, P = 6.67×10^-16^) which is also associated with both disorders separately^3,14^. We identified 17 shared loci that have not been reported for either disorder. Evaluating the concordant ADHD-CUD index variants in a Phenome-wide association study (PheWAS) revealed that the variants are especially associated with phenotypes within domains related to smoking, weight and cardiometabolic diseases (Supplementary Table 11), which are domains that are also linked to the single disorders separately^3,14^.

We identified 13 genome-wide significant loci associated with ADHD-CU (Supplementary Table 9; Supplementary Figure 3.b.) with concordant direction of effect, 11 of which demonstrated a high contribution from both phenotypes (m-value > 0.85; Supplementary Table 9). Ten loci were genome-wide significantly associated with discordant direction of effects (Supplementary Table 10; Supplementary Figure 4.b).

Five loci were identified in both ADHD-CUD and ADHD-CU GWASs with concordant direction of effects.

Regional association plots for concordant loci can be found in Extended Data 1 and forest plots and m-value plots in Extended Data 2.

### Tissue specific expression of ADHD-CUD and ADHD-CU associated genes

We evaluated the expression of ADHD-CUD and ADHD-CU risk genes across a range of tissues to identify affected tissues. First gene-based associations across the entire genome were calculated using MAGMA^25^ (including only concordant variants), second, we tested for a relationship between the gene-based associations and tissue-specific gene expressions in 54 tissues from GTEx^26^. This showed a significant association of ADHD-CUD genes with high expression in four brain tissues including the cortex (Supplementary Figure 5.a), but no increased expression of ADHD-CU genes in brain tissues were observed (Supplementary Figure 5.b).

### Expression of ADHD-CUD and ADHD-CU risk genes across brain developmental stages

To further explore the differences in expression of ADHD-CUD and ADHD-CU genes, we identified candidate risk genes and linked those to gene expression across brain developmental stages. First concordant ADHD-CUD and ADHD-CU genome-wide significant risk loci with high posterior probability of contributions from both phenotypes, were fine-mapped to identify sets of Bayesian credible variants^27^, with each set most likely (probability > 95%) including a causal variant (Supplementary Tables 12 and 13). Subsequently, credible variants were linked to genes using the online platform FUMA^28^ and data on genomic position, information about expression quantitative trait loci (eQTLs) and chromatin interaction mapping in human brain tissue (datasets selected are listed in the Supplementary Note). We identified 195 potential risk genes for ADHD-CUD (Supplementary Table 14) and 17 for ADHD-CU (Supplementary Table 15). The expression of identified candidate genes was then evaluated across 12 brain developmental stages using RNA-sequencing data of neocortex brain developmental stages from BrainSpan. ADHD-CUD genes (167 genes with high quality data in BrainSpan) demonstrated significant increased expression across all neocortex brain developmental stages when compared to background genes (Supplementary Table 16). When comparing expression of ADHD-CUD risk genes to ADHD-CU risk genes (16 genes with high quality data in BrainSpan; 4 overlap the ADHD-CUD gene set), ADHD-CUD genes demonstrated the highest mean expression at all brain developmental stages and significant overall mean expression when compared to the mean expression of ADHD-CU risk genes (P=2.73×10^-123^; Figure 1; Supplementary Table 16). Random down-sampling of ADHD-CUD genes 10,000 times to sets corresponding to the size of the ADHD-CU gene set (12 genes), strongly supported the finding of ADHD-CUD genes having significantly higher neocortical gene-expression than ADHD-CU genes no matter which genes were randomly assembled into the subset. 95% of the 1000 down sampled ADHD-CUD gene sets had significantly different expression than the ADHD-CU genes and for 91% of the down sampled gene sets it was due to an over-expression of ADHD-CUD genes compared to ADHD-CU genes (Supplementary Figure 6.a. and 6.b.).

**Figure 1.**
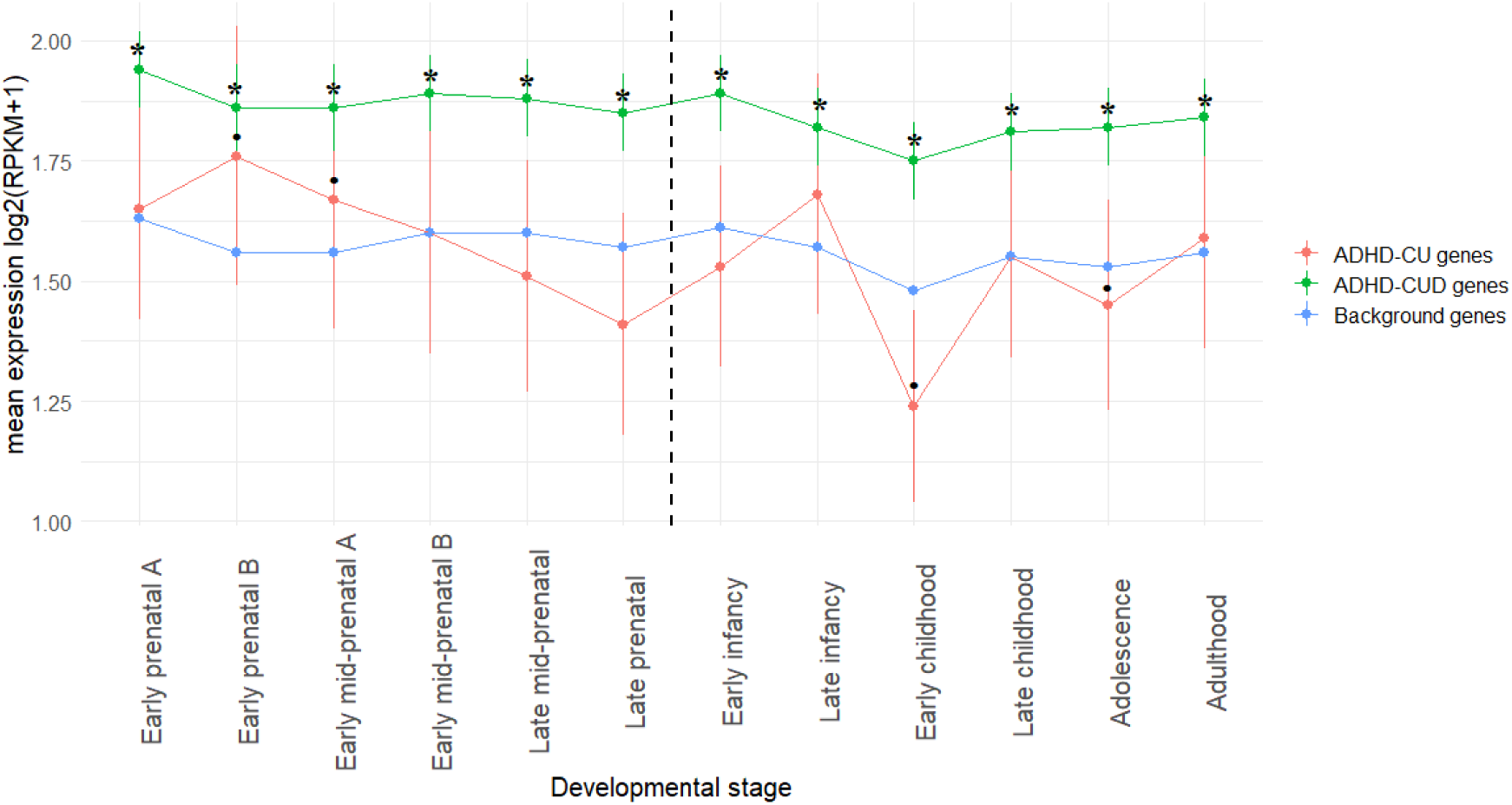
ADHD-CUD and ADHD-CU risk gene expression across brain developmental stages. Mean expression (measured as Reads Per Kilobase of transcript per Million mapped reads (RPKM)) on the y-axis of ADHD-CUD candidate risk genes (167 genes with high quality data in BrainSpan) in green, and ADHD-CU genes (16 genes with high quality data in BrainSpan) in red, across neocortex brain developmental stages in BrainSpan, on the x-axis. Vertical lines represent standard errors and the dotted vertical line in the middle indicate pre- or postnatal stages. *Indicate significant different expression compared to background genes (P-values less than P = 0.05). Expression of background genes, in blue, include expressed BrainSpan genes not in either the ADHD-CUD and/or in the ADHD-CU gene-sets. See also Supplementary Figure 6 for results evaluating expression differences when down-sampling the ADHD-CUD risk gene set to the same size as the ADHD-CU risk gene set.

### Transcriptome-wide association studies (TWAS)

We performed TWAS of the imputed gene-expressions to identify gene expressions associated with ADHD-CUD and ADHD-CU. This was done using S-prediXcan prediction models trained on GTEx^26^ (v. 8) data for 13 brain tissues, and summary statistics including both concordant and discordant variants. Subsequently, individual S-prediXcan^29^ results were combined in a meta-analysis using S-MulTiXcan^30^ to estimate overall association of the genetically regulated gene-expression in brain with the phenotypes. The expression of 70 genes were significantly associated with ADHD-CUD overall in brain tissues (TWAS- supplementary Tables 1), 36 were also exome-wide significantly associated with ADHD-CUD in MAGMA gene-based association analysis (Supplementary Table 19), 49 were also identified when linking credible variants to functional genomics data in FUMA (Supplementary Table 17), and 34 genes were identified by all three methods (TWAS S-MulTiXcan, MAGMA and FUMA; TWAS Supplementary Table 3). For ADHD-CU, 30 significant TWAS genes for association of overall brain expression with ADHD-CU were identified (TWAS-supplementary Tables 2), only one overlapped MAGMA genes (Supplementary Table 18) and genes identified using functional genomics data in FUMA (Supplementary Table 15). Thus, there were larger consistency in risk genes identified across methods and approaches for ADHD-CUD than for ADHD-CU.

### SNP-heritability and genetic correlations with other phenotypes

Genetic correlation analyses were performed to evaluate the extent to which the genetics of ADHD-CUD and ADHD-CU overlap other phenotypes. We calculated the genetic correlations (r_g_) of ADHD-CUD and ADHD-CU with 10 phenotypes related to (I) psychiatric disorders: schizophrenia (SZ)^31^, autism spectrum disorder (ASD)^32^, major depressive disorder (MDD)^33^, SUD (alcohol use disorder [AUD]^34^, opioid use disorder [OUD] ^35^), (II) cognition: educational attainment^36^ (III) substance use: smoking initiation^37^, drinks per week^37^ and (IV) other phenotypes highly correlated with ADHD: insomnia^38^, age at first birth (AFB)^39^). This was done using Linkage disequilibrium score regression (LDSC)^40^ and summary statistics only containing variants with concordant direction of effect in the GWAS of ADHD-CUD and ADHD-CU. For comparison r_g_ of ADHD with the 10 phenotypes were also calculated (Supplementary Table 19).

We identified significantly less negative r_g_ of ADHD-CU with educational attainment (r_g_ = - 0.19, s.e. = 0.04) and AFB (r_g_ = -0.45, s.e. = 0.03) compared to ADHD-CUD (r_gEA_ = -0.57, s.e. = 0.03, r_gAFB_ = -0.72, s.e. = 0.03) and ADHD (r_gEA_ = -0.53, s.e. = 0.02, r_gAFB_ = -0.66, s.e. = 0.02) (Supplementary Table 20, Figure 2). Both ADHD-CUD and ADHD-CU demonstrated a significant larger genetic correlation with SZ (r_gADHD-CUD_ = 0.42, s.e. =0.04; r_gADHD-CU_ = 0.33, s.e. = 0.04) than observed for ADHD (r_g_ = 0.19, s.e. 0.03). For SUD and substance use phenotypes, we identified a significantly higher r_g_ of ADHD-CUD with AUD (r_g_ = 0.72, s.e. = 0.04) and OUD (r_g_ = 0.76, s.e. = 0.04) than observed for both ADHD-CU (r_gAUD_ = 0.50, s.e. = 0.05; r_gOUD_ = 0.50, s.e. = 0.07) and ADHD (r_gAUD_ = 0.38, s.e. = 0.04; r_gOUD_ = 0.34, s.e. = 0.05), while both ADHD-CUD and ADHD-CU demonstrated significantly higher r_g_ with substance use i.e., with drinks per week (r_gADHD-CUD_ = 0.36, s.e. = 0.04; r_gADHD-CU_ = 0.43, s.e. = 0.04) and smoking initiation (r_gADHD-CUD_ = 0.77, s.e. = 0.02; r_gADHD-CU_ = 0.80, s.e. = 0.02) than observed for ADHD (r_gDrinks_per_week_ = 0.15, s.e. = 0.03, r_gSmoking_ = 0.60, s.e. = 0.02)(Supplementary Table 20). These results indicate that the shared genetic components of ADHD with CUD and CU are similarly related to substance use (smoking and drinks per week), while they differ with respect to SUD phenotypes where ADHD-CUD concordant variants increase r_g_ and ADHD-CU concordant variants decrease r_g_ relative to the r_g_ observed for ADHD (Figure 2).

**Figure 2.**
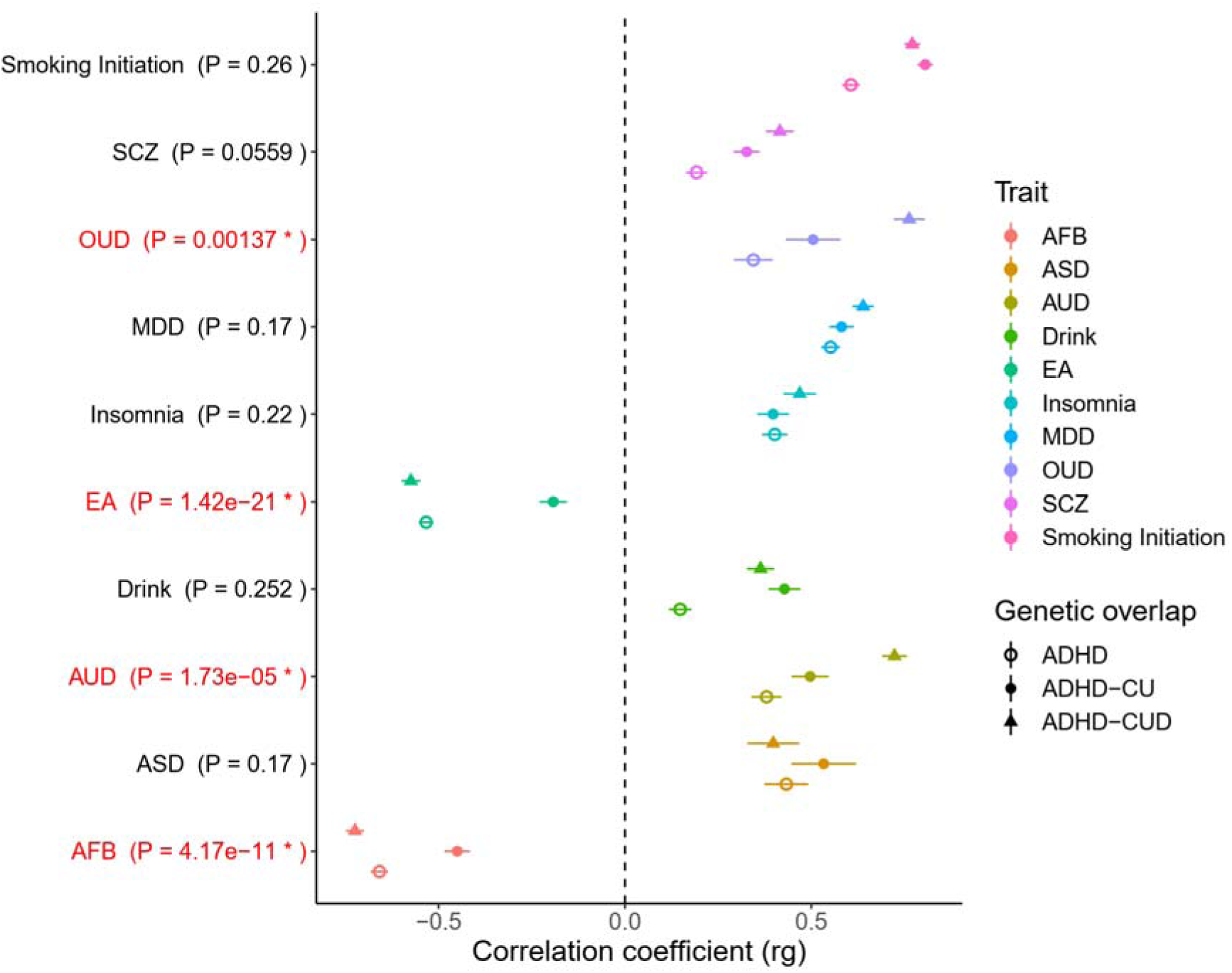
Genetic correlation of ADHD-CUD, ADHD-CU and ADHD with other phenotypes. Genetic correlation (r_g_) of ADHD-CUD (marked by a triangle; N=1,150,250), ADHD-CU (marked by a filled circle; N=387,616) and ADHD (marked by a circle with no fill; N = 225,534) with 10 other phenotypes: age at first birth (AFB), autism spectrum disorder (ASD), alcohol use disorder (AUD), drinks per week (Drink), educational attainment (EA), insomnia, major depressive disorder (MDD), opioid use disorder (OUD), schizophrenia (SCZ), smoking initiation. P-values in parentheses indicate significant difference in r_g_ of the phenotype with ADHD-CUD compared to the r_g_ of the phenotype with ADHD-CU. The symbols indicate r_g_ and the horizontal lines the standard error of r_g_. All pair-wise differences in r_g_ can be found in Supplementary Table 20.

### Load of rare variants in individuals with ADHD and comorbid CUD

We assessed the load of rare deleterious variants in individuals with ADHD and comorbid CUD in exome sequencing data from the Danish iPSYCH cohort. We compared the load of rare protein truncating variants (PTVs) and rare severe damaging missense variants (SevereDMVs) in ADHD-CUD (N=333) to individuals with ADHD without comorbid CUD (N=3,483) and to control individuals (N=8,951). None of the groups – ADHD-CUD, ADHD-only or controls - included individuals diagnosed with intellectual disability (ID), bipolar disorder (BPD), ASD, or SZ. We identified a higher overall load of rare PTVs and rare SevereDMVs in ADHD-CUD compared to ADHD-only (P = 0.03; Figure 3; Supplementary Table 21). In sub-analyses in sets of genes grouped by their probability of loss-of-function intolerance, the load of rare PTVs+SevereDMVs in evolutionary constrained genes (pLI > 0.9^41^, 2,811 autosomal genes) in ADHD-CUD was similar to what was found for ADHD-only (Supplementary Figure 7), however not significantly increased compared to controls, probably due to low power. Rare PTVs+SevereDMVs in less constrained genes (0 < pLI < 0.5; 14,267 autosomal genes) were significantly higher in ADHD-CUD compared to ADHD-only (P = 0.014; Figure 3; Supplementary Table 21). This suggests that the main rare variant component driving the overall higher load of rare PTVs+SevereDMVs in ADHD-CUD compared to ADHD without comorbidities are variants in genes with high tolerance to deleterious mutations.

**Figure 3.**
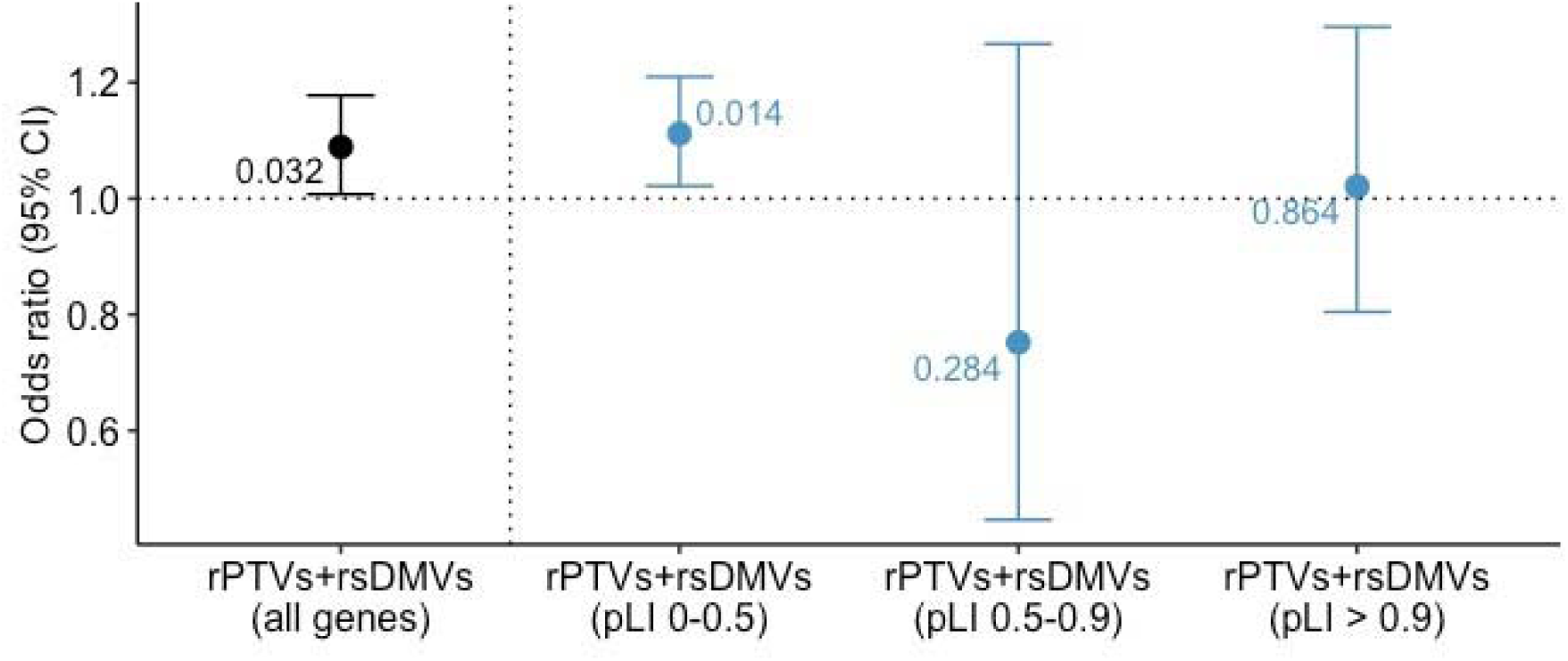
Rare variant load in ADHD-CUD compared to ADHD without CUD. Odds ratio (and corresponding 95% confidence intervals represented by vertical lines) on the y-axis from logistic regression testing for the load of rare (rPTVs and rare severe damaging missense variants (rare SevereDMVs) in ADHD-CUD (N=333) compared to ADHD without CUD (ADHD-only; N=3,483) in all genes (marked in black). Values next to the vertical lines are P-values. Sub-analyses (marked in blue) of the load of rare PTVs+SevereDMVs in ADHD-CUD vs ADHD-only in genes stratified by their pLI score: (I) genes with high tolerance to loss of function mutations (0 < pLI < 0.5), (II) less constrained genes (0.5 < pLI < 0.9), (III) evolutionarily constrained genes (pLI > 0.9). For the three sub-analyses P < 0.0167 is considered significant.

### Dissecting the genetic architecture of CUD comorbidity in ADHD using PGS

Individuals diagnosed with ADHD are at an increased risk of developing CUD, but not much is known about the specific genetic architecture of those that develop the comorbid condition compared to those that do not. To explore this, we performed PGS analyses using individual level genotype data from individuals in the Danish iPSYCH cohort^18,19^ with ADHD-CUD (N= 2,079) and individuals with ADHD without CUD (N = 23,466; other comorbidities are not excluded) benchmarked against a control group (N= 37,246). Twelve PGSs were analysed within the domains of psychiatric disorders (ADHD, ASD, SZ, MDD, BPD), cognition (educational attainment), SUD (CUD, AUD, OUD), substance use (smoking initiation, drinks per week, CU) (Supplementary Table 22). Comorbid ADHD-CUD individuals demonstrated a higher PGS for all evaluated scores (except ASD) compared to ADHD without CUD (or more negative for educational attainment) (Figure 4; Supplementary Table 23). When comparing the PGS of females and males with ADHD-CUD no sex differences were observed (Supplementary Figure 8, Supplementary Table 24). In summary, individuals with both ADHD and CUD were found to have higher polygenic risk load not only for SUD but also for a range of other psychiatric disorders and a decreased load of variants associated with increased educational performance compared to ADHD individuals without CUD, while no differences were observed between males and females with ADHD- CUD.

**Figure 4.**
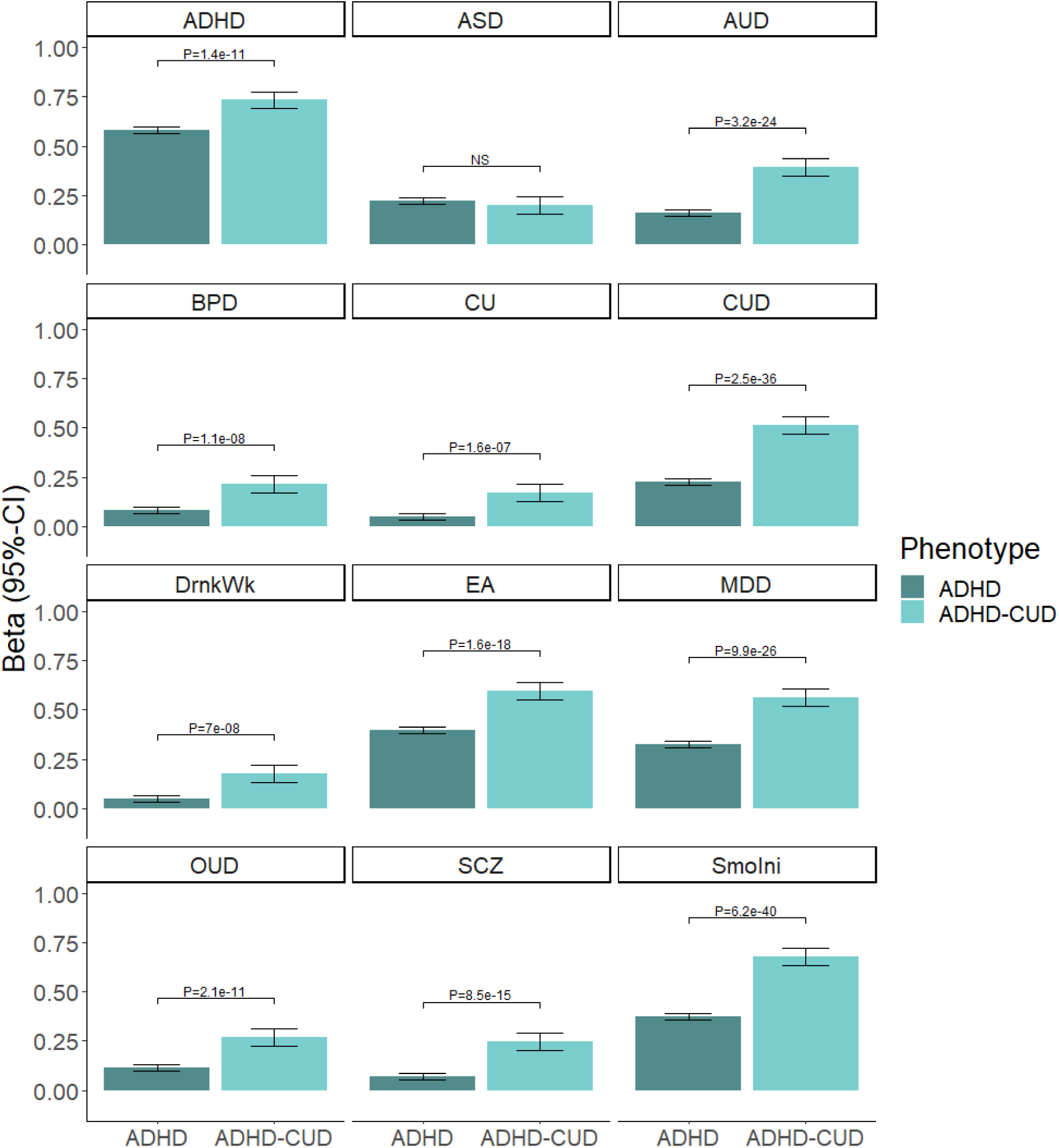
Multivariate PGS analyses in ADHD-CUD and ADHD without CUD. PGS analyses of ADHD-CUD (N = 2,079) vs ADHD without CUD (ADHD; N = 23,466), on the x-axis bench marked against controls (N = 37,246). The slope (beta) of the linear regression (95% confidence interval [CI]) is shown on the y-axis. Significant difference (P-value) between beta for ADHD-CUD and ADHD without CUD is indicated with a horizontal line with P-value above, i.e., the Wald test of equal group effect (see also Supplementary Table 23). NS indicate no significant difference. PGSs analysed are ADHD, autism spectrum disorder (ASD), alcohol use disorder (AUD), bipolar disorder (BPD), CU, CUD, drinks per week (DrkWk), educational attainment (EA), major depressive disorder (MDD), opioid use disorder (OUD), schizophrenia (SCZ), smoking initiation (SmokIni).

### Risk of comorbid CUD in individuals with ADHD stratified by their PGS

iPSYCH is a population-based case-cohort including all individuals diagnosed with ADHD in Denmark born between 1981 and 2008, with longitudinal information about comorbid diagnoses from the Danish registries. This gives the unique possibility of estimating the absolute risk that an individual with ADHD will develop CUD before a certain age depending on the PGS. To do this we stratified individuals with ADHD in the iPSYCH cohort (N=25,545) into quantiles depending on their PGS load of the 12 PGSs described above (Supplementary Table 22). Subsequently, cox regression was performed to calculate the cumulative absolute risk and hazard rate ratios (HRR) for developing comorbid CUD from 10 years to 30 years of age within PGS groups. For comparison the same analysis was done in population-based controls excluding individuals with ADHD (N=37,246). We found that the HRR of comorbid CUD generally increased with increasing PGS, and the highest HRR was observed in the 4^th^ CUD-PRS quantile with a HRR_4/1_ = 1.99 (s.e. = 0.07, P = 1.7×10^-21^) compared to the first decile (Supplementary Figure 9, Supplementary Table 25). The highest absolute risk was observed for the smoking-PRS with the trajectory for the 4^th^ quantile reaching an absolute risk of 23.2% at the age of 30 years, which was significantly higher than the absolute risk of 12.7% reached in the lowest quantile (Supplementary Table 25). The results were similar for the CUD-PGS with an absolute risk of 21.9% in the 4^th^ quantile (Figure 5) and for the ADHD-PGS with an absolute risk of 19.9% in the 4^th^ quantile, however the ADHD-PGS was less able to stratify individuals into four distinguishable risk groups (Supplementary Figure 9). For comparison, the absolute risk for CUD among population-based controls in the high-risk smoking-PGS and CUD-PGS groups reached 1.8% and 1.6% (Figure 5) respectively.

**Figure 5.**
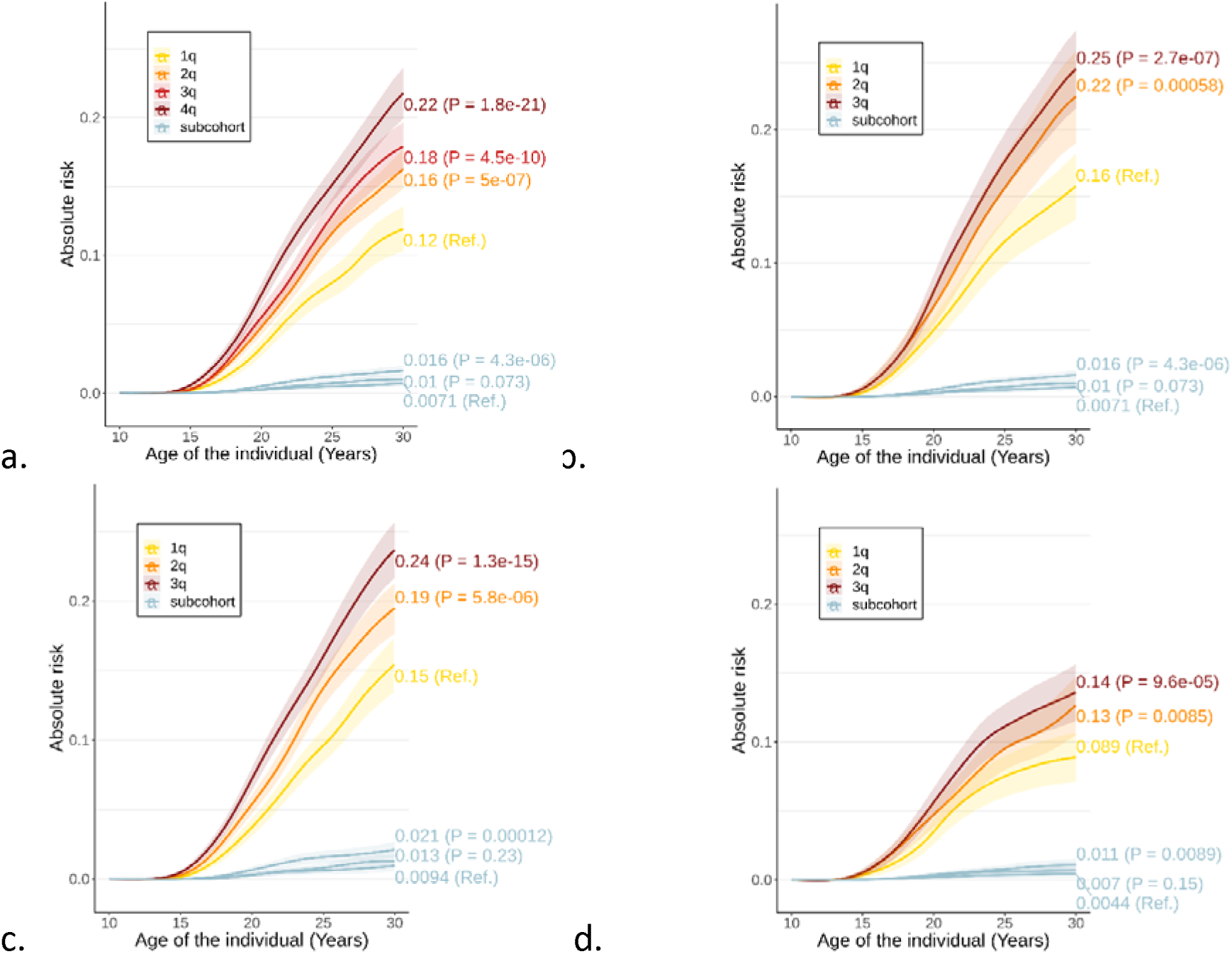
Absolute risk of comorbid CUD among individuals with ADHD stratified by their CUD-PGS. Absolute risk (95% CI) (on the y-axis) over time (on the x-axis) of CUD among individuals stratified into quartiles (1q-4q) or tertials (1q-3q) based on their CUD-PGS in the iPSYCH cohort. The colored values at the right on the figures indicate absolute risk of CUD at age 30 and the P-values in the parentheses indicate difference in relative risk of comorbid CUD in each PGS group relative to quartile/tertial one (yellow line (ref)) **(a)** all individuals with ADHD (N=25545) **(b)** among individuals with ADHD having at least one parent diagnosed with a psychiatric disorder (N=6,999) **(c)** among males with ADHD (N=17,682) **(d)** among females with ADHD (N = 7,863). Absolute risk (95% CI) of CUD among individuals in the population-based sub-cohort (without ADHD) stratified into tertials based on their CUD-PGS (N = 37,246), is shown in light blue (the blue values indicate absolute risk of CUD at age 30 and corresponding P-values indicate difference in relative risk of CUD in a tertial relative to tertial one (ref)). In the sex-specific analyses the controls are restricted to either males (N = 18,960) or females (N = 18,607).

When estimating the risk for comorbid CUD among individuals with ADHD who have one or both parents diagnosed with a psychiatric disorder (N= 6,999) the absolute risk increased further to 25% in the 4^th^ quantile for both the CUD-PGS (Figure 5) and the smoking-PGS (Supplementary Figure 10, Supplementary Table 26). Sex-specific analyses identified substantial differences in the absolute risk of comorbid CUD between males (N = 17,682) and females (N = 7,863) with ADHD, with a ∼10% higher CUD risk among males than females in both the high-risk CUD-PGS group and smoking-PGS group (PGS-CUD: absolute risk for males 24% and 14% among females; Figure 5, Supplementary Figure 11 and 12 and Supplementary Table 27).

## Discussion

Here we dissect the shared genetic architecture of ADHD, CUD and CU through large-scale GWAS cross-disorder analyses based on the most recent GWAS of the single disorders and novel exome sequencing data. First, we evaluated the polygenic overlap between the phenotypes, and identified a higher r_g_ between ADHD and CUD (r_g_ = 0.57) than between ADHD and CU (r_g_ = 0.20). This was reflected in a larger sharing of concordant influencing variants between ADHD and CUD (7,000 variants; 75% shared and concordant directional effect) than between ADHD and CU (5,100 variants; 60% shared and concordant), refining the previously reported genetic overlap^14^ appreciably. We also found that most of the effect of ADHD genetics on CUD is direct and only a small proportion (12%) is mediated by CU genetics, which suggests CU to be less a risk factor for CUD than a genetic predisposition of ADHD. MR analyses identified significant causal bidirectional relationships between ADHD and CUD with a larger effect of CUD on ADHD than the reverse. MR analyses have previously reported a causal effect of ADHD on CU^42,43^. The large number of shared variants between ADHD and CUD makes it important to address pleiotropy in the MR analyses as pleiotropy violates MR assumptions. Even though we performed sensitivity analyses we did not apply the exhaustive list of methods to handle, detect or control for pleiotropic instrumental variables, and we cannot completely exclude potential biases introduced by pleiotropy.

We identified 36 genome-wide significant loci for ADHD-CUD, with concordant direction of effect of these 17 are novel loci i.e., they did not reach genome-wide significance in the single disorders. The most strongly associated of these is located on the p-arm of chromosome 3 (rs3774800, OR= 1.05, s.e. = 0.006, P = 1.79×10^-15^), in a broad LD region, containing credible variants that were linked to 69 genes through integration with functional genomics data. The second most strongly associated new locus is located on the q-arm of chromosome 3, intragenic in *CADM2* (rs11915747, OR = 1.05, s.e.= 0.006, P = 6.93×10^-14^). In the cross-disorder GWAS of ADHD-CU, 23 loci were identified, 11 of them with concordant direction of effects and strong contributions to the association signal from both phenotypes. The most strongly associated concordant locus (rs62263912, OR = 1.06, s.e. = 0.007, P= 7.82×10^-16^) was also located intragenic in *CADM2*. *CADM2* encodes the synaptic cell adhesion molecule 1, and was previously identified in a GWAS of CU^21^ and has also been linked to sensation seeking^44^ and impulsive behaviors^45^.

The larger number of concordant risk loci for ADHD-CUD goes well in line with the MiXeR analyses finding that 75% of shared variants have concordant direction of effect, and the observation that a little less than half of the genome-wide significant loci for ADHD-CU variants have discordant directions corresponds well with the estimate that around 40% of the shared influencing variants have discordant effects on ADHD and CU.

Besides *CADM2* four other loci were genome-wide significantly associated with both ADHD-CUD and ADHD-CU with concordant directions of effect, including an interesting locus on chromosome 11. This locus is also associated with both CUD and CU^21^, and has been linked to tobacco smoking^37^ and alcohol dependence^46^. In the present study both index variants were located in *NCAM1*, but for ADHD-CUD, three independent genome-wide significant variants (two remained significant after conditional analyses) were observed downstream *NCAM1* representing a secondary independent locus not associated with ADHD- CU. One of these variants is located in *DRD2*, and the other two variants are located downstream *DRD2* but were linked to *DRD2* by functional annotations (rs6589386 is a significant eQTL for *DRD2* in the cerebellar hemisphere [P=6.5×10^-6^] and rs2014920 is located in a chromatin region interacting with *DRD2*; see also Supplementary Table 14). *DRD2* was also one of the 34 genes identified as a risk gene for ADHD-CUD by three different methods (gene-based magma test, by integration with functional genomics data in FUMA and by TWAS). Identification of *DRD2* as a risk gene for ADHD-CUD is of high interest considering the role of dopamine in CUD^47^ and in ADHD, where ADHD symptoms have been linked to dysfunctional dopamine regulation in the brain^48^. We have previously identified a significant association of ADHD risk genes with high expression in dopaminergic neurons^3^, but the present result is the first to directly link a single locus implicating a dopamine receptor/transporter to ADHD (subgroups) based on GWAS results. The TWAS identified an overall significantly decreased expression of *DRD2* in the meta-analysis of expression across brain regions (P= 4.8×10^-9^, Z_mean_ = -0.19) with the most significantly decreased expression found in the cerebellar hemisphere (1.57×10^-8^, Z = -5.65), a brain region that has been found to be involved in reward and cognitive processing^49,50^.

ADHD demonstrated a significant genetic correlation with both CUD and CU, but our analyses suggest several differences in the shared concordant genetics of ADHD and CUD from the shared concordant genetics of ADHD and CU. ADHD-CUD associated genes have a significant increased expression in several brain regions (Supplementary Figure 5), and a significant increased expression across all brain developmental stages (Figure 1), which is not observed for ADHD-CU genes. The higher expression of ADHD-CUD genes across brain regions and developmental stages could indicate that these genes are more important for brain development and function than ADHD-CU genes. It should be noted that GWAS of ADHD^3^ and CUD^14^ have previously linked the disorders to genes with high expression in early brain development, while we observed increased expression of ADHD-CUD risk genes across all brain developmental stages. Our approach was different from what was used in the single disorder GWAS, so we cannot exclude that other approaches might link ADHD-CUD to genes with different patters with regard to prenatal and postnatal expressions.

When evaluating genetic correlations we found that variants associated with educational attainment, age at first birth, and SUD (AUD, OUD) contributed to differences in the concordant polygenic architecture of ADHD-CUD from ADHD-CU. ADHD-CUD has a significant higher r_g_ with SUD phenotypes (r_g_AUD_ = 0.72; r_g_OUD_ = 0.76) and significant lower r_g_ with educational attainment (r_g_EA_ = -0.58) and age at first birth (r_g_AFB_ = -0.73) when compared to those for ADHD-CU (r_g_AUD_ = 0.5; r_g_OUD_ = 0.5, r_g_EA_ = -0.19, r_g_AFB_ = -0.45) (Figure 2). These results are in line with previous findings suggesting that the genetic architecture of drug use is different from that of diagnosed SUD^14,17^, including opposite genetic correlations of EA with substance use (positive r_g_ ^14,17^) and SUD (negative r_g_ ^14,17^). Our results suggest that these genetic differences are also reflected in the genetics shared with ADHD. It could be speculated that ADHD-CUD risk variants affect pathological mechanisms shared across ADHD and SUD, while ADHD-CU associated variants influence less detrimental biological aspects of ADHD and substance use related to sensation seeking and risky behavior which have positive genetic correlations with educational attainment^51,52^.

A limitation of this study is potential heterogeneity and lack of specificity of the CU phenotype. The phenotype definition is life-time use, and therefore includes a range of users, from those that have used cannabis just once to those that have used it regularly and a minority who will progress to CUD. The impact of CUD is assumed to be small as the prevalence of CU is 22% in UK Biobank (which compose 78.2% of the samples in the CU GWAS), and thus much higher than the prevalence of CUD in UK which is around 1.2%^53^. Additionally, the different samples varied regarding the age of the participants, the prevalence of cannabis use due to different cohort ascertainments, and access to cannabis due to differences in policy regarding cannabis use between countries. These factors may have introduced some heterogeneity and thus reduced power to detect genetic associations, but despite that the study identified eight genome-wide significant loci. Even though the CU phenotype is somewhat unspecific, and could be influenced by other traits related to cannabis use, like risky behaviour and novelty seeking, the phenotype does capture genetic aspects of substance use, since CU demonstrate significant higher r_g_ with substance use and SUD phenotypes than what was found for ADHD.

We have previously found an increased load of rare loss of function variants in individuals with ADHD compared to controls^54^ but very little is known about the role of rare variants in CUD. Here we identified a significant increased load of rare PTVs+SevereDMVs in individuals with ADHD-CUD compared to ADHD-only when considering all autosomal genes, and in particular, in evolutionarily less constrained genes (i.e., genes with 0 < pLI < 0.5). This suggests that the genetic risk component contributing to CUD, besides what is shared with ADHD, involves genes that are expressed in fewer tissues and are less important for cell survival, as these features characterize less constrained genes compared to evolutionary constrained genes^41,55^. Besides increased load of rare PTVs+SevereDMVs, PGS analyses in the iPSYCH cohort also identified the load of common risk variants associated with other psychiatric disorders (except ASD) and SUD to be increased in comorbid individuals (Figure 4). Stratification of iPSYCH individuals with ADHD into PGS quantiles found an absolute risk of 22% in the high-risk CUD-PGS group, which was strikingly higher than observed for controls in the high-risk CUD-PGS group, where the absolute risk of CUD reached 1.6% (Figure 5). When evaluating the cumulative absolute risk among individuals having one or more parents diagnosed with a psychiatric disorder the absolute risk increased further to 24% in the high CUD-PRS group.

In the PGS analyses we did not find significant differences between males and females with ADHD-CUD however we identified substantial sex differences in the absolute risk, with males having almost twice the risk for comorbid CUD in all CUD-PGS groups compared to females, also among controls. This means that given the same genetic risk load a much larger proportion of males develop SUD, suggesting that males are more susceptible to some environmental factors than females. Overall, our findings provide a statistically significant basis for considering the use of PGSs in clinical settings to identify individuals with ADHD at high risk for comorbid CUD. These predictions could be further elaborated by incorporating other relevant information that modifies the risk, like parental diagnosis with a psychiatric disorder or sex. For instance, targeted efforts could be envisaged towards males with the highest CUD-PGS. This could include preventive programs and/or monitoring to identify potential early stages of CUD for early intervention^56^ to prevent the manifestation of severe CUD. The use of PGS in clinical settings with such potential applications is warranted^57,58^.

Overall, our results suggest that ADHD shares a larger number of concordant influencing variants with CUD than with CU and that ADHD-CUD associated variants affect genes with high expression across several brain tissues and developmental stages, including *DRD2*. ADHD with comorbid CUD is associated with increased polygenic risk load of both common and rare deleterious variants, and this information could potentially be incorporated into prediction tools to identify vulnerable individuals with ADHD at high risk for CUD.

## Methods

An overview of the workflow and methods are illustrated in Supplementary Figures 13-15. The study was approved by the local scientific ethics committees and IRBs. The iPSYCH study was approved by the Scientific Ethics Committee in the Central Denmark Region (Case No 1-10-72-287-12) and the Danish Data Protection Agency. In accordance with Danish legislation, the Danish Scientific Ethics Committee has, for this study, waived the need for specific informed consent in biomedical research based on existing biobanks.

### GWAS summary statistics

This study is based on the largest available summary statistics from GWAS of ADHD, CUD and CU, of European ancestry. The GWAS meta-analysis of ADHD^3^ included data on 38,691 individuals with ADHD and 186,843 controls from iPSYCH, deCODE, and ten ADHD cohorts collected by the Psychiatric Genomics Consortium. The CUD GWAS^20^ meta-analysis included data 42,281 individuals with CUD and 843,744 controls from the Department of Veterans Affairs, Million Veteran Program (MVP), iPSYCH, deCODE, Yale-Penn 3, Partners Healthcare Biobank and 17 cohorts from the Psychiatric Genomics Consortium. The CU GWAS meta-analysis^21^ included data on 162,082 individuals with self-reported lifetime cannabis use from 16 cohorts from the International Cannabis Consortium and UK Biobank. Most of the individuals with ADHD or CUD in the published GWAS were diagnosed according to the ICD or DSM diagnosis criteria. An overview of the cohorts included in the GWAS, and a brief overview of the diagnostic criteria can be found in Supplementary Table 28. Detailed information about cohort ascertainment and phenotype definitions are described elsewhere (see references in Supplementary Table 28). In the CU GWAS data were available for all individuals on whether an individual reported having ever used cannabis during their lifetime: yes versus no. Although phrasing of the question slightly differed between cohorts, the answer reflected lifetime cannabis use in all the samples (a short description can be found in Supplementary Table 28). Due to differences in recruitment strategies, cultural and temporal differences, combined with differences in cannabis availability between countries, there was a range in the percentage of lifetime use with a mean of 36,1% across cohorts.

The published GWAS of ADHD and CUD include analyses that evaluate potential genetic heterogeneity across cohorts due to differences in diagnostic criteria, ascertainment strategies or other differentiating factors. The studies found significant high genetic correlations between cohorts in both the ADHD GWAS (r_g_ = 0.82 – 0.92) and the CUD GWAS (r_g_ = 0.71 - 0.87) supporting a high consistency in the polygenic architecture underlying the phenotypes across samples. Genetic correlations between cohorts included in the CU GWAS are not reported in the published study and we can therefore not rule out heterogeneity between cohorts. However, the study identified eight genome-wide significant loci, which indicates that genetic heterogeneity is not a major issue, since this would most likely lead to decreased power.

### Genetic overlap between ADHD, CUD and CU

The number of shared common variants between ADHD and either CUD or CU were estimated using bivariate mixture modelling MiXeR^23^. MiXeR quantifies the polygenic overlap (i.e., total number of variants that explain 90% of the SNP heritability) irrespective of the directions of effect on the two phenotypes, using summary statistics from GWAS. First summary statistics from GWAS meta-analysis of ADHD^3^, CU^21^ and CUD were prepared by filtering out variants in the region on chromosome 6: 26000000-34000000 bases, and prepared with the munge function from LDSC^40^ using effective sample sizes calculated as N_eff_=4/(1/N_cas_ + 1/N_con_). The MiXeR analysis was run in two steps with default settings. First univariate Gaussian Mixture Modelling analysis was run to determine the polygenicity (proportion of non-zero effect SNPs) and discoverability (effect size variance) separately for ADHD, CU, and CUD. In a second step, a bivariate analysis uses estimates from the univariate analysis to determine the number of causal variants shared between the two phenotypes, the number of variants specific to each of the two phenotypes and variants with zero-effects on both phenotypes. These parameters were combined to quantify the polygenic overlap and genetic correlations between ADHD and either CU or CUD.

### Genomic SEM, mediation and Mendelian randomization analyses

We used structural equation modelling implemented in Genomic SEM^22^ to estimate the loadings of ADHD, CU and CUD on a shared latent genetic factor. For this we used summary statistics of ADHD, CUD and CU and harmonized the files using the munge function in LDSC^40^ as described above. We applied a common factor model, where the latent factor connecting the three phenotypes, represents the shared genetic architecture. This model was also constructed with a user-defined model, where the model was constructed to handle a Heywood case (i.e., a negative variance) observed for CUD, which was solved by fixing the variance to zero. Diagonally Weighted Least Square (DWLS) was chosen as the estimation method and the variance of the latent factor was constrained to be 1. Fit indices are not reported as this was a fully saturated model (i.e., *df* = 0).

We also used Genomic SEM to test whether the common variant liability to ADHD effects CUD directly or if the relationship is mediated through common variant liability to cannabis use. Multivariable LD-Score regression was run to obtain the genetic covariance matrix (S) and the corresponding sampling covariance matrix (V), where the population prevalence was set to 0.05 for ADHD^1^, 0.01 for CUD (the prevalence of CUD in Denmark^59^) and 0.44 for CU (estimated based on information in the paper^21^). LD scores and weights for the European population were obtained from the original LD score package (see URLs). In the model CUD (V3) was regressed on both CU (V2) and ADHD (V1), and CU (V2) was also regressed on ADHD (V1). This creates a path from ADHD to CUD directly and a path from ADHD via CU to CUD. A user specified mediation model was subsequently fitted using Genomic SEM. The mediation models were run with DWLS as the estimation method.

### Mendelian randomization

MR is a method that uses genetic variants as proxy for an exposure to evaluate the extent to which the exposure has a causal impact on an outcome. The causal relationship between ADHD and CUD was examined using bidirectional MR analyses i.e., we tested for the causal impact of ADHD (as exposure) on CUD (the outcome) and the reverse.

Instrumental variables in the analysis with ADHD as the exposure were genome-wide significant index variants identified in the newest ADHD GWAS^3^. The instrumental variables for CUD as the exposure were defined as independent genome-wide significant variants located with a distance of more than with kb > 500 kb and with a correlation (r^2^) < 0.001. Before MR was run, a harmonization step was performed to ensure effects were coded corresponding to the same reference allele in both the outcome and exposure dataset. After harmonization, 12 instrumental variables were identified for ADHD and 20 instrumental variables were identified for CUD. Inverse variance weighted MR was performed along with four sensitivity analyses: simple mode MR, weighted mode MR, weighted median MR, MR Egger regression and Generalized Summary-data-based MR method (GSMR). These analyzes were performed to handle potential problems with horizontal pleiotropy. Moreover, Q-heterogeneity tests, single variant analyses, leave-one (variant) -out analyses, funnel plots and a test of the MR Egger regression intercept were performed to detect potential problems with horizontal pleiotropy. The harmonization step, MR analyses and sensitivity analyses were done using the R-package TwoSampleMR (see URL) with default settings, except from the GSMR^60^ which was implemented in GCTA^61^. GSMR was performed with the newest version of the HEIDI-outlier filtering method. GSMR was run with a 1000 Genomes phase 3 European reference file and default settings.

In all MR analyses, the Q-heterogeneity tests were significant, indicating the MR analyzes could be biased by horizontal pleiotropy. The IVW MR and the sensitivity analyzes along with the Q heterogeneity test and the test of the MR Egger intercept were therefore rerun without variants detected as outliers by HEIDI in the GSMR.

To ensure no bias from weak instruments we calculated F-statistics for each of the SNPs and an overall F-statistic for ADHD and CUD. An F-statistic above 10 is recommended to avoid problems with weak instruments. The F-statistics were estimated using the proportion of variance in the exposure explained by instrumental variables, also called the R^2^. The r^2^ for each instrumental variable was calculated as:

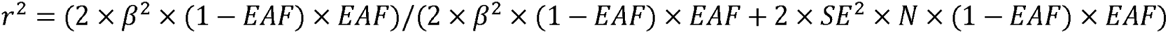

Where β is the effect size of the exposure, SE is the standard error, EAF is the effect allele frequency and N is the sample size.

An F-statistic was then calculated for each SNP:

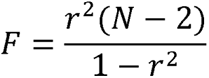

An overall F statistic for the risk factor was calculated by summing the individual R^2^ and using the formula:

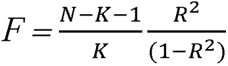

Where N is sample size and K is the number of instrumental variables. Besides calculating a F statistic for the risk factor, a lower limit of a one-sided 95 % confidence interval was estimated for the F parameter as seen in (REF)^62^.

### Cross-disorder GWAS meta-analyses

Two cross-disorder GWAS were performed for (1) ADHD^3^ (38,691 with ADHD; 186,843 controls) and CUD^20^ (42,281 with CUD; 843,744 controls) and (2) ADHD^3^ and CU^21^ (N= 162,082 individuals) using a fixed effect meta-analysis approach implemented in ASSET^24^. Unlike traditional meta-analysis methods, ASSET takes into account the direction of effects a variant has on multiple disorders, and outputs subsets of variants with the same directions of effect on the analyzed phenotypes or subsets of variants whose effect sizes are in the opposite direction on the analyzed phenotypes.

Before ASSET was run, the ADHD, CUD and CU summary statistics were harmonized and aligned to a reference genome to ensure effect alleles and effect sizes were aligned between the three summary statistics. The harmonization was performed with the Bioconductor package MungeSumstats using the reference genomes SNPlocs.Hsapiens.dbSNP144.GRCH37 and BSgenome.Hsapies.1000genomes.hs37d5 also from Bioconductor. ASSET was run using the h.traits() function and one-sided and two-sided meta-analyses were conducted with default settings. The analyses were constrained to variants shared between the ADHD summary statistics and either the CU or CUD summary statistics. We corrected for sample overlap by including an inter-study phenotypic correlation matrix. This was derived by estimating the intercepts from genetic correlation analysis in LDSC^40^ which were used as rough estimates of sample overlap between ADHD and respectively CUD and CU. Two datasets were created for each of the two cross-disorder GWAS. One dataset was created based on results from the one-sided subset test, where only variants having the same direction of effect for both phenotypes were kept (the concordant variants) and another dataset was created from the two-sided subset test, where only variants with opposite directions of effect on the two phenotypes were kept (the discordant variants). A P-value = 5×10^-5^ was used to declare variants genome-wide significantly associated.

Independent index variants and secondary lead variants were defined as variants with a P-value < 5×10^-8^ and low linkage disequilibrium (LD) between them (r^2^ < 0.1). LD blocks of independent variants with a distance less than 250 kb of each other were merged into one risk locus. One genome-wide significant risk locus in the ADHD-CU GWAS and four genome-wide significant risk loci in the ADHD-CUD GWAS had more than one independent lead variant. COJO^63^ implemented GCTA v1.94.1^61^ was used to examine if the secondary lead variants remained genome-wide significant after conditioning on the index variant in the locus.

Potential heterogeneity in effect size estimates between the phenotypes included in the cross-disorder GWAS was evaluated for all genome wide significant index variants and secondary lead variants. Heterogeneity was quantified by estimating the posterior probabilities (the m-value^64^) that the effect exists in both studies, using METASOFT^65^. Loci with m-values below 0.85 for one of the phenotypes were removed. METASOFT was run with an alpha set to 1.5 and sigma set to 0.4.

We performed a PheWAS, using PhennoScanner v2^66,67^ (see ULRs) to evaluate associations of ADHD-CUD index variants (and independent secondary variants) across more than 5000 GWAS studies implemented in the curated database.

### Tissue specific expression of ADHD-CUD and ADHD-CU genes

MAGMA v1.08^25^ implemented in FUMA v1.4.2 (Ref.^28^) was used to perform gene-based association analysis using the full summary statistics from the GWAS meta-analyses. Genome-wide significance was assessed through Bonferroni correction for the number of genes tested (CUD: *P* = 0.05/15,536 = 3.22×10^-6^; CU: *P* = 0.05/16,083 = 3.11×10^-6^). The relationships between tissue specific gene expression profiles and ADHD-gene associations was tested using MAGMA gene-property analysis of expression data from GTEx^26^ (54 tissue types) available in FUMA^28^ (See Supplementary Information for data sets selected in FUMA). Bonferroni correction was applied to correct for the number of tissues tested.

### Mapping of risk genes and their expression across brain developmental stages

In order to identify sets of causal variants we fine-mapped each genome-wide significant locus from both cross-disorder GWAS using three fine-mapping tools, FINEMAP v. 1.3.1(Ref. ^68^), PAINTOR v.3.0 (Ref.^69^) and CAVIARBF v.0.2.1 (Ref.^70^), using CAUSALdb-finemapping-pip downloaded from https://github.com/mulinlab/CAUSALdb-finemapping-pip^27^.

Genetic variants within 1 million base pairs of the index variants were used as input for the pipeline. We used a threshold 95% for the total posterior probability of the variants included in the credible sets and only variants claimed to be within the set by all three methods were included in the final credible set for each locus. For secondary lead variants the pipeline was run by first removing variants with stronger association signal than the secondary independent lead variants from the input file to get reliable results.

Credible variants were linked to genes based on genomic position and functional annotations in FUMA^28^. Protein coding genes were mapped if they were located with a distance of 10Kb up- or downstream index variants or if a credible variant was annotated to the gene based on eQTL data or chromatin interaction data from human brain (data sets used in the mapping can be found in the Supplementary Note). Gene mapping was performed with functional annotation filters set with default settings and no additional variant filtering by functional annotation was applied in the eQTL and chromatin interaction mapping.

The expression of mapped ADHD-CUD genes (N=195) and ADHD-CU genes (N=17) across neocortex brain developmental stages was evaluated in bulk RNA-sequencing data (v. 10) from BrainSpan (see URLs). Following (Ref.^71^) we only analyzed the following neocortical regions: dorsolateral prefrontal cortex, ventrolateral prefrontal cortex , medial prefrontal cortex, orbitofrontal cortex, primary motor cortex, primary somatosensory cortex, primary association cortex, inferior parietal cortex, superior temporal cortex, inferior temporal cortex, and primary visual cortex. The different developmental stages were defined as in the BrainSpan documentation (see URLs). Samples with poor quality were removed (RNA integrity number (RIN) < 7). Genes were defined as expressed if the Reads Per Kilobase of transcript per Million mapped reads (RPKM) was minimum 1 in at least 80 % of the samples for at least one neocortical region in one major temporal epoch. The expression data kept after filtering was log-transformed (log_2_[RPKM+1]). A two-sided paired t-test was used to test for differential expression of ADHD-CUD and ADHD-CU genes against a background gene set at each developmental stage and across all developmental stages. Due to small sample sizes for the individual developmental stages, the data was tested to see if the assumptions for a paired t-test were met. If the assumptions of the paired t-test were not met a Wilcoxon signed-rank test was performed instead. The background gene set included expressed BrainSpan genes not found in either the ADHD-CUD or ADHD-CU gene set. A two-sided paired t-test was also performed to determine differential expression between ADHD-CUD genes and ADHD-CU genes across developmental stages. Genes shared between the two gene sets were removed before the test (four genes). Bonferroni correction was applied correcting for the number of brain developmental stages analyzed (12 stages). Due to the large difference in sample size between the ADHD-CUD (167 genes) and the ADHD-CU (16 genes) gene sets, the ADHD-CUD gene set was randomly down-sampled 10,000 times to the same size as the ADHD-CU genes and tested using paired t-test, for difference in mean expression from the ADHD-CU genes.

### Transcriptome-wide association studies (TWAS) with S-prediXcan and S-MulTiXcan

TWAS of the genetically regulated gene expression was done using S-prediXcan^29^. This method use GWAS summary statistics as input and imputes the genetically regulated gene expression based on prediction models trained on external transcriptome data and test for association of the imputed gene expression with a phenotype. We used the and summary statistics from GWAS of ADHD-CUD and ADHD-CU including all variants, and gene expression predictions models based on mashr trained on GTEx data (v. 8), for 13 brain tissues: Amygdala, Anterior cingulate cortex (BA24), Caudate basal ganglia, Cerebellar hemisphere, Cerebellum, Cortex, Frontal cortex (BA9), Hippocampus, Hypothalamus, Nucleus accumbens basal ganglia, Putamen basal ganglia, Spinal cord cervical (c-1) and Substantia nigra. Prior to S-prediXcan analyses, the summary statistics were prepared in a harmonization, imputation and post-imputation step. In the harmonization step, the data was made compatible with a 1000 Genome Phase 3 European reference panel^72^ and updated to Human Genome hg38 format. Next, in the imputation and post-imputation steps missing variants in the summary statistics were imputed using the same 1000 Genomes Phase 3 reference data. S-prediXcan results for the 13 brain tissues listed above were a meta-analyzed using S-MulTiXcan^30^, to calculate gene expression associations across different brain tissues while handling potential correlations. The harmonization step, imputation step, S-prediXcan and S-MulTiXcan were run with default settings (see URLs).

### SNP-heritability and genetic correlations

SNP-heritabilities (h^2^_SNP_ ) and genetic correlations (r_g_ ) with 10 other phenotypes were estimated for ADHD-CUD and ADHD-CU using LDSC^40^ and summary statistics only containing concordant variants. The phenotypes included in the r_g_ analyses were within the domains of SUD (OUD, AUD), substance use (drinks per week, smoking initiation), psychiatric disorders (ASD, MDD, SZ) or highly genetically correlated with ADHD (insomnia, educational attainment, age at first birth) (see references in Supplementary Table 17). We applied Bonferroni correction and declared genetic correlations significantly different from zero if the P-value was lower than P = 0.0017 (0.05/30 tests). For comparison r_g_ between the above-mentioned phenotypes were also estimated with ADHD^3^. Block-jackknife method^73^ was used to test for significant differences between estimated genetic correlations. The block-jackknife method is a resampling method, where the difference between resampling genetic correlations is used to calculate a jackknife standard error. From this standard error a Z-statistic is estimated and used in a two-tailed Z-test to determine if the difference between two genetic correlations is significant different from zero. Here 200 resampling estimates were obtained, and Bonferroni correction was applied to correct for multiple testing (P < (0.05/30) = 0.002).

All analyzes were constrained to SNPs from the 1000 Genomes Phase 3 European reference panel^72^ with a MAF above 0.05, and LD scores which we calculated based on the European Haplotype reference consortium using LDSC with a 1 centiMorgan window.

### Load of rare variants in individuals with ADHD and comorbid CUD

We analyzed whole-exome sequencing data from iPSYCH to evaluate the role of rare- deleterious variants in ADHD comorbid with CUD. The sequencing data were obtained for a subset (n=34,544) of iPSYCH cohort, which was described in details in our previous study of the role of rare variants in childhood, persistent and late diagnosed ADHD^74^ including description of data quality control (QC), functional annotation, and rare variant burden analysis. In brief, the sequencing was executed across multiple phases — Pilot1, Wave 1, Wave 2, and Wave 3 — using an Illumina HiSeq platform at the Broad Institute’s Genomics Platform. After stringent QC excluding individuals with low quality sequencing, related individuals (one of each related pair with an piHAT value > 0.2 was excluded), and genetic outliers were removed based on PCA of a set of pruned high-quality variants with maf > 0.05. After QC 28,448 individuals remained for further analysis. By cross-referencing with diagnoses recorded in the Danish Psychiatric Central Research Registry^75^ and the Danish Patient Registry^76^ up to the year 2016, we categorized 333 individuals with comorbid ADHD and CUD (ADHD-CUD; ICD-10 codes: F90.0, F90.1, F98.8 for ADHD and F12.1-9 for CUD), 3,483 individuals with ADHD without CUD (ADHD-only), and 8,951 controls. None of these groups included individuals diagnosed with intellectual disability (ICD10: F70, F71, F72, F73, F78, F79), ASD (ICD10: F84.0, F84.1, F84.5, F84.8, F84.9), bipolar disorder (ICD10: F30-F31) or schizophrenia (ICD10: F20).

We defined rare variants as those with an allele count no greater than five across our iPSYCH dataset and the non-Finnish Europeans from non-psychiatric exome subset of the gnomAD (N = 44,779^55^). PTVs were defined by being annotated as frame-shift, splice-site or stop-gained and predicted with LOF flag by SnpEff^77^. Severe damaging missense variants (SevereDMVs) were classified based on an MPC score ≥ 3^78^.

We employed logistic regression to assess the impact of rare deleterious variants (rare PTVs+ SevereDMVs) in ADHD-CUD compared to ADHD-only in all autosomal genes. Then we proceeded to perform sub-analyses evaluating the load of rare deleterious variants (rare PTVs+ SevereDMVs) in ADHD-CUD, compared to ADHD-only in bins of genes stratified by their pLI score: (I) in genes with high tolerance to loss of function mutations (0 < pLI < 0.5), (II) in less constrained genes (0.5 < pLI < 0.9), and in (III) evolutionarily constrained genes (pLI > 0.9). In the sub-analyses we corrected for multiple testing using Bonferroni correction and considered a P-value of 0.0167 as significant (0.05 / 3 (number of pLI bin) = 0.0167).

For comparison we also tested for the load of rare PTVs+SevereDMVs in ADHD-CUD compared to controls and in ADHD-only compared to controls. Load of rare synonymous variants in constrained genes (pLI > 0.9) was also tested as a sensitivity analysis as we would expect no difference between groups for this category of variants.

Covariates included in the regression were birth year, sex, the first ten PCs from PCA (performed after excluding non-European samples), total number of variants, number of rare synonymous variants, percentage of exome target covered at a read depth of at least 20, mean read depth at sites within the exome target passing VQSR and sequencing wave (one-hot encoded).

### Polygenic score analyses

A PGS is a score that summarizes an individual’s polygenic load of common variants associated with a phenotype, based on individual level genotypes. PGS analyses were performed using individual level genotype data from individuals in the iPSYCH cohort^18,19^. Individuals with ADHD (ICD10 diagnosis codes: F90.0, F90.1, F98.8) and CUD (ICD10 diagnosis codes: F12.1-9) were identified in the Danish registries^75,76^, controls were randomly selected population-based controls without CUD and ADHD.

Quality control and imputation of genotypes have been described in detail elsewhere (REF^3^). Related individuals were removed using PLINK^79^ identity by states analyses and genetic outliers were identified using Eigensoft^80^ implemented in the Ricopili pipeline^81^. Genetic outliers were identified in principal component analysis (PCA) using best-guess genotypes. The principal component (PC) 1 and PC2 values from the first round of PCA were used to define an ellipsoid with a center based on the mean values of PC1 and PC2 of a subsample. Individuals with PC1 or PC2 values greater than six standard deviations from the center of the ellipsoid were removed as genetic outliers. The sub-sample used to define the ellipsoid consisted of individuals who themselves and their parents had Denmark registered as their birthplace. Two additional PCAs were run to further filter and inspect data for genetic outliers. After non-Europeans and related individuals were removed, the iPSYCH dataset contained 37,246 controls, 25,545 ADHD cases, and 5,490 CUD cases.

PGSs were estimated using summary statistics from large GWAS of phenotypes related to SUD, substance use, psychiatric disorders, and educational attainment (see Supplementary Table 22) using LDpred2-auto^82^ (automatic model). For studies including iPSYCH data in the original GWAS we used results with iPSYCH excluded (MDD, SZ, CUD), or generated the PGS as described below (for ADHD and ASD). The method automatically estimates the sparsity *p* and the h^2^_SNP_, and thus validation data are not required to tune hyper-parameters. We ran 30 Gibbs chains with the h^2^_SNP_ estimate from LDSC as the initial value for h^2^_SNP_ and a sequence of equally spaced and log scaled P-values between 1×10^-4^ and 0.5. A Gibbs chain was kept if the absolute difference between the standard deviation of the predictors and the median standard deviation were within three median absolute deviations. The average effect size of retained models was used to calculate PGSs in the iPSYCH sample. Only variants with minor allele frequency MAF > 0.01 and INFO score > 0.8 (if this information was available) in the summary statistics were used, and analyses were constrained to HapMap3+ variants as recommended in (REF^82^). All generated PGSs were standardized using means and standard deviations estimated based on individuals from the iPSYCH control sub cohort with European ancestry.

Due to lack of an external large ADHD GWAS and ASD GWAS, the ADHD-PGS and ASD-PGS were partly internally derived following the approach previously described^4,74^. In short 50 GWAS were run, where 1/50 of the individuals on turn were excluded from the training GWAS. LDpred2 was run 50 times with these training GWASs where PGSs in each run were estimated for the 1/50 of individuals not included in that specific GWAS. The PGSs were first standardized in each fold by using the mean and standard deviation of the PGSs in that fold. Next the 50 PGS-folds were combined into one sample and standardized using the mean and the standard deviation estimated for sub cohort individuals with European ancestry.

Logistic regression models were run for each of the standardized PGSs to estimate how much variance in the phenotype each PGS explains (ADHD vs controls and ADHD-CUD vs controls) estimated by Nagelkerke R^2^ and McFadden R^2^. The first 10 principal components and a batch variable (iPSYCH1 or iPSYCH2) were included in the models. Multivariate multivariable regression was done to evaluate the load of all PGSs in ADHD and ADHD-CUD benchmarked against a control group without ADHD or CUD in one analysis. We applied the multivariate multivariable regression framework, described in detail in (REF^32^). In short the method simultaneously run one linear regression model per PGS on the phenotypes, while handling potential correlations between PGSs by fitting the variance-covariance matrixes jointly. The first 10 principal components and a batch indicator for iPSYCH1 and iPSYCH2 were included as covariates in the model. We applied Bonferroni correction correcting for 36 pair-wise comparisons of PGS load in the groups.

Cox regression was performed to estimate the relative HRR and the absolute risk of developing CUD among individuals with ADHD stratified by their PGS, using an in-house pipeline incorporating the two R-packages survival and survminer (see ULRs). Besides the PGS, the 10 first PCs and a batch variable (iPSYCH1/iPSYCH2) were included in the cox models. Three models were run (1) including all individuals diagnosed with ADHD (stratified into PGS-quantiles), where the model examined the risk of developing CUD from the age of 10 years to 30 years of age. Four PGSs (ADHD, CUD, smoking and EA) were evaluated in the last models (2) the same model but run separately for females and males (stratified into PGS-tertiles) (3) a model constrained to individuals (stratified into PGS- tertiles) having at least one parent diagnosed with a psychiatric disorder (ADHD, SZ, BP, MDD) or any substance use disorder (i.e., alcohol, opioids, cannabinoids, sedatives or hypnotics, cocaine, stimulants, hallucinogens, tobacco, volatile solvents, or multiple drug use and use of other psychoactive substances corresponding to ICD10 diagnoses F10-F19). Cases of acute intoxication were excluded from the analysis (ICD10 diagnoses F1X.0). For each of the three models the risk of CUD among ADHD cases within each PGS group was calculated relative to the low PGS-group and for model 1 HRRs with a P-value less than P = 0.001 (Bonferroni correction correcting for 36 pair-wise comparisons) were considered significantly different from 1. For model 2 and 3 HRRs with a P-value less than P = 0.004 (Bonferroni correction correcting for 12 pair-wise comparisons) were considered significantly different from 1.

For comparison we estimated the relative and absolute risk of developing CUD in the general Danish population with European ancestry by estimating the risk of CUD in the iPSYCH sub cohort excluding individuals with ADHD. Cox regression was done as explained above with individuals stratified into tertiles. All graphs generated for cox regression was smoothed in R with the geom_smooth() method “loess” in order to obfuscate individual data points.

## URLs

Mendelian Randomization: https://mrcieu.github.io/TwoSampleMR/

M-value calculation: METASOFT/updated software MEGASOFT: https://github.com/DGU-CBLAB/MEGASOFT

BrianSpan: http://www.brainspan.org/static/download

BrainSpan documentation: https://www.brainspan.org/static/download.html

TWAS: https://github.com/hakyimlab/MetaXcan/wiki/Tutorial:-GTEx-v8-MASH-models-integration-with-a-Coronary-Artery-Disease-GWAS

1000 Genome Phase 3: https://data.broadinstitute.org/alkesgroup/LDSCORE/1000G_Phase3_frq.tgz

Survival R-package: https://CRAN.R-project.org/package=survival

Survminer R-package: https://CRAN.R-project.org/package=survminer

CAUSALdb finemapping pipeline: https://github.com/mulinlab/CAUSALdb-finemapping-pip

FUMA v.1.3.7: https://fuma.ctglab.nl/.

S-PrediXcan: https://github.com/hakyimlab/MetaXcan.

LDpred2: https://privefl.github.io/bigsnpr/articles/LDpred2.html.

MiXeR v.1.3: https://github.com/precimed/mixer.

## Author contributions

T.T.N., J.D., were responsible for analysis. Sample and/or data provider and processing was carried out by D.F.L., G.B.W., E.C. J., T.T., M.V.P., T.W., P.B.M., H.S., K.S., D.M.H., A.A., J.G., J.G., A.D, D.D. Writing was the responsibility of D.D. Study design and direction was the responsibility of DD. All authors contributed to critical revision of the paper.

## Supporting information

TWAS_Supplementary _Tables

## Acknowledgements

D.D. is supported by the Novo Nordisk Foundation (NNF20OC0065561 and NNFC for Genomic Mechanisms of Disease - NNF21SA0072102), the Lundbeck Foundation (R344-2020-1060) the European Union’s Horizon 2020 research and innovation programme under grant agreement No. 965381(TIMESPAN). D.F.L. is supported by a Career Development Award CDA-2 from the Veterans Affairs Office of Research and Development (1IK2BX005058-01A2). We acknowledge support from VA Merit Award CX001849-01 (Jo.G.). E.C.J. was supported by NIDA (K01DA051759). The iPSYCH team was supported by grants from the Lundbeck Foundation (R102-A9118, R155-2014-1724, and R248-2017-2003), NIH/NIMH (1U01MH109514-01 and 1R01MH124851-01 to A.D.B.) and the Universities and University Hospitals of Aarhus and Copenhagen. The Danish National Biobank resource was supported by the Novo Nordisk Foundation. High-performance computer capacity for handling and statistical analysis of iPSYCH data on the GenomeDK HPC facility was provided by the Center for Genomics and Personalized Medicine and the Centre for Integrative Sequencing, iSEQ, Aarhus University, Denmark (grant to A.D.B.). A.D.B. was also supported by the EU’s HORIZON-HLTH-2021-STAYHLTH-01programme, project number 101057385: Risk and Resilience in Developmental Diversity and Mental Health (R2D2-MH). 23andMe data was included in the MDD summary statistics used in this study, we would like to thank the research participants and employees of 23andMe, inc. for making this work possible. This publication does not represent the views of the Department of Veteran Affairs or the United States Government.

## Data availability

The summary statics of ADHD-CUD and ADHD-CU are available for public download from https://ipsych.dk/en/research/downloads. All relevant iPSYCH data are available from the authors after approval by the iPSYCH Data Access Committee and can only be accessed on the secured Danish server (GenomeDK, https://genome.au.dk) as the data are protected by Danish legislation. For data access please contact D.D. or A.D.B.

## Code availability

No previously unreported custom computer codes or algorithms were used to generate results. The remaining authors declare no competing interests.

## References

1 Faraone, et al. Attention-deficit/hyperactivity disorder. Nature Reviews Disease Primers 15020 2015

2 Faraone, et al. Genetics of attention deficit hyperactivity disorder. Mol Psychiatry 2018

3 Demontis, et al. Genome-wide analyses of ADHD identify 27 risk loci, refine the genetic architecture and implicate several cognitive domains. Nat Genet 2023

4 Demontis, et al. Discovery of the first genome-wide significant risk loci for attention deficit/hyperactivity disorder. Nat Genet 51 63–75 2019

5 Plana-Ripoll, et al. Exploring Comorbidity Within Mental Disorders Among a Danish National Population. JAMA Psychiatry 76 259–70 2019

6 van Emmerik-van Oortmerssen et al. Prevalence of attention-deficit hyperactivity disorder in substance use disorder patients: a meta-analysis and meta-regression analysis. Drug Alcohol Depend 122 11–9 2012

7 Dalsgaard, et al. Mortality in children, adolescents, and adults with attention deficit hyperactivity disorder: a nationwide cohort study. Lancet 385 2190–6 2015

8 Lee, et al. Prospective association of childhood attention-deficit/hyperactivity disorder (ADHD) and substance use and abuse/dependence: a meta-analytic review. Clin Psychol Rev 31 328–41 2011

9 Sundhedsstyrelsen. Stofmisbrugsbehandling - efterspørgsel og tilgængelighed. Narkotikasituationen i Danmark - delrapport 3. 2022

10 Argyriou, et al. Age and impulsive behavior in drug addiction: A review of past research and future directions. Pharmacol Biochem Behav 164 106–17 2018

11 Kozak, et al. The neurobiology of impulsivity and substance use disorders: implications for treatment. Ann N Y Acad Sci 1451 71–91 2019

12 Calakos, et al. Assessment of transient dopamine responses to smoked cannabis. Drug Alcohol Depend 227 108920 2021

13 Faraone. The pharmacology of amphetamine and methylphenidate: Relevance to the neurobiology of attention-deficit/hyperactivity disorder and other psychiatric comorbidities. Neurosci Biobehav Rev 87 255–70 2018

14 Johnson, et al. A large-scale genome-wide association study meta-analysis of cannabis use disorder. Lancet Psychiatry 2020

15 Demontis, et al. Genome-wide association study implicates CHRNA2 in cannabis use disorder. Nat Neurosci 22 1066–74 2019

16 Abdellaoui, et al. Genomic relationships across psychiatric disorders including substance use disorders. Drug Alcohol Depend 220 108535 2021

17 Kranzler, et al. Genome-wide association study of alcohol consumption and use disorder in 274,424 individuals from multiple populations. Nat Commun 10 1499 2019

18 Pedersen, et al. The iPSYCH2012 case-cohort sample: new directions for unravelling genetic and environmental architectures of severe mental disorders. Mol Psychiatry 2017

19 Bybjerg-Grauholm, et al. The iPSYCH2015 Case-Cohort sample: updated directions for unravelling genetic and environmental architectures of severe mental disorders. medRxiv 2020

20 Levey, et al. Multi-ancestry genome-wide association study of cannabis use disorder yields insight into disease biology and public health implications. Nat Genet 55 2094–103 2023

21 Pasman, et al. GWAS of lifetime cannabis use reveals new risk loci, genetic overlap with psychiatric traits, and a causal influence of schizophrenia. Nat Neurosci 21 1161–70 2018

22 Grotzinger, et al. Genomic structural equation modelling provides insights into the multivariate genetic architecture of complex traits. Nat Hum Behav 2019

23 Frei, et al. Bivariate causal mixture model quantifies polygenic overlap between complex traits beyond genetic correlation. Nat Commun 10 2417 2019

24 Bhattacharjee, et al. A subset-based approach improves power and interpretation for the combined analysis of genetic association studies of heterogeneous traits. Am J Hum Genet 90 821–35 2012

25 de Leeuw, et al. MAGMA: generalized gene-set analysis of GWAS data. PLoS Comput Biol 11 e1004219 2015

26 GTEx Consortium. Human genomics. The Genotype-Tissue Expression (GTEx) pilot analysis: multitissue gene regulation in humans. Science 348 648–60 2015

27 Wang, et al. CAUSALdb: a database for disease/trait causal variants identified using summary statistics of genome-wide association studies. Nucleic Acids Res 48 D807–D16 2020

28 Watanabe, et al. Functional mapping and annotation of genetic associations with FUMA. Nat Commun 8 1826 2017

29 Barbeira, et al. Exploring the phenotypic consequences of tissue specific gene expression variation inferred from GWAS summary statistics. Nat Commun 9 1825 2018

30 Barbeira, et al. Integrating predicted transcriptome from multiple tissues improves association detection. PLoS Genet 15 e1007889 2019

31 Trubetskoy, et al. Mapping genomic loci implicates genes and synaptic biology in schizophrenia. Nature 604 502–8 2022

32 Grove, et al. Identification of common genetic risk variants for autism spectrum disorder. Nat Genet 51 431–44 2019

33 Als et al. Identification of 64 new risk loci for major depression, refinement of the genetic architecture and risk prediction of recurrence and comorbidities. medRxiv 2022

34 Zhou, et al. Genome-wide meta-analysis of problematic alcohol use in 435,563 individuals yields insights into biology and relationships with other traits. Nat Neurosci 2020

35 Kember, et al. Cross-ancestry meta-analysis of opioid use disorder uncovers novel loci with predominant effects in brain regions associated with addiction. Nat Neurosci 25 1279–87 2022

36 Okbay, et al. Polygenic prediction of educational attainment within and between families from genome-wide association analyses in 3 million individuals. Nat Genet 54 437–49 2022

37 Liu, et al. Association studies of up to 1.2 million individuals yield new insights into the genetic etiology of tobacco and alcohol use. Nat Genet 2019

38 Jansen, et al. Genome-wide analysis of insomnia in 1,331,010 individuals identifies new risk loci and functional pathways. Nat Genet 51 394–403 2019

39 Mills, et al. Identification of 371 genetic variants for age at first sex and birth linked to externalising behaviour. Nat Hum Behav 5 1717-30 2021

40 Bulik-Sullivan, et al. LD Score regression distinguishes confounding from polygenicity in genome-wide association studies. Nat Genet 47 291–5 2015

41 Lek, et al. Analysis of protein-coding genetic variation in 60,706 humans. Nature 536 285–91 2016

42 Treur, et al. Investigating causality between liability to ADHD and substance use, and liability to substance use and ADHD risk, using Mendelian randomization. Addict Biol 26 e12849 2021

43 Soler Artigas et al. Attention-deficit/hyperactivity disorder and lifetime cannabis use: genetic overlap and causality. Mol Psychiatry 2019

44 Arends, et al. Associations between the CADM2 gene, substance use, risky sexual behavior, and self-control: A phenome-wide association study. Addict Biol 26 e13015 2021

45 Sanchez-Roige, et al. CADM2 is implicated in impulsive personality and numerous other traits by genome- and phenome-wide association studies in humans and mice. Transl Psychiatry 13 167 2023

46 Yang, et al. Association of haplotypic variants in DRD2, ANKK1, TTC12 and NCAM1 to alcohol dependence in independent case control and family samples. Hum Mol Genet 16 2844–53 2007

47 Bloomfield, et al. The effects of Delta9-tetrahydrocannabinol on the dopamine system. Nature 539 369–77 2016

48 Klein, et al. Dopamine: Functions, Signaling, and Association with Neurological Diseases. Cell Mol Neurobiol 39 31–59 2019

49 Carta, et al. Cerebellar modulation of the reward circuitry and social behavior. Science 363 2019

50 Wagner, et al. Cerebellar granule cells encode the expectation of reward. Nature 544 96–100 2017

51 Sanchez-Roige, et al. Genome-Wide Association Studies of Impulsive Personality Traits (BIS-11 and UPPS-P) and Drug Experimentation in up to 22,861 Adult Research Participants Identify Loci in the CACNA1I and CADM2 genes. J Neurosci 39 2562-72 2019

52 Karlsson Linner et al. Genome-wide association analyses of risk tolerance and risky behaviors in over 1 million individuals identify hundreds of loci and shared genetic influences. Nat Genet 51 245–57 2019

53 Manthey, et al. Public health monitoring of cannabis use in Europe: prevalence of use, cannabis potency, and treatment rates. Lancet Reg Health Eur 10 100227 2021

54 Satterstrom, et al. Autism spectrum disorder and attention deficit hyperactivity disorder have a similar burden of rare protein-truncating variants. Nat Neurosci 22 1961–5 2019

55 Karczewski, et al. The mutational constraint spectrum quantified from variation in 141,456 humans. Nature 581 434–43 2020

56 Carney, et al. Effectiveness of early interventions for substance-using adolescents: findings from a systematic review and meta-analysis. Subst Abuse Treat Prev Policy 7 25 2012

57 Polygenic Risk Score Task Force of the International Common Disease. Responsible use of polygenic risk scores in the clinic: potential benefits, risks and gaps. Nat Med 27 1876–84 2021

58 Murray, et al. Could Polygenic Risk Scores Be Useful in Psychiatry?: A Review. JAMA Psychiatry 78 210–9 2021

59 Hjorthoj, et al. Association between cannabis use disorder and schizophrenia stronger in young males than in females. Psychol Med 1–7 2023

60 Zhu, et al. Causal associations between risk factors and common diseases inferred from GWAS summary data. Nat Commun 9 224 2018

61 Yang, et al. GCTA: a tool for genome-wide complex trait analysis. Am J Hum Genet 88 76–82 2011

62 Burgess, et al. Bias due to participant overlap in two-sample Mendelian randomization. Genet Epidemiol 40 597–608 2016

63 Yang, et al. Conditional and joint multiple-SNP analysis of GWAS summary statistics identifies additional variants influencing complex traits. Nat Genet 44 369–75, S1-3 2012

64 Han, et al. Interpreting meta-analyses of genome-wide association studies. PLoS Genet 8 e1002555 2012

65 Han, et al. Random-effects model aimed at discovering associations in meta-analysis of genome-wide association studies. Am J Hum Genet 88 586–98 2011

66 Staley, et al. PhenoScanner: a database of human genotype-phenotype associations. Bioinformatics 32 3207–9 2016

67 Kamat, et al. PhenoScanner V2: an expanded tool for searching human genotype-phenotype associations. Bioinformatics 35 4851–3 2019

68 Benner, et al. FINEMAP: efficient variable selection using summary data from genome-wide association studies. Bioinformatics 32 1493–501 2016

69 Greenbaum, et al. A Statistical Approach to Fine Mapping for the Identification of Potential Causal Variants Related to Bone Mineral Density. J Bone Miner Res 32 1651–8 2017

70 Chen, et al. Fine Mapping Causal Variants with an Approximate Bayesian Method Using Marginal Test Statistics. Genetics 200 719–36 2015

71 Satterstrom, et al. Large-Scale Exome Sequencing Study Implicates Both Developmental and Functional Changes in the Neurobiology of Autism. Cell 180 568–84 e23 2020

72 Sudmant, et al. An integrated map of structural variation in 2,504 human genomes. Nature 526 75–81 2015

73 Bulik-Sullivan, et al. An atlas of genetic correlations across human diseases and traits. Nat Genet 47 1236–41 2015

74 Rajagopal, et al. Differences in the genetic architecture of common and rare variants in childhood, persistent and late-diagnosed attention-deficit hyperactivity disorder. Nat Genet 54 1117–24 2022

75 Mors, et al. The Danish Psychiatric Central Research Register. Scand J Public Health 39 54–7 2011

76 Lynge, et al. The Danish National Patient Register. Scand J Public Health 39 30–3 2011

77 Cingolani et al. A program for annotating and predicting the effects of single nucleotide polymorphisms, SnpEff: SNPs in the genome of Drosophila melanogaster strain w1118; iso-2; iso-3. Fly (Austin) 6 80–92 2012

78 Samocha et al. Regional missense constraint improves variant deleteriousness prediction. bioRxiv 2017

79 Purcell, et al. PLINK: a tool set for whole-genome association and population-based linkage analyses. Am J Hum Genet 81 559–75 2007

80 Wu, et al. A comparison of association methods correcting for population stratification in case-control studies. Ann Hum Genet 75 418–27 2012

81 Lam, et al. RICOPILI: Rapid Imputation for COnsortias PIpeLIne. Bioinformatics 2019 82

82 Prive et al. LDpred2: better, faster, stronger. Bioinformatics 2020

