## Extended_Data1 for "Disentangling the shared genetics of ADHD, cannabis use disorder and cannabis use and prediction of cannabis use disorder in ADHD"

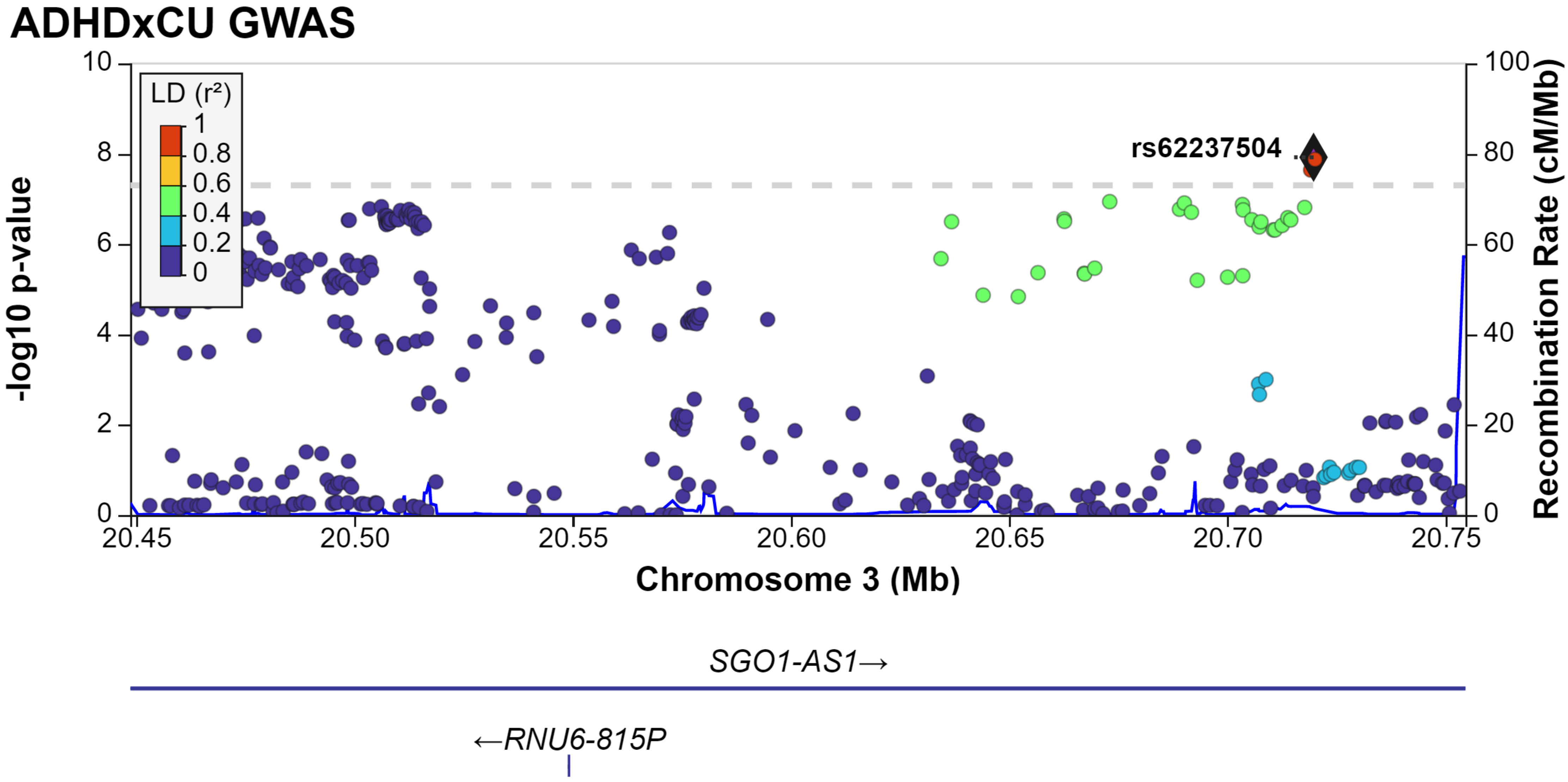

ADHDxCU GWAS
LD (r²)
1
0.8
0.6
0.4
0.2
0
rs62237504
-log10 p-value
Recombination Rate (cM/Mb)
Chromosome 3 (Mb)
20.45
20.50
20.55
20.60
20.65
20.70
20.75
SGO1-AS1→
←RNU6-815P

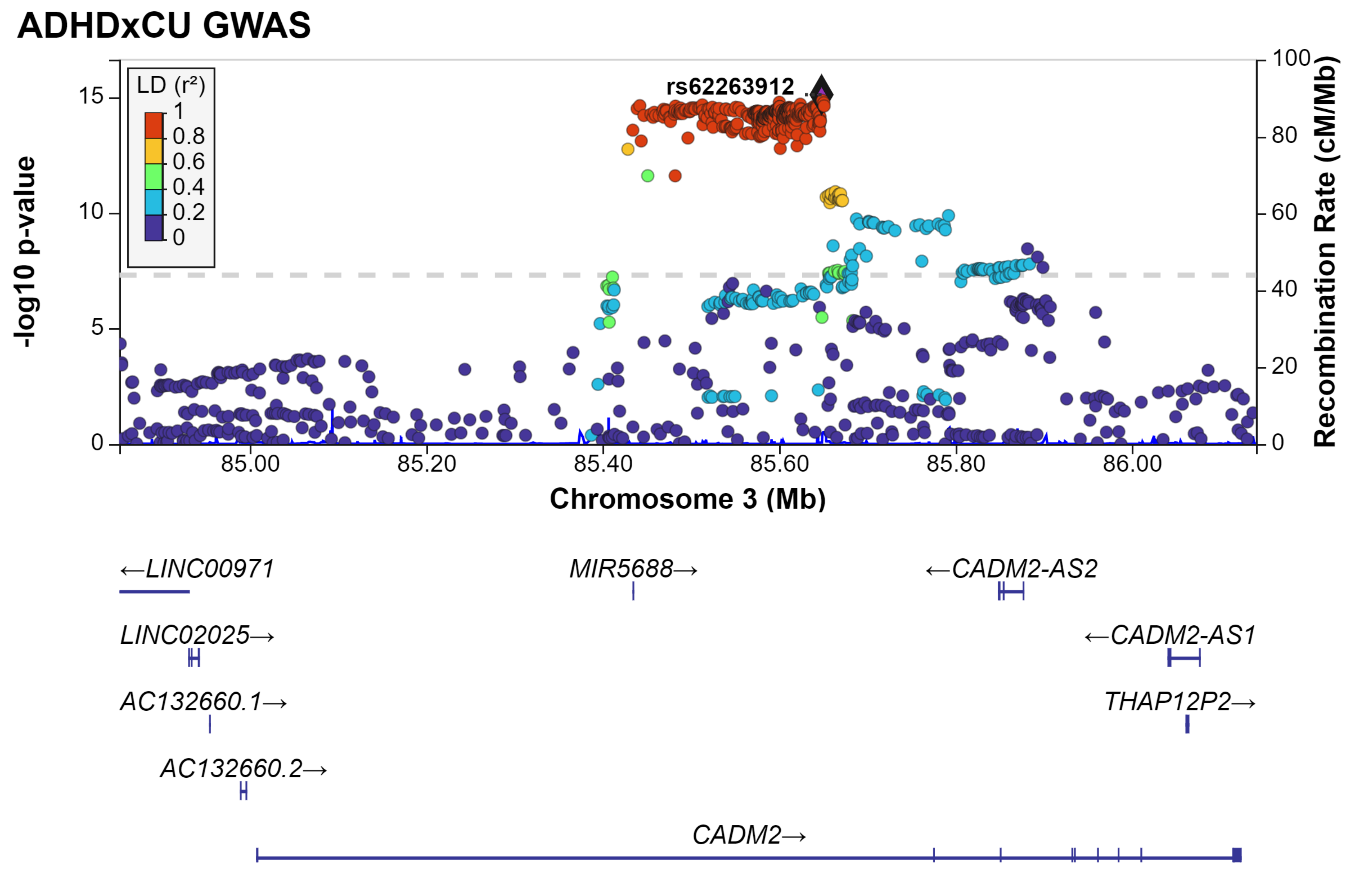
ADHDxCU GWAS
LD (r²)
1
0.8
0.6
0.4
0.2
0
rs62263912
-log10 p-value
15
10
5
0
100
80
60
40
20
0
Recombination Rate (cM/Mb)
85.00
85.20
85.40
85.60
85.80
86.00
Chromosome 3 (Mb)
←LINC00971
MIR5688→
←CADM2-AS2
LINC02025→
←CADM2-AS1
AC132660.1→
THAP12P2→
AC132660.2→
CADM2→

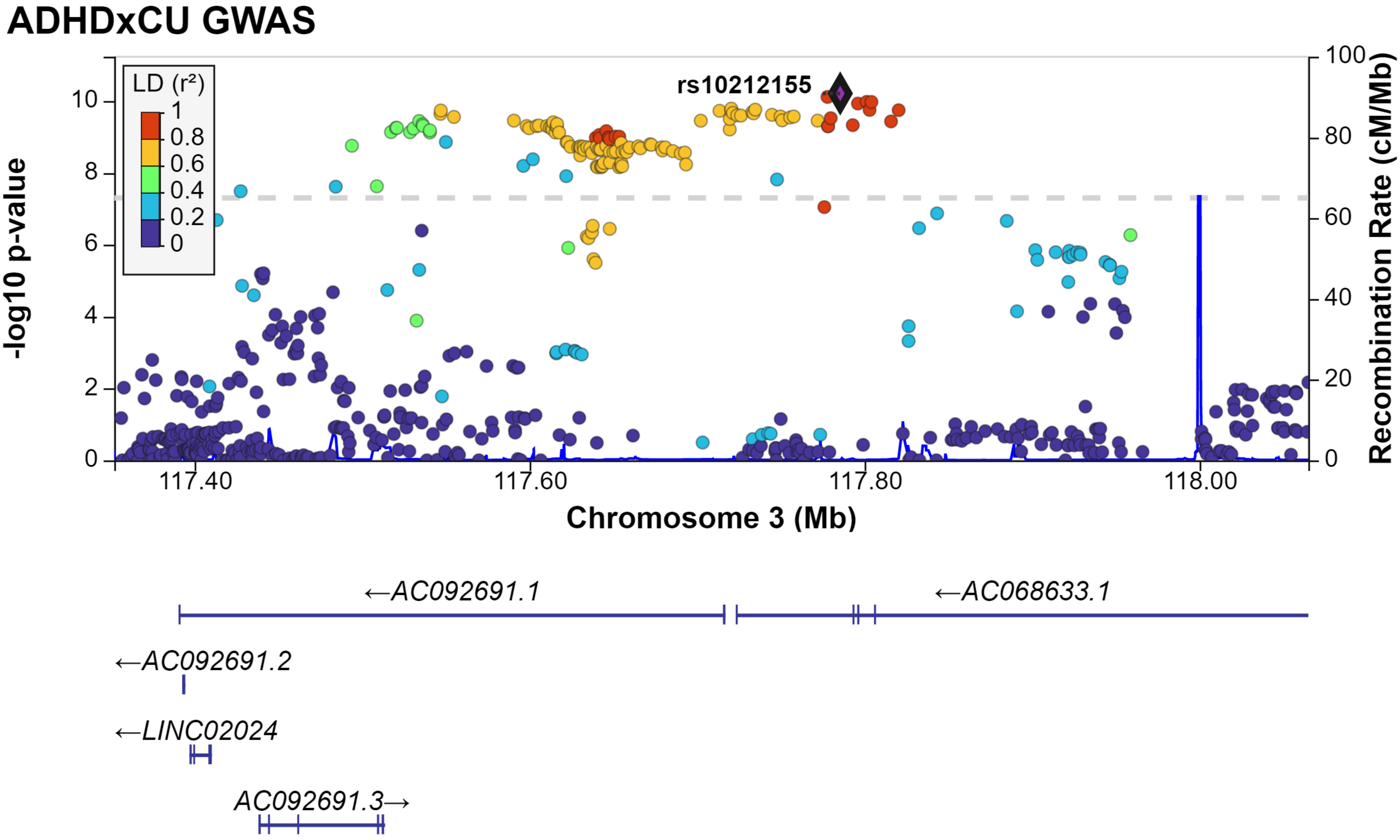
ADHDxCU GWAS
LD (r²)
1
0.8
0.6
0.4
0.2
0
rs10212155
-log10 p-value
Recombination Rate (cM/Mb)
117.40
117.60
117.80
118.00
Chromosome 3 (Mb)
←AC092691.1
←AC068633.1
←AC092691.2
←LINC02024
AC092691.3→

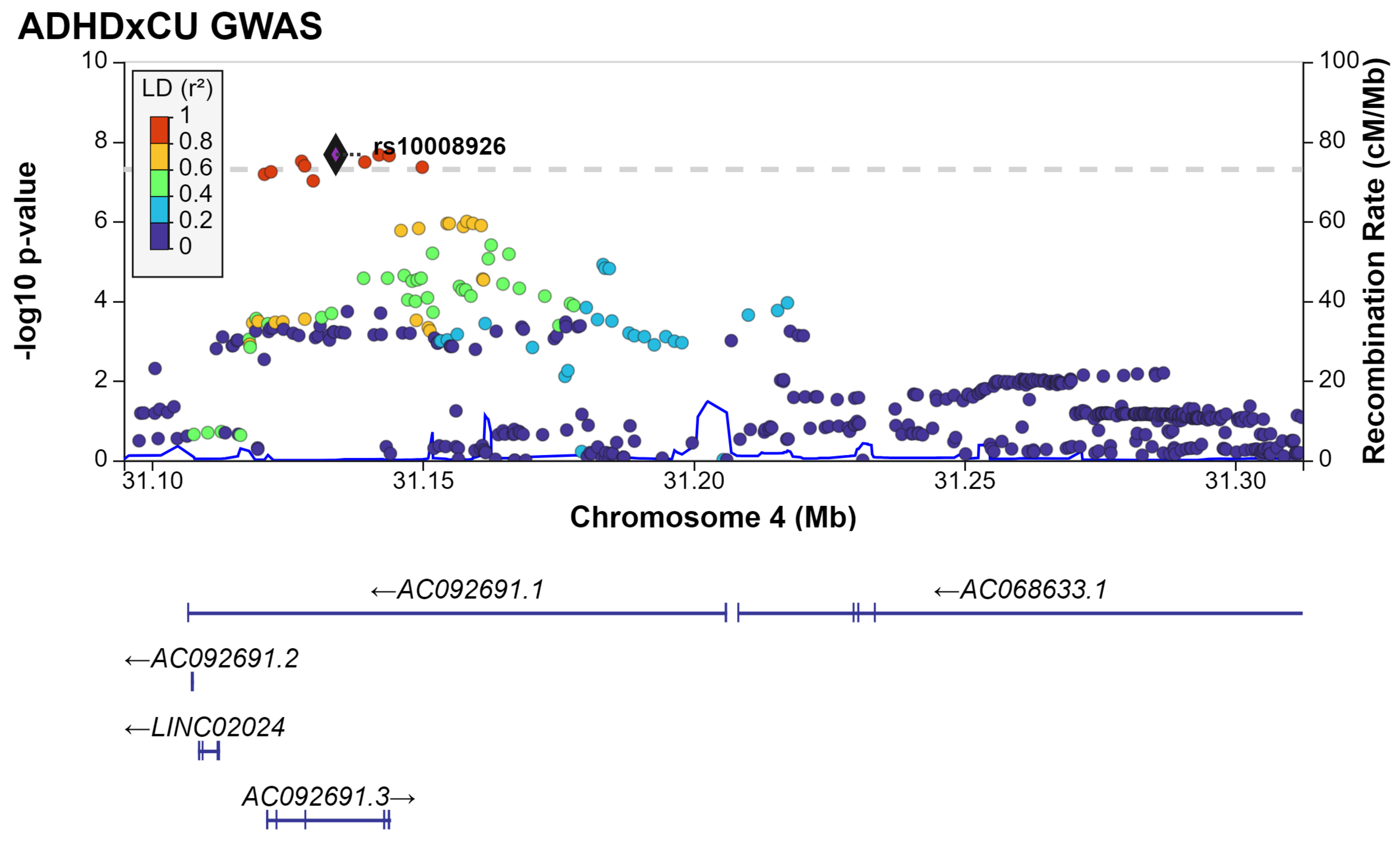
ADHDxCU GWAS
LD (r²)
1
0.8
0.6
0.4
0.2
0
rs10008926
-log10 p-value
Recombination Rate (cM/Mb)
Chromosome 4 (Mb)
31.10
31.15
31.20
31.25
31.30
←AC092691.1
←AC068633.1
←AC092691.2
←LINC02024
AC092691.3→

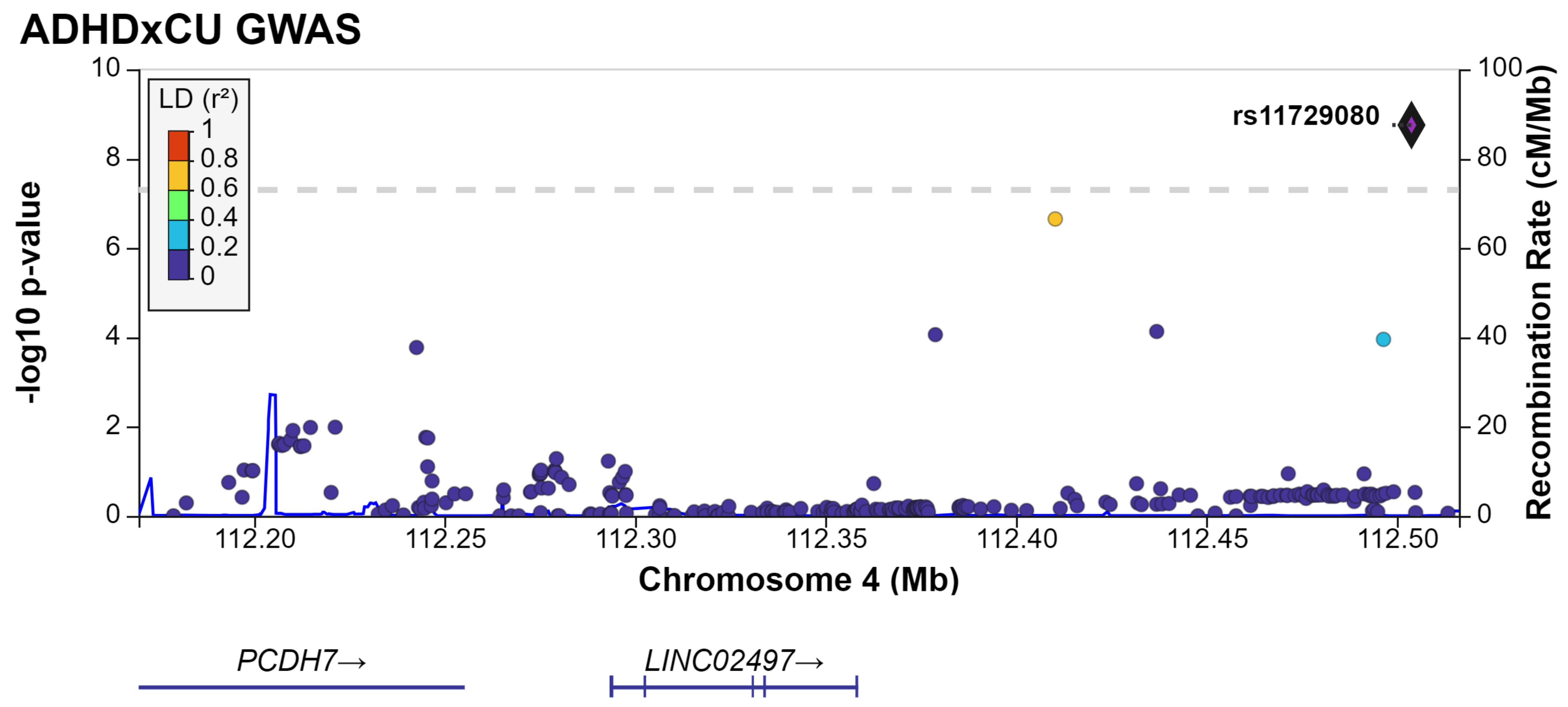
ADHDxCU GWAS
-log10 p-value
Recombination Rate (cM/Mb)
LD (r²)
1
0.8
0.6
0.4
0.2
0
rs11729080
0
2
4
6
8
10
0
20
40
60
80
100
112.20
112.25
112.30
112.35
112.40
112.45
112.50
Chromosome 4 (Mb)
PCDH7→
LINC02497→

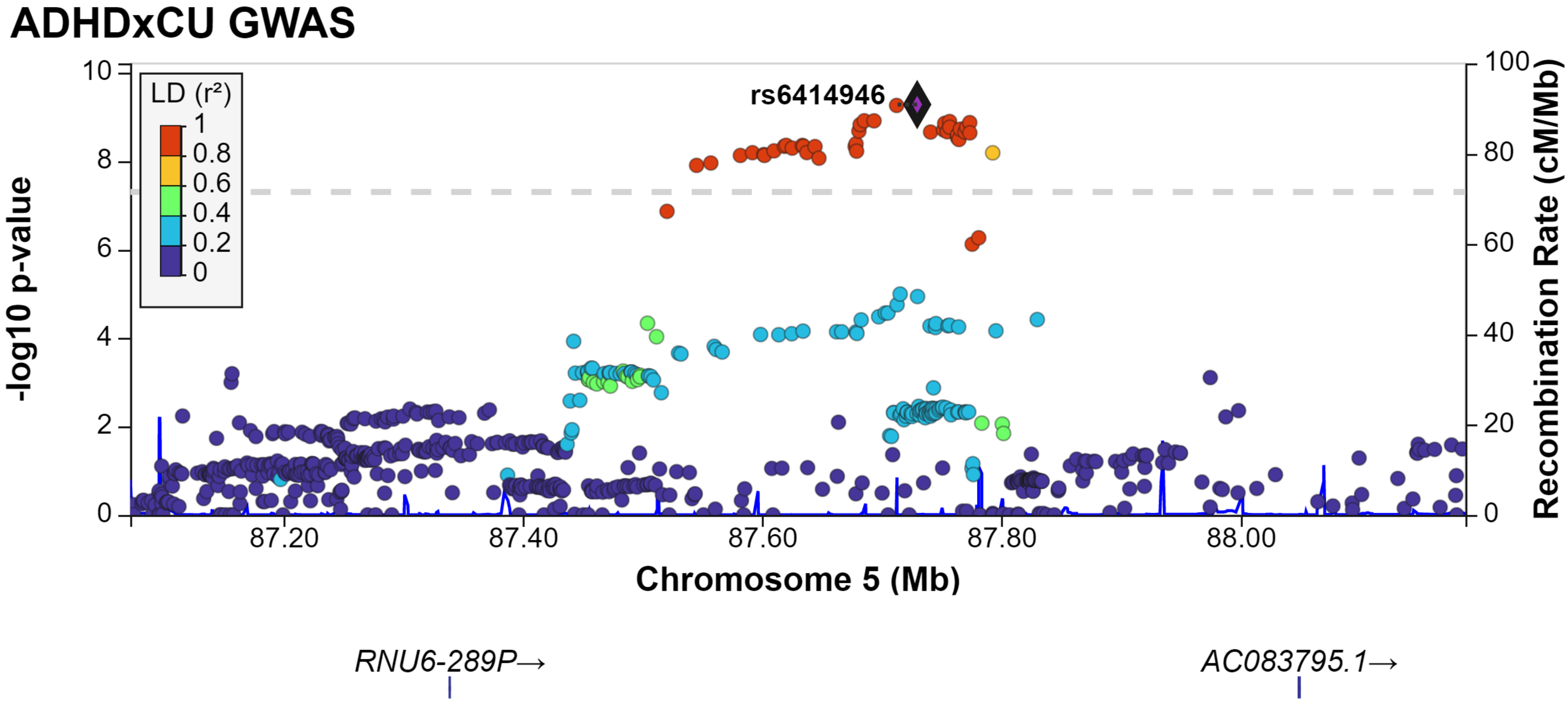

ADHDxCU GWAS
LD (r²)
1
0.8
0.6
0.4
0.2
0
rs6414946
-log10 p-value
Recombination Rate (cM/Mb)
Chromosome 5 (Mb)
87.20
87.40
87.60
87.80
88.00
RNU6-289P→
AC083795.1→

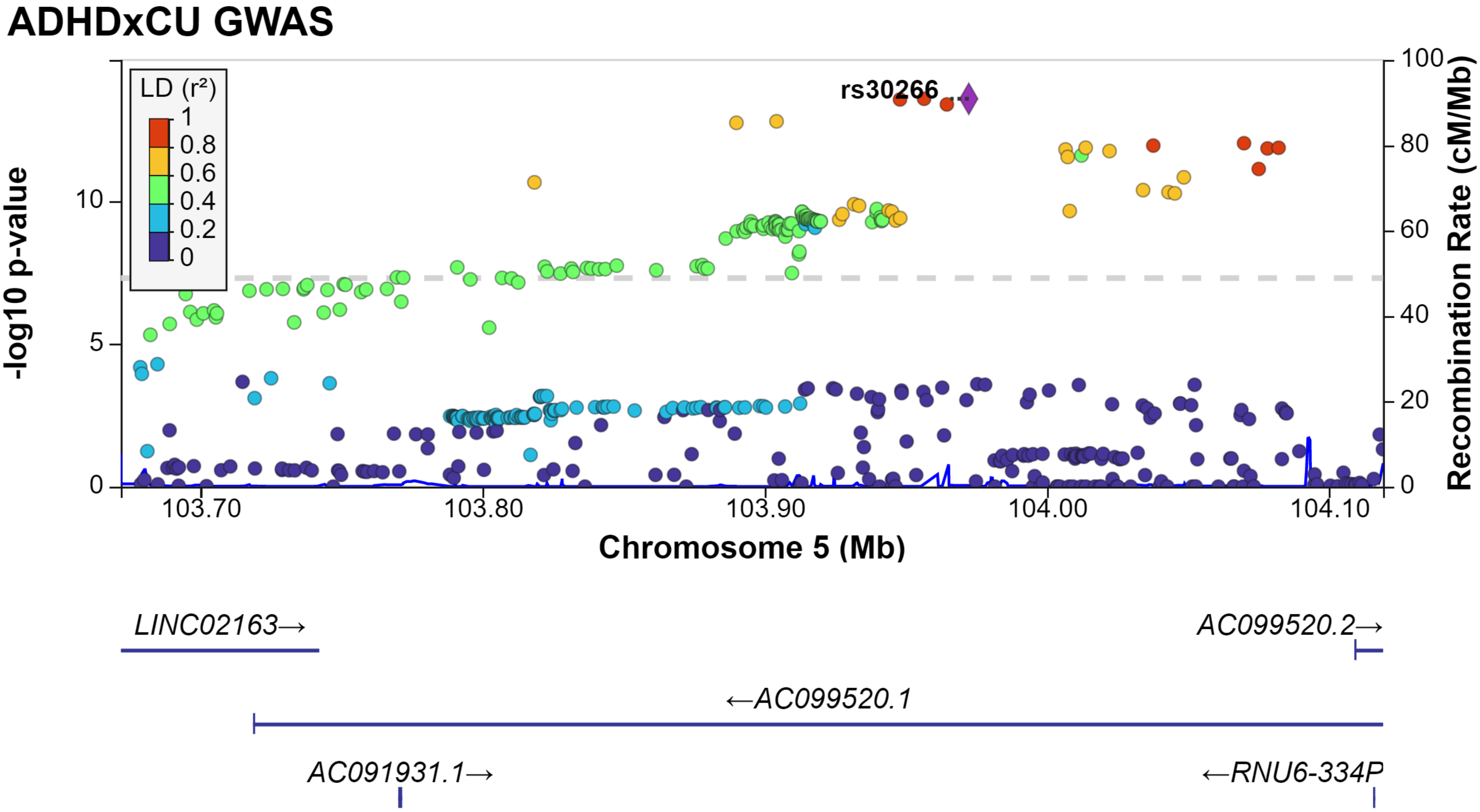
ADHDxCU GWAS
LD (r²)
1
0.8
0.6
0.4
0.2
0
rs30266
-log10 p-value
10
5
0
100
80
60
40
20
0
Recombination Rate (cM/Mb)
103.70
103.80
103.90
104.00
104.10
Chromosome 5 (Mb)
LINC02163→
AC099520.2→
←AC099520.1
AC091931.1→
←RNU6-334P

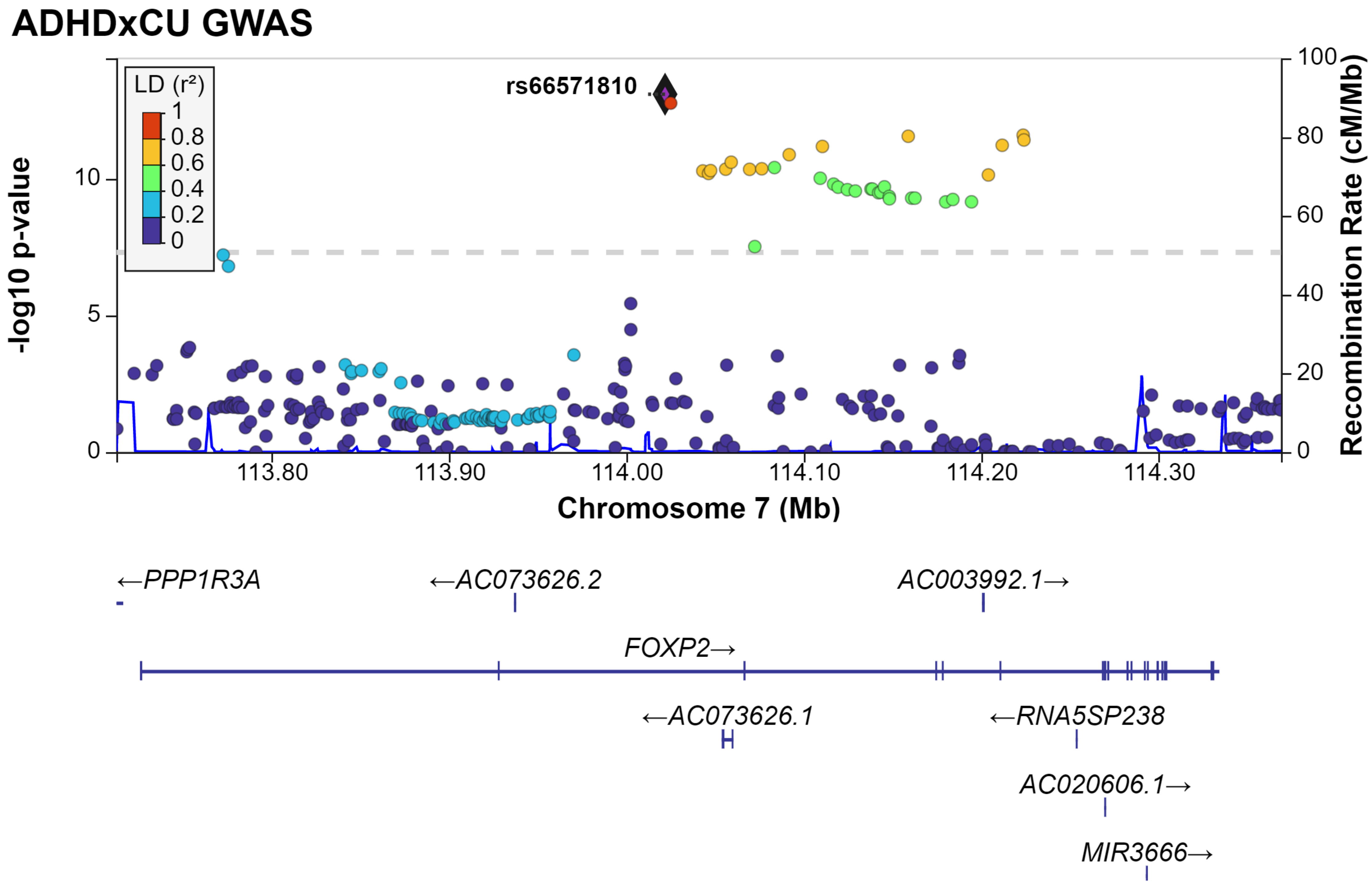

ADHDxCU GWAS
LD (r²)
1
0.8
0.6
0.4
0.2
0
rs66571810
-log10 p-value
Recombination Rate (cM/Mb)
Chromosome 7 (Mb)
113.80
113.90
114.00
114.10
114.20
114.30
←PPP1R3A
←AC073626.2
AC003992.1→
FOXP2→
←AC073626.1
←RNA5SP238
AC020606.1→
MIR3666→

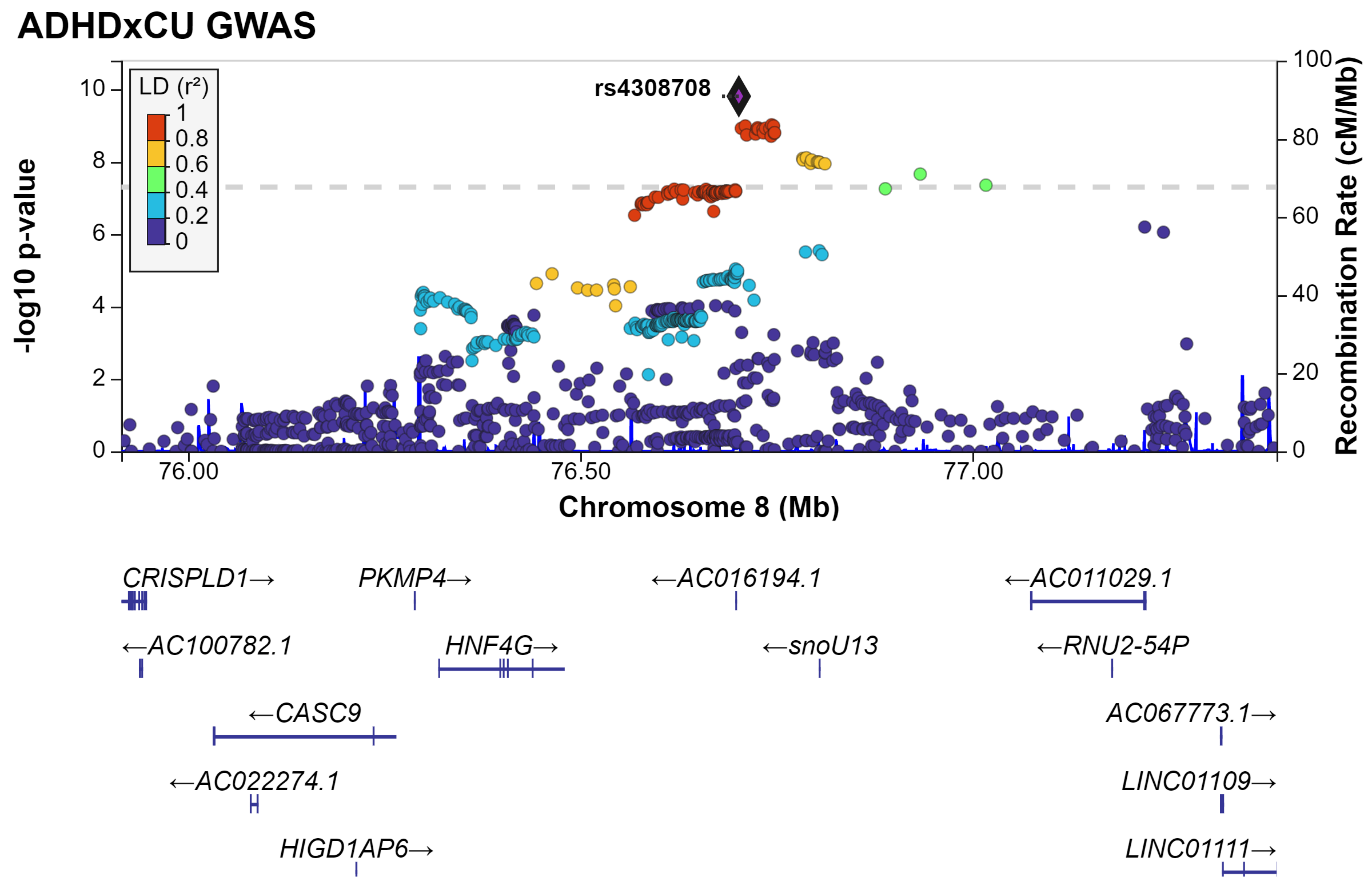
ADHDxCU GWAS
LD (r²)
1
0.8
0.6
0.4
0.2
0
rs4308708
10
8
6
4
2
0
-log10 p-value
100
80
60
40
20
0
Recombination Rate (cM/Mb)
76.00
76.50
77.00
Chromosome 8 (Mb)
CRISPLD1→
PKMP4→
←AC016194.1
←AC011029.1
←AC100782.1
HNF4G→
←snoU13
←RNU2-54P
←CASC9
AC067773.1→
←AC022274.1
LINC01109→
HIGD1AP6→
LINC01111→

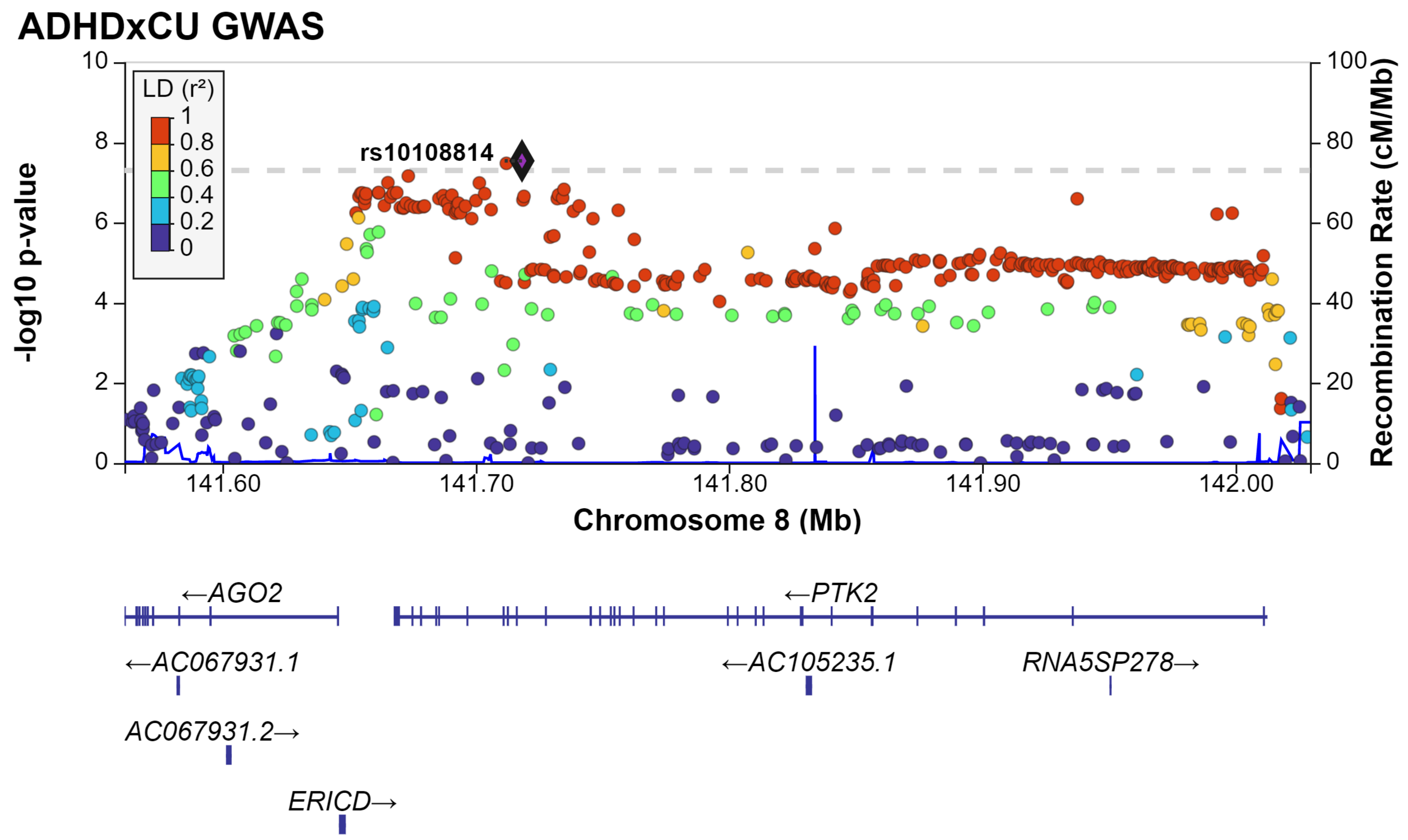

ADHDxCU GWAS
LD (r²)
1
0.8
0.6
0.4
0.2
0
rs10108814
-log10 p-value
Recombination Rate (cM/Mb)
10
8
6
4
2
0
100
80
60
40
20
0
141.60
141.70
141.80
141.90
142.00
Chromosome 8 (Mb)
←AGO2
←PTK2
←AC067931.1
←AC105235.1
RNA5SP278→
AC067931.2→
ERICD→

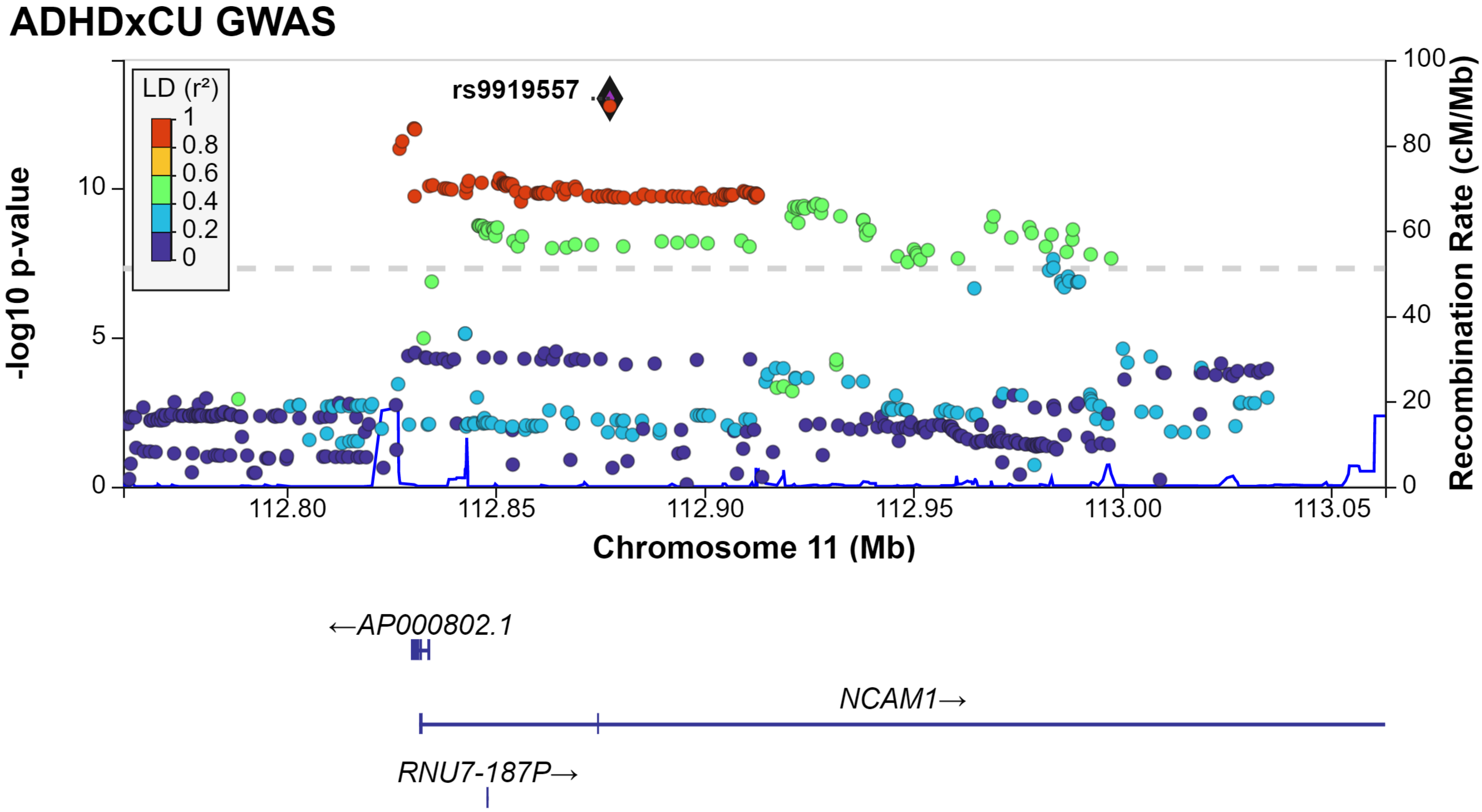
ADHDxCU GWAS
LD (r²)
1
0.8
0.6
0.4
0.2
0
rs9919557
-log10 p-value
10
5
0
100
80
60
40
20
0
Recombination Rate (cM/Mb)
112.80
112.85
112.90
112.95
113.00
113.05
Chromosome 11 (Mb)
←AP000802.1
NCAM1→
RNU7-187P→

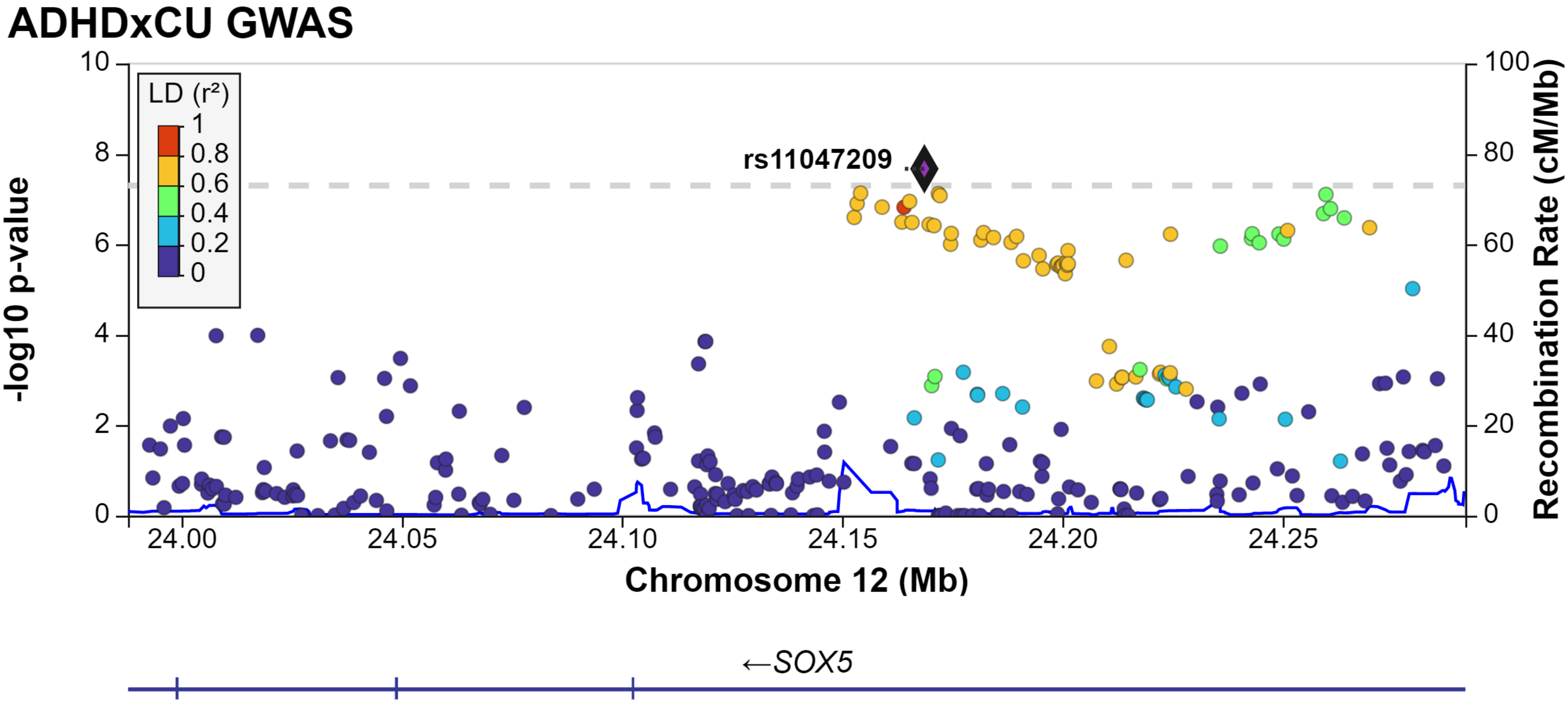
ADHDxCU GWAS
LD (r²)
1
0.8
0.6
0.4
0.2
0
rs11047209
-log10 p-value
Recombination Rate (cM/Mb)
Chromosome 12 (Mb)
24.00
24.05
24.10
24.15
24.20
24.25
←SOX5

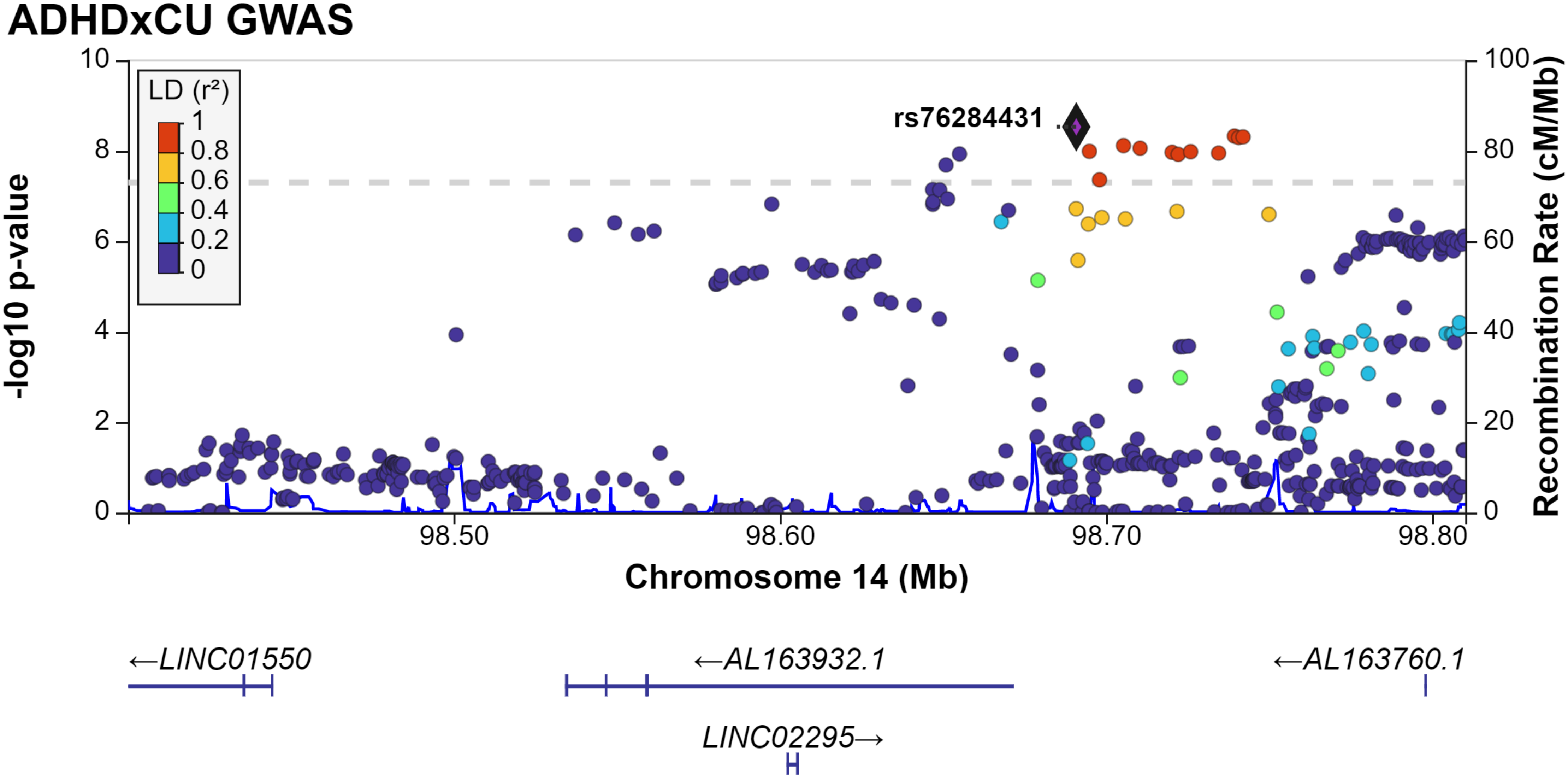

ADHDxCU GWAS
LD (r²)
1
0.8
0.6
0.4
0.2
0
rs76284431
-log10 p-value
Recombination Rate (cM/Mb)
Chromosome 14 (Mb)
98.50
98.60
98.70
98.80
←LINC01550
←AL163932.1
←AL163760.1
LINC02295→

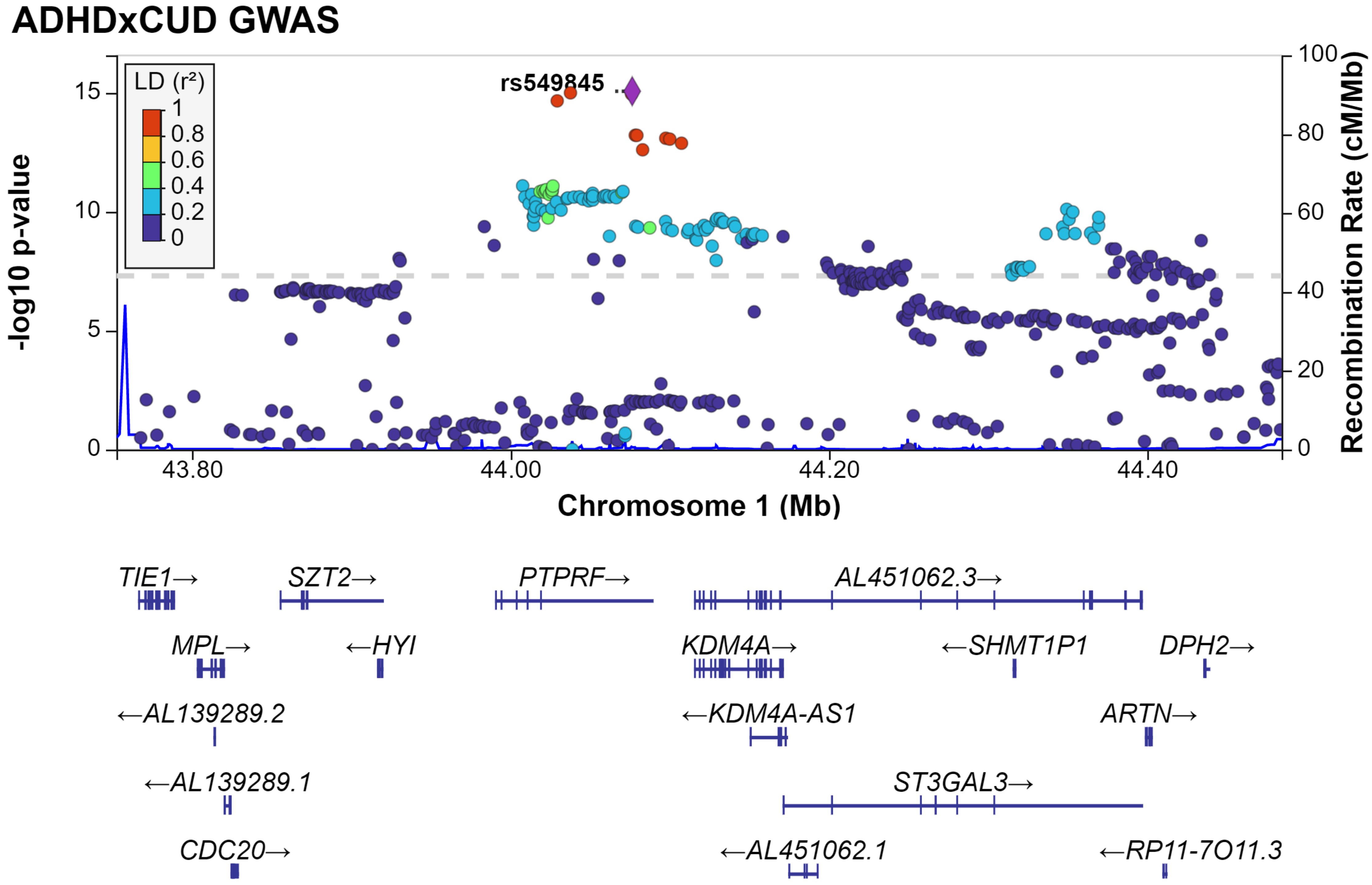

ADHDxCUD GWAS
LD (r²)
1
0.8
0.6
0.4
0.2
0
rs549845
-log10 p-value
Recombination Rate (cM/Mb)
Chromosome 1 (Mb)
43.80
44.00
44.20
44.40
TIE1→
SZT2→
PTPRF→
AL451062.3→
MPL→
←HYI
KDM4A→
←SHMT1P1
DPH2→
←AL139289.2
←KDM4A-AS1
ARTN→
←AL139289.1
ST3GAL3→
CDC20→
←AL451062.1
←RP11-7O11.3

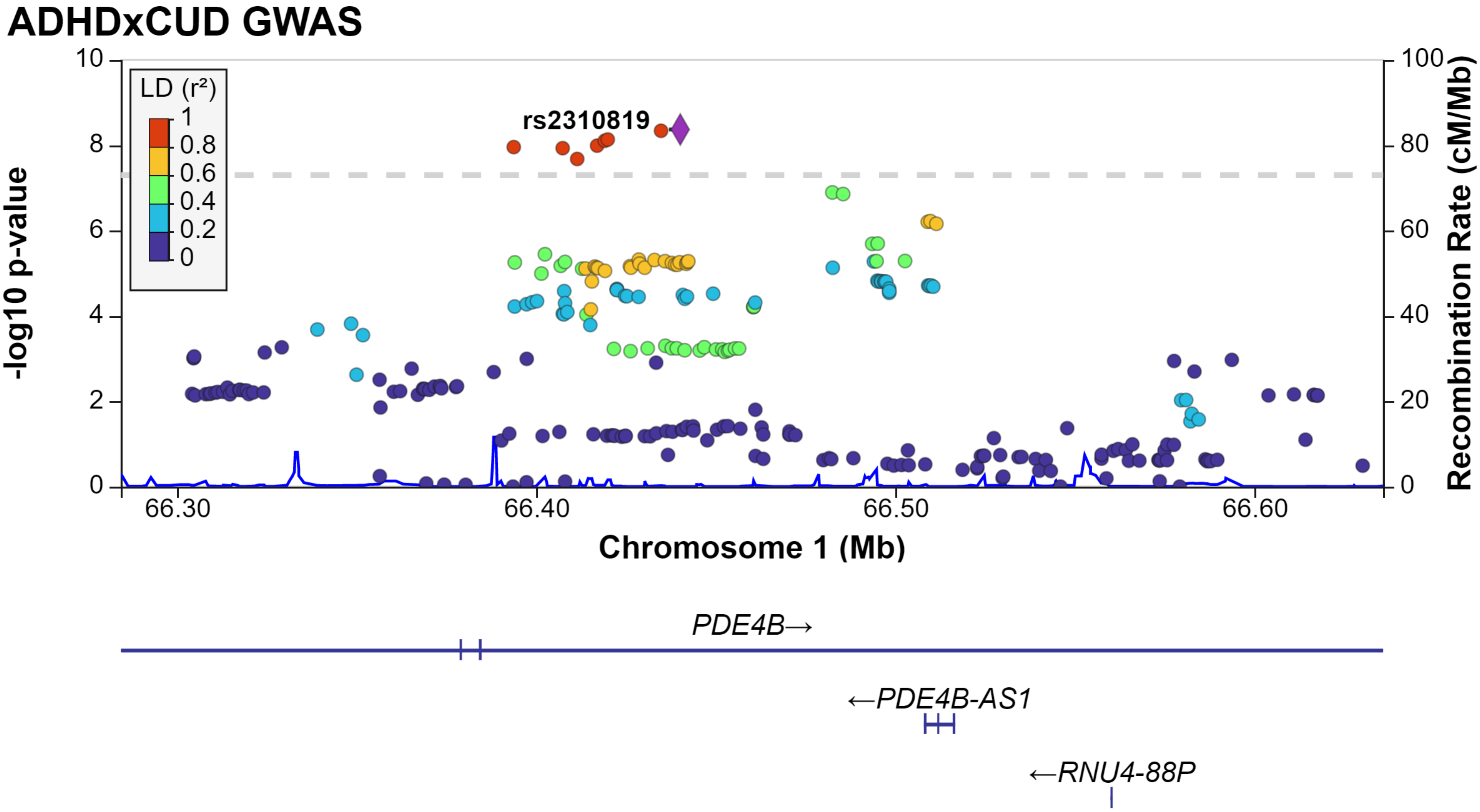
ADHDxCUD GWAS
LD (r²)
1
0.8
0.6
0.4
0.2
0
rs2310819
-log10 p-value
Recombination Rate (cM/Mb)
10
8
6
4
2
0
100
80
60
40
20
0
66.30
66.40
66.50
66.60
Chromosome 1 (Mb)
PDE4B→
←PDE4B-AS1
←RNU4-88P

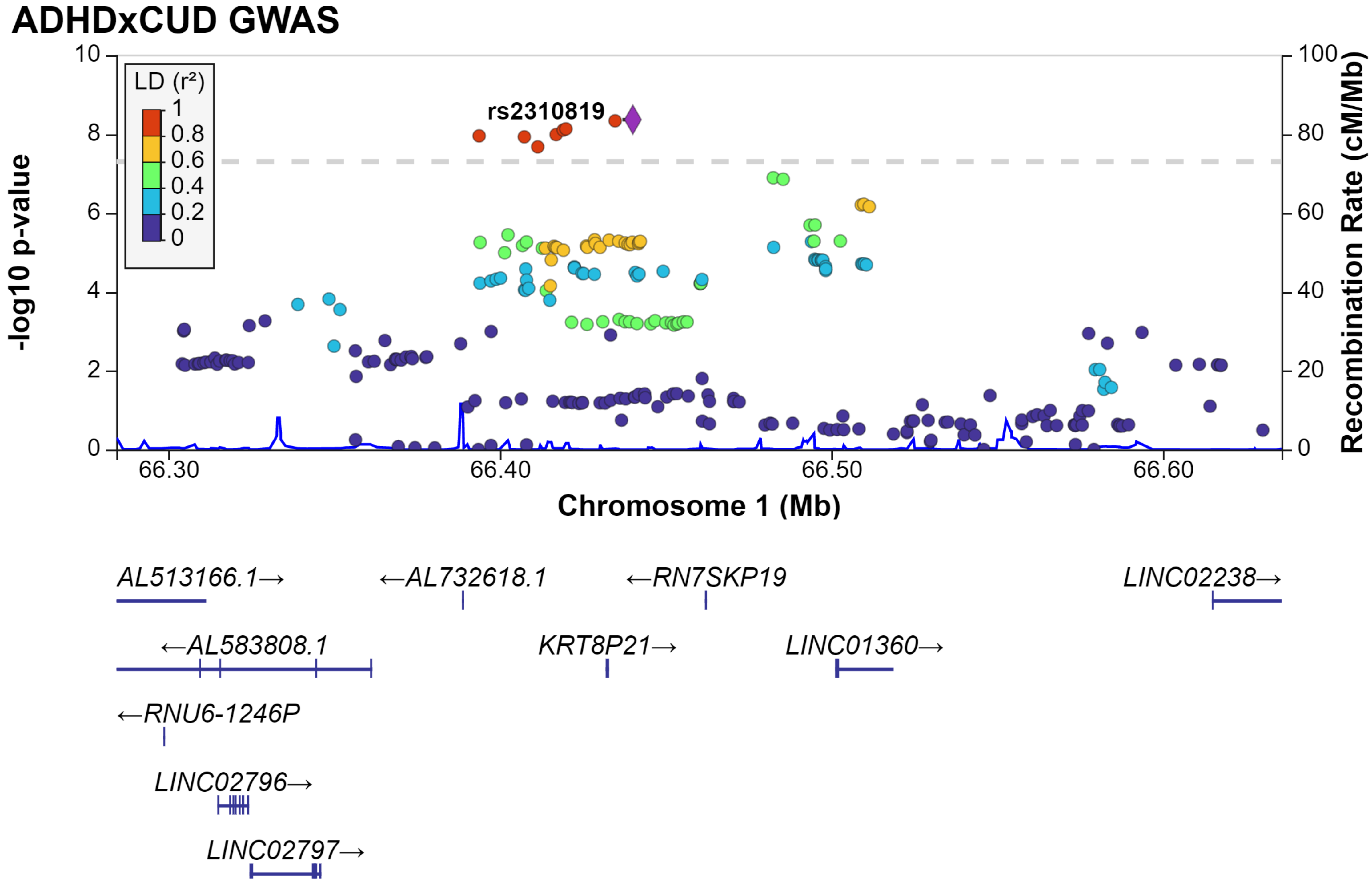
ADHDxCUD GWAS
LD (r²)
1
0.8
0.6
0.4
0.2
0
rs2310819
-log10 p-value
10
8
6
4
2
0
Recombination Rate (cM/Mb)
100
80
60
40
20
0
66.30
66.40
66.50
66.60
Chromosome 1 (Mb)
AL513166.1→
←AL732618.1
←RN7SKP19
LINC02238→
←AL583808.1
KRT8P21→
LINC01360→
←RNU6-1246P
LINC02796→
LINC02797→

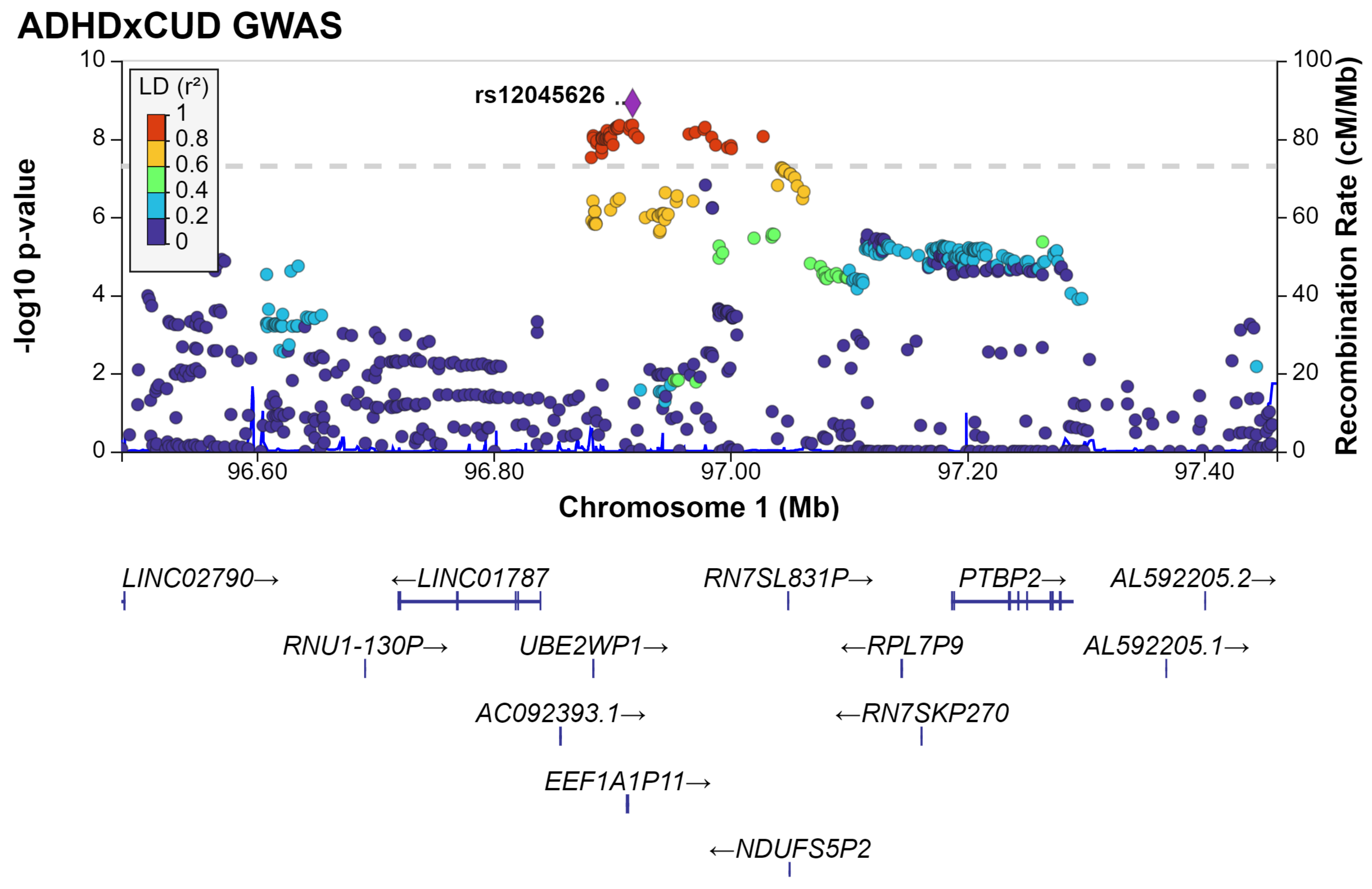

ADHDxCUD GWAS
-log10 p-value
Recombination Rate (cM/Mb)
LD (r²)
1
0.8
0.6
0.4
0.2
0
rs12045626
10
8
6
4
2
0
100
80
60
40
20
0
96.60
96.80
97.00
97.20
97.40
Chromosome 1 (Mb)
LINC02790→
←LINC01787
RN7SL831P→
PTBP2→
AL592205.2→
RNU1-130P→
UBE2WP1→
←RPL7P9
AL592205.1→
AC092393.1→
←RN7SKP270
EEF1A1P11→
←NDUFS5P2

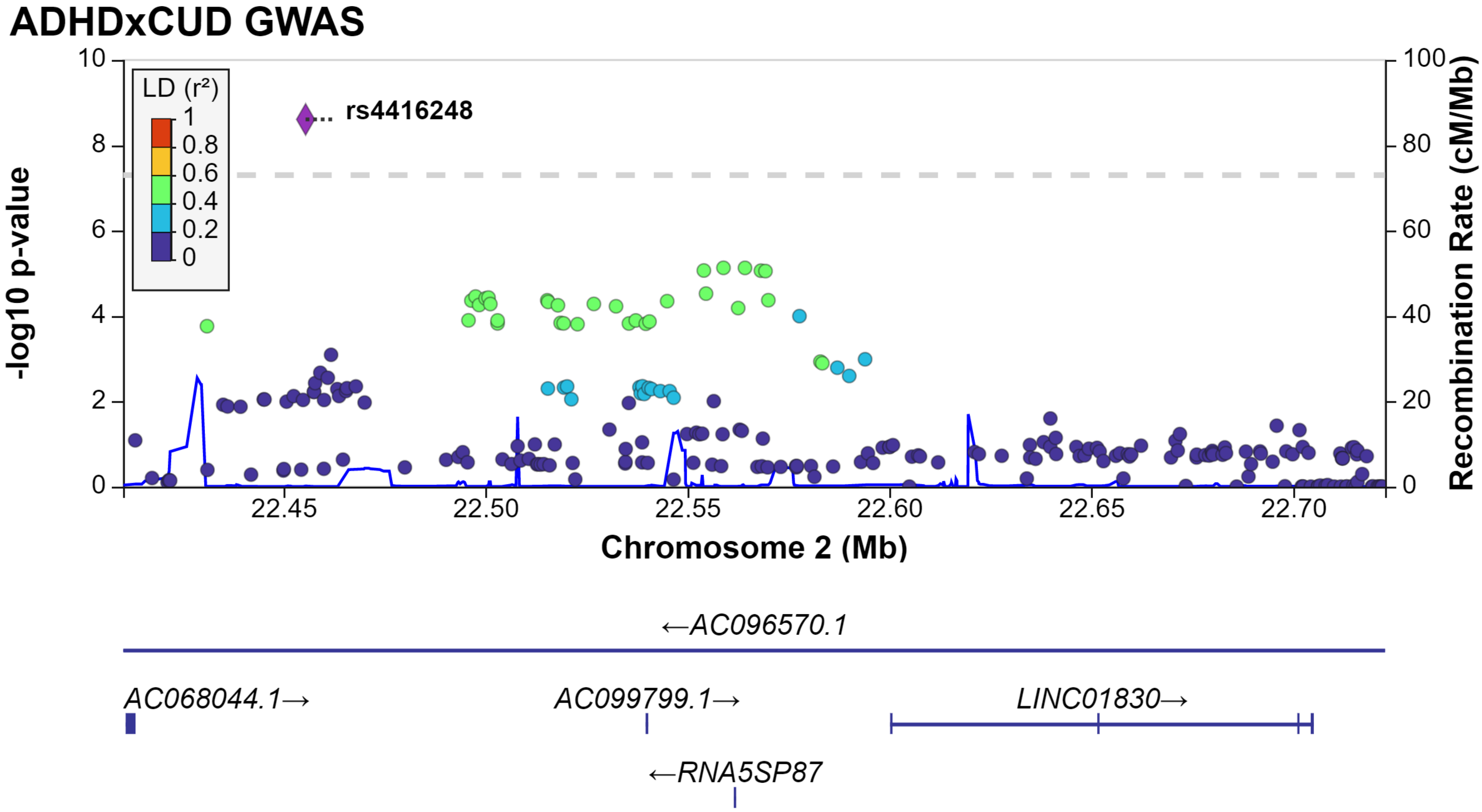

ADHDxCUD GWAS
LD (r²)
1
0.8
0.6
0.4
0.2
0
rs4416248
-log10 p-value
Recombination Rate (cM/Mb)
10
8
6
4
2
0
100
80
60
40
20
0
22.45
22.50
22.55
22.60
22.65
22.70
Chromosome 2 (Mb)
←AC096570.1
AC068044.1→
AC099799.1→
LINC01830→
←RNA5SP87

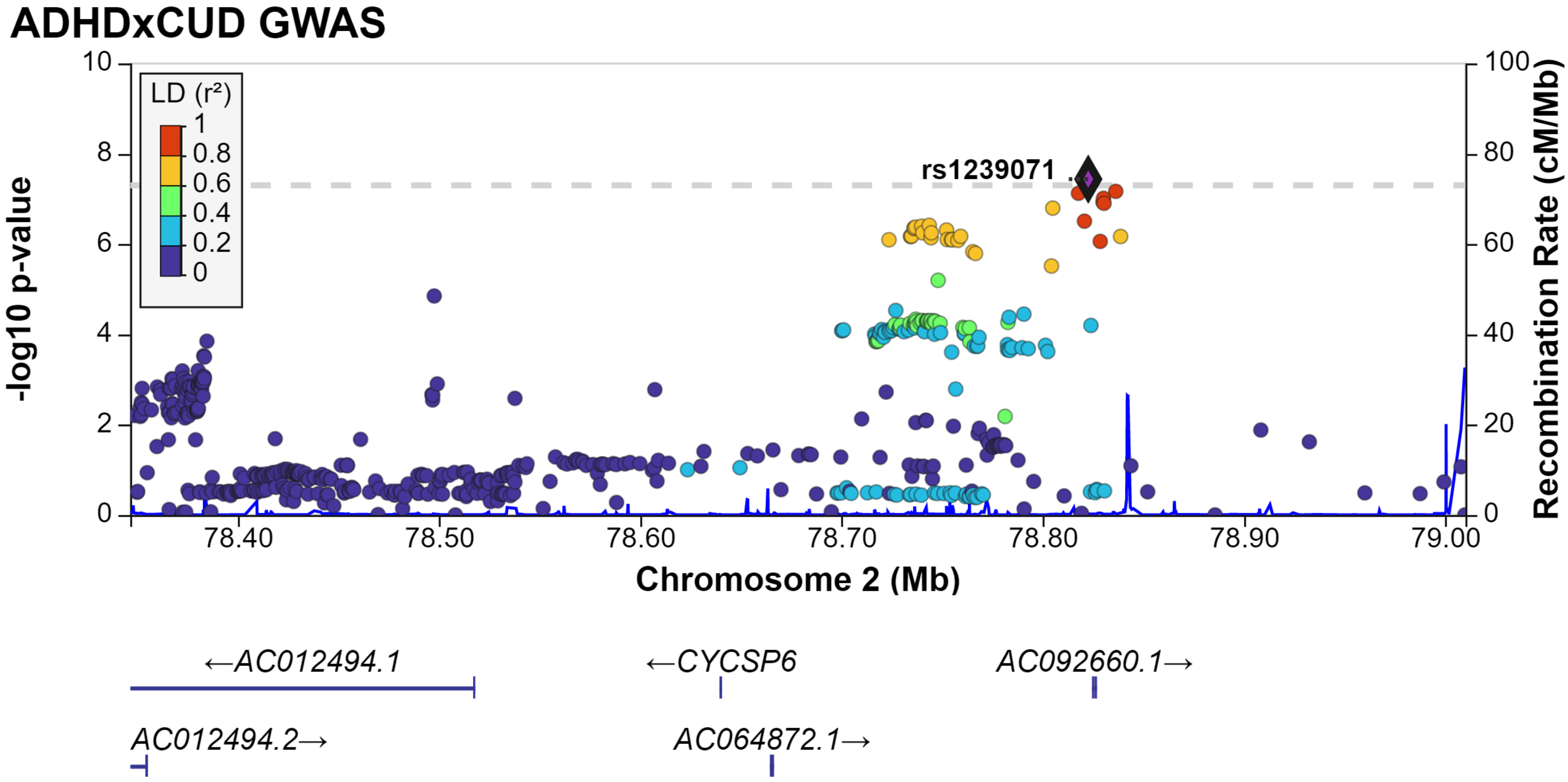
ADHDxCUD GWAS
LD (r²)
1
0.8
0.6
0.4
0.2
0
rs1239071
-log10 p-value
Recombination Rate (cM/Mb)
Chromosome 2 (Mb)
78.40
78.50
78.60
78.70
78.80
78.90
79.00
←AC012494.1
←CYCSP6
AC092660.1→
AC012494.2→
AC064872.1→

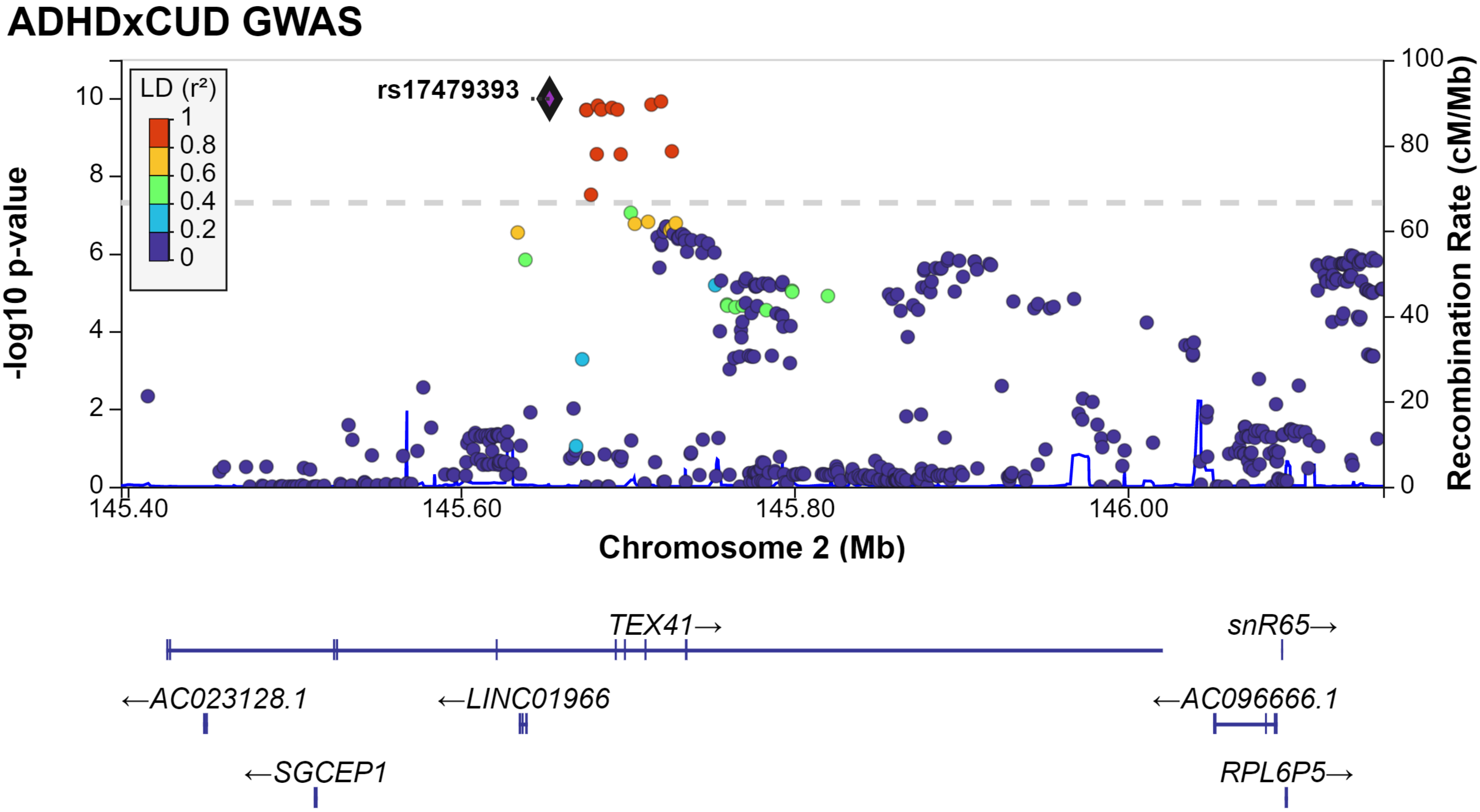

ADHDxCUD GWAS
LD (r²)
1
0.8
0.6
0.4
0.2
0
rs17479393
-log10 p-value
Recombination Rate (cM/Mb)
Chromosome 2 (Mb)
145.40
145.60
145.80
146.00
TEX41→
snR65→
←AC023128.1
←LINC01966
←AC096666.1
←SGCEP1
RPL6P5→

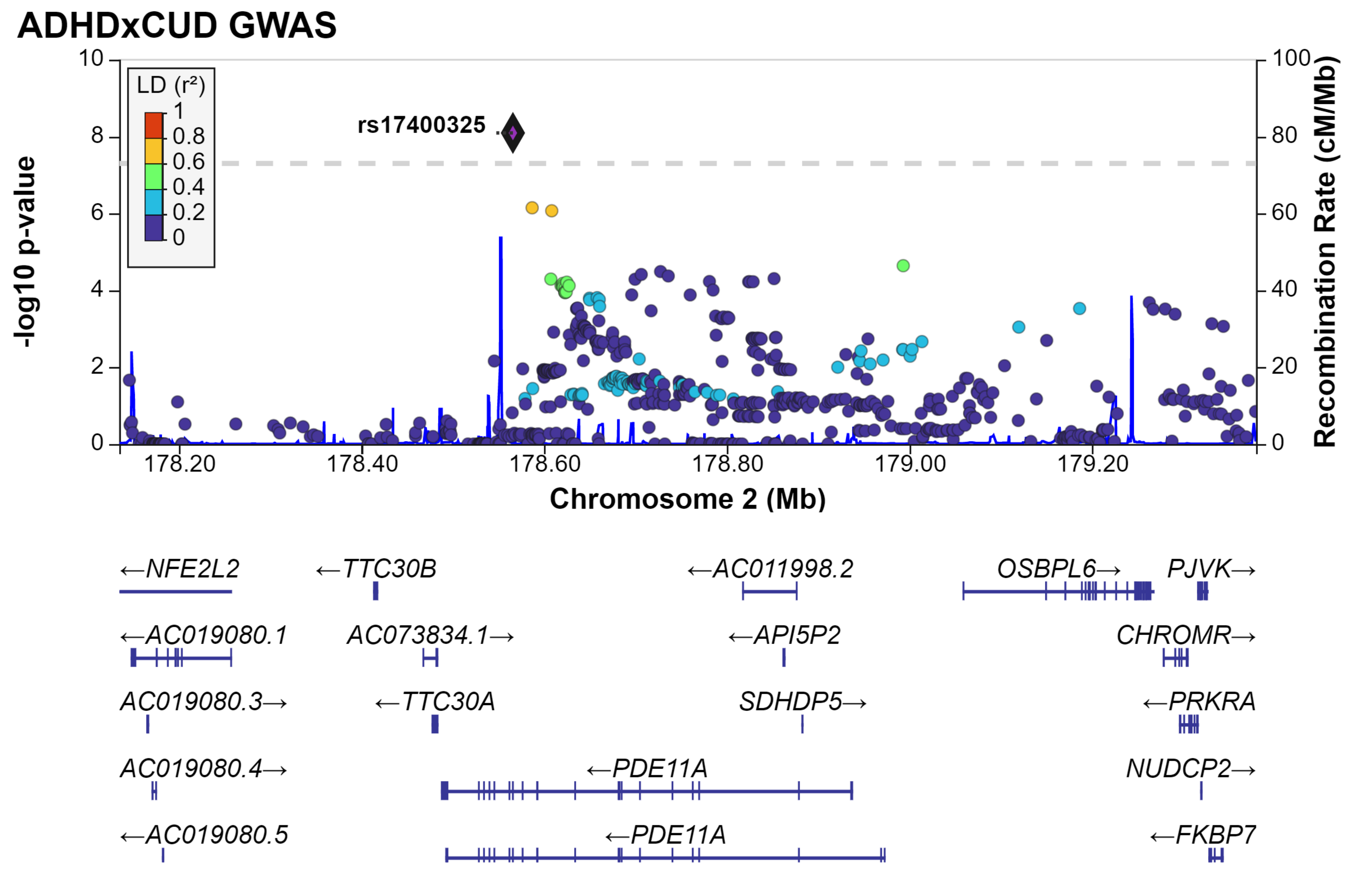

ADHDxCUD GWAS
LD (r²)
1
0.8
0.6
0.4
0.2
0
rs17400325
-log10 p-value
Recombination Rate (cM/Mb)
Chromosome 2 (Mb)
178.20
178.40
178.60
178.80
179.00
179.20
0
2
4
6
8
10
20
40
60
80
100
←NFE2L2
←TTC30B
←AC011998.2
OSBPL6→
PJVK→
←AC019080.1
AC073834.1→
←API5P2
CHROMR→
AC019080.3→
←TTC30A
SDHDP5→
←PRKRA
AC019080.4→
←PDE11A
NUDCP2→
←AC019080.5
←PDE11A
←FKBP7

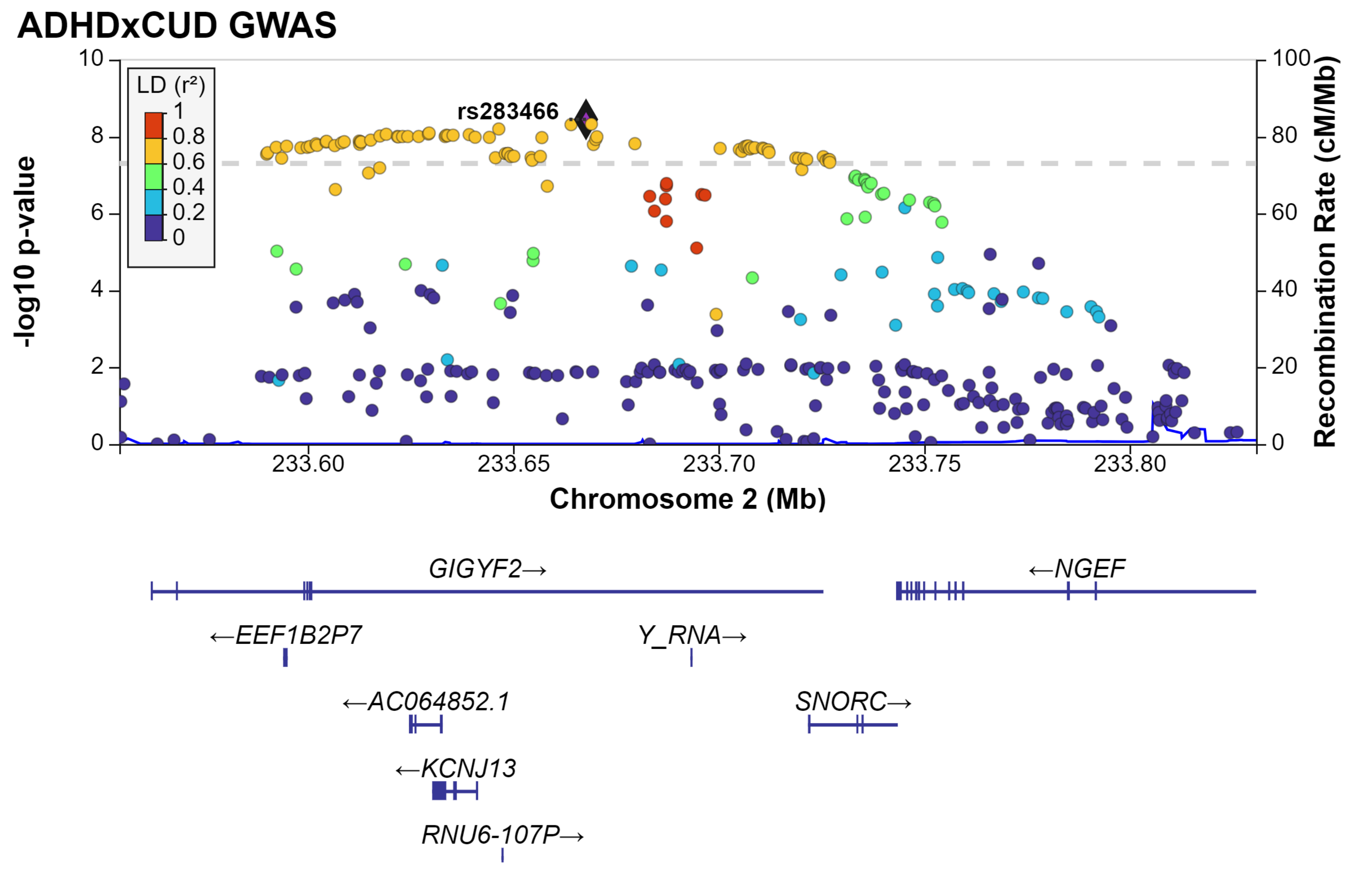

ADHDxCUD GWAS
LD (r²)
1
0.8
0.6
0.4
0.2
0
rs283466
-log10 p-value
Recombination Rate (cM/Mb)
Chromosome 2 (Mb)
233.60
233.65
233.70
233.75
233.80
GIGYF2→
←NGEF
←EEF1B2P7
Y_RNA→
←AC064852.1
SNORC→
←KCNJ13
RNU6-107P→

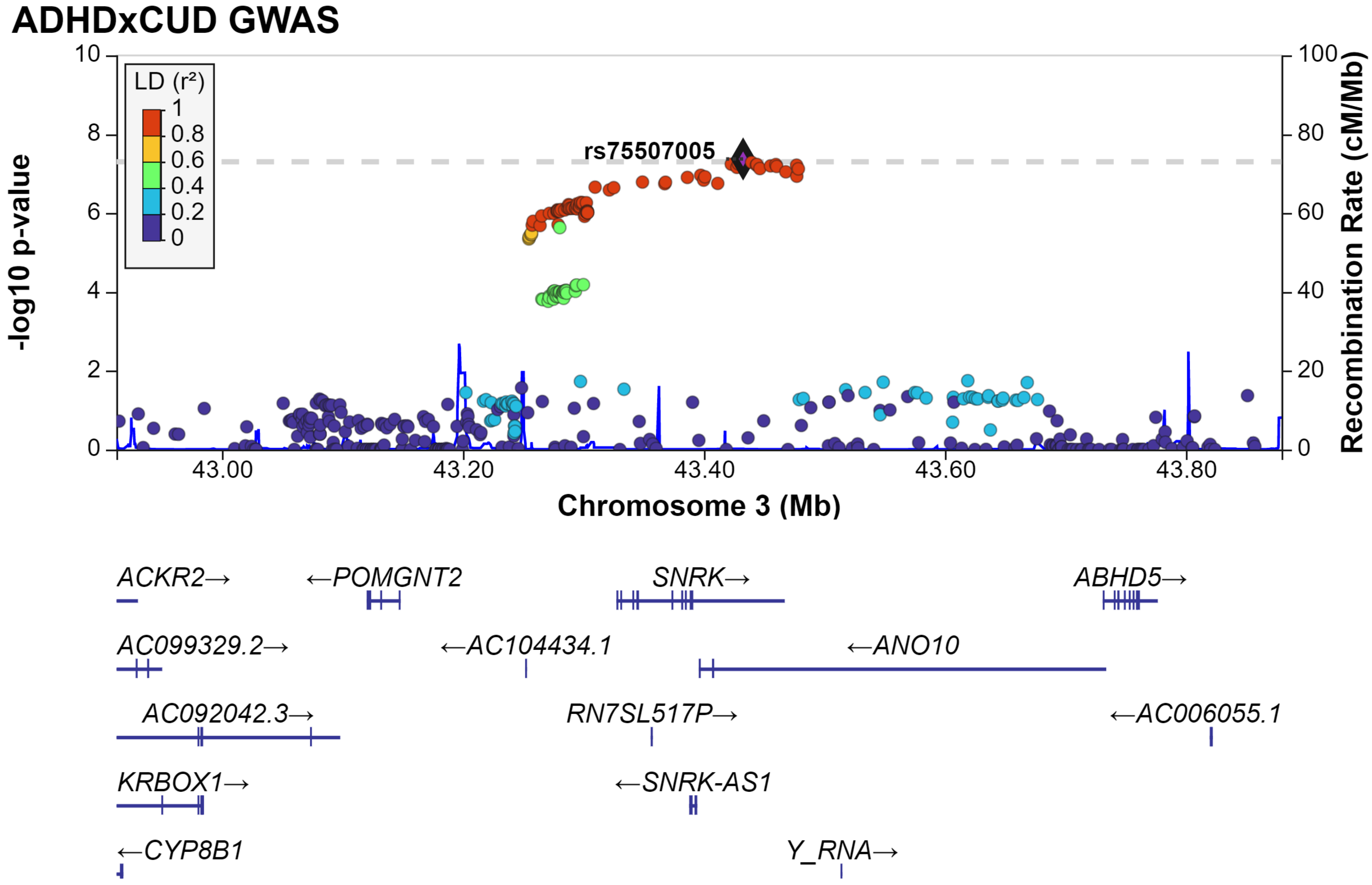

ADHDxCUD GWAS
LD (r²)
1
0.8
0.6
0.4
0.2
0
rs75507005
-log10 p-value
Recombination Rate (cM/Mb)
Chromosome 3 (Mb)
43.00
43.20
43.40
43.60
43.80
ACKR2→
←POMGNT2
SNRK→
ABHD5→
AC099329.2→
←AC104434.1
←ANO10
AC092042.3→
RN7SL517P→
←AC006055.1
KRBOX1→
←SNRK-AS1
←CYP8B1
Y_RNA→

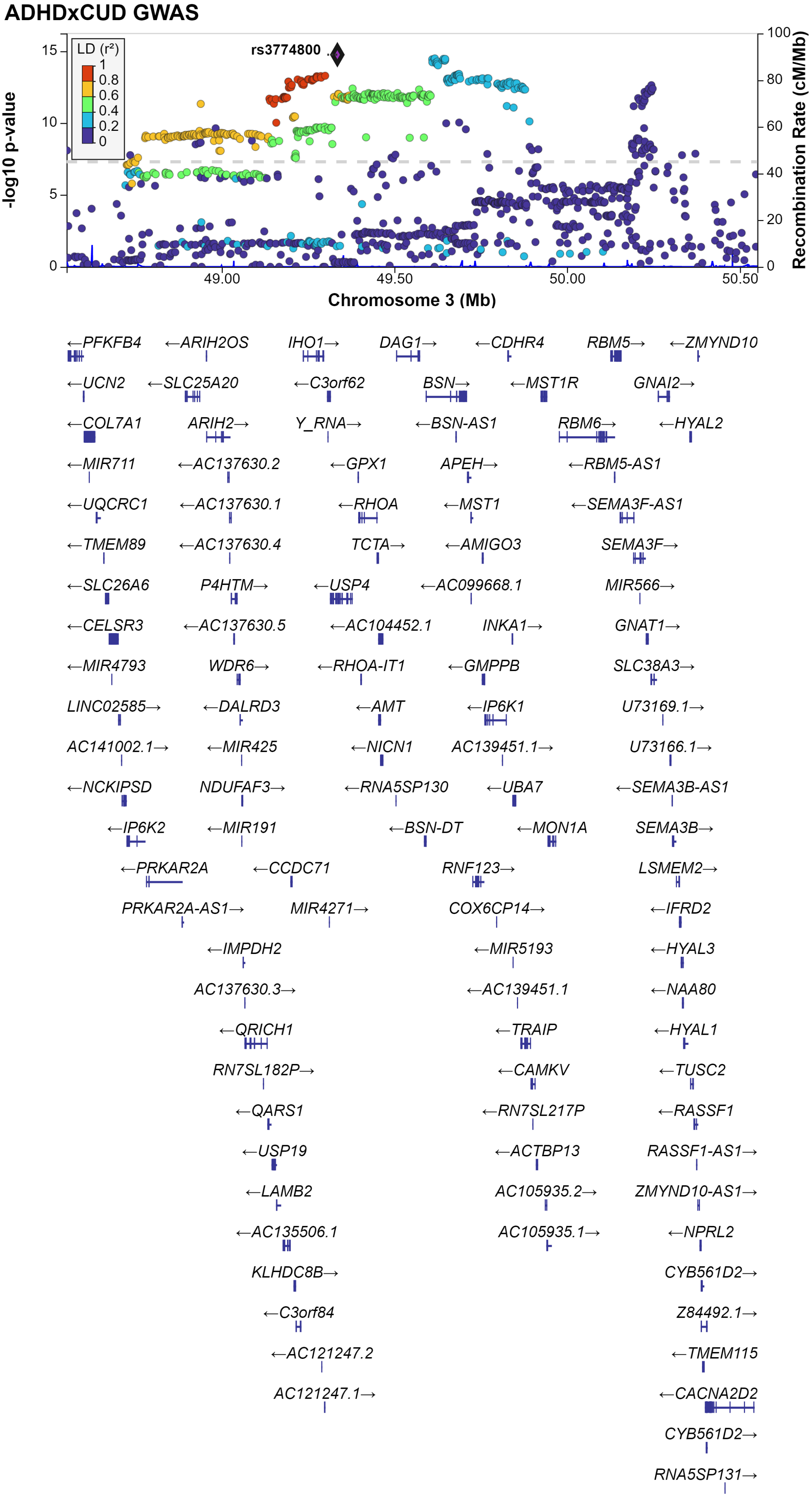

ADHDxCUD GWAS
LD (r²)
1
0.8
0.6
0.4
0.2
0
rs3774800
-log10 p-value
15
10
5
0
100
80
60
40
20
0
Recombination Rate (cM/Mb)
49.00
49.50
50.00
50.50
Chromosome 3 (Mb)
←PFKFB4
←ARIH2OS
IHO1→
DAG1→
←CDHR4
RBM5→
←ZMYND10
←UCN2
←SLC25A20
←C3orf62
BSN→
←MST1R
GNAI2→
←COL7A1
ARIH2→
Y_RNA→
←BSN-AS1
RBM6→
←HYAL2
←MIR711
←AC137630.2
←GPX1
APEH→
←RBM5-AS1
←UQCRC1
←AC137630.1
←RHOA
←MST1
←SEMA3F-AS1
←TMEM89
←AC137630.4
TCTA→
←AMIGO3
SEMA3F→
←SLC26A6
P4HTM→
←USP4
←AC099668.1
MIR566→
←CELSR3
←AC137630.5
←AC104452.1
INKA1→
GNAT1→
←MIR4793
WDR6→
←RHOA-IT1
←GMPPB
SLC38A3→
LINC02585→
←DALRD3
←AMT
←IP6K1
U73169.1→
AC141002.1→
←MIR425
←NICN1
AC139451.1→
U73166.1→
←NCKIPSD
NDUFAF3→
←RNA5SP130
←UBA7
←SEMA3B-AS1
←IP6K2
←MIR191
←BSN-DT
←MON1A
SEMA3B→
←PRKAR2A
←CCDC71
RNF123→
LSMEM2→
PRKAR2A-AS1→
MIR4271→
COX6CP14→
←IFRD2
←IMPDH2
←MIR5193
←HYAL3
AC137630.3→
←AC139451.1
←NAA80
←QRICH1
←TRAIP
←HYAL1
RN7SL182P→
←CAMKV
←TUSC2
←QARS1
←RN7SL217P
←RASSF1
←USP19
←ACTBP13
RASSF1-AS1→
←LAMB2
AC105935.2→
ZMYND10-AS1→
←AC135506.1
AC105935.1→
←NPRL2
KLHDC8B→
CYB561D2→
←C3orf84
Z84492.1→
←AC121247.2
←TMEM115
AC121247.1→
←CACNA2D2
CYB561D2→
RNA5SP131→

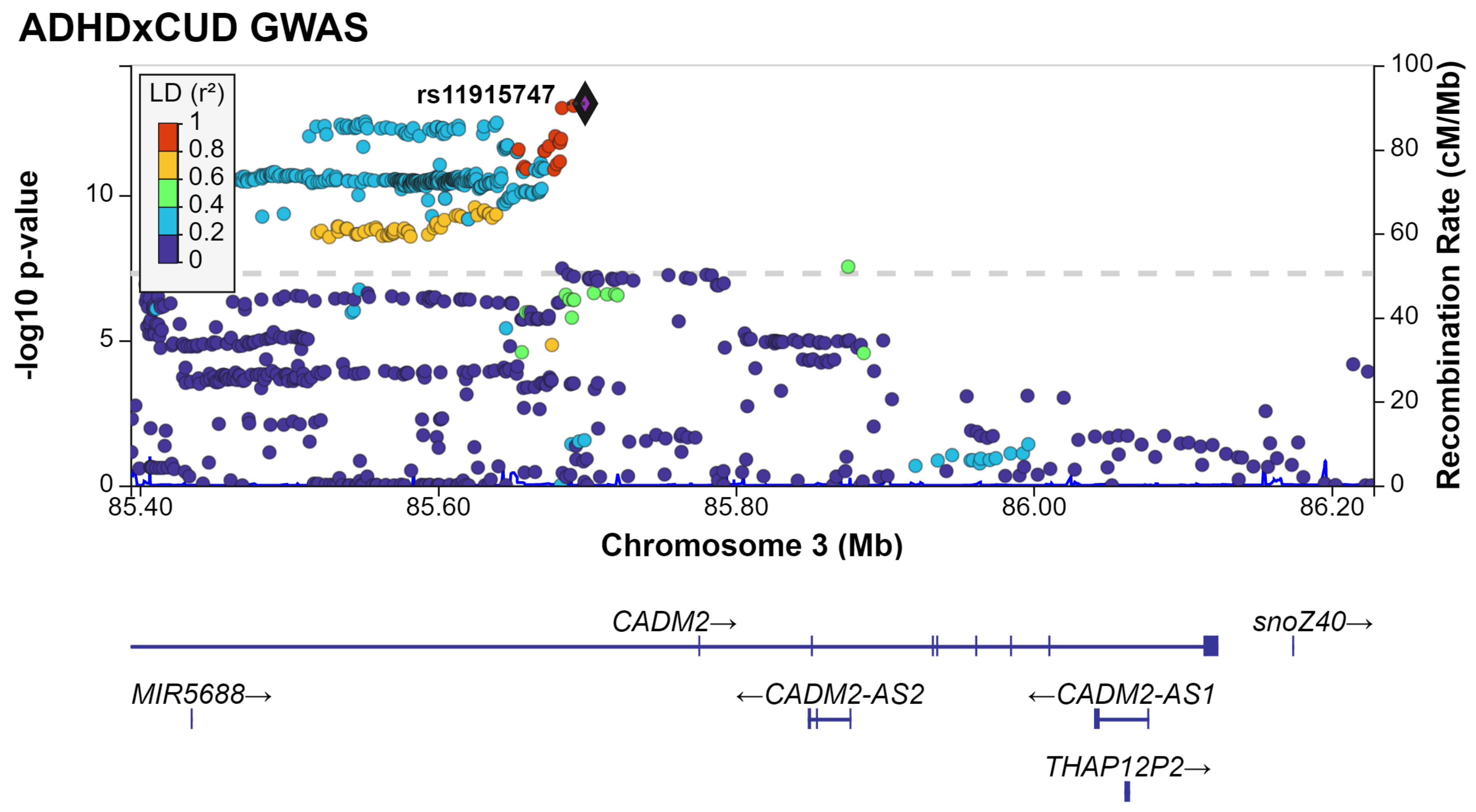
ADHDxCUD GWAS
LD (r²)
1
0.8
0.6
0.4
0.2
0
rs11915747
-log10 p-value
0
5
10
Recombination Rate (cM/Mb)
0
20
40
60
80
100
85.40
85.60
85.80
86.00
86.20
Chromosome 3 (Mb)
CADM2→
snoZ40→
MIR5688→
←CADM2-AS2
←CADM2-AS1
THAP12P2→

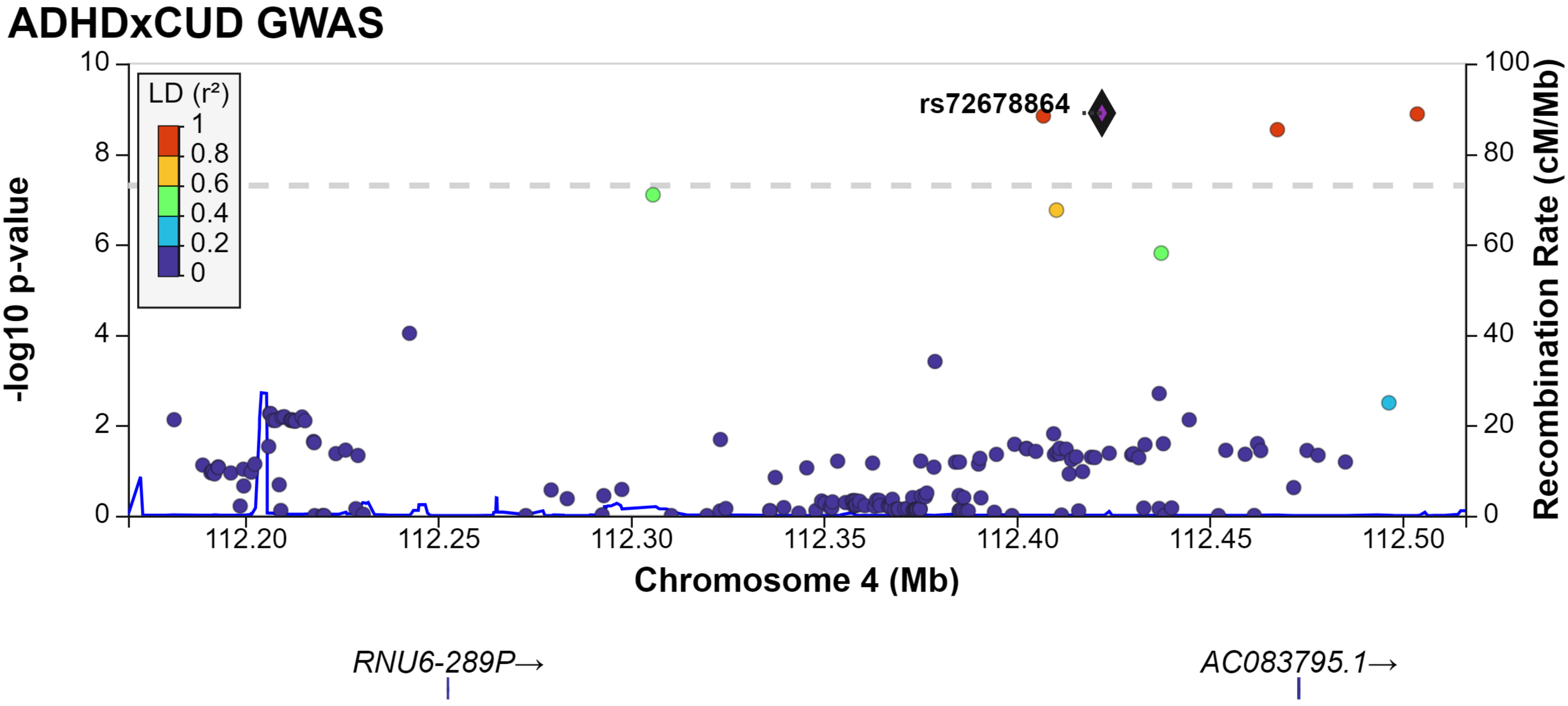
ADHDxCUD GWAS
LD (r²)
1
0.8
0.6
0.4
0.2
0
rs72678864
-log10 p-value
Recombination Rate (cM/Mb)
Chromosome 4 (Mb)
112.20
112.25
112.30
112.35
112.40
112.45
112.50
RNU6-289P→
AC083795.1→

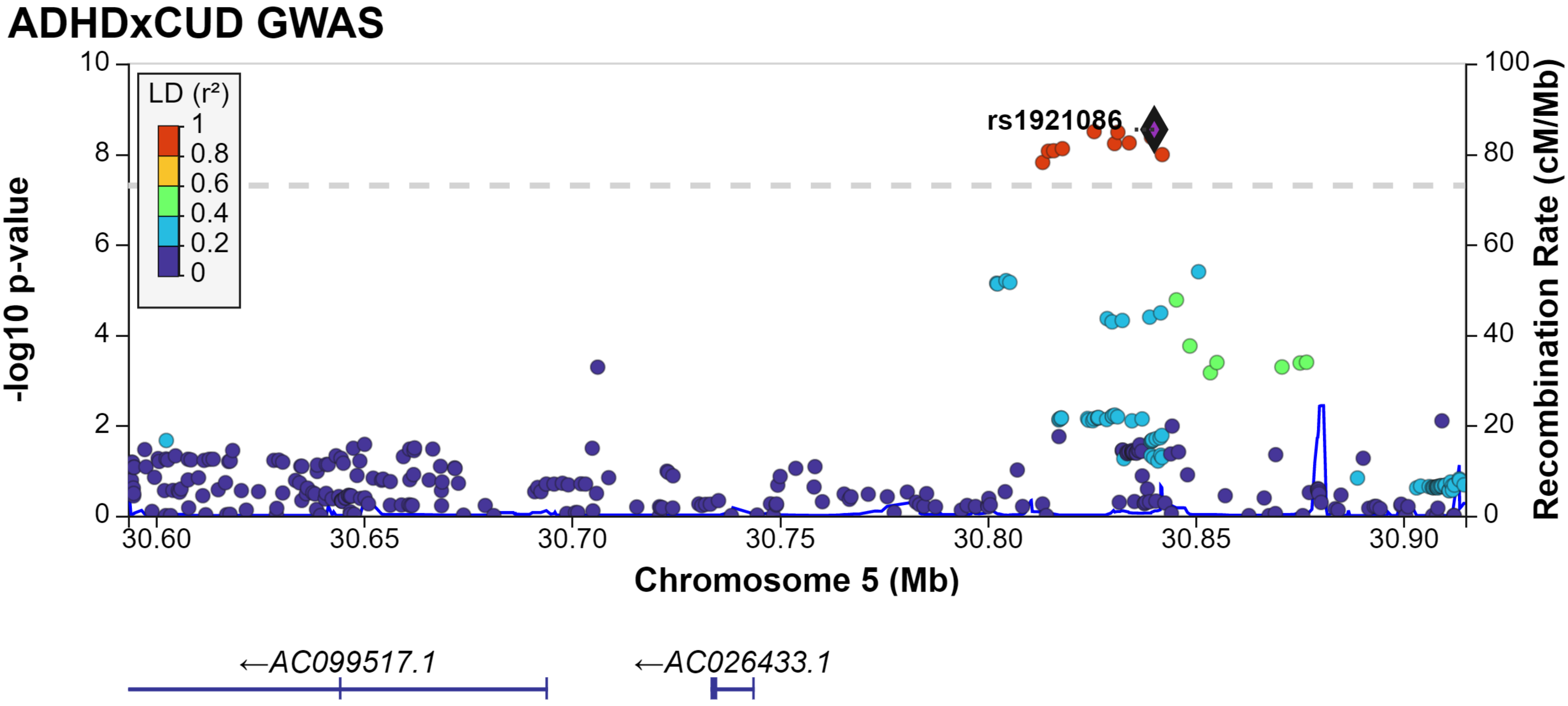
ADHDxCUD GWAS
LD (r²)
1
0.8
0.6
0.4
0.2
0
rs1921086
-log10 p-value
Recombination Rate (cM/Mb)
10
8
6
4
2
0
100
80
60
40
20
0
30.60
30.65
30.70
30.75
30.80
30.85
30.90
Chromosome 5 (Mb)
←AC099517.1
←AC026433.1

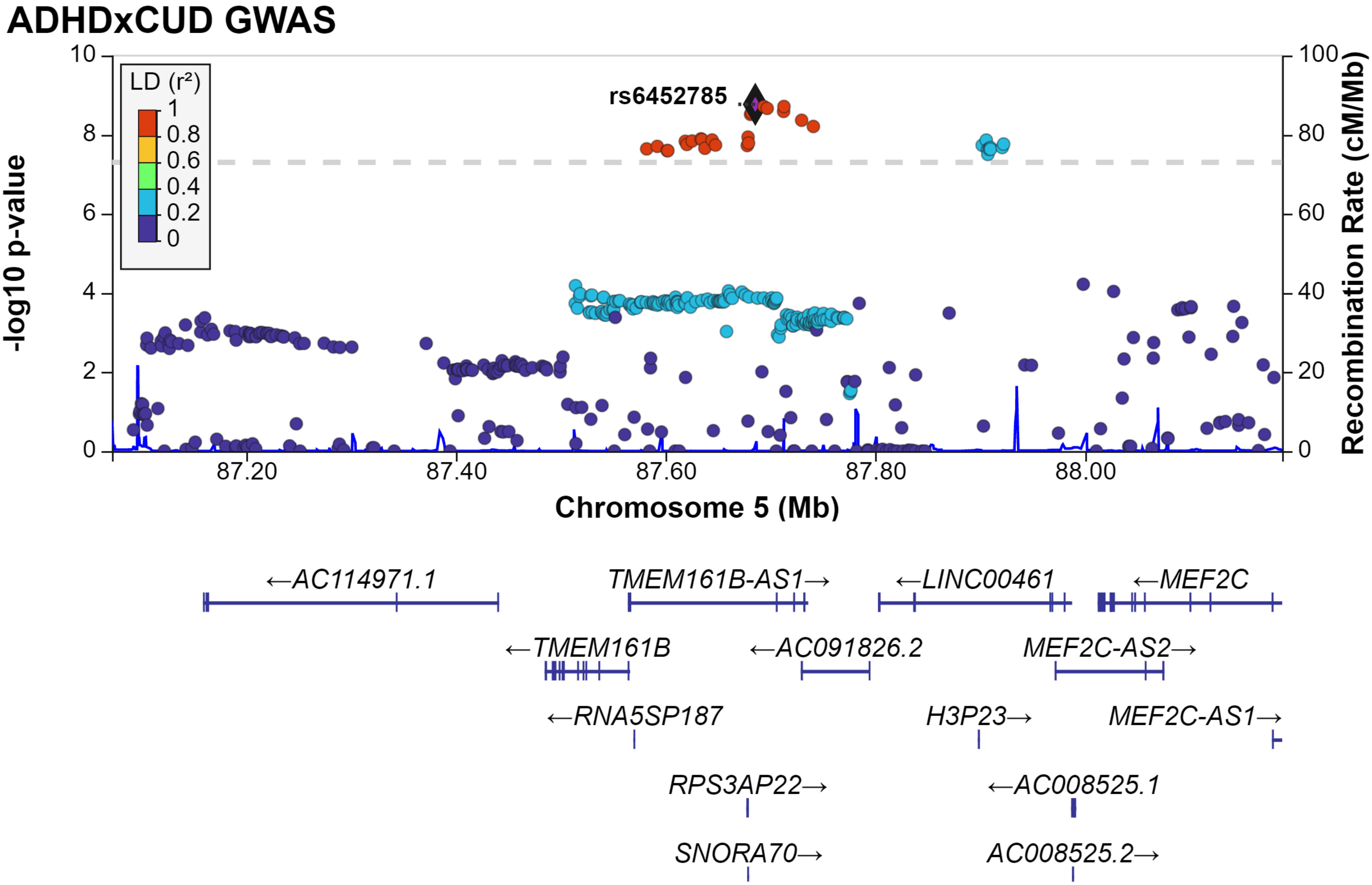
ADHDxCUD GWAS
LD (r²)
1
0.8
0.6
0.4
0.2
0
rs6452785
-log10 p-value
Recombination Rate (cM/Mb)
10
8
6
4
2
0
100
80
60
40
20
0
87.20
87.40
87.60
87.80
88.00
Chromosome 5 (Mb)
←AC114971.1
TMEM161B-AS1→
←LINC00461
←MEF2C
←TMEM161B
←AC091826.2
MEF2C-AS2→
←RNA5SP187
H3P23→
MEF2C-AS1→
RPS3AP22→
←AC008525.1
SNORA70→
AC008525.2→

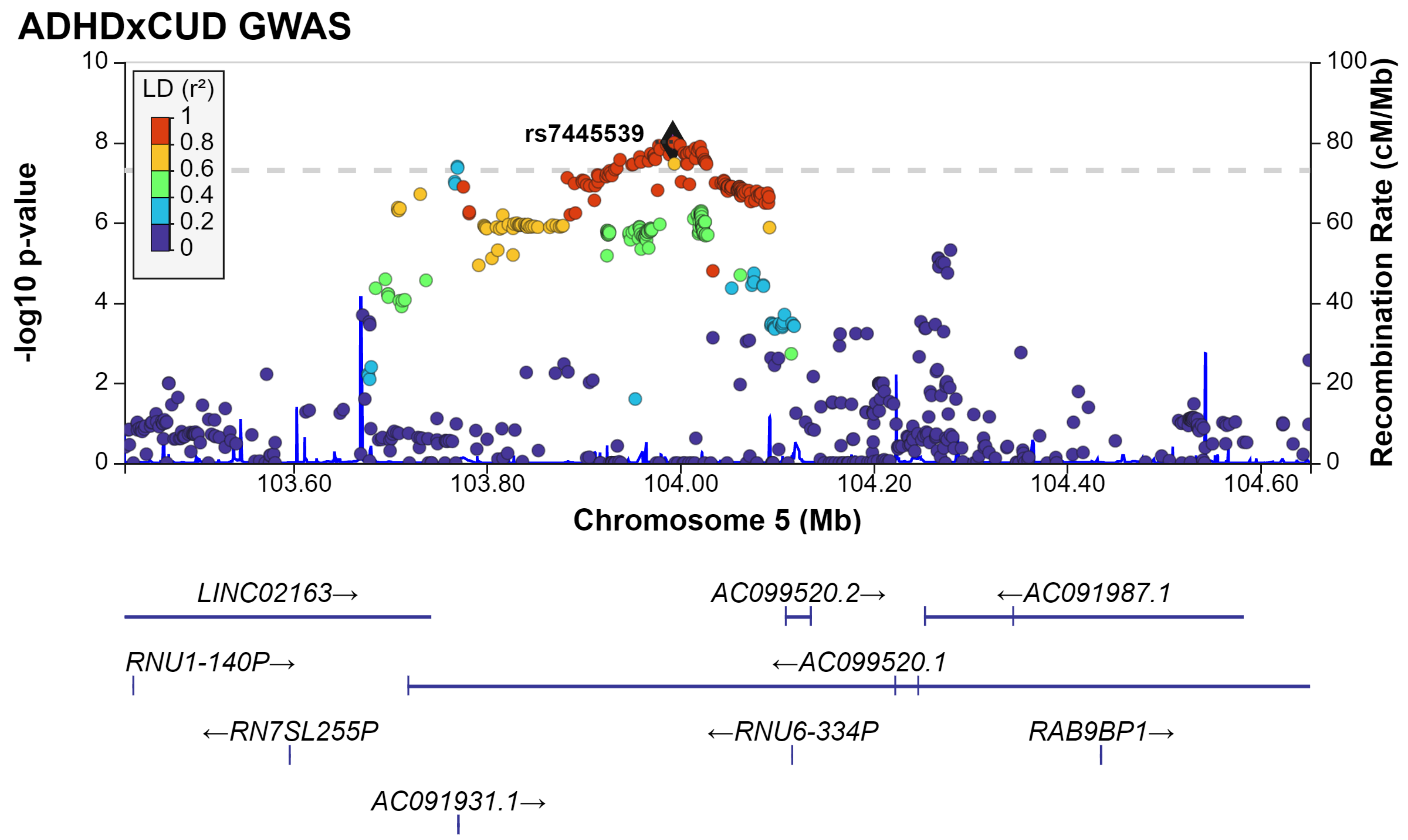
ADHDxCUD GWAS
LD (r²)
1
0.8
0.6
0.4
0.2
0
rs7445539
-log10 p-value
Recombination Rate (cM/Mb)
Chromosome 5 (Mb)
103.60
103.80
104.00
104.20
104.40
104.60
LINC02163→
AC099520.2→
←AC091987.1
RNU1-140P→
←AC099520.1
←RN7SL255P
←RNU6-334P
RAB9BP1→
AC091931.1→

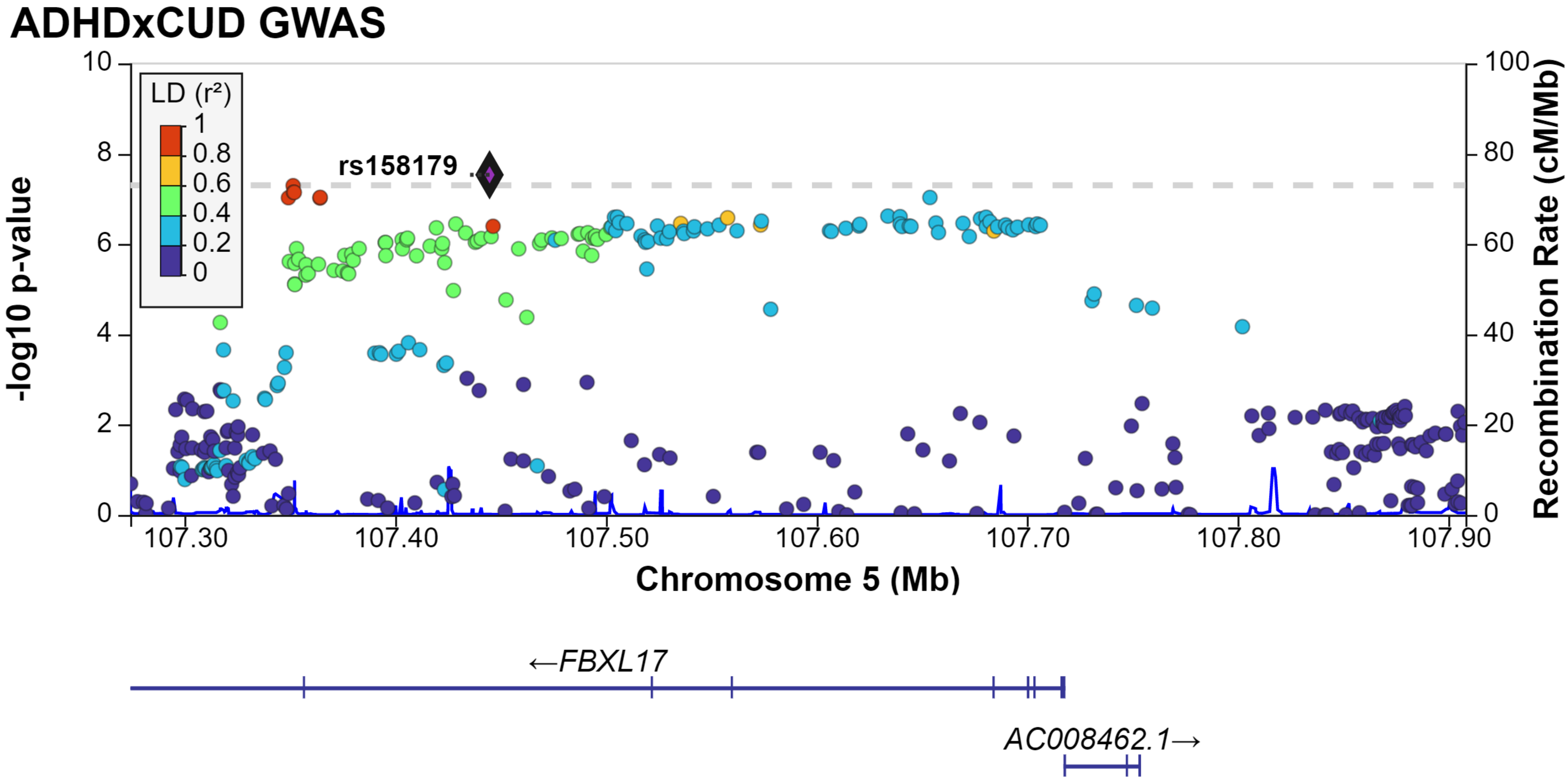
ADHDxCUD GWAS
LD (r²)
1
0.8
0.6
0.4
0.2
0
rs158179
-log10 p-value
Recombination Rate (cM/Mb)
Chromosome 5 (Mb)
107.30
107.40
107.50
107.60
107.70
107.80
107.90
←FBXL17
AC008462.1→

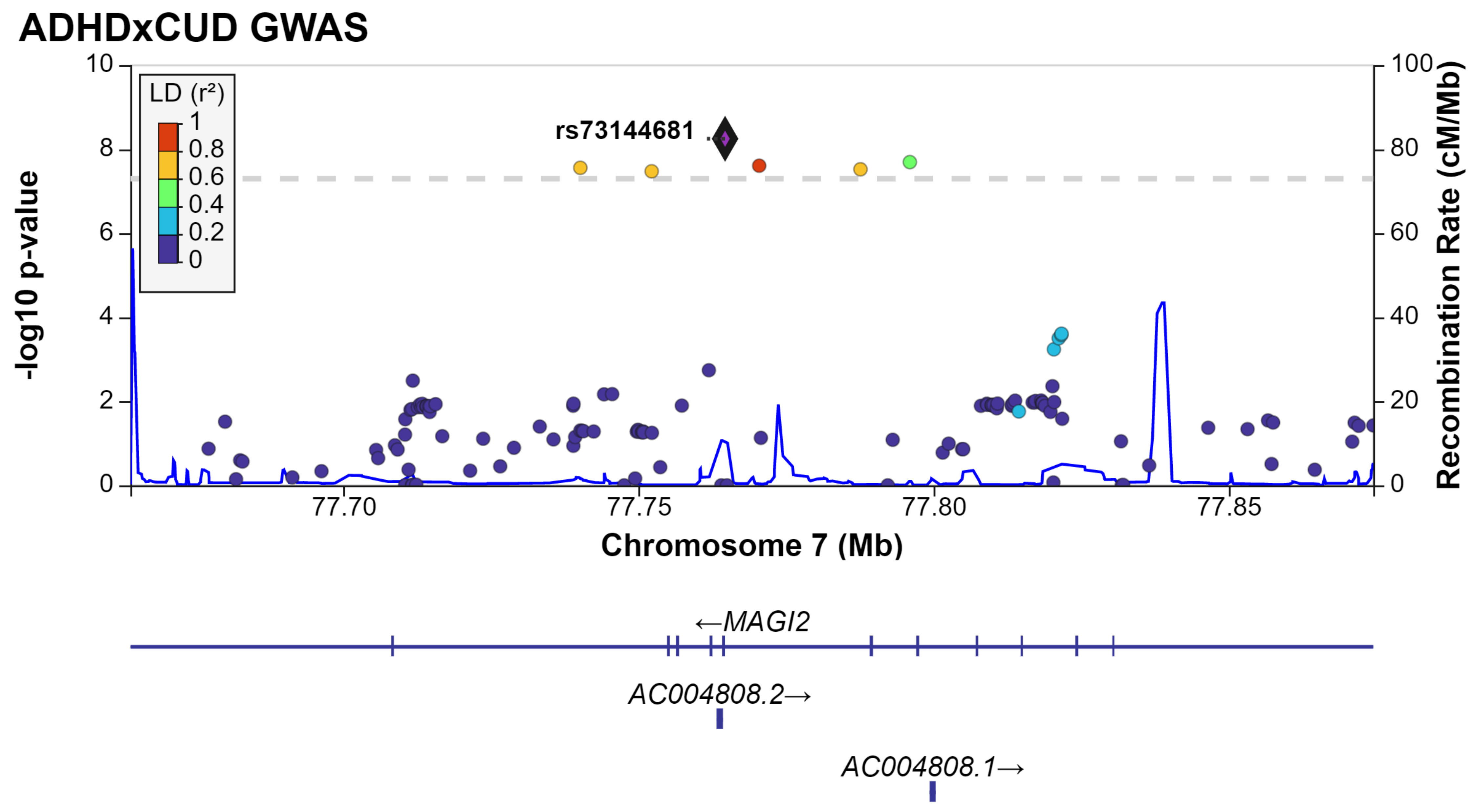
ADHDxCUD GWAS
LD (r²)
1
0.8
0.6
0.4
0.2
0
rs73144681
10
8
6
4
2
0
-log10 p-value
100
80
60
40
20
0
Recombination Rate (cM/Mb)
77.70
77.75
77.80
77.85
Chromosome 7 (Mb)
←MAGI2
AC004808.2→
AC004808.1→

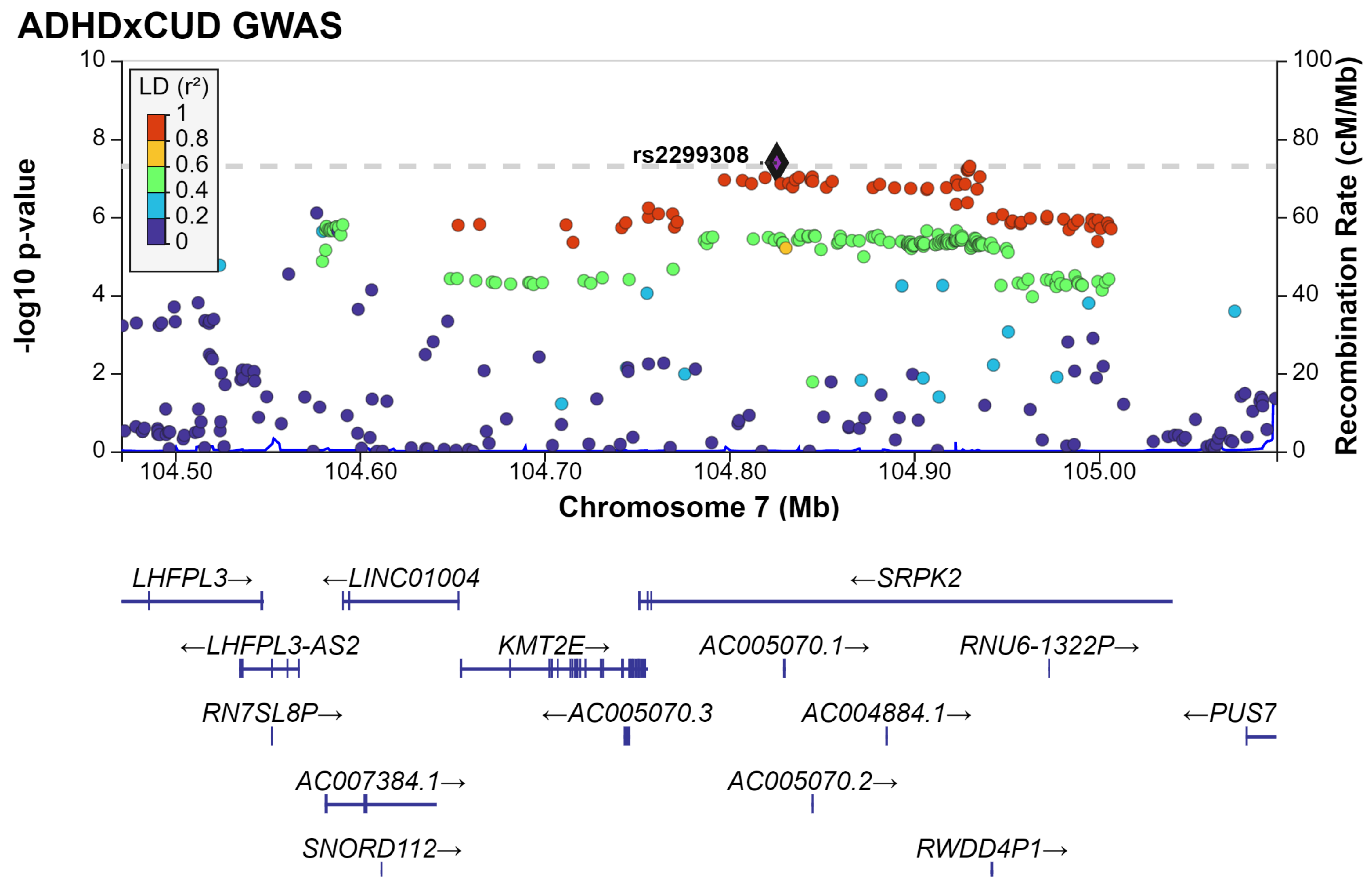
ADHDxCUD GWAS
LD (r²)
1
0.8
0.6
0.4
0.2
0
rs2299308
-log10 p-value
Recombination Rate (cM/Mb)
Chromosome 7 (Mb)
0
2
4
6
8
10
0
20
40
60
80
100
104.50
104.60
104.70
104.80
104.90
105.00
LHFPL3→
←LINC01004
←SRPK2
←LHFPL3-AS2
KMT2E→
AC005070.1→
RNU6-1322P→
RN7SL8P→
←AC005070.3
AC004884.1→
←PUS7
AC007384.1→
AC005070.2→
SNORD112→
RWDD4P1→

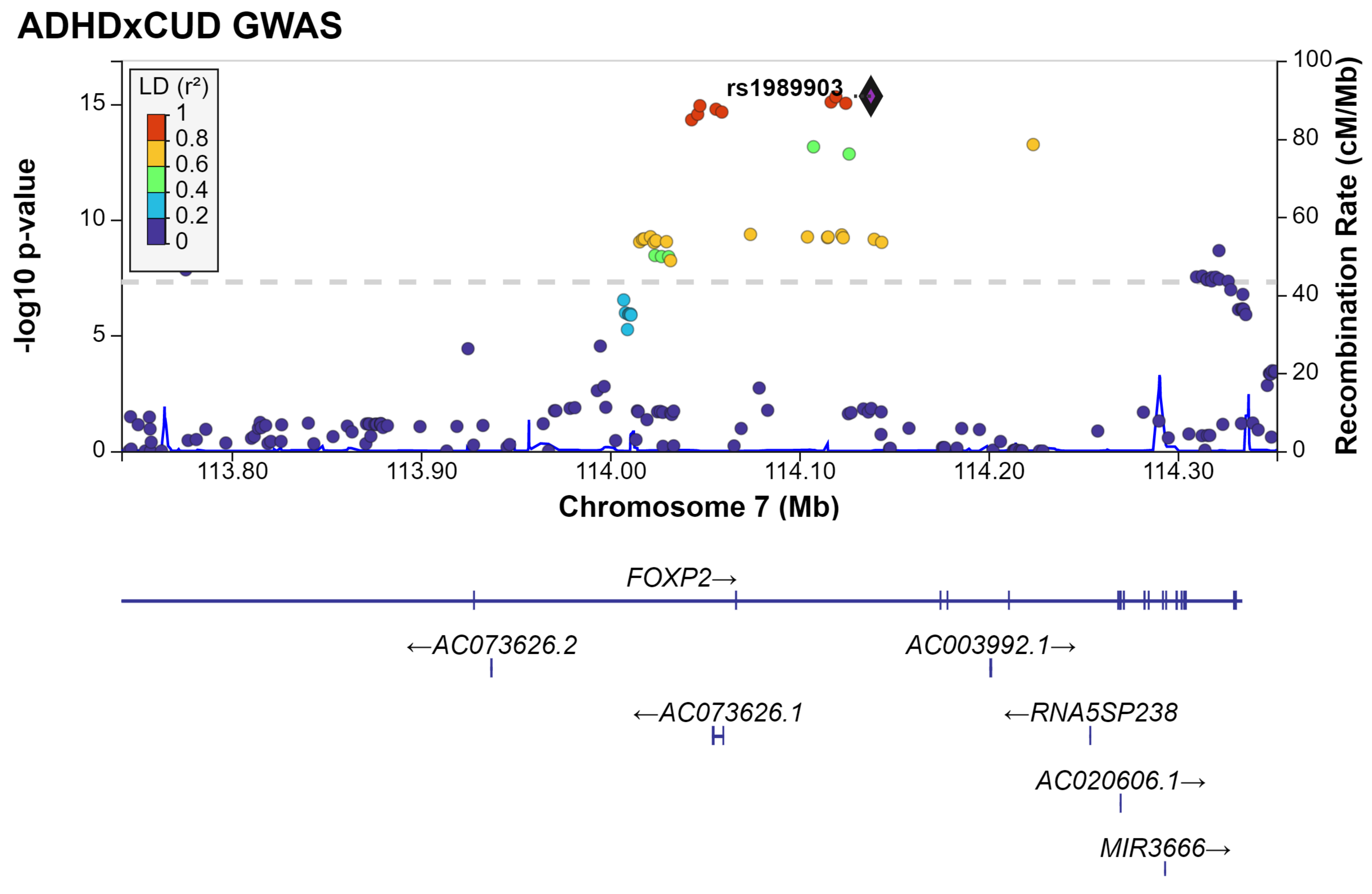

ADHDxCUD GWAS
LD (r²)
1
0.8
0.6
0.4
0.2
0
rs1989903
-log10 p-value
0
5
10
15
Recombination Rate (cM/Mb)
0
20
40
60
80
100
113.80
113.90
114.00
114.10
114.20
114.30
Chromosome 7 (Mb)
FOXP2→
←AC073626.2
AC003992.1→
←AC073626.1
←RNA5SP238
AC020606.1→
MIR3666→

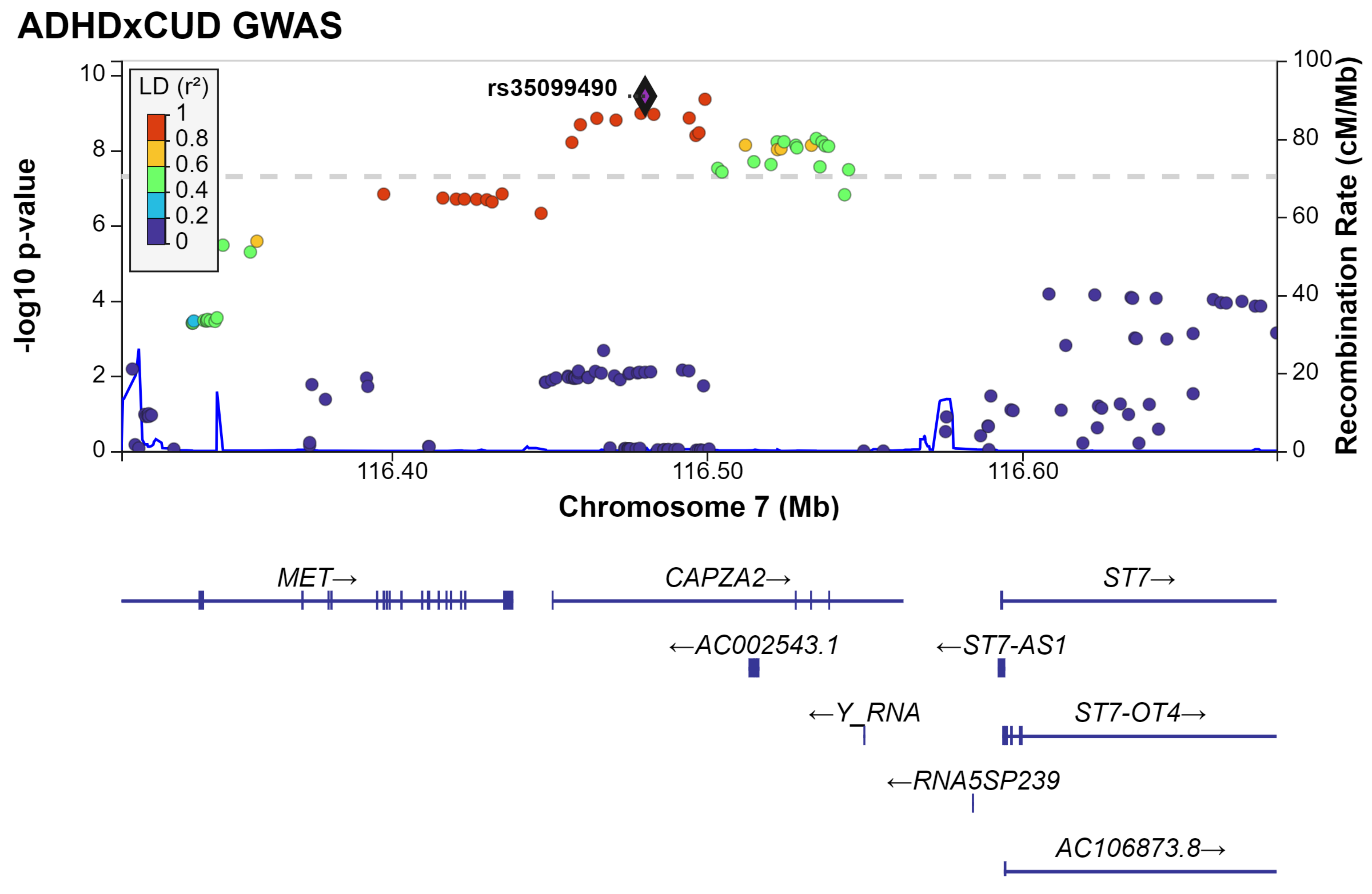

ADHDxCUD GWAS
LD (r²)
1
0.8
0.6
0.4
0.2
0
rs35099490
-log10 p-value
10
8
6
4
2
0
Recombination Rate (cM/Mb)
100
80
60
40
20
0
116.40
116.50
116.60
Chromosome 7 (Mb)
MET→
CAPZA2→
ST7→
←AC002543.1
←ST7-AS1
←Y_RNA
ST7-OT4→
←RNA5SP239
AC106873.8→

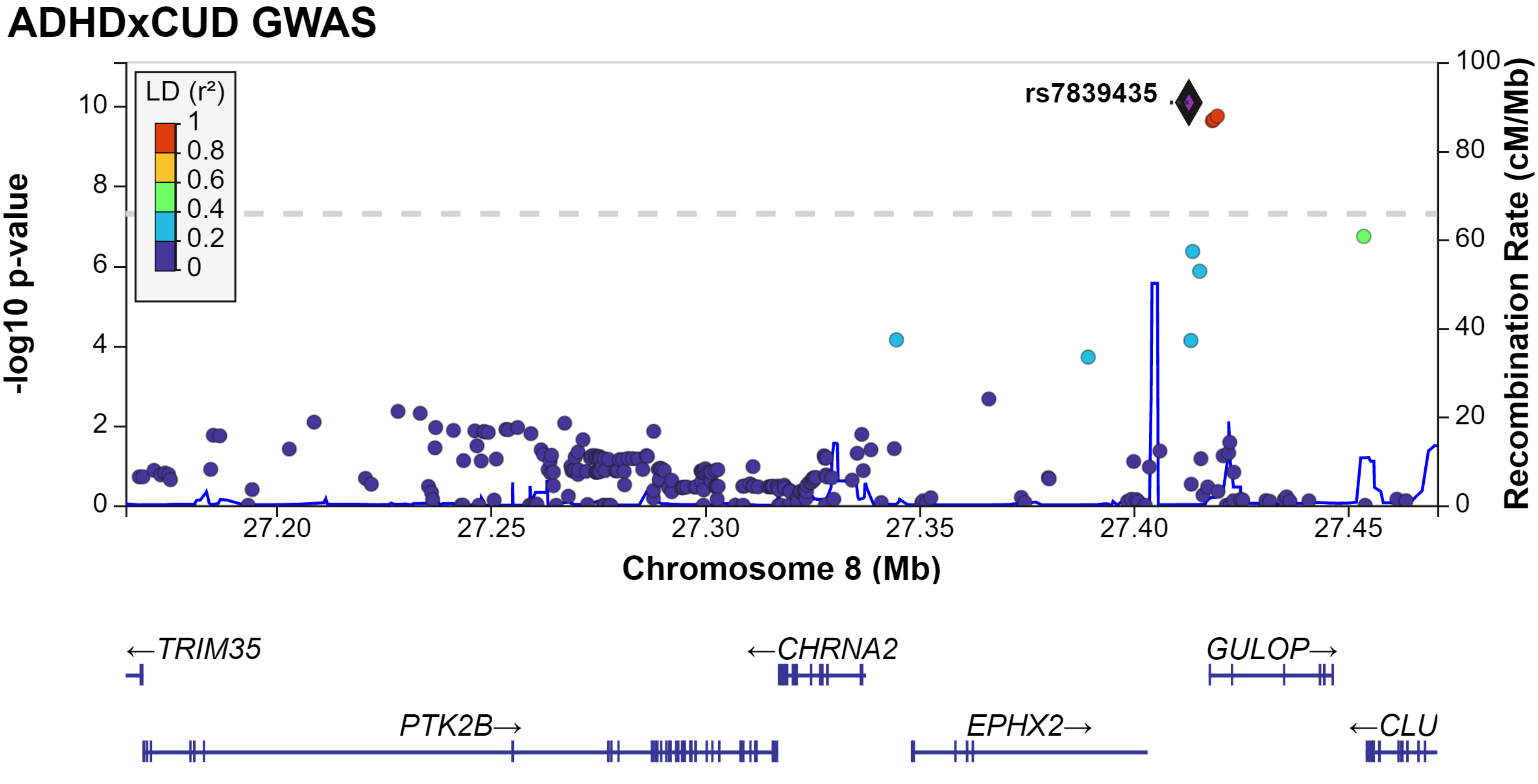

ADHDxCUD GWAS
LD (r²)
1
0.8
0.6
0.4
0.2
0
rs7839435
-log10 p-value
Recombination Rate (cM/Mb)
Chromosome 8 (Mb)
27.20
27.25
27.30
27.35
27.40
27.45
←TRIM35
←CHRNA2
GULOP→
PTK2B→
EPHX2→
←CLU

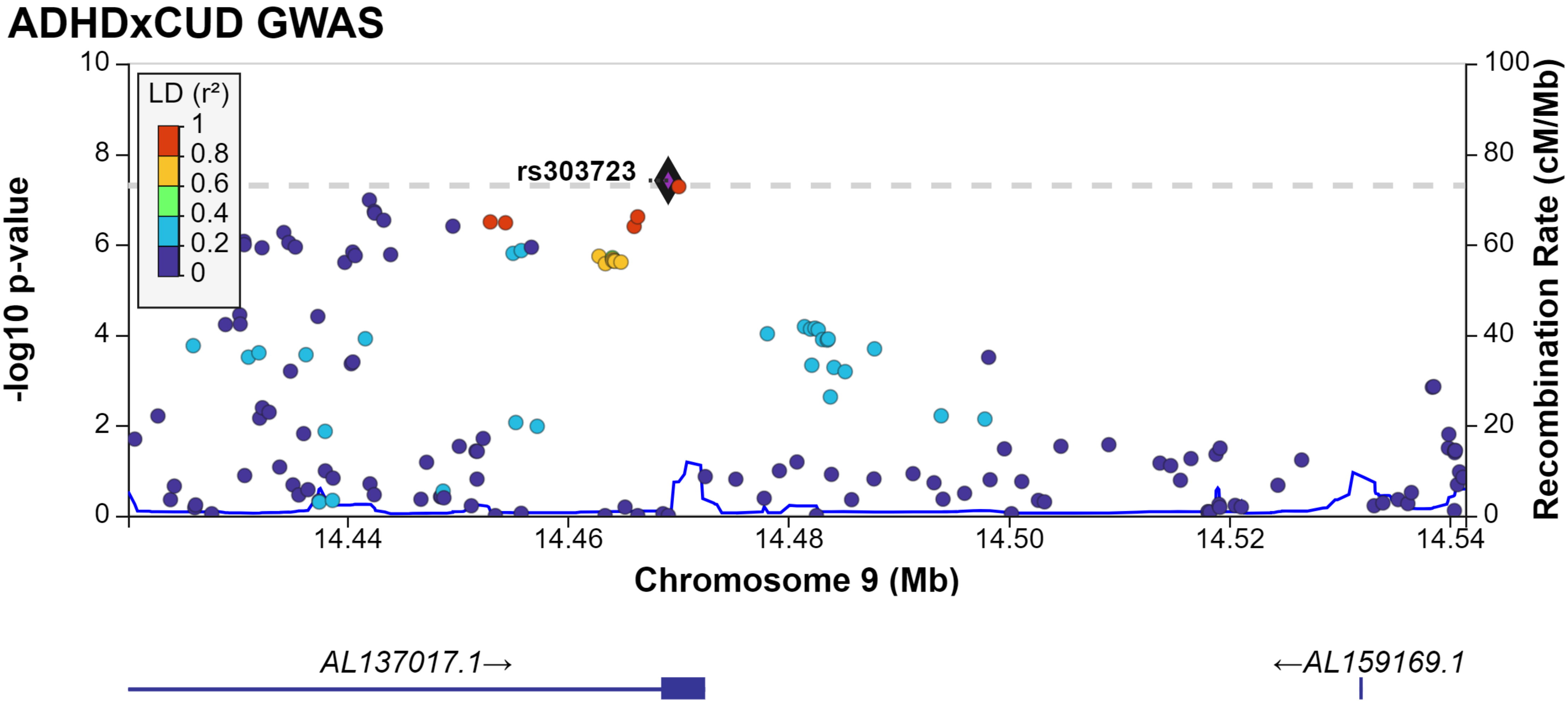
ADHDxCUD GWAS
-log10 p-value
Recombination Rate (cM/Mb)
LD (r²)
1
0.8
0.6
0.4
0.2
0
rs303723
10
8
6
4
2
0
100
80
60
40
20
0
14.44
14.46
14.48
14.50
14.52
14.54
Chromosome 9 (Mb)
AL137017.1→
←AL159169.1

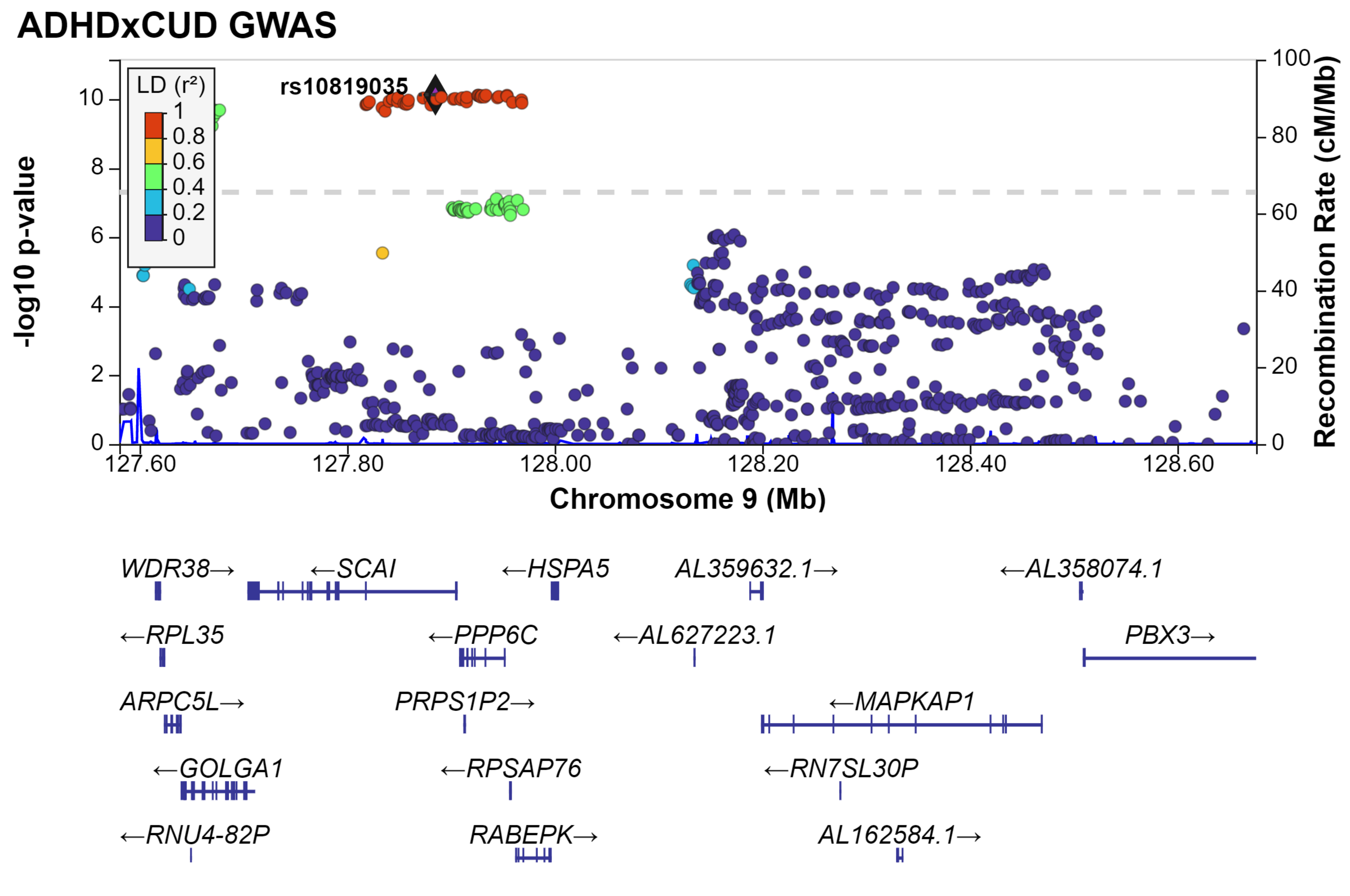

ADHDxCUD GWAS
rs10819035
LD (r²)
1
0.8
0.6
0.4
0.2
0
-log10 p-value
Recombination Rate (cM/Mb)
Chromosome 9 (Mb)
127.60
127.80
128.00
128.20
128.40
128.60
WDR38→
←SCAI
←HSPA5
AL359632.1→
←AL358074.1
←RPL35
←PPP6C
←AL627223.1
PBX3→
ARPC5L→
PRPS1P2→
←MAPKAP1
←GOLGA1
←RPSAP76
←RN7SL30P
←RNU4-82P
RABEPK→
AL162584.1→

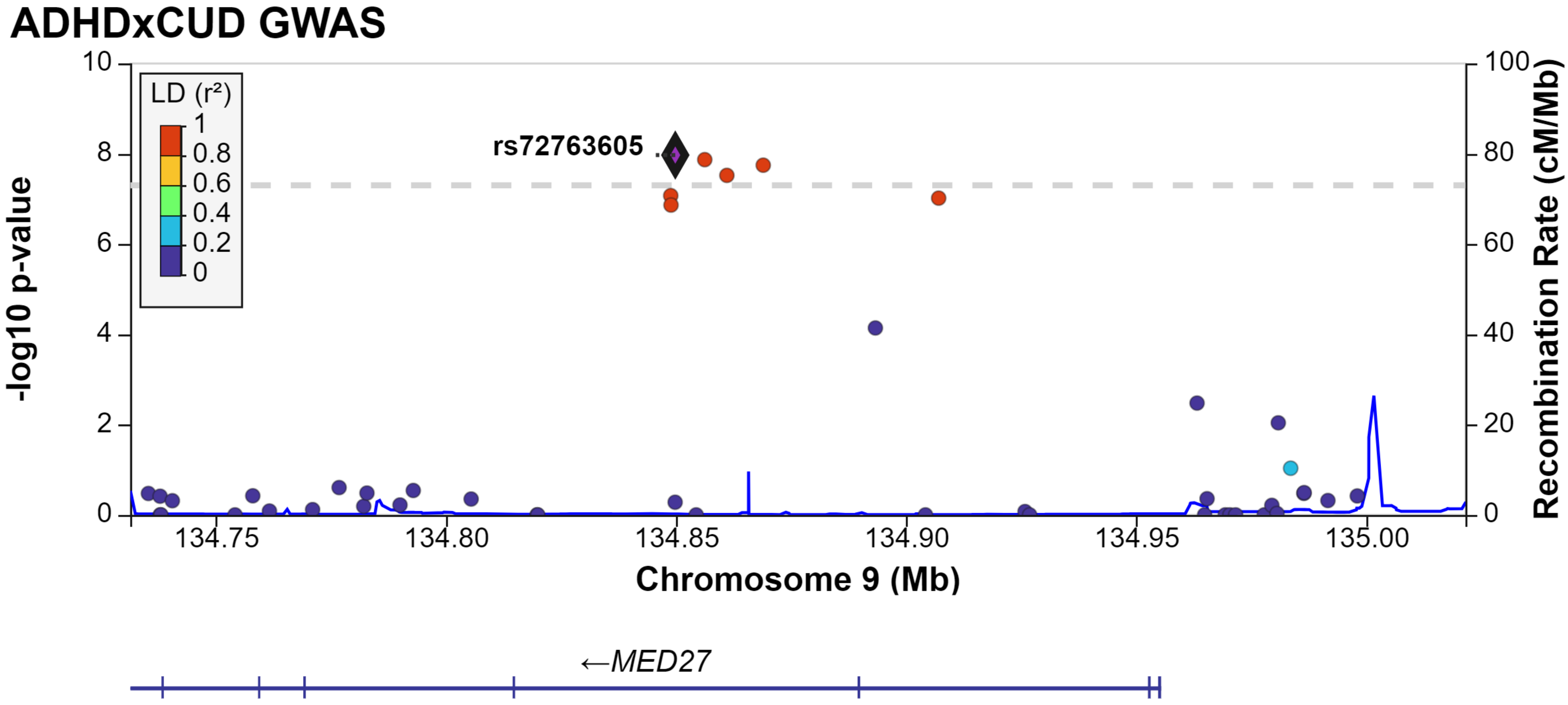
ADHDxCUD GWAS
-log10 p-value
Recombination Rate (cM/Mb)
LD (r²)
1
0.8
0.6
0.4
0.2
0
rs72763605
134.75
134.80
134.85
134.90
134.95
135.00
Chromosome 9 (Mb)
←MED27

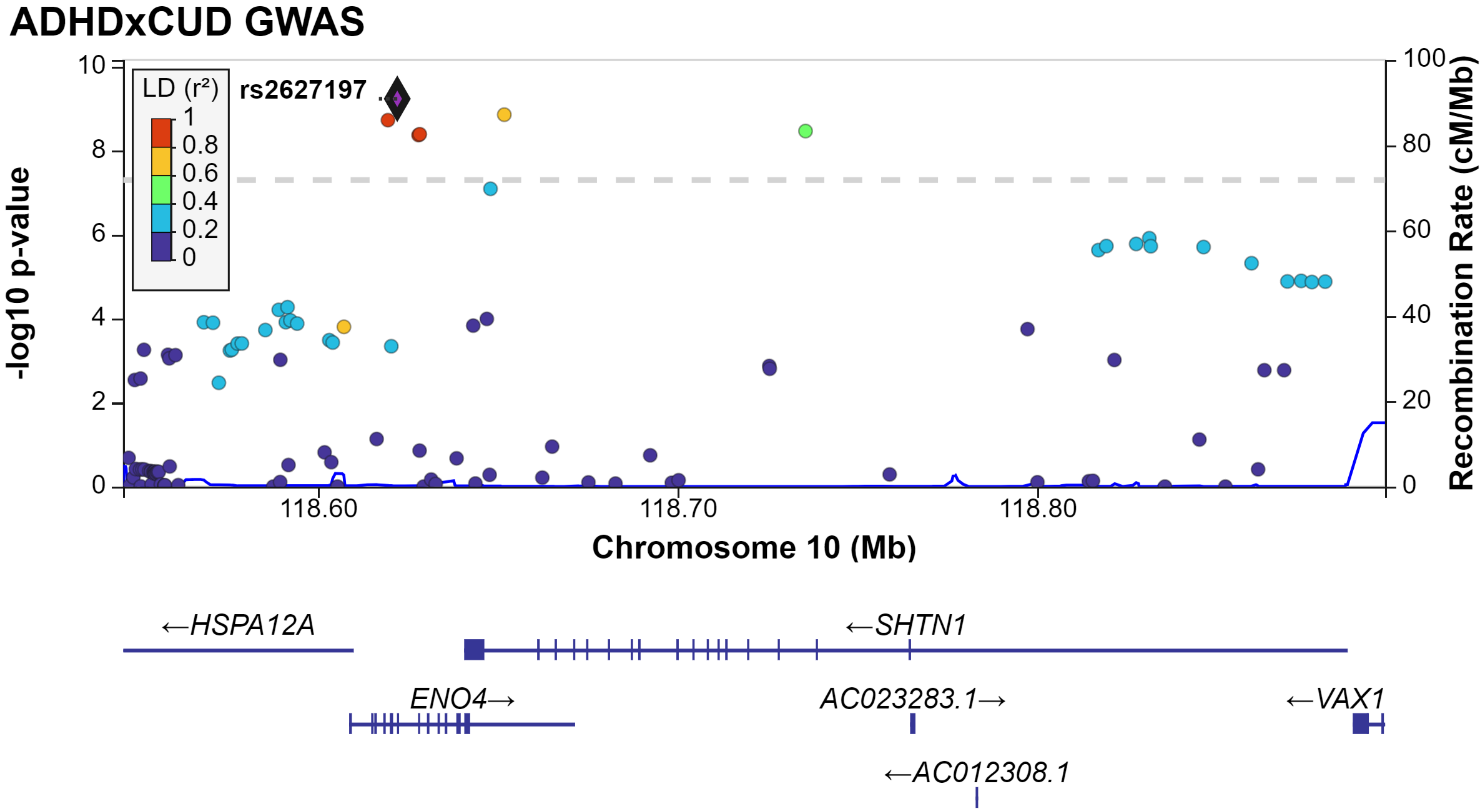

ADHDxCUD GWAS
LD (r²)
1
0.8
0.6
0.4
0.2
0
rs2627197
-log10 p-value
Recombination Rate (cM/Mb)
0
2
4
6
8
10
0
20
40
60
80
100
118.60
118.70
118.80
Chromosome 10 (Mb)
←HSPA12A
←SHTN1
ENO4→
AC023283.1→
←VAX1
←AC012308.1

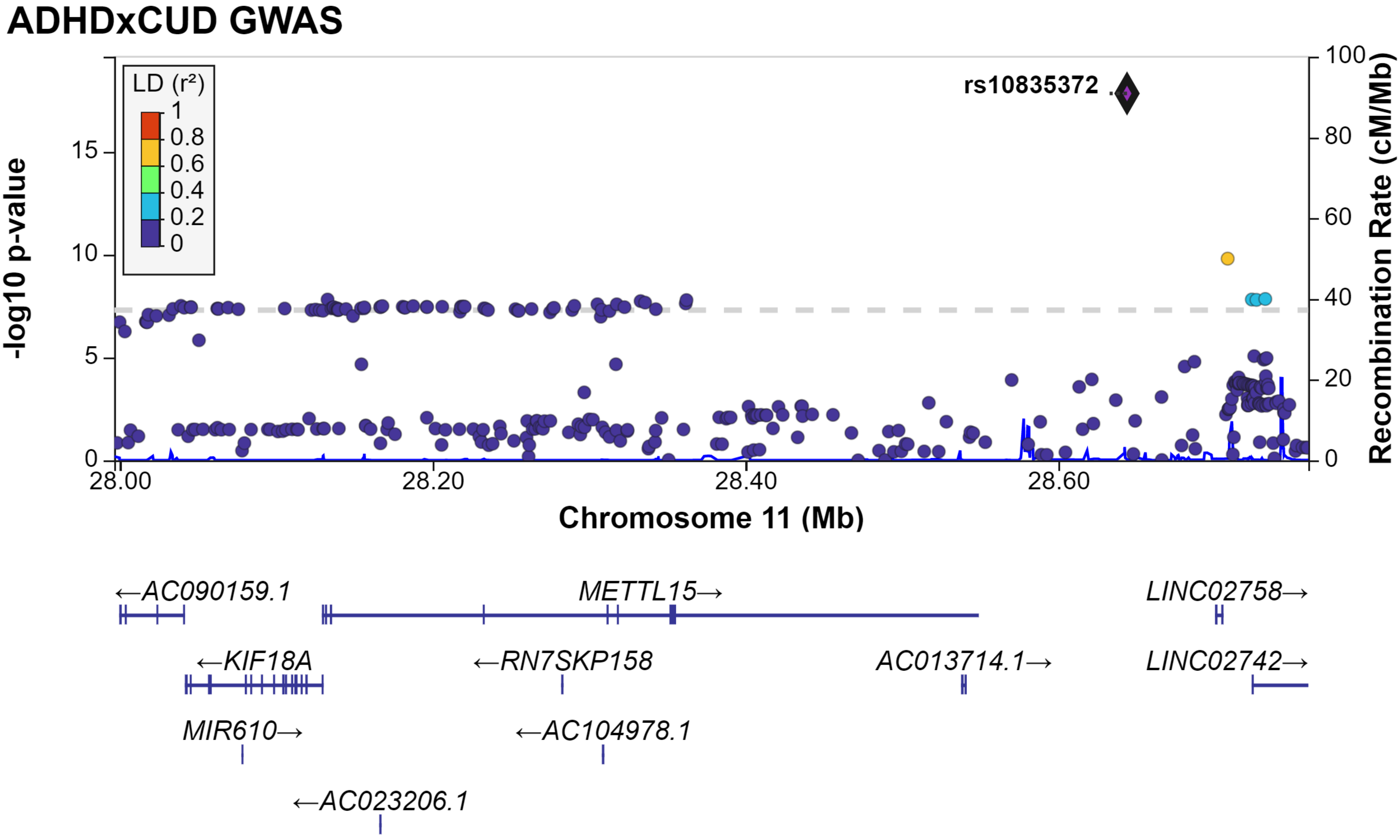
ADHDxCUD GWAS
LD (r²)
1
0.8
0.6
0.4
0.2
0
rs10835372
-log10 p-value
15
10
5
0
Recombination Rate (cM/Mb)
100
80
60
40
20
0
28.00
28.20
28.40
28.60
Chromosome 11 (Mb)
←AC090159.1
METTL15→
LINC02758→
←KIF18A
←RN7SKP158
AC013714.1→
LINC02742→
MIR610→
←AC104978.1
←AC023206.1

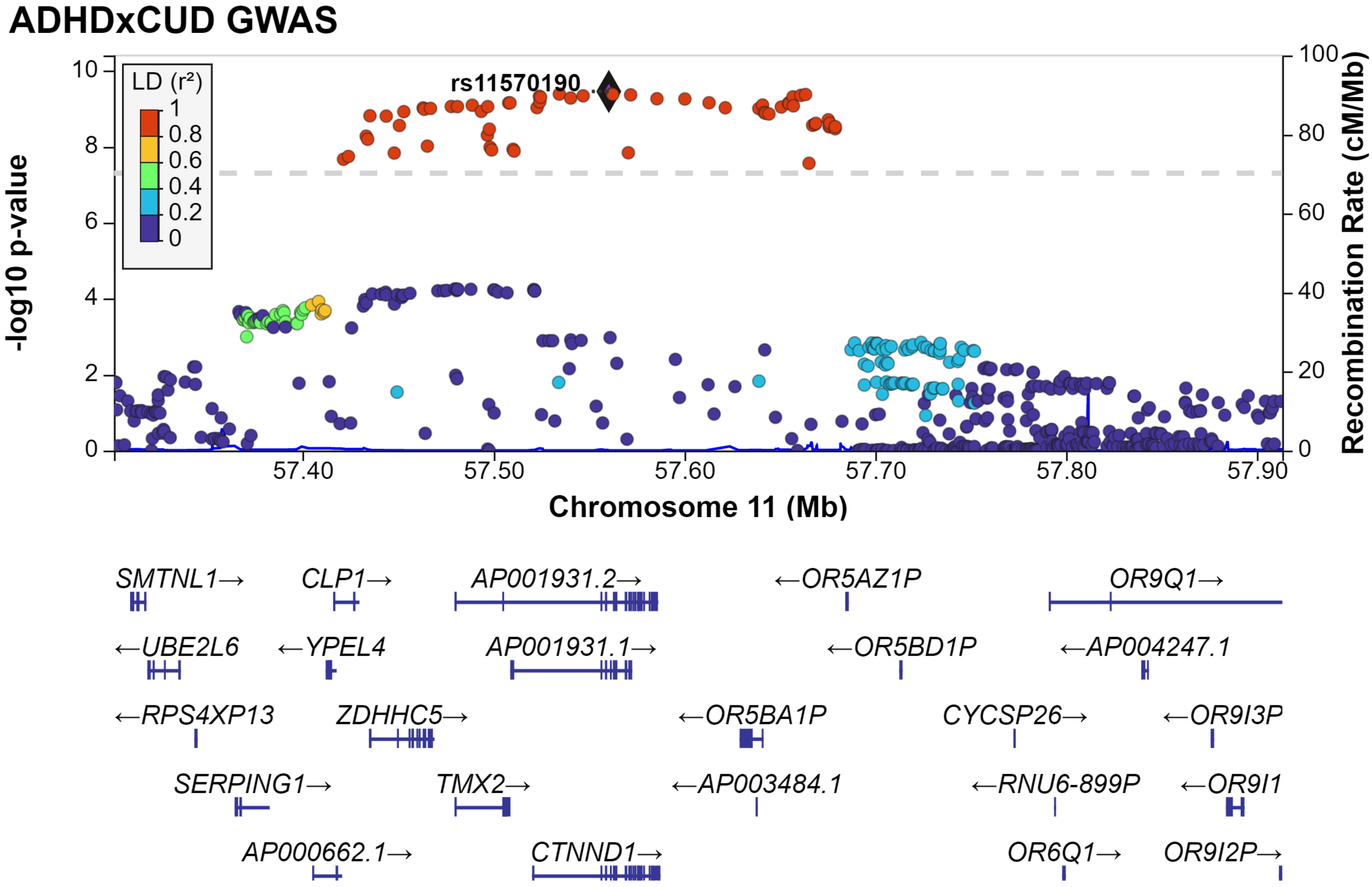
ADHDxCUD GWAS
rs11570190
LD (r²)
1
0.8
0.6
0.4
0.2
0
-log10 p-value
Recombination Rate (cM/Mb)
Chromosome 11 (Mb)
57.40
57.50
57.60
57.70
57.80
57.90
SMTNL1→
CLP1→
AP001931.2→
←OR5AZ1P
OR9Q1→
←UBE2L6
←YPEL4
AP001931.1→
←OR5BD1P
←AP004247.1
←RPS4XP13
ZDHHC5→
←OR5BA1P
CYCSP26→
←OR9I3P
SERPING1→
TMX2→
←AP003484.1
←RNU6-899P
←OR9I1
AP000662.1→
CTNND1→
OR6Q1→
OR9I2P→

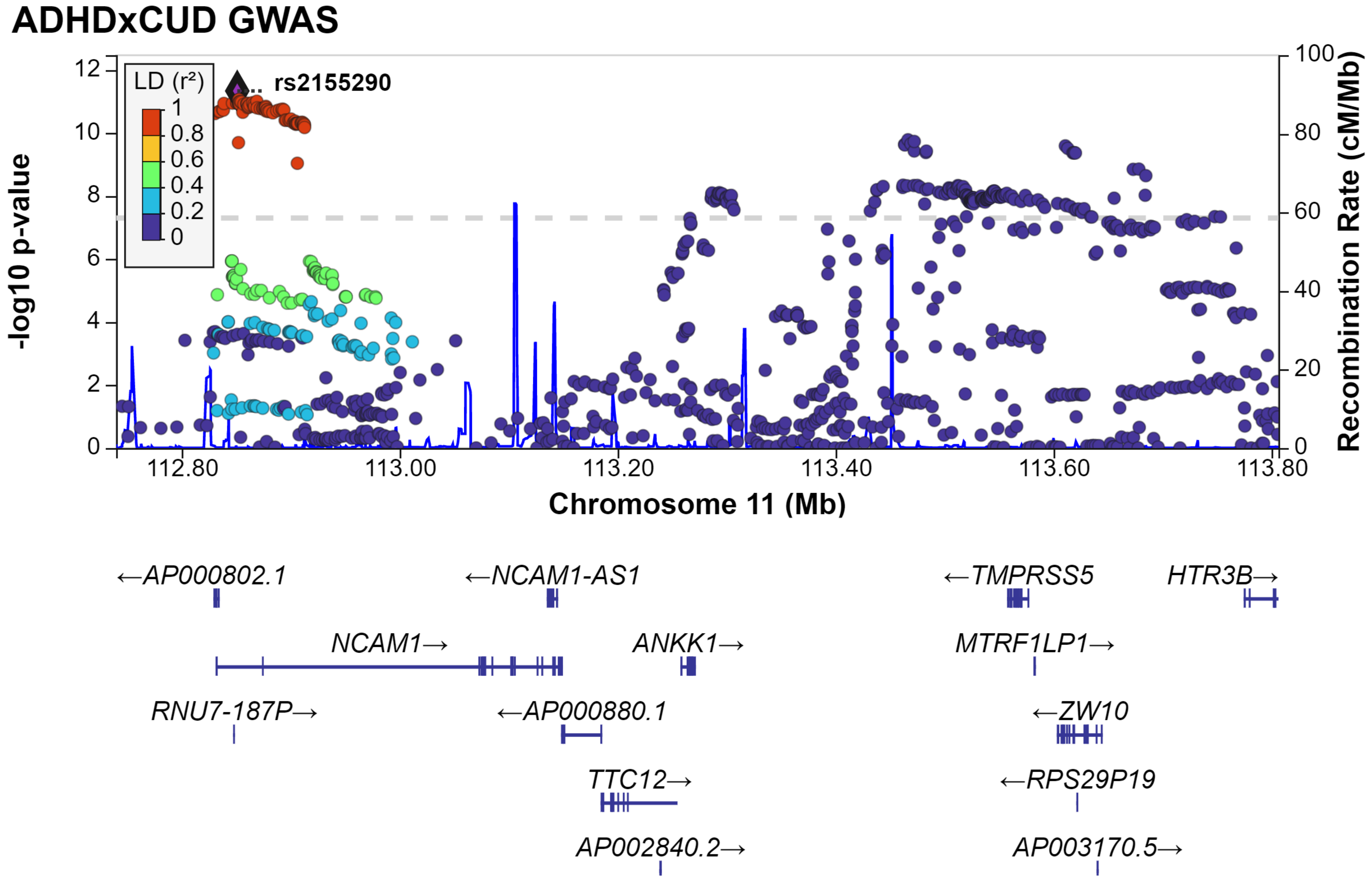
ADHDxCUD GWAS
rs2155290
LD (r²)
1
0.8
0.6
0.4
0.2
0
-log10 p-value
Recombination Rate (cM/Mb)
Chromosome 11 (Mb)
112.80
113.00
113.20
113.40
113.60
113.80
←AP000802.1
←NCAM1-AS1
←TMPRSS5
HTR3B→
NCAM1→
ANKK1→
MTRF1LP1→
RNU7-187P→
←AP000880.1
←ZW10
TTC12→
←RPS29P19
AP002840.2→
AP003170.5→

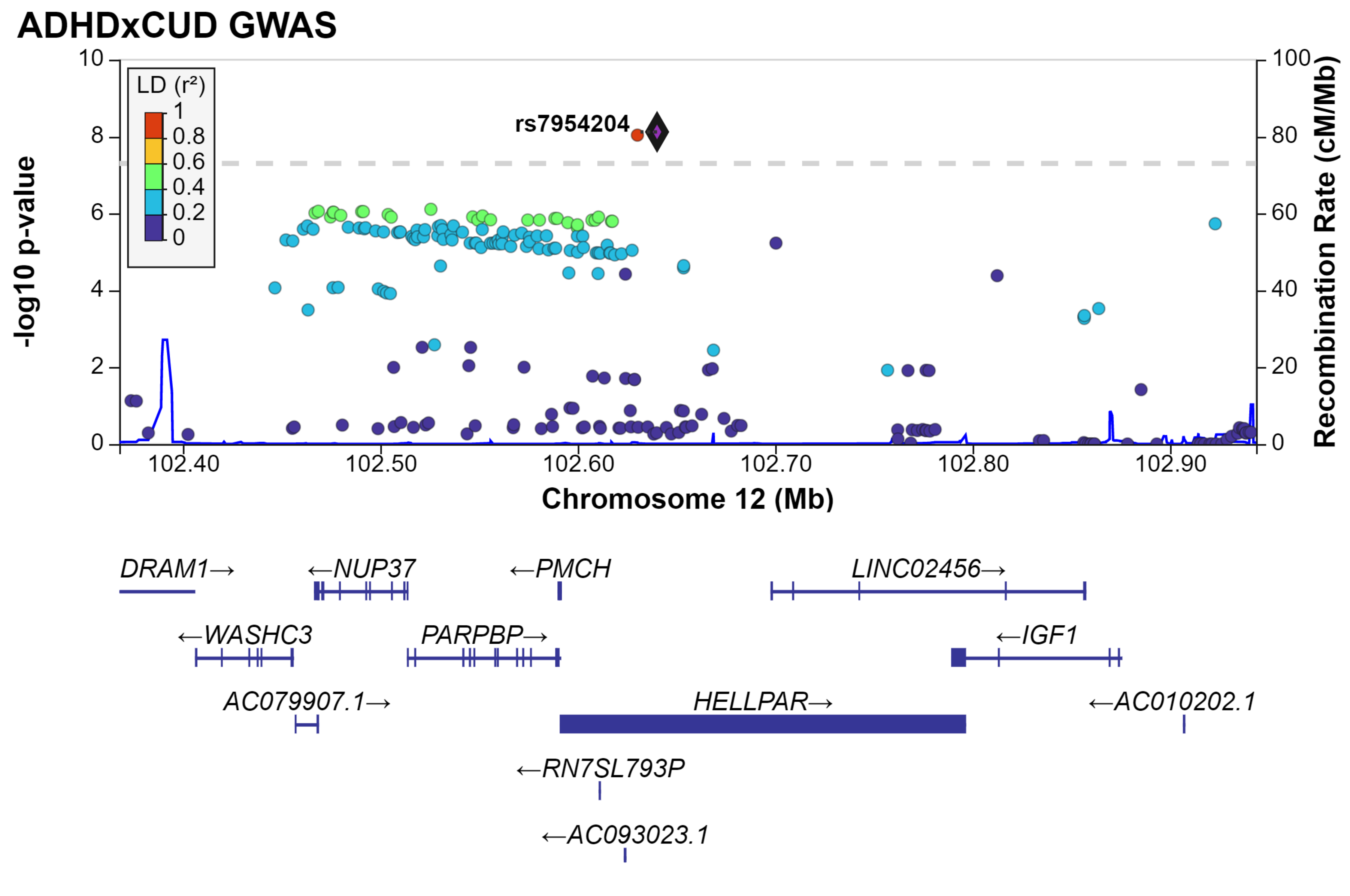
ADHDxCUD GWAS
LD (r²)
1
0.8
0.6
0.4
0.2
0
rs7954204
-log10 p-value
Recombination Rate (cM/Mb)
Chromosome 12 (Mb)
102.40
102.50
102.60
102.70
102.80
102.90
DRAM1→
←NUP37
←PMCH
LINC02456→
←WASHC3
PARPBP→
←IGF1
AC079907.1→
HELLPAR→
←AC010202.1
←RN7SL793P
←AC093023.1

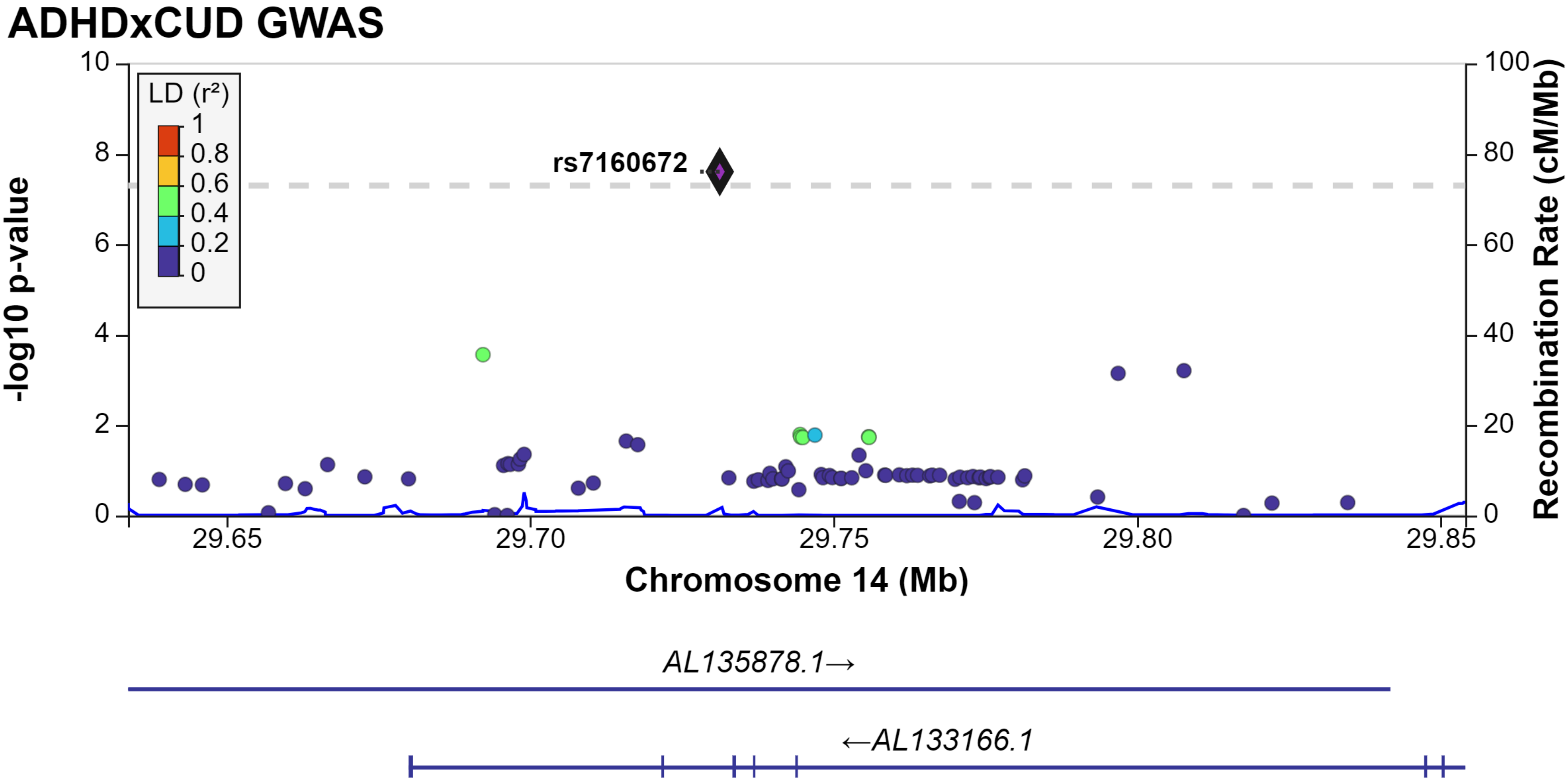

ADHDxCUD GWAS
LD (r²)
1
0.8
0.6
0.4
0.2
0
rs7160672
-log10 p-value
Recombination Rate (cM/Mb)
10
8
6
4
2
0
100
80
60
40
20
0
29.65
29.70
29.75
29.80
29.85
Chromosome 14 (Mb)
AL135878.1→
←AL133166.1

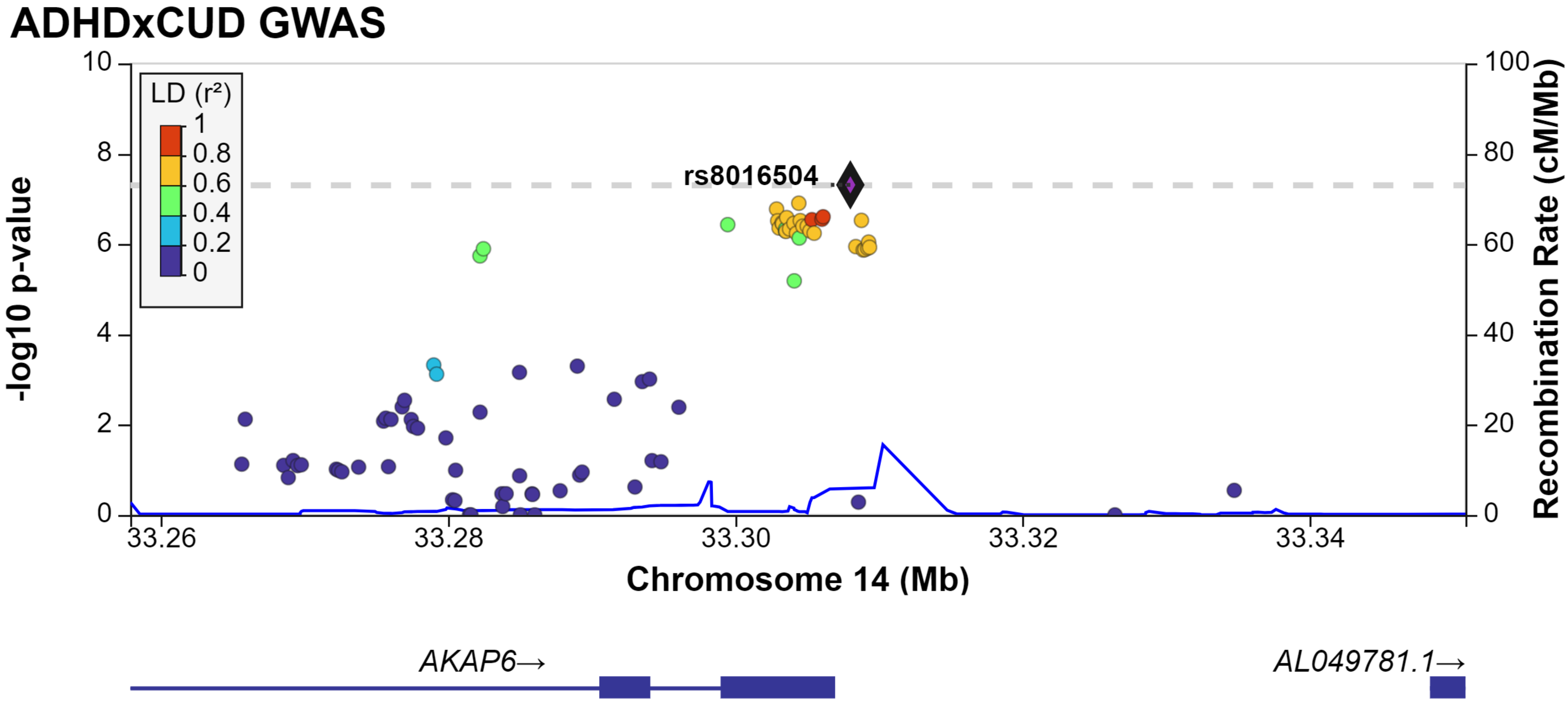
ADHDxCUD GWAS
LD (r²)
1
0.8
0.6
0.4
0.2
0
rs8016504
-log10 p-value
Recombination Rate (cM/Mb)
Chromosome 14 (Mb)
33.26
33.28
33.30
33.32
33.34
AKAP6→
AL049781.1→

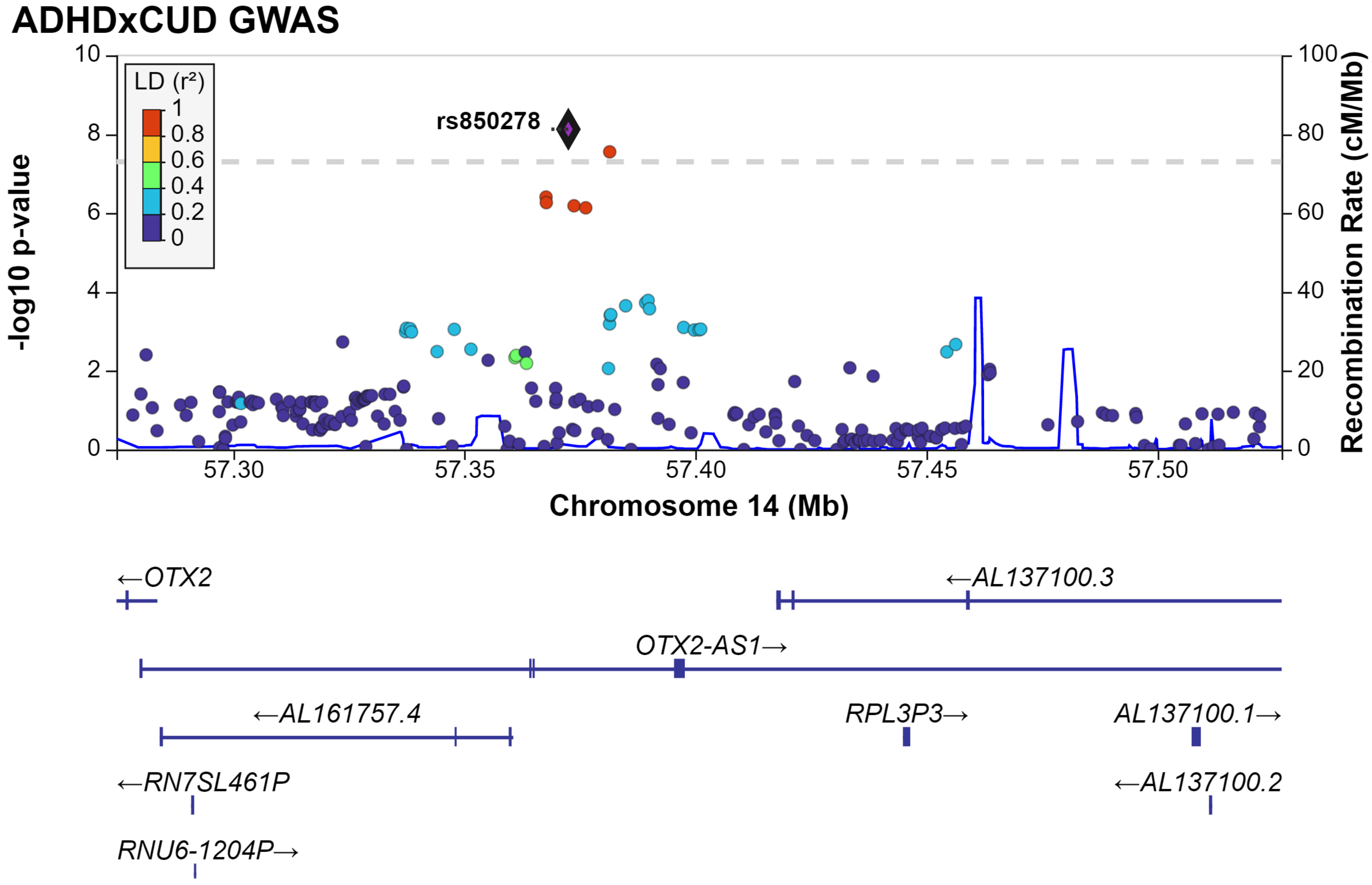
ADHDxCUD GWAS
LD (r²)
1
0.8
0.6
0.4
0.2
0
rs850278
-log10 p-value
Recombination Rate (cM/Mb)
10
8
6
4
2
0
100
80
60
40
20
0
57.30
57.35
57.40
57.45
57.50
Chromosome 14 (Mb)
←OTX2
←AL137100.3
OTX2-AS1→
←AL161757.4
RPL3P3→
AL137100.1→
←RN7SL461P
←AL137100.2
RNU6-1204P→

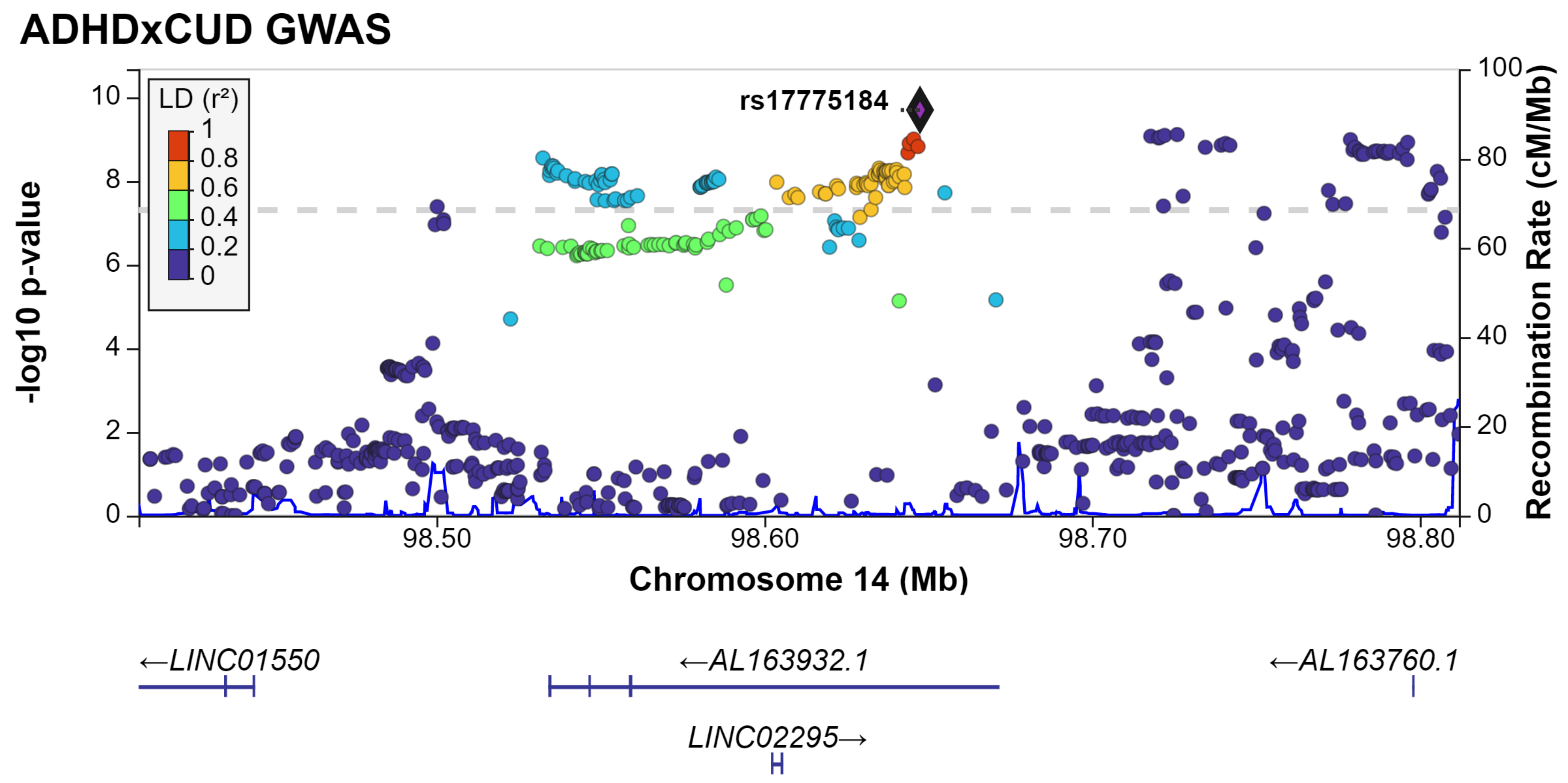
ADHDxCUD GWAS
rs17775184
LD (r²)
1
0.8
0.6
0.4
0.2
0
-log10 p-value
Recombination Rate (cM/Mb)
Chromosome 14 (Mb)
98.50
98.60
98.70
98.80
←LINC01550
←AL163932.1
←AL163760.1
LINC02295→

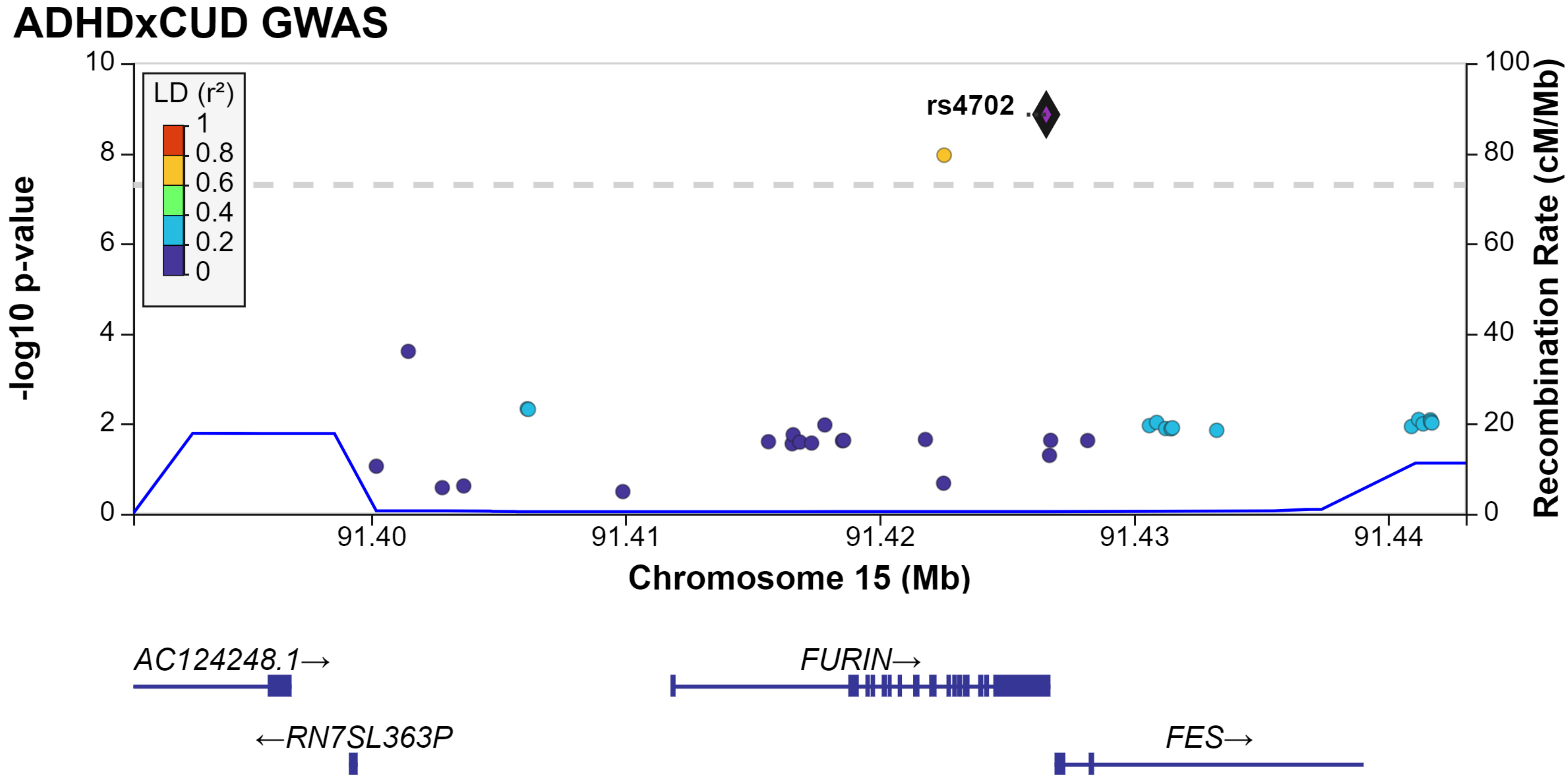
ADHDxCUD GWAS
rs4702
LD (r²)
1
0.8
0.6
0.4
0.2
0
-log10 p-value
Recombination Rate (cM/Mb)
Chromosome 15 (Mb)
91.40
91.41
91.42
91.43
91.44
AC124248.1→
FURIN→
←RN7SL363P
FES→

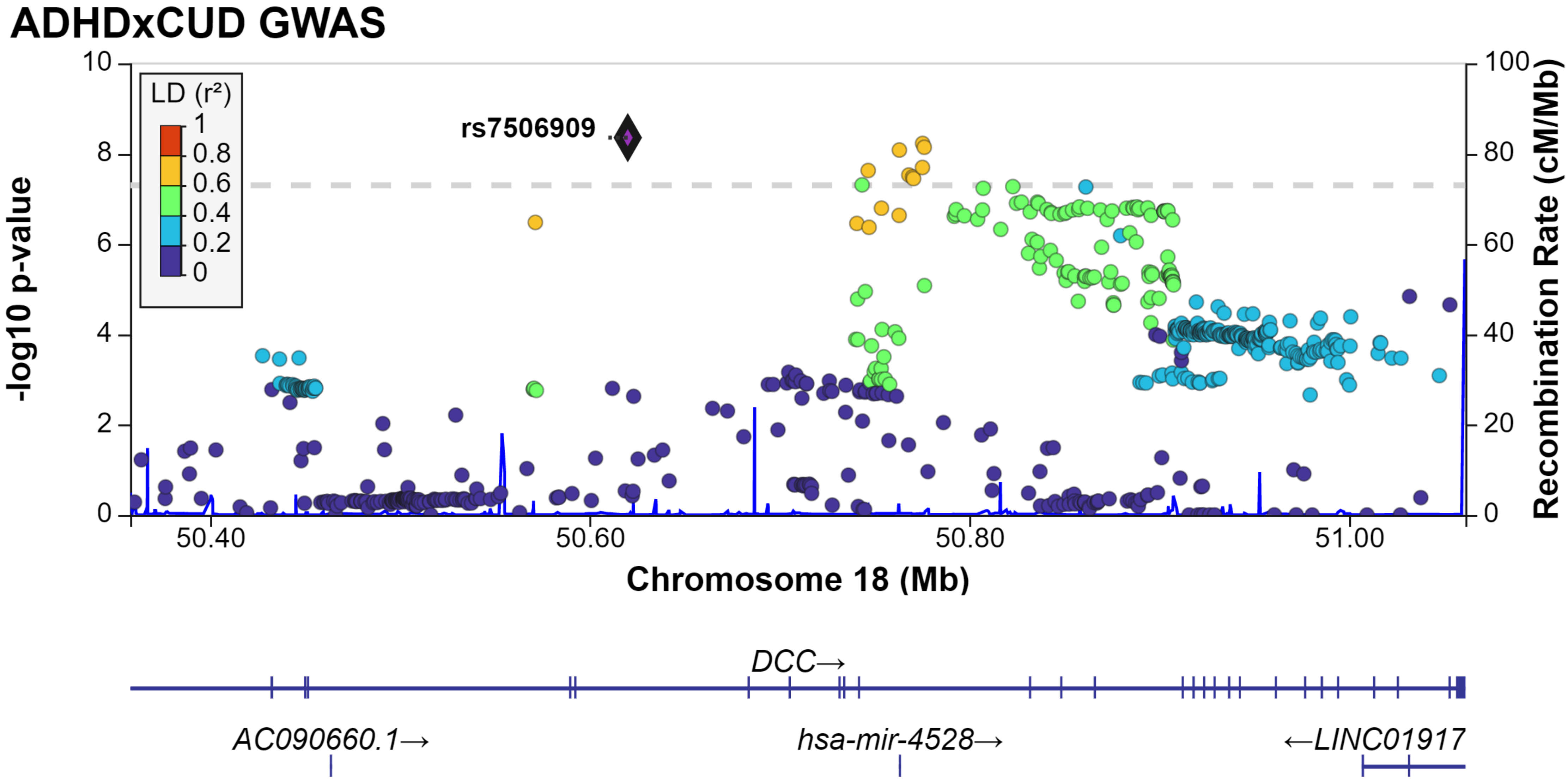

ADHDxCUD GWAS
LD (r²)
1
0.8
0.6
0.4
0.2
0
rs7506909
-log10 p-value
Recombination Rate (cM/Mb)
0
2
4
6
8
10
20
40
60
80
100
50.40
50.60
50.80
51.00
Chromosome 18 (Mb)
DCC→
AC090660.1→
hsa-mir-4528→
←LINC01917
