## Extended_Data2 for "Disentangling the shared genetics of ADHD, cannabis use disorder and cannabis use and prediction of cannabis use disorder in ADHD"

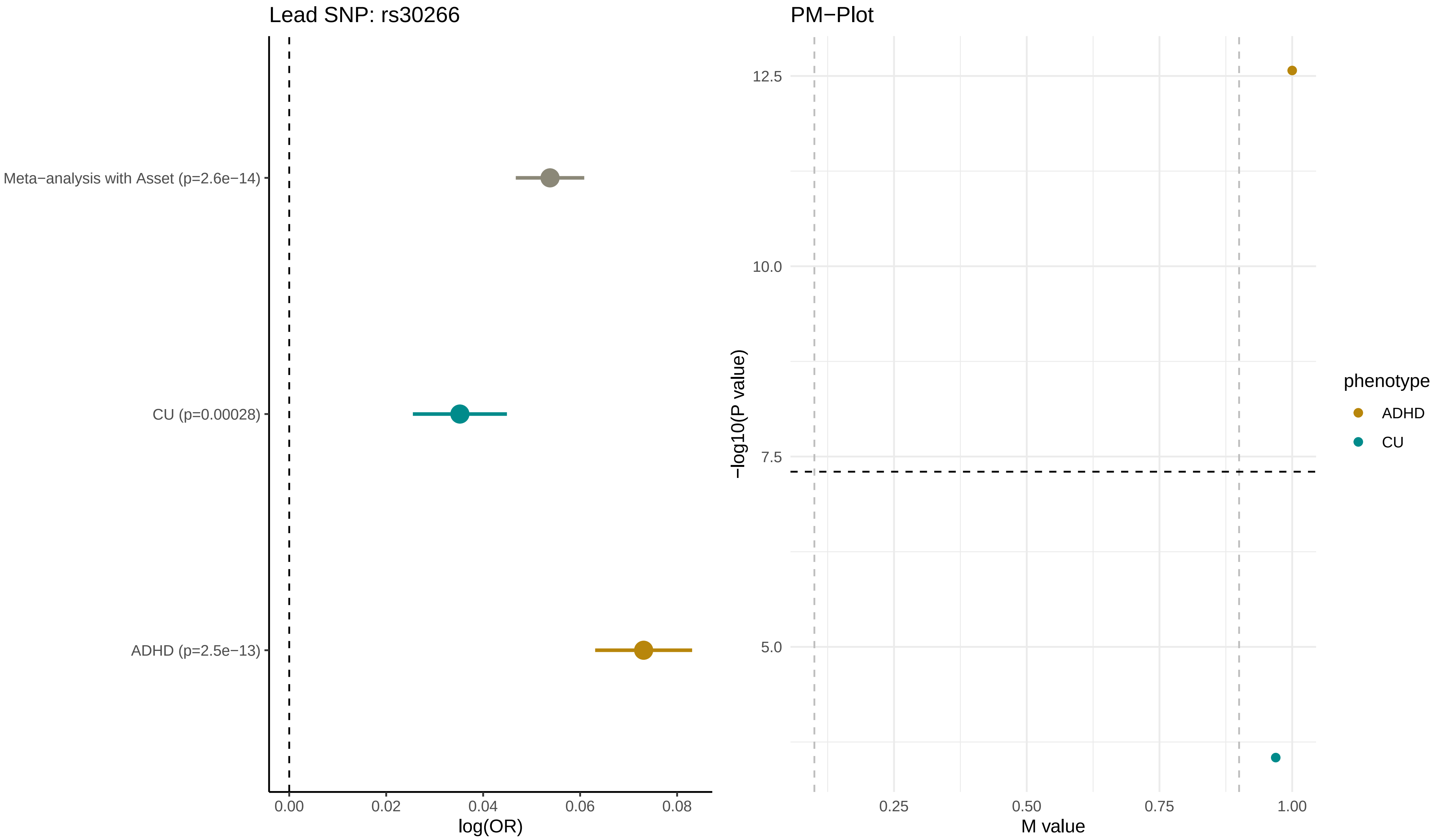
Lead SNP: rs30266
Meta−analysis with Asset (p=2.6e−14)
CU (p=0.00028)
ADHD (p=2.5e−13)
0.00
0.02
0.04
0.06
0.08
log(OR)
PM−Plot
12.5
10.0
7.5
5.0
−log10(P value)
0.25
0.50
0.75
1.00
M value
phenotype
ADHD
CU

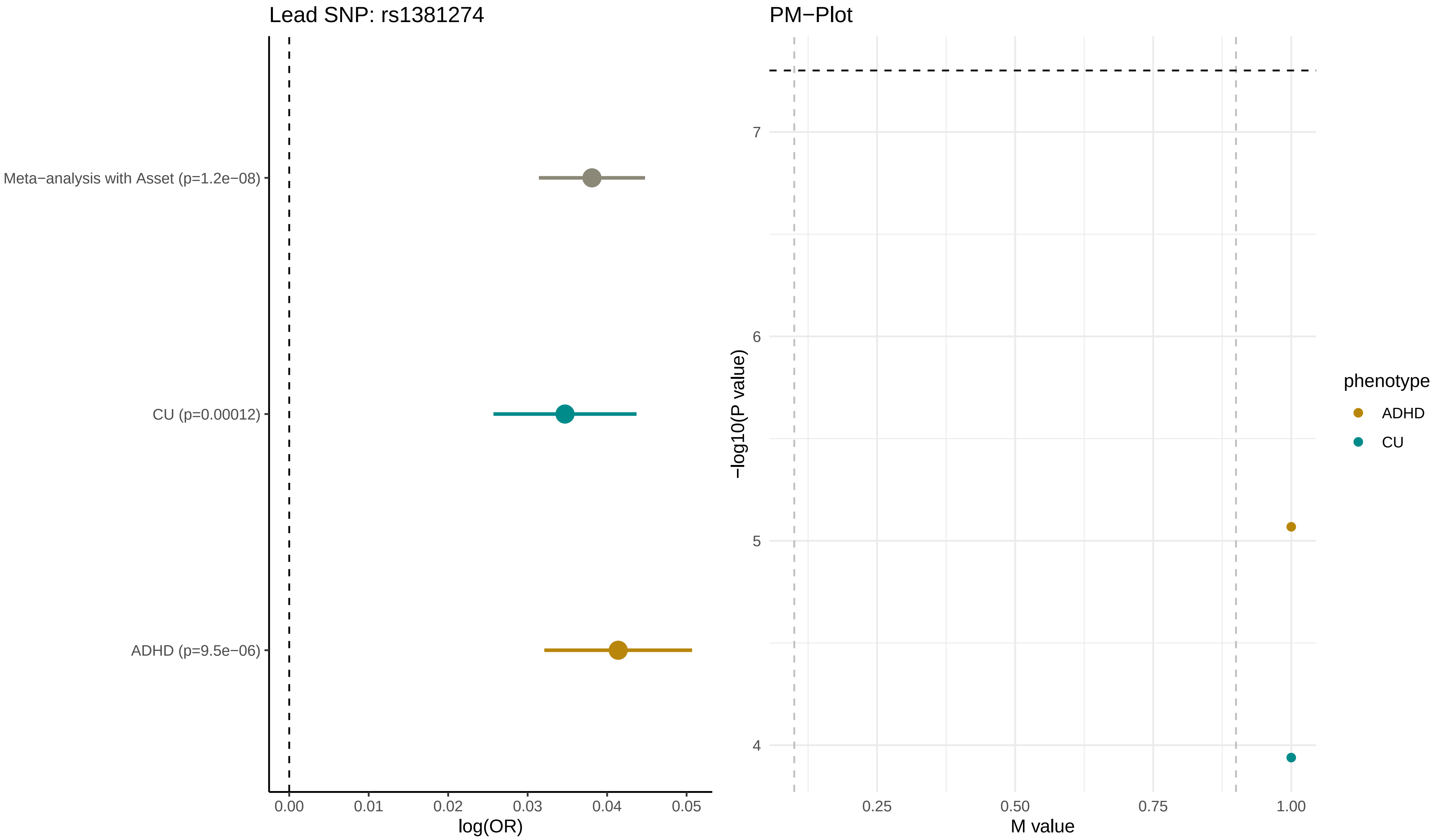

Lead SNP: rs1381274
Meta−analysis with Asset (p=1.2e−08)
CU (p=0.00012)
ADHD (p=9.5e−06)
0.00
0.01
0.02
0.03
0.04
0.05
log(OR)
PM−Plot
−log10(P value)
7
6
5
4
0.25
0.50
0.75
1.00
M value
phenotype
ADHD
CU

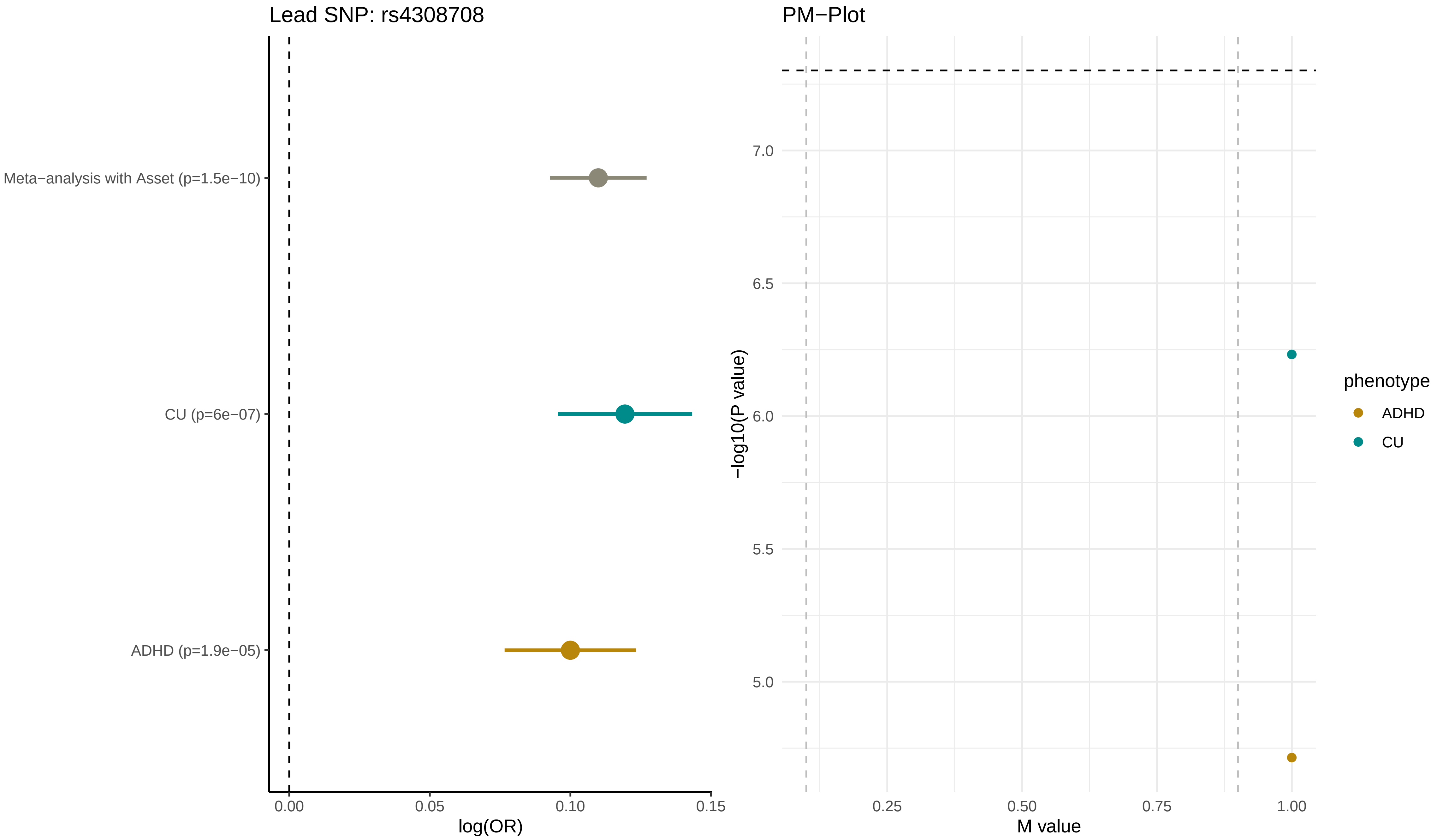
Lead SNP: rs4308708
Meta−analysis with Asset (p=1.5e−10)
CU (p=6e−07)
ADHD (p=1.9e−05)
0.00
0.05
0.10
0.15
log(OR)
PM−Plot
7.0
6.5
6.0
5.5
5.0
−log10(P value)
0.25
0.50
0.75
1.00
M value
phenotype
ADHD
CU

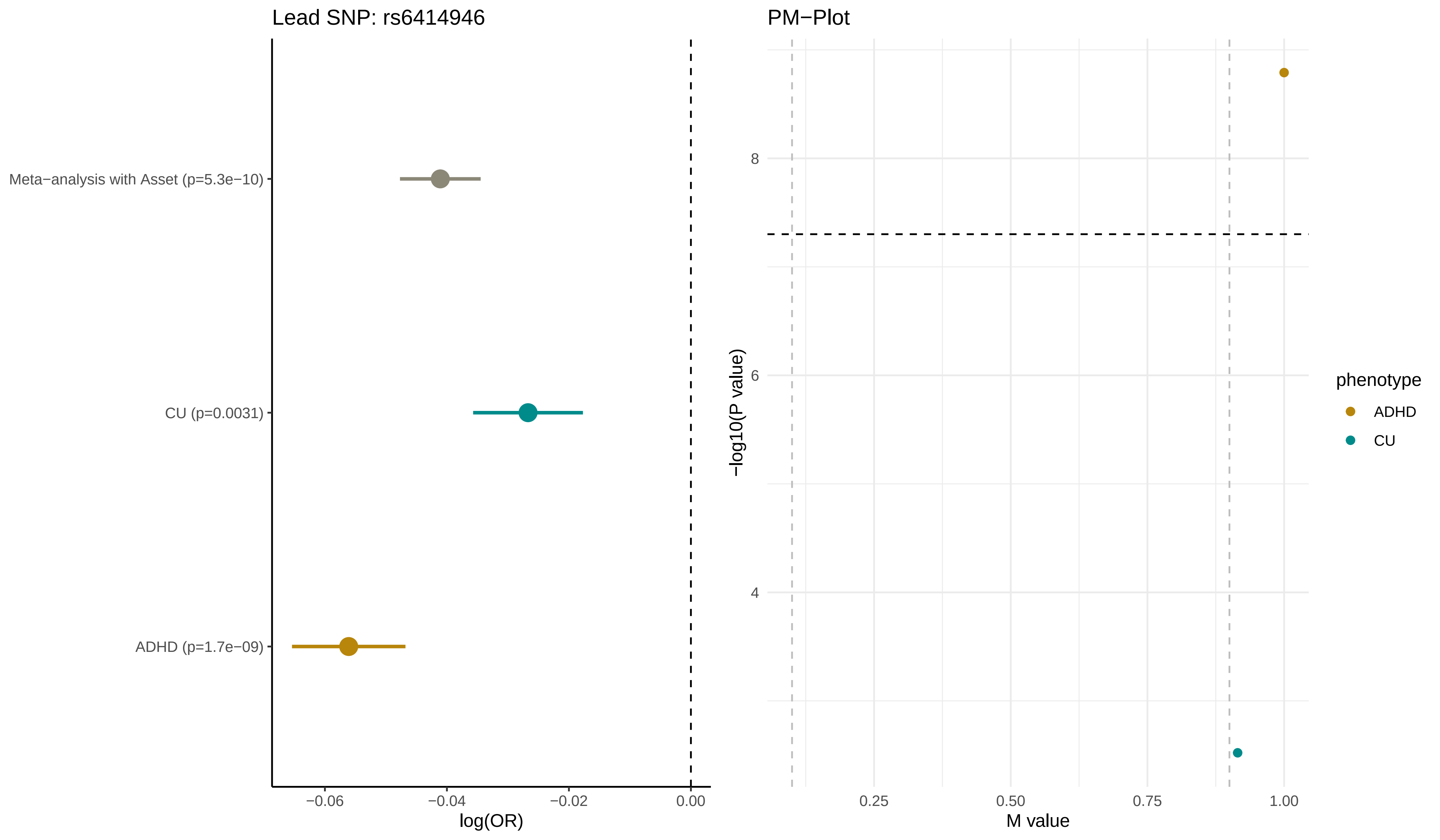
Lead SNP: rs6414946
Meta−analysis with Asset (p=5.3e−10)
CU (p=0.0031)
ADHD (p=1.7e−09)
−0.06
−0.04
−0.02
0.00
log(OR)
PM−Plot
8
6
4
−log10(P value)
0.25
0.50
0.75
1.00
M value
phenotype
ADHD
CU

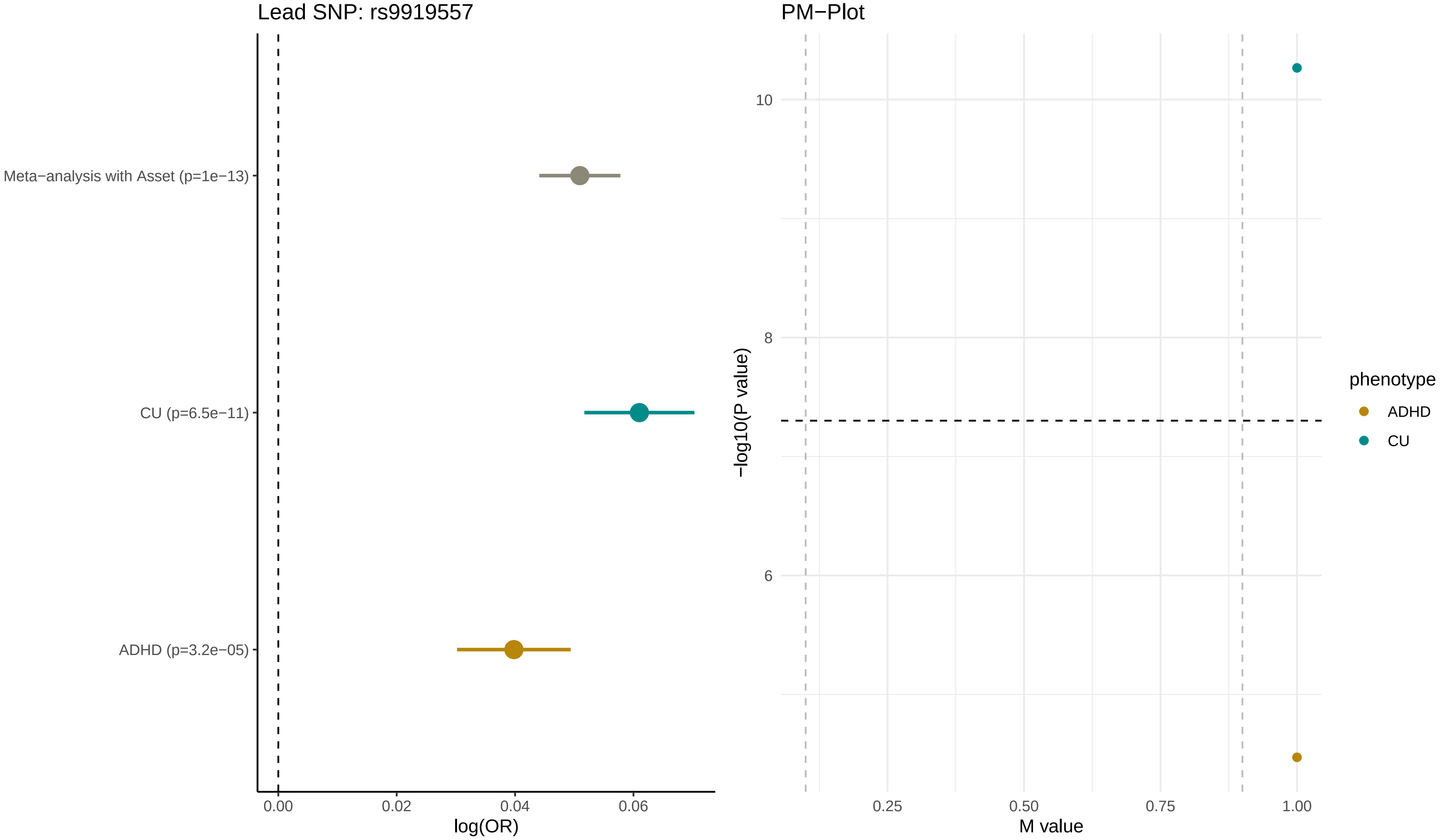

Lead SNP: rs9919557
Meta−analysis with Asset (p=1e−13)
CU (p=6.5e−11)
ADHD (p=3.2e−05)
0.00
0.02
0.04
0.06
log(OR)
PM−Plot
10
8
6
−log10(P value)
0.25
0.50
0.75
1.00
M value
phenotype
ADHD
CU

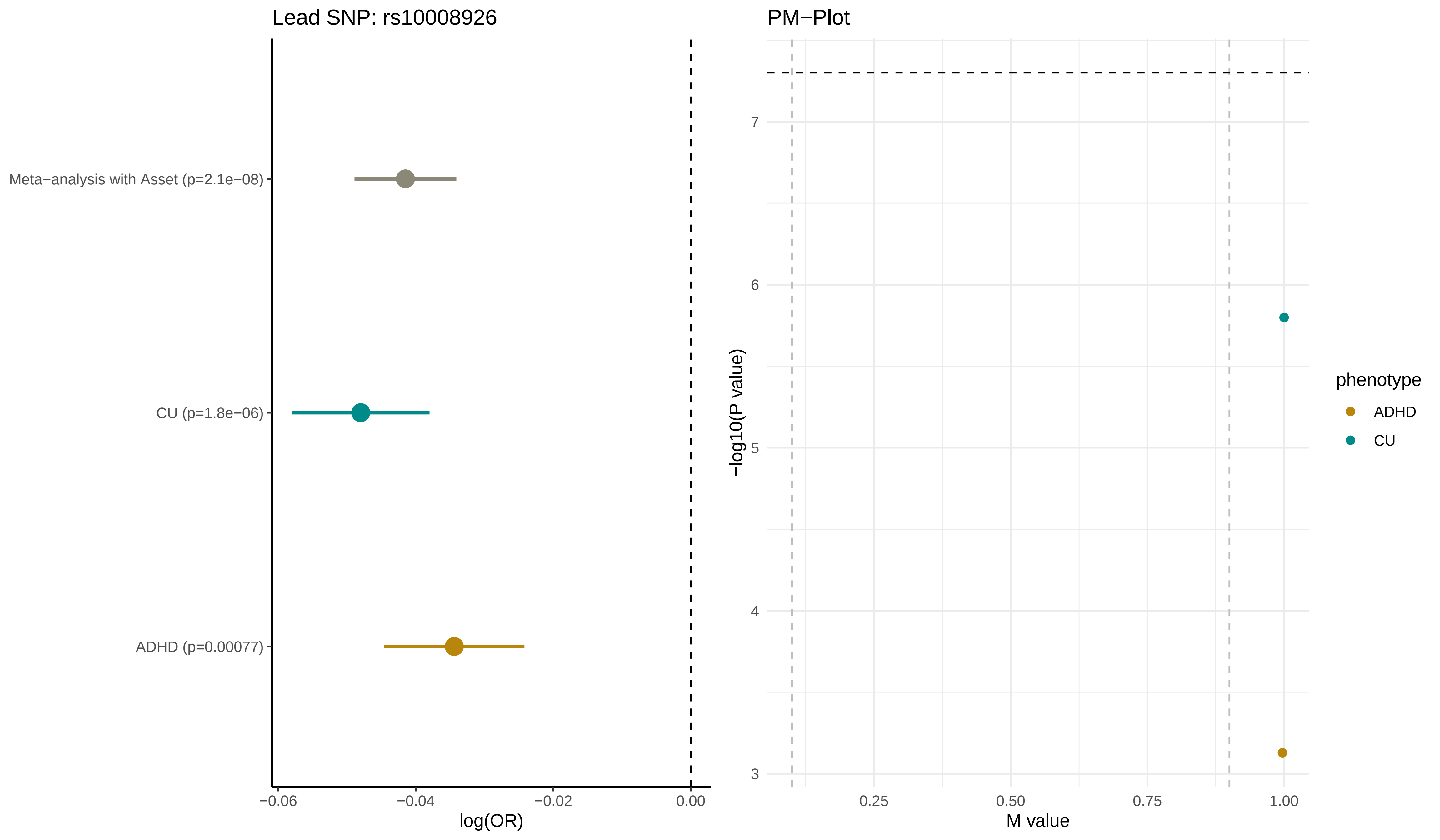

Lead SNP: rs10008926
Meta−analysis with Asset (p=2.1e−08)
CU (p=1.8e−06)
ADHD (p=0.00077)
−0.06
−0.04
−0.02
0.00
log(OR)
PM−Plot
7
6
5
4
3
−log10(P value)
0.25
0.50
0.75
1.00
M value
phenotype
ADHD
CU

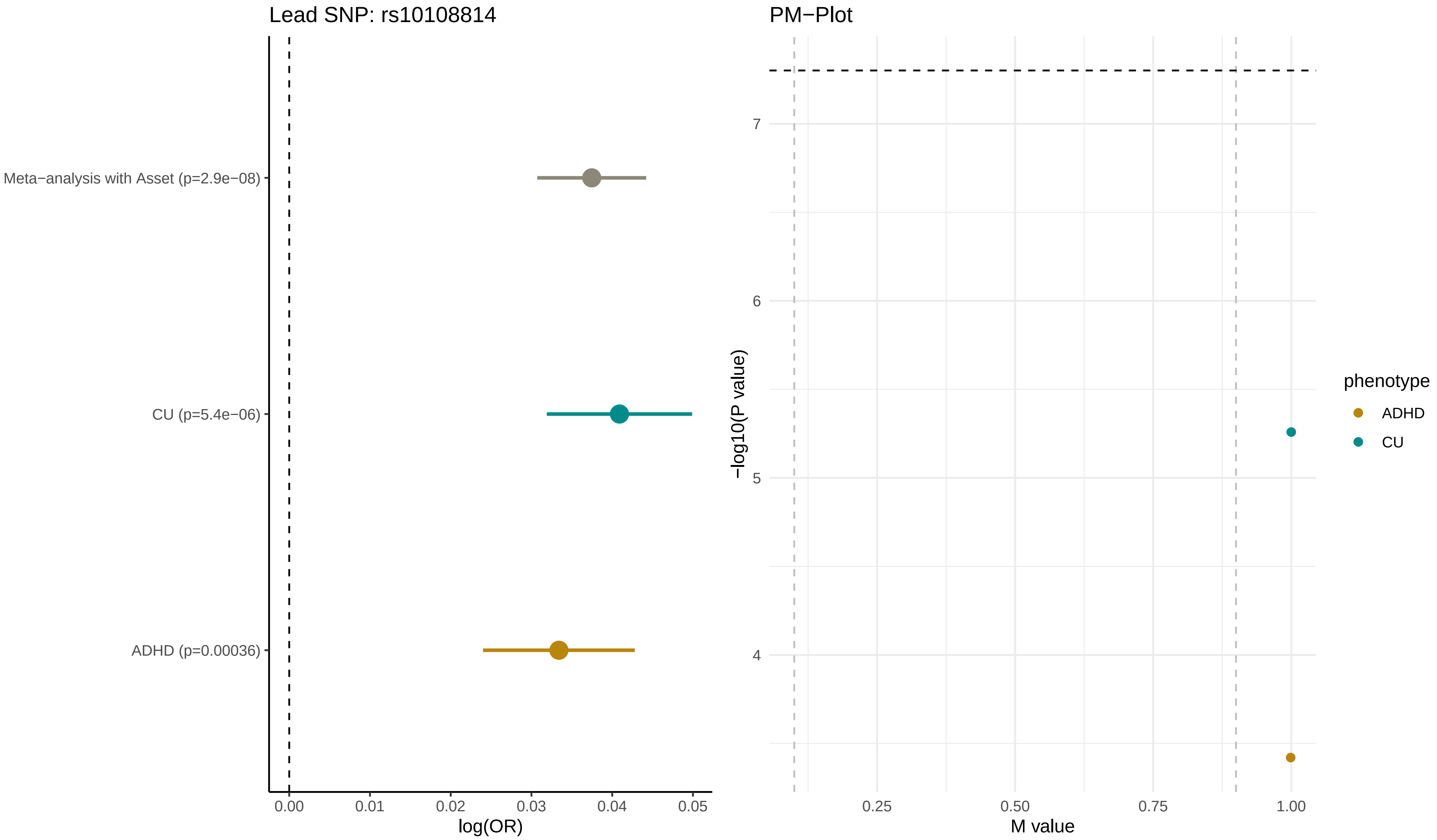

Lead SNP: rs10108814
Meta−analysis with Asset (p=2.9e−08)
CU (p=5.4e−06)
ADHD (p=0.00036)
0.00
0.01
0.02
0.03
0.04
0.05
log(OR)
PM−Plot
−log10(P value)
7
6
5
4
0.25
0.50
0.75
1.00
M value
phenotype
ADHD
CU

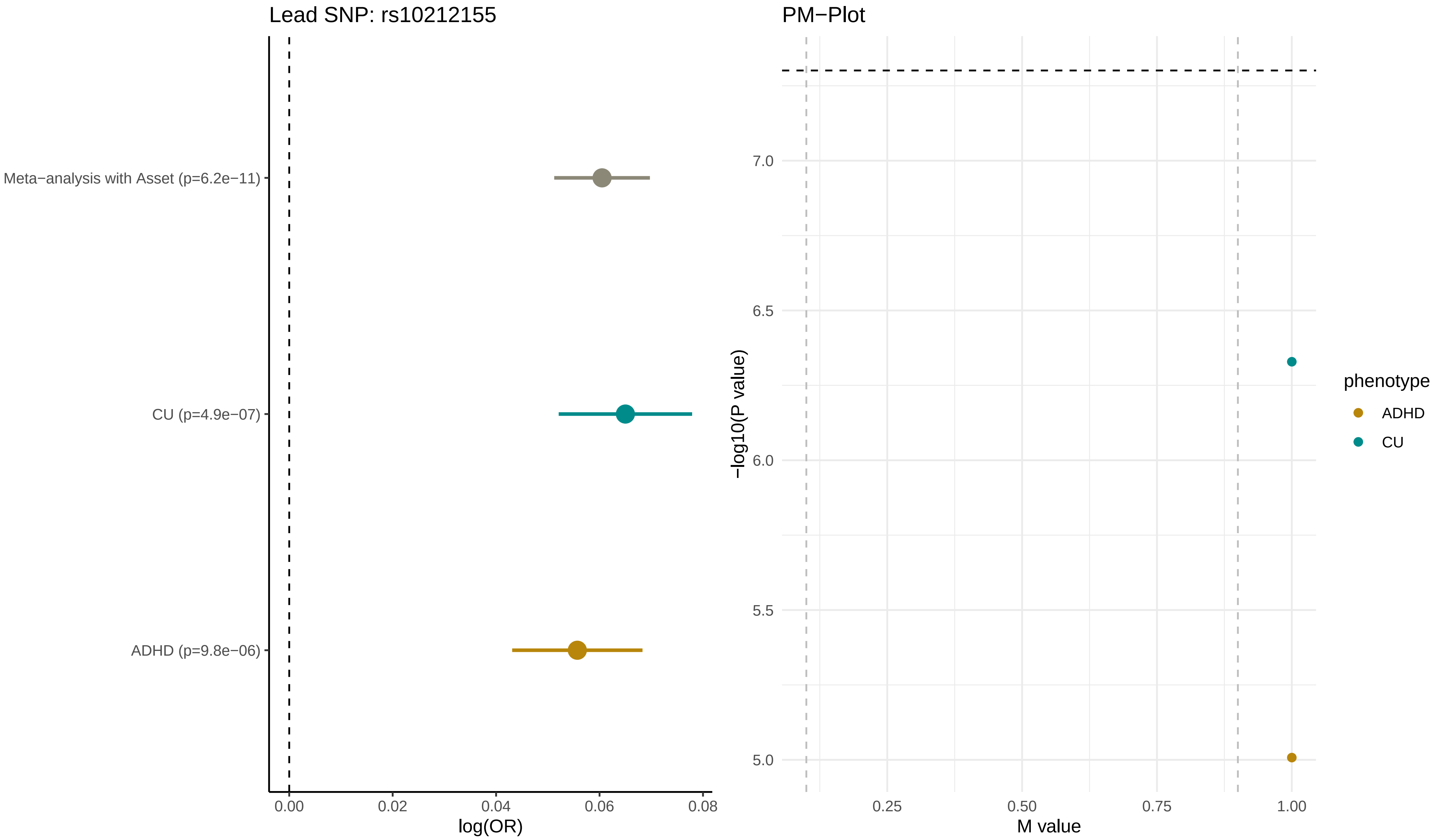

Lead SNP: rs10212155
Meta−analysis with Asset (p=6.2e−11)
CU (p=4.9e−07)
ADHD (p=9.8e−06)
0.00
0.02
0.04
0.06
0.08
log(OR)
PM−Plot
7.0
6.5
6.0
5.5
5.0
−log10(P value)
0.25
0.50
0.75
1.00
M value
phenotype
ADHD
CU

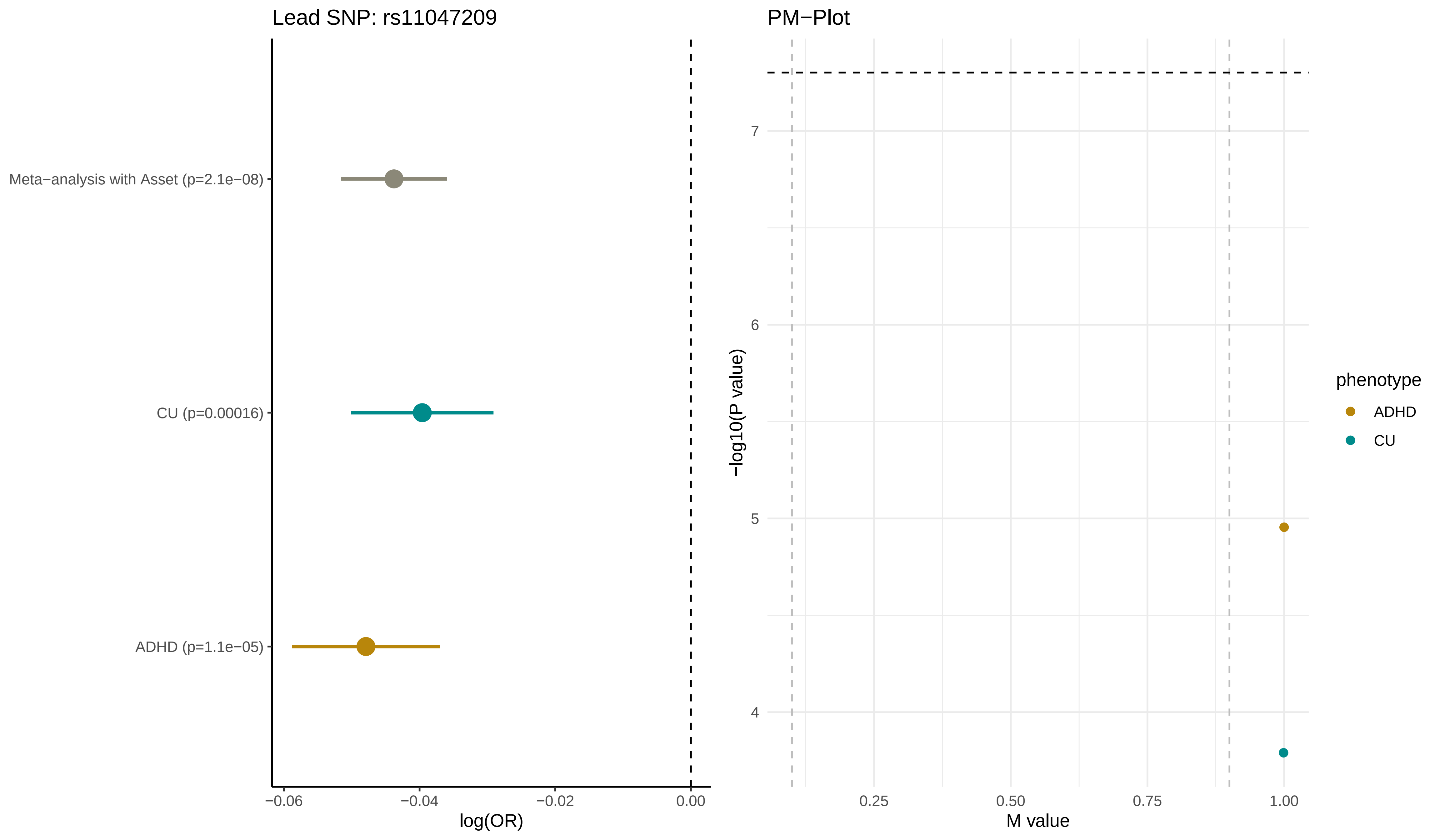

Lead SNP: rs11047209
Meta−analysis with Asset (p=2.1e−08)
CU (p=0.00016)
ADHD (p=1.1e−05)
−0.06
−0.04
−0.02
0.00
log(OR)
PM−Plot
7
6
5
4
−log10(P value)
0.25
0.50
0.75
1.00
M value
phenotype
ADHD
CU

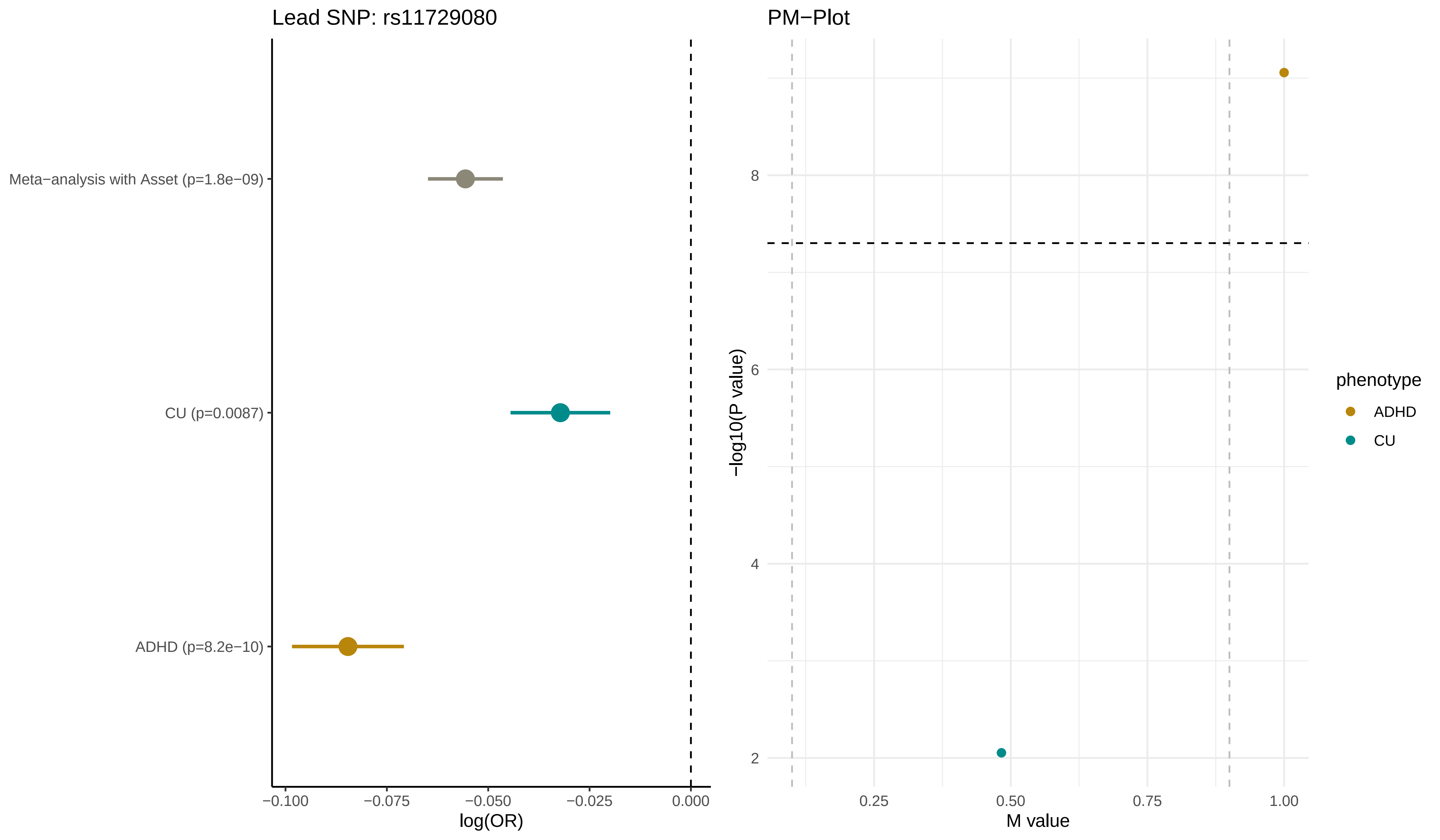

Lead SNP: rs11729080
Meta−analysis with Asset (p=1.8e−09)
CU (p=0.0087)
ADHD (p=8.2e−10)
−0.100
−0.075
−0.050
−0.025
0.000
log(OR)
PM−Plot
8
6
4
2
−log10(P value)
0.25
0.50
0.75
1.00
M value
phenotype
ADHD
CU

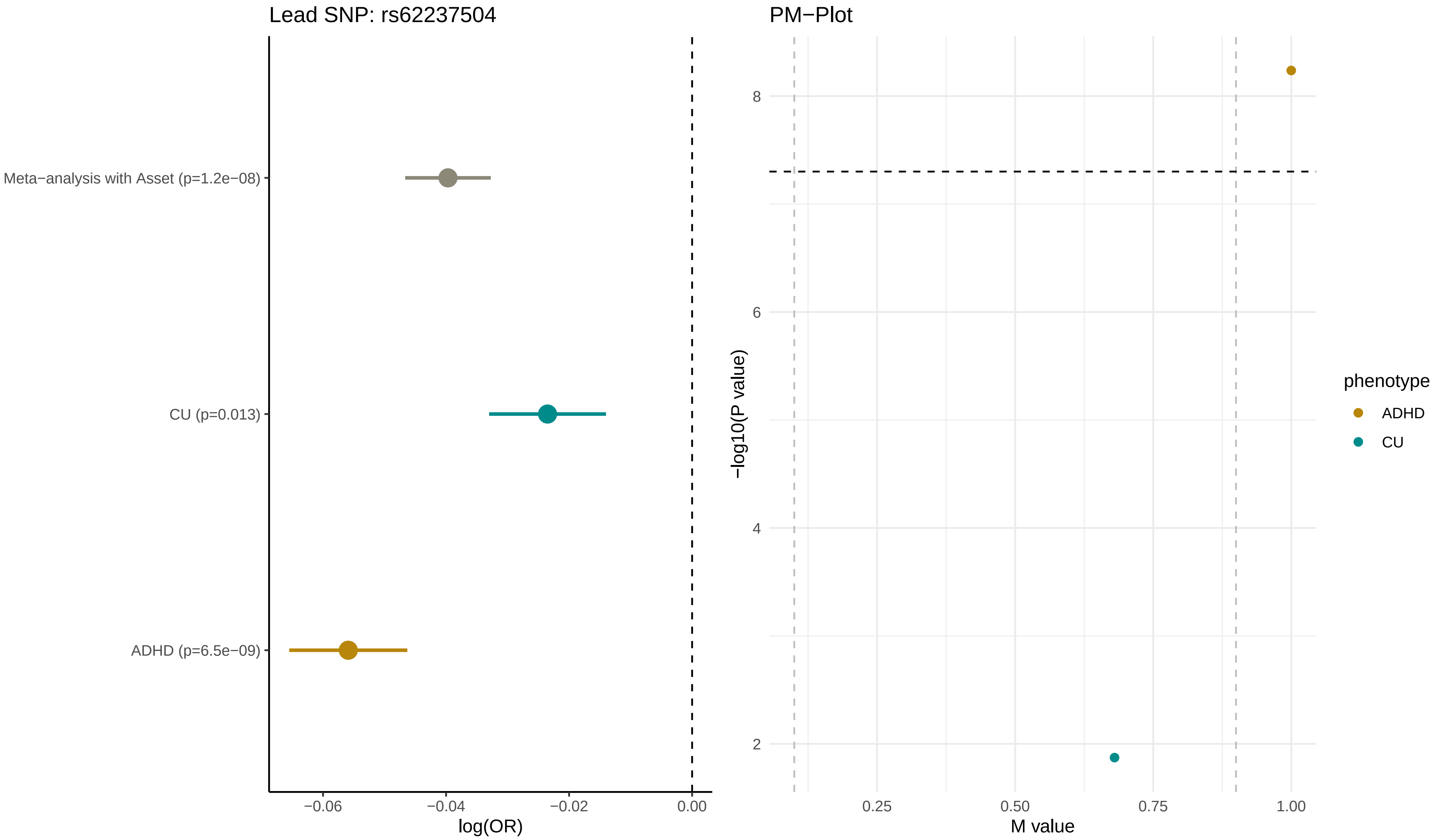

Lead SNP: rs62237504
Meta−analysis with Asset (p=1.2e−08)
CU (p=0.013)
ADHD (p=6.5e−09)
−0.06
−0.04
−0.02
0.00
log(OR)
PM−Plot
8
6
4
2
−log10(P value)
0.25
0.50
0.75
1.00
M value
phenotype
ADHD
CU

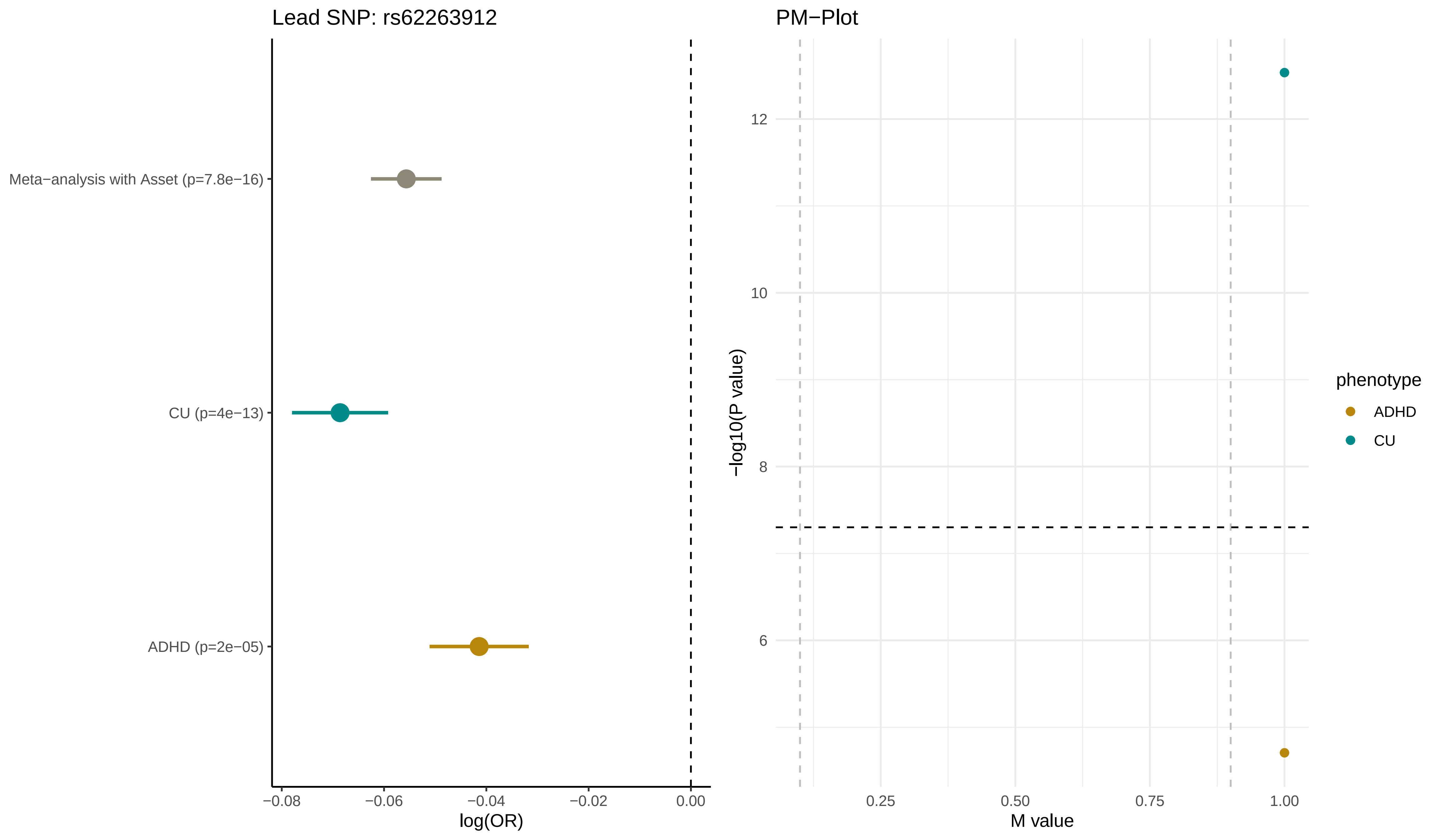

Lead SNP: rs62263912
Meta−analysis with Asset (p=7.8e−16)
CU (p=4e−13)
ADHD (p=2e−05)
−0.08
−0.06
−0.04
−0.02
0.00
log(OR)
PM−Plot
12
10
8
6
−log10(P value)
0.25
0.50
0.75
1.00
M value
phenotype
ADHD
CU

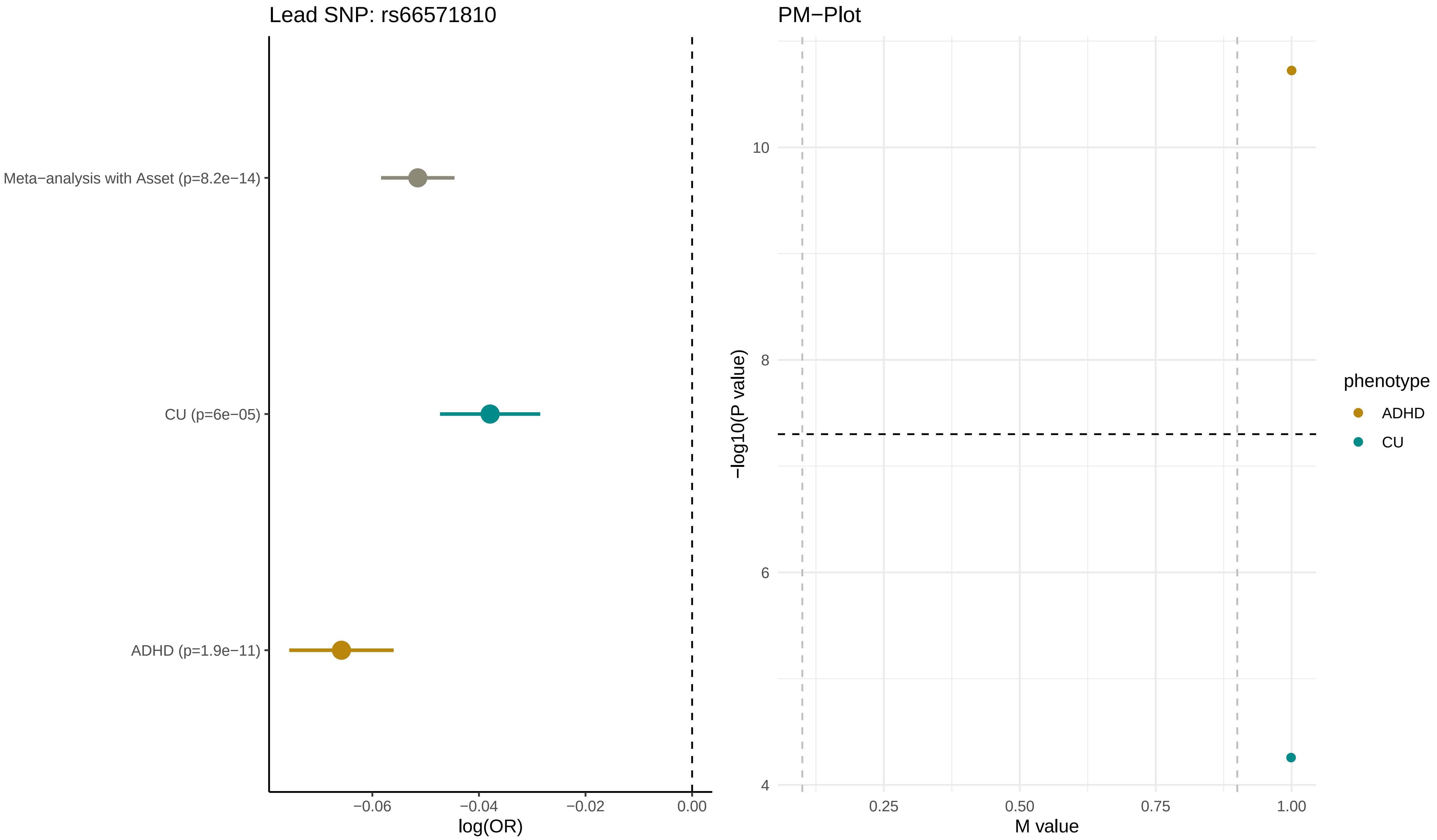

Lead SNP: rs66571810
Meta−analysis with Asset (p=8.2e−14)
CU (p=6e−05)
ADHD (p=1.9e−11)
−0.06
−0.04
−0.02
0.00
log(OR)
PM−Plot
−log10(P value)
10
8
6
4
0.25
0.50
0.75
1.00
M value
phenotype
ADHD
CU

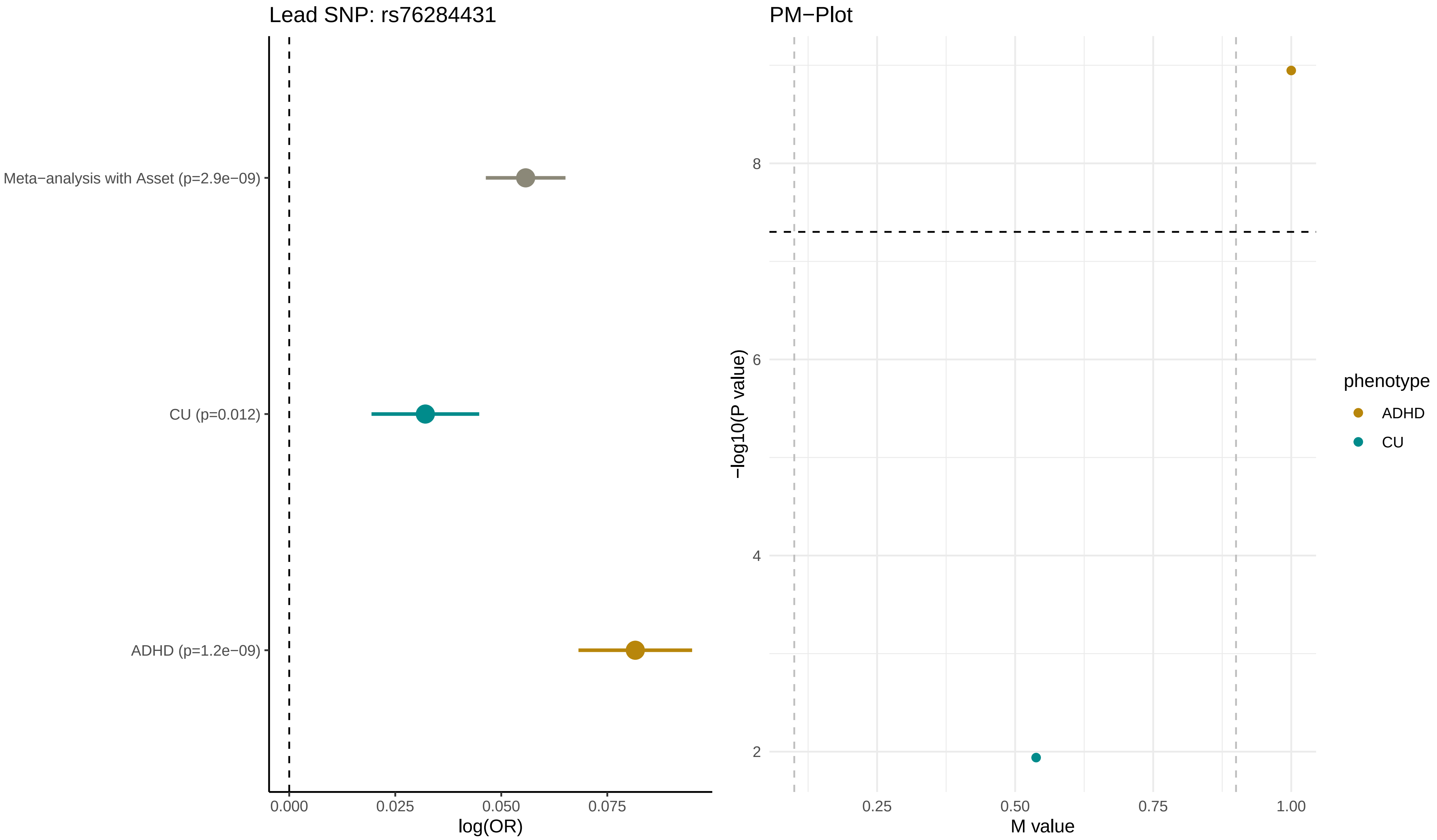

Lead SNP: rs76284431
Meta−analysis with Asset (p=2.9e−09)
CU (p=0.012)
ADHD (p=1.2e−09)
0.000
0.025
0.050
0.075
log(OR)
PM−Plot
−log10(P value)
8
6
4
2
0.25
0.50
0.75
1.00
M value
phenotype
ADHD
CU

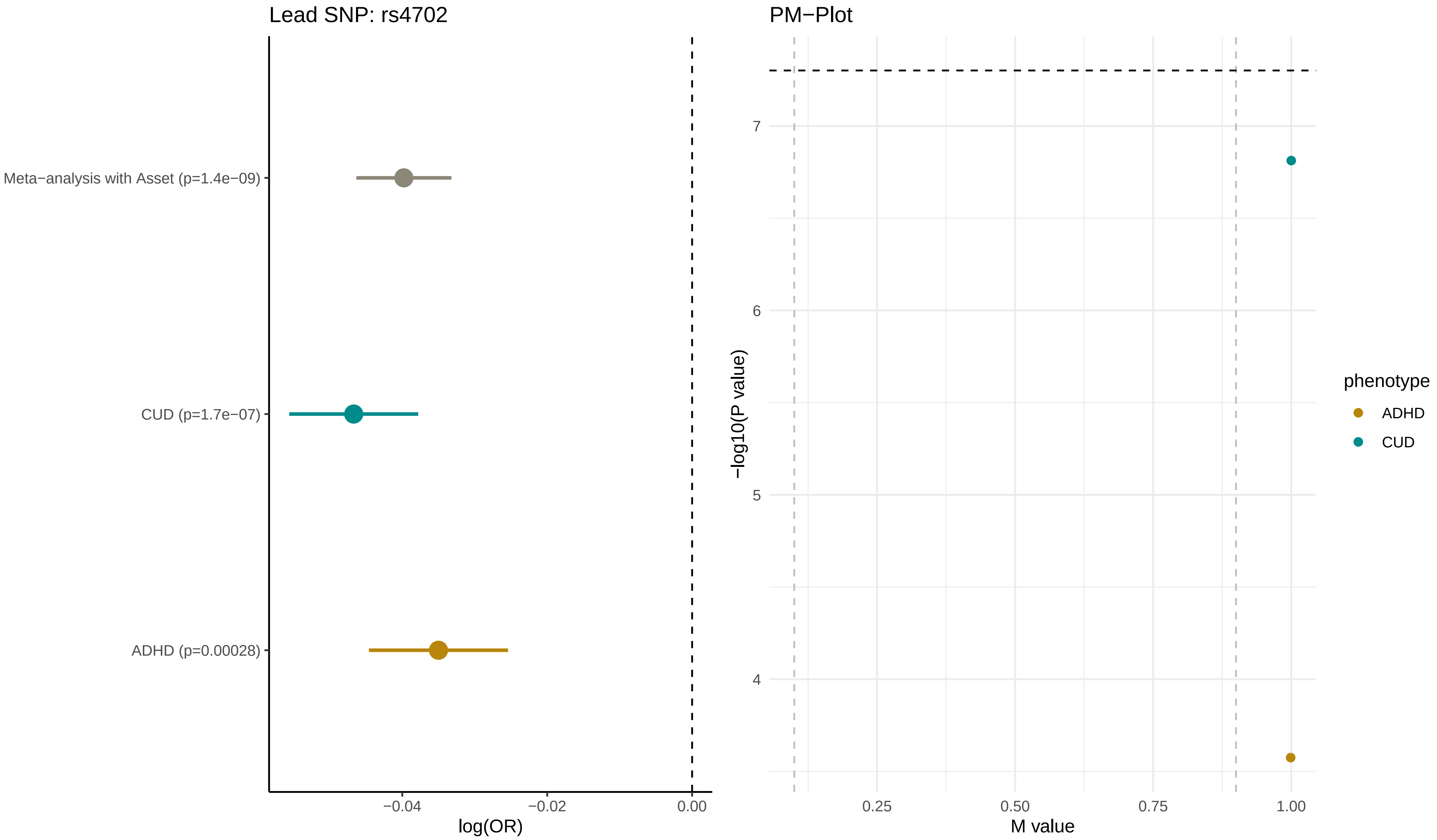
Lead SNP: rs4702
Meta−analysis with Asset (p=1.4e−09)
CUD (p=1.7e−07)
ADHD (p=0.00028)
−0.04
−0.02
0.00
log(OR)
PM−Plot
7
6
5
4
−log10(P value)
0.25
0.50
0.75
1.00
M value
phenotype
ADHD
CUD

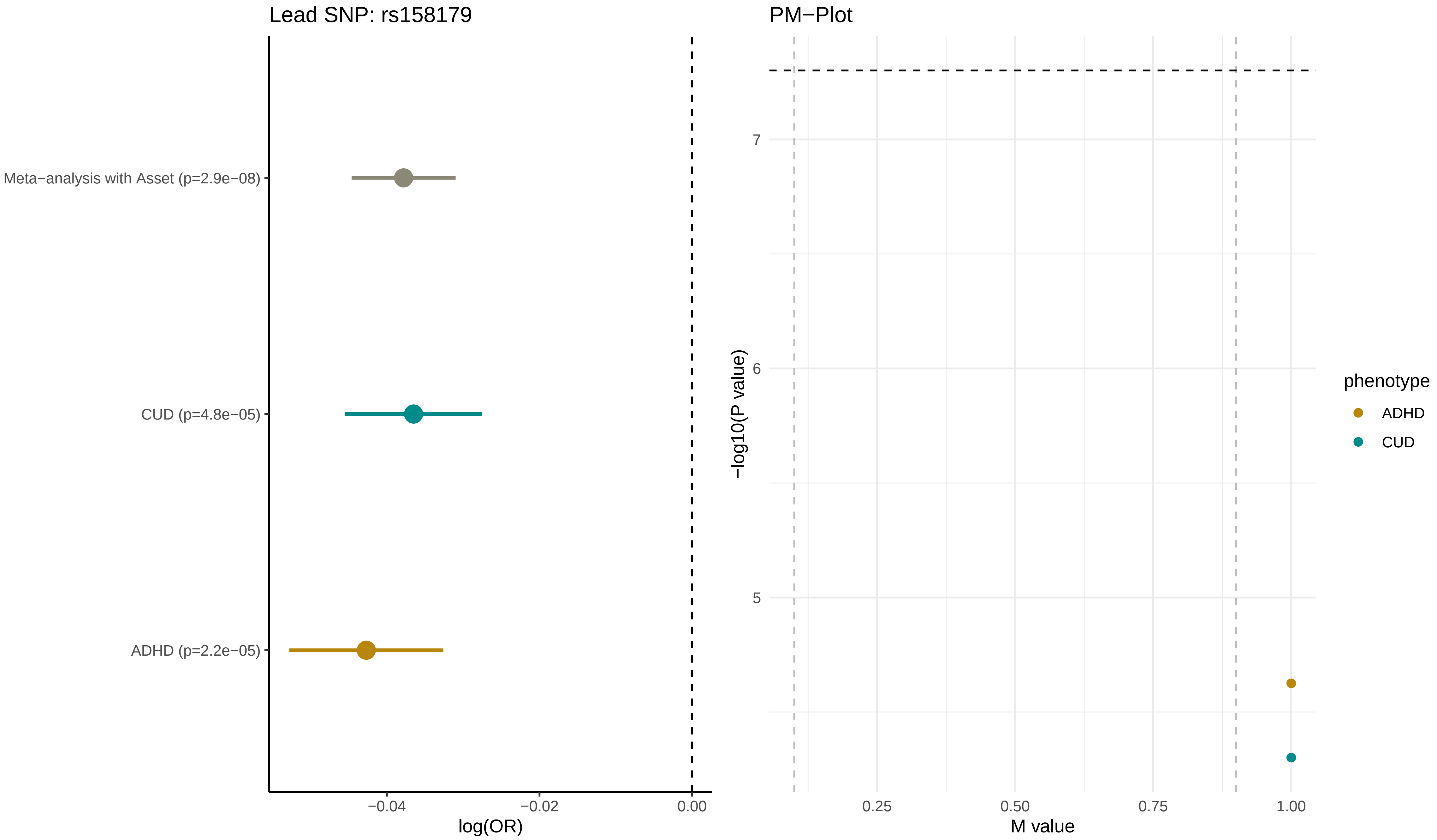

Lead SNP: rs158179
Meta−analysis with Asset (p=2.9e−08)
CUD (p=4.8e−05)
ADHD (p=2.2e−05)
−0.04
−0.02
0.00
log(OR)
PM−Plot
7
6
5
−log10(P value)
0.25
0.50
0.75
1.00
M value
phenotype
ADHD
CUD

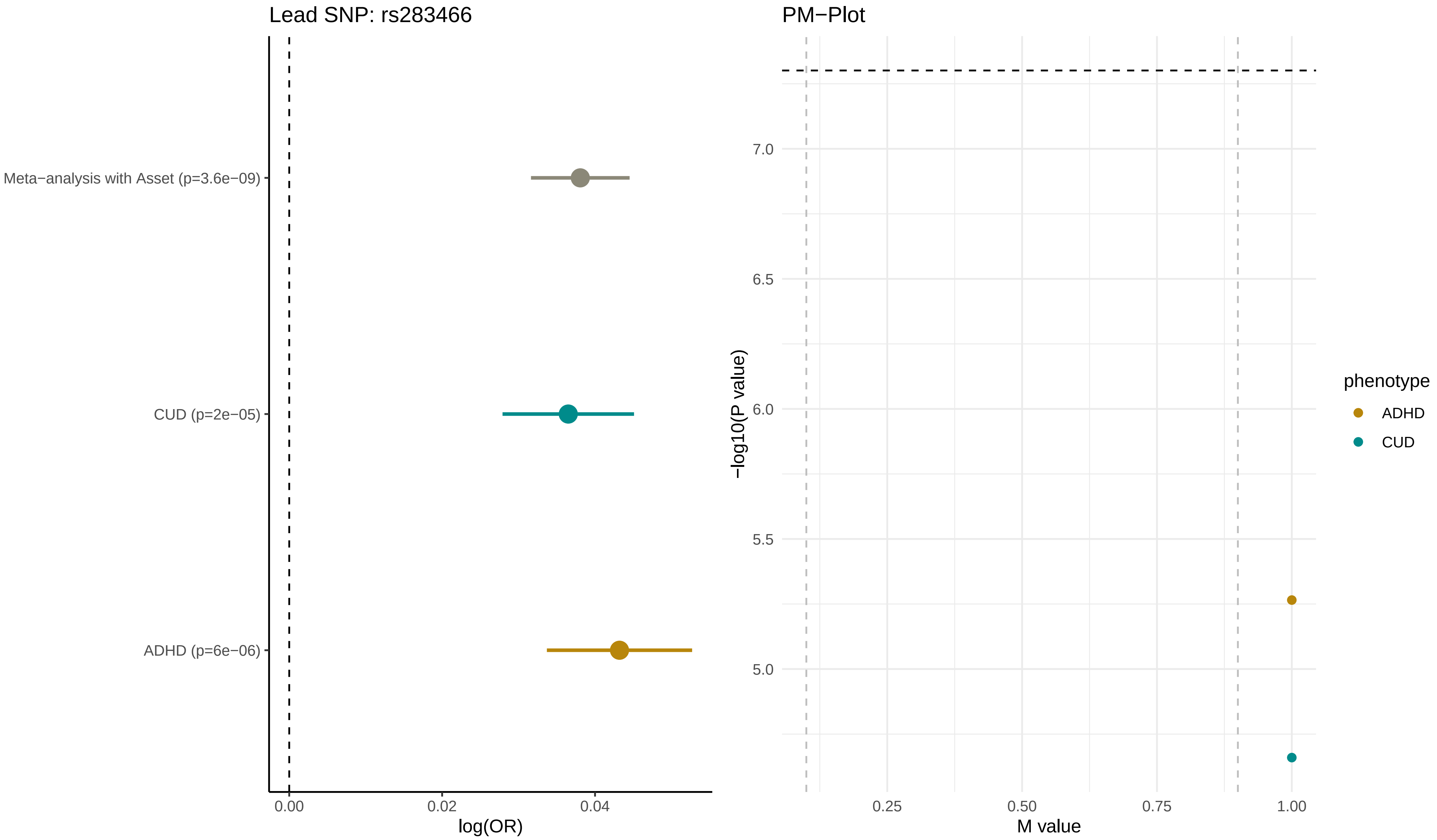
Lead SNP: rs283466
Meta−analysis with Asset (p=3.6e−09)
CUD (p=2e−05)
ADHD (p=6e−06)
0.00
0.02
0.04
log(OR)
PM−Plot
7.0
6.5
6.0
5.5
5.0
−log10(P value)
0.25
0.50
0.75
1.00
M value
phenotype
ADHD
CUD

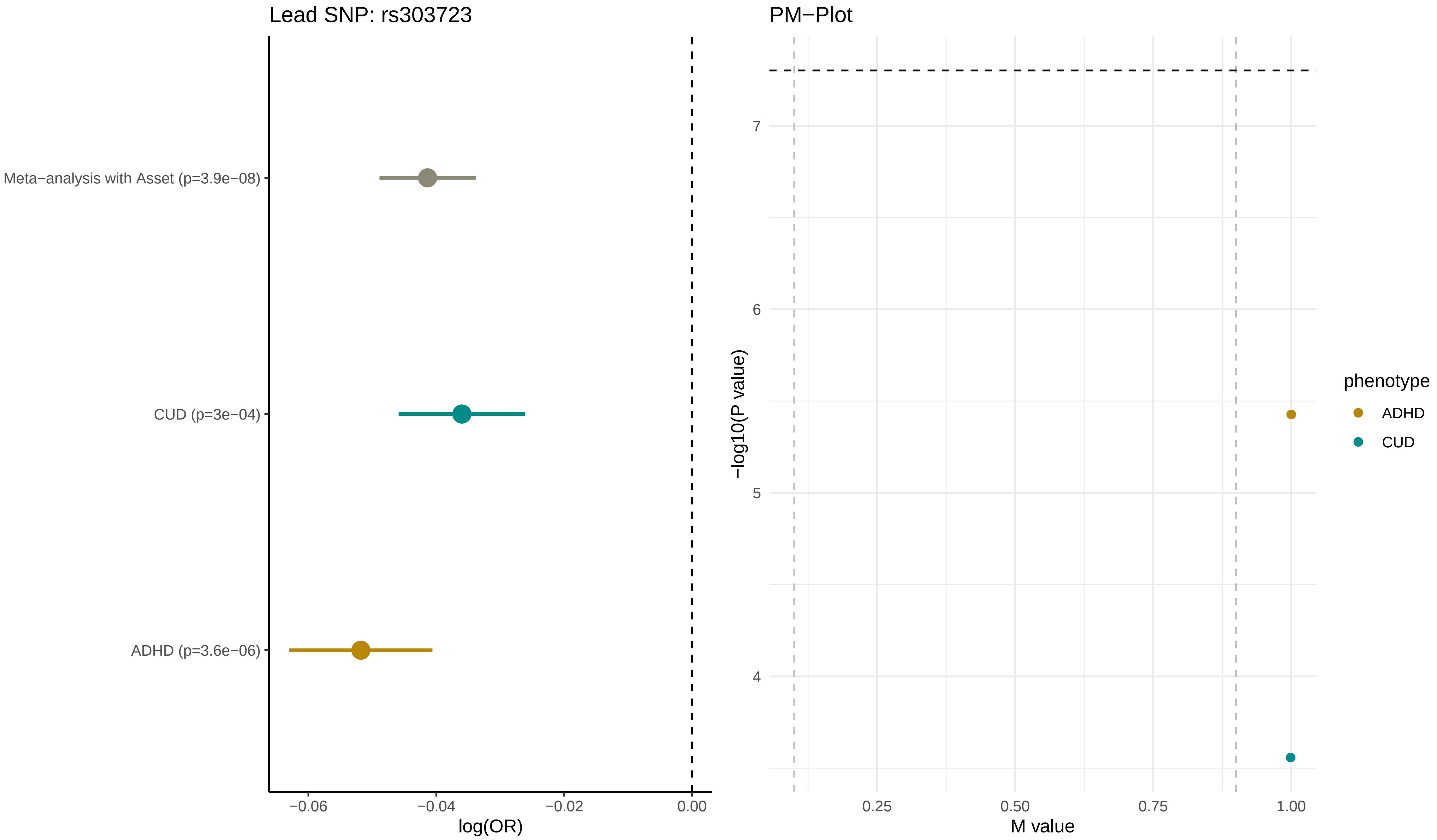

Lead SNP: rs303723
Meta−analysis with Asset (p=3.9e−08)
CUD (p=3e−04)
ADHD (p=3.6e−06)
−0.06
−0.04
−0.02
0.00
log(OR)
PM−Plot
−log10(P value)
7
6
5
4
0.25
0.50
0.75
1.00
M value
phenotype
ADHD
CUD

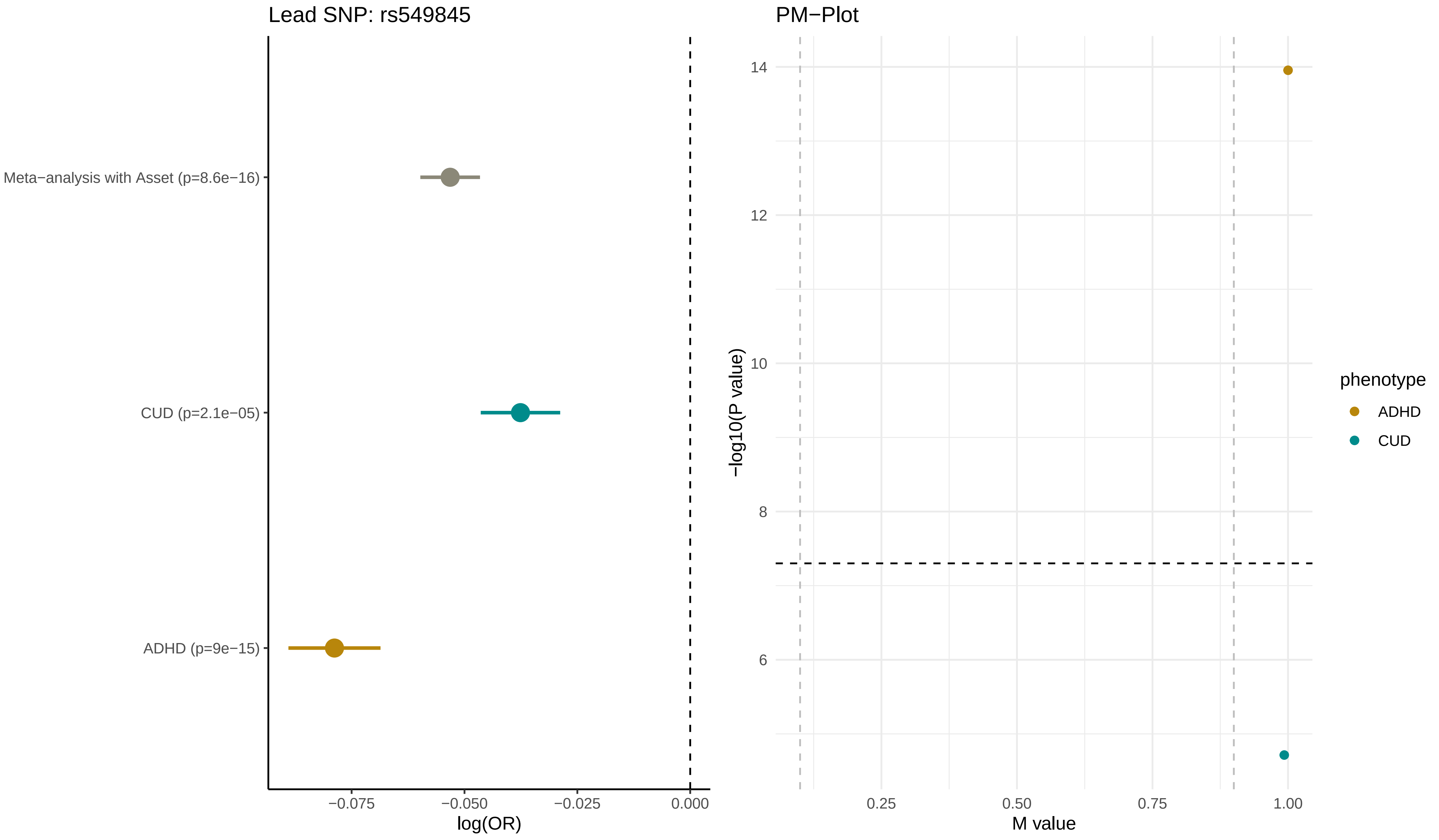

Lead SNP: rs549845
Meta−analysis with Asset (p=8.6e−16)
CUD (p=2.1e−05)
ADHD (p=9e−15)
−0.075
−0.050
−0.025
0.000
log(OR)
PM−Plot
14
12
10
8
6
−log10(P value)
0.25
0.50
0.75
1.00
M value
phenotype
ADHD
CUD

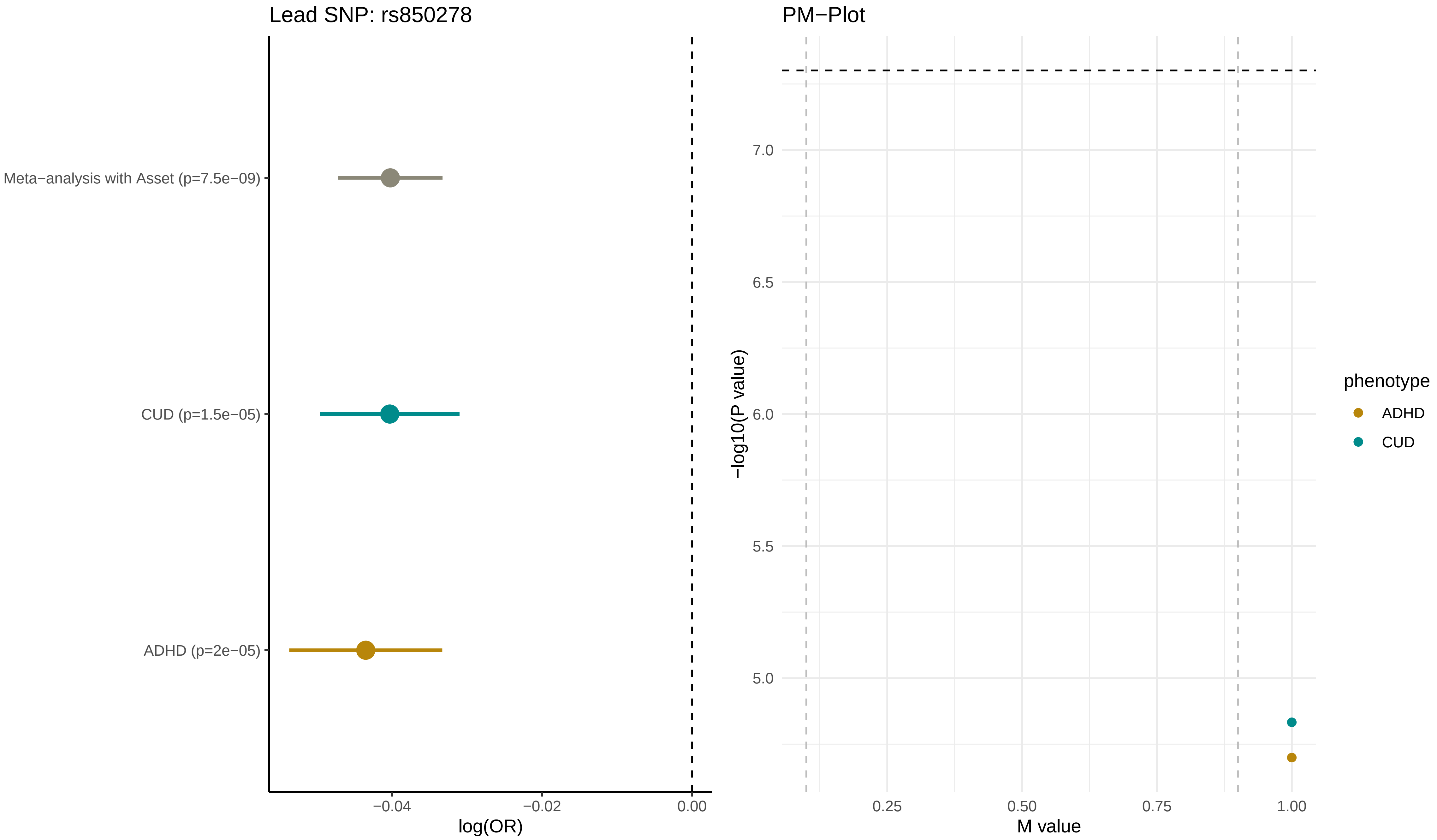

Lead SNP: rs850278
Meta−analysis with Asset (p=7.5e−09)
CUD (p=1.5e−05)
ADHD (p=2e−05)
−0.04
−0.02
0.00
log(OR)
PM−Plot
7.0
6.5
6.0
5.5
5.0
−log10(P value)
0.25
0.50
0.75
1.00
M value
phenotype
ADHD
CUD

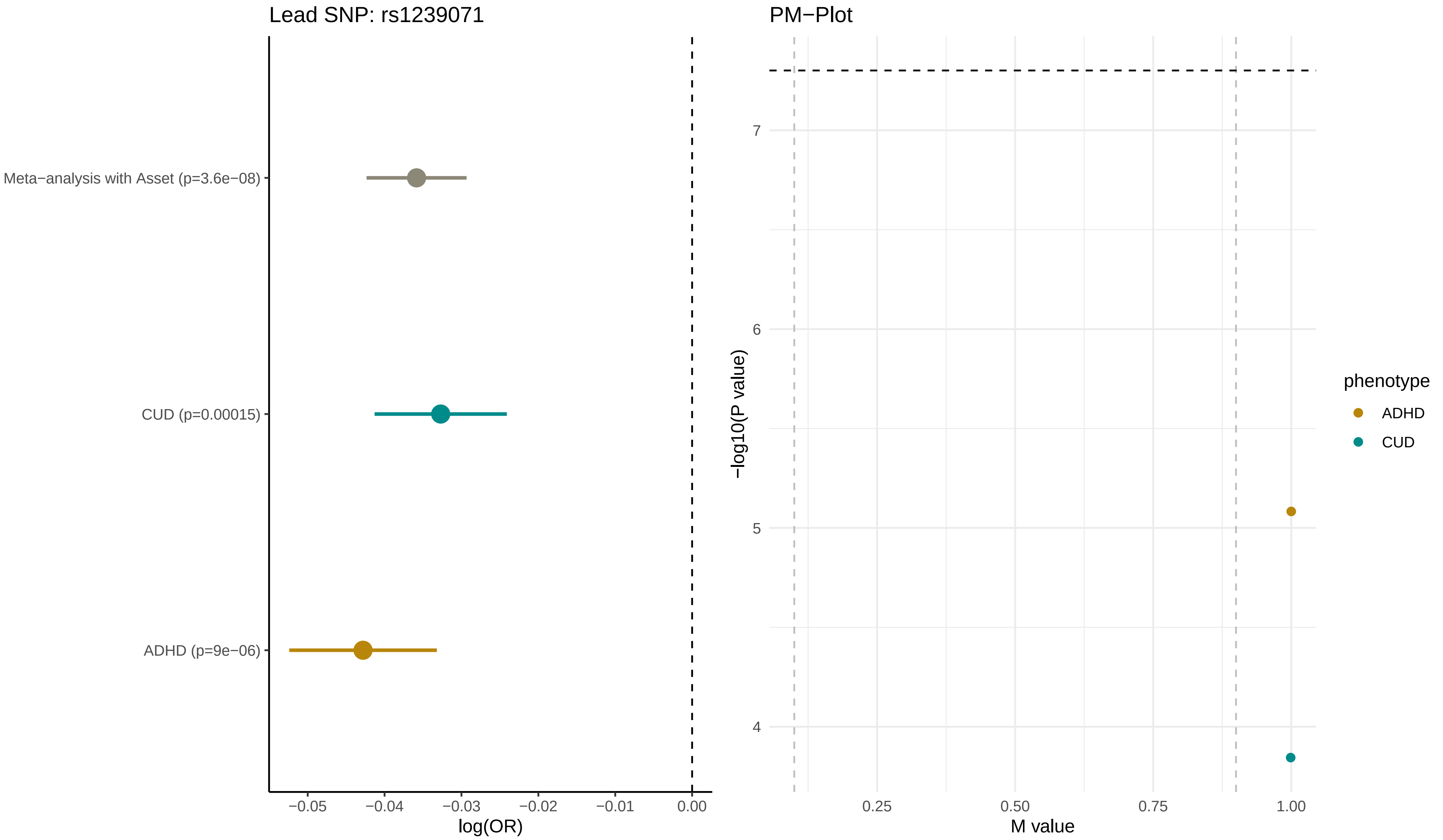

Lead SNP: rs1239071
Meta−analysis with Asset (p=3.6e−08)
CUD (p=0.00015)
ADHD (p=9e−06)
−0.05
−0.04
−0.03
−0.02
−0.01
0.00
log(OR)
PM−Plot
7
6
5
4
−log10(P value)
0.25
0.50
0.75
1.00
M value
phenotype
ADHD
CUD

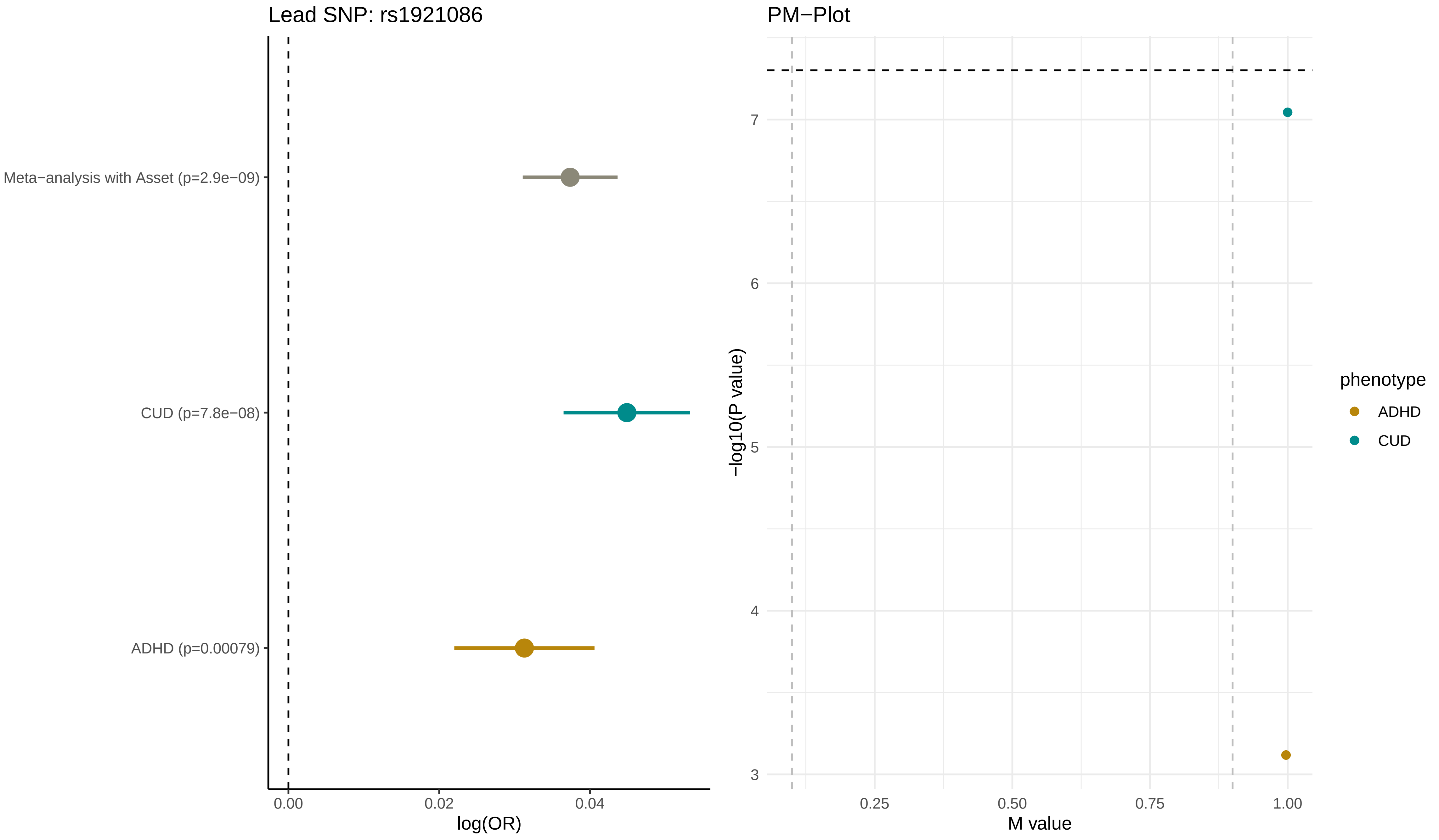
Lead SNP: rs1921086
Meta−analysis with Asset (p=2.9e−09)
CUD (p=7.8e−08)
ADHD (p=0.00079)
0.00
0.02
0.04
log(OR)
PM−Plot
−log10(P value)
7
6
5
4
3
0.25
0.50
0.75
1.00
M value
phenotype
ADHD
CUD

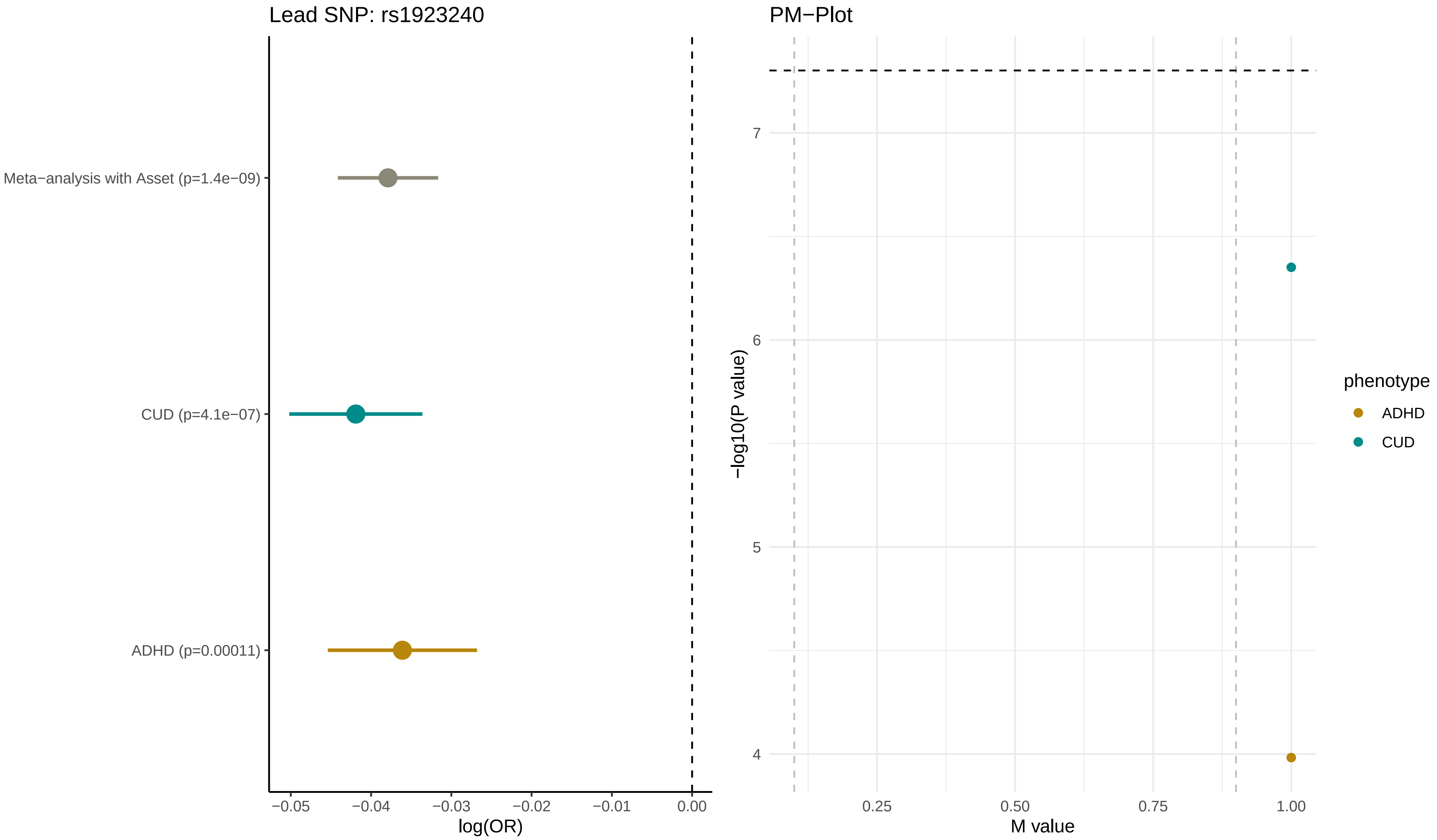
Lead SNP: rs1923240
Meta−analysis with Asset (p=1.4e−09)
CUD (p=4.1e−07)
ADHD (p=0.00011)
−0.05
−0.04
−0.03
−0.02
−0.01
0.00
log(OR)
PM−Plot
−log10(P value)
7
6
5
4
0.25
0.50
0.75
1.00
M value
phenotype
ADHD
CUD

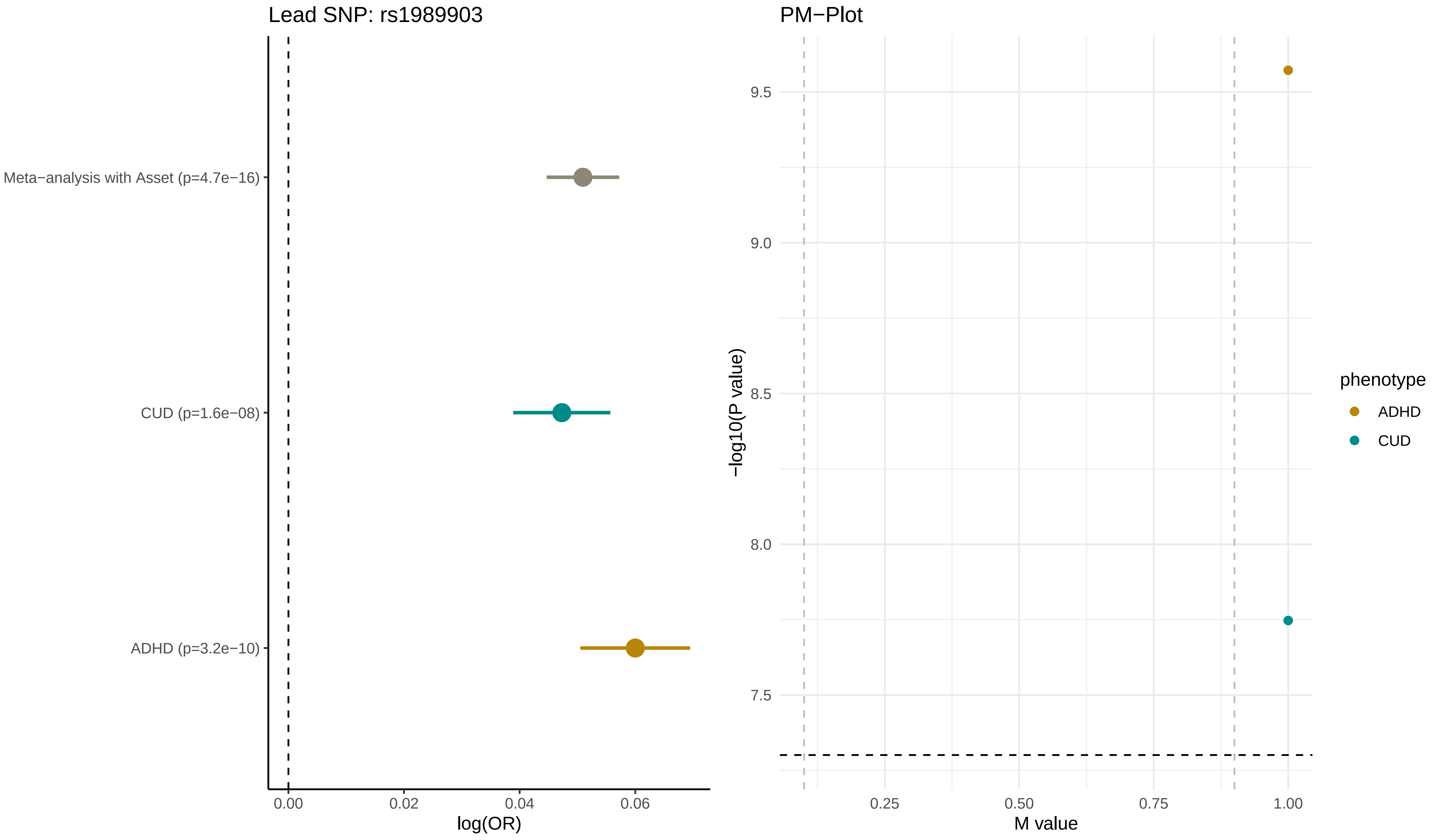

Lead SNP: rs1989903
Meta−analysis with Asset (p=4.7e−16)
CUD (p=1.6e−08)
ADHD (p=3.2e−10)
0.00
0.02
0.04
0.06
log(OR)
PM−Plot
9.5
9.0
8.5
8.0
7.5
−log10(P value)
0.25
0.50
0.75
1.00
M value
phenotype
ADHD
CUD

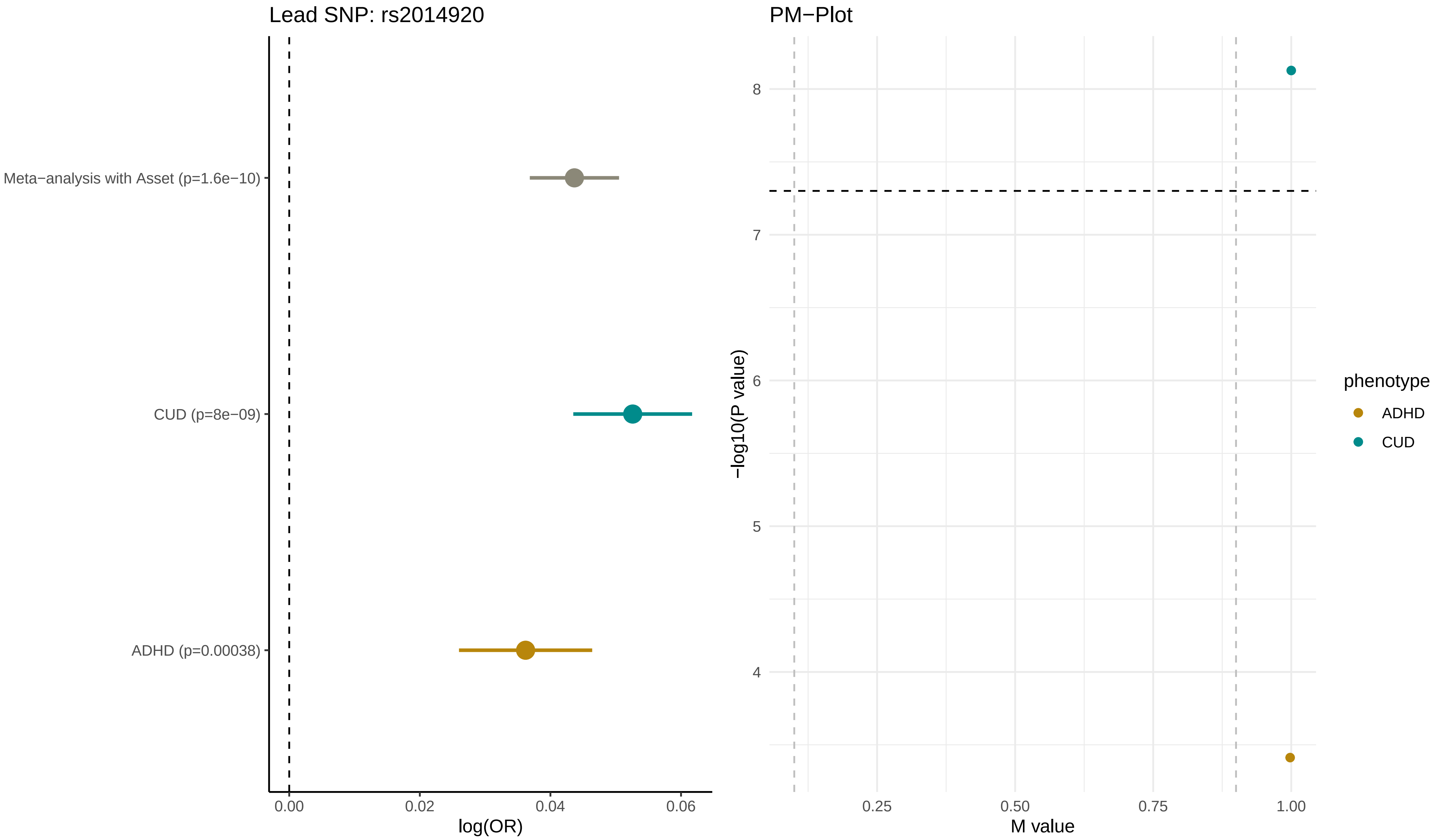

Lead SNP: rs2014920
Meta−analysis with Asset (p=1.6e−10)
CUD (p=8e−09)
ADHD (p=0.00038)
0.00
0.02
0.04
0.06
log(OR)
PM−Plot
8
7
6
5
4
−log10(P value)
0.25
0.50
0.75
1.00
M value
phenotype
ADHD
CUD

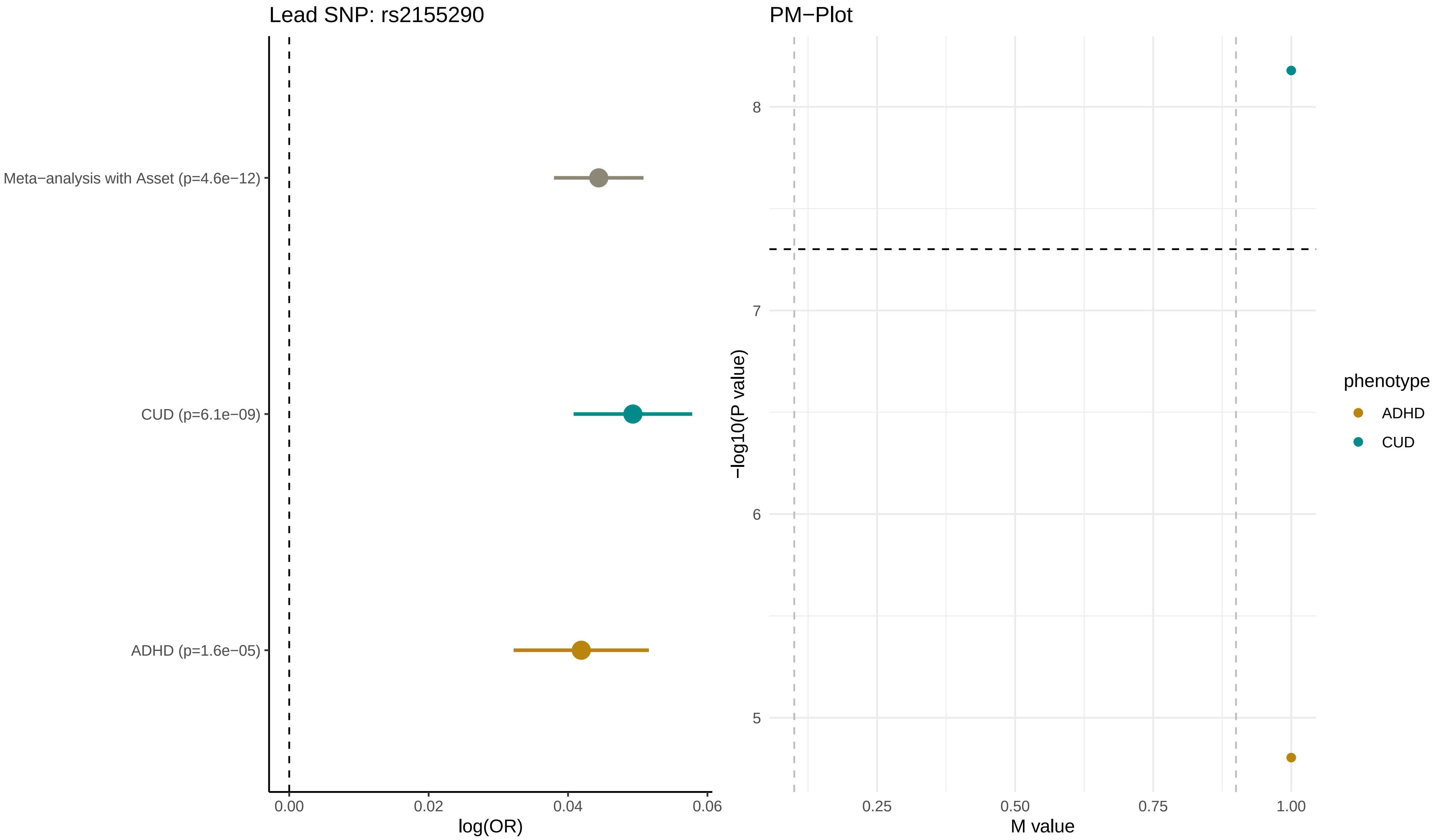
Lead SNP: rs2155290
Meta−analysis with Asset (p=4.6e−12)
CUD (p=6.1e−09)
ADHD (p=1.6e−05)
0.00
0.02
0.04
0.06
log(OR)
PM−Plot
8
7
6
5
−log10(P value)
0.25
0.50
0.75
1.00
M value
phenotype
ADHD
CUD

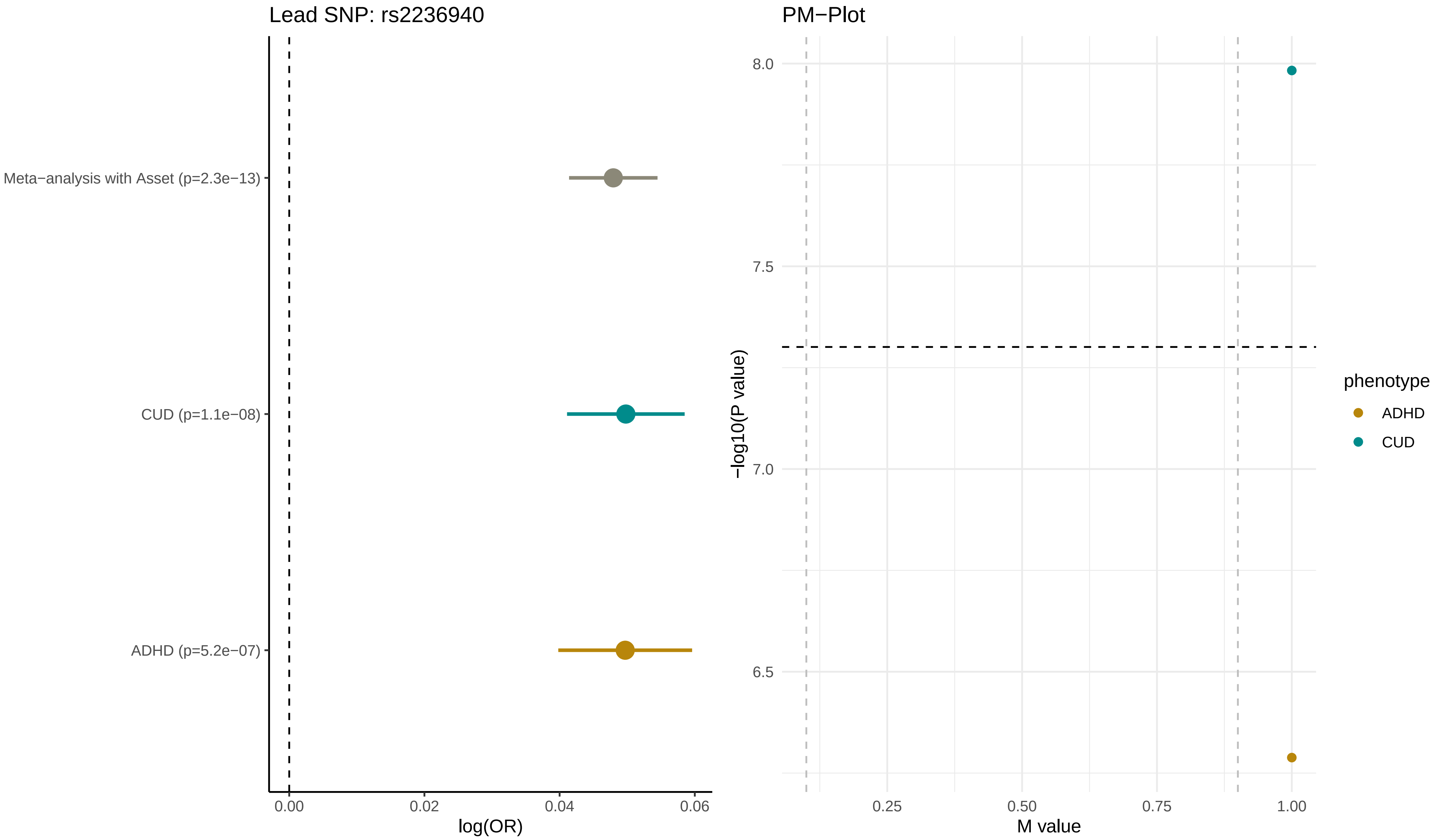

Lead SNP: rs2236940
Meta−analysis with Asset (p=2.3e−13)
CUD (p=1.1e−08)
ADHD (p=5.2e−07)
0.00
0.02
0.04
0.06
log(OR)
PM−Plot
8.0
7.5
7.0
6.5
−log10(P value)
0.25
0.50
0.75
1.00
M value
phenotype
ADHD
CUD

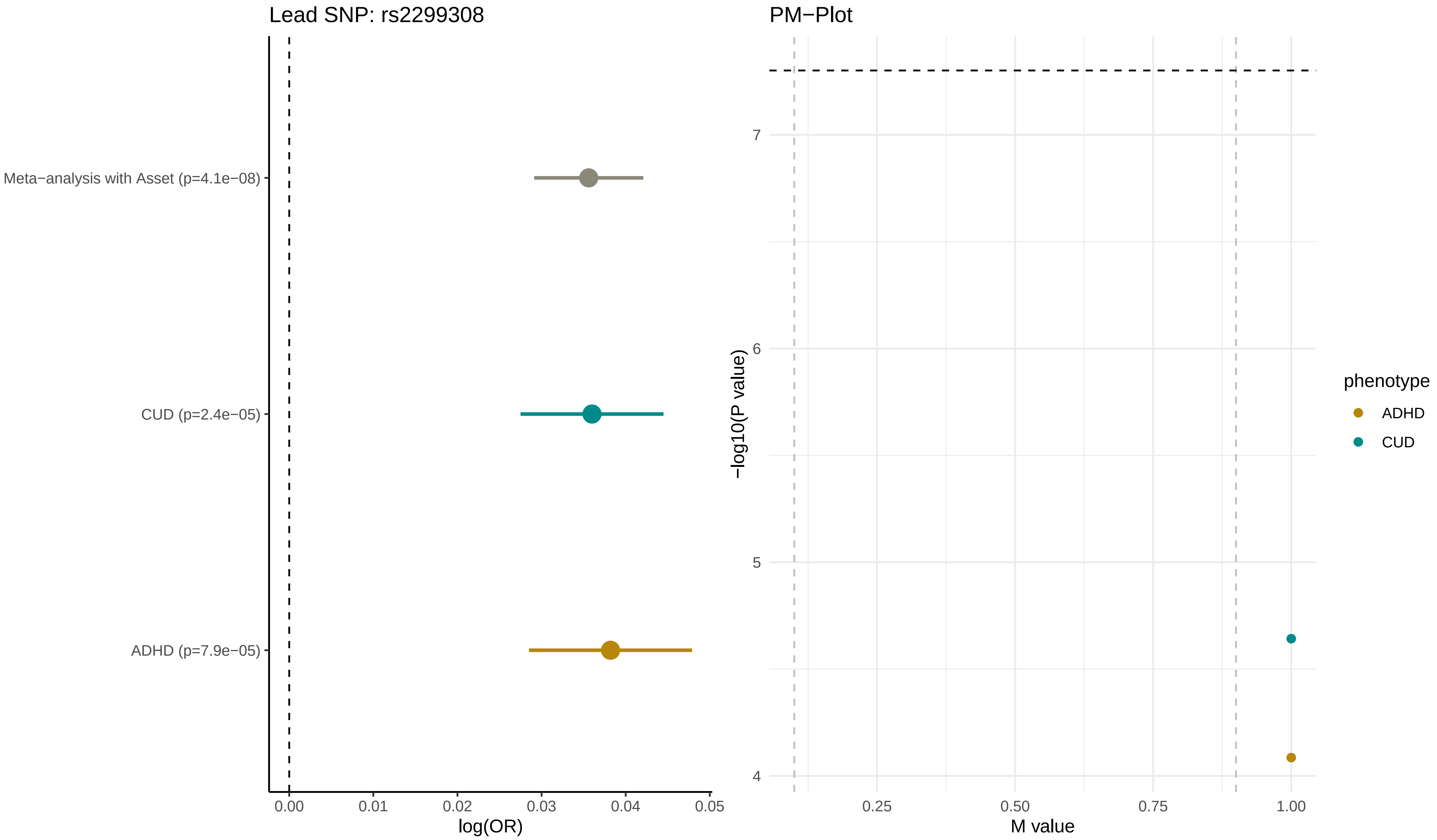

Lead SNP: rs2299308
Meta−analysis with Asset (p=4.1e−08)
CUD (p=2.4e−05)
ADHD (p=7.9e−05)
0.00
0.01
0.02
0.03
0.04
0.05
log(OR)
PM−Plot
−log10(P value)
7
6
5
4
0.25
0.50
0.75
1.00
M value
phenotype
ADHD
CUD

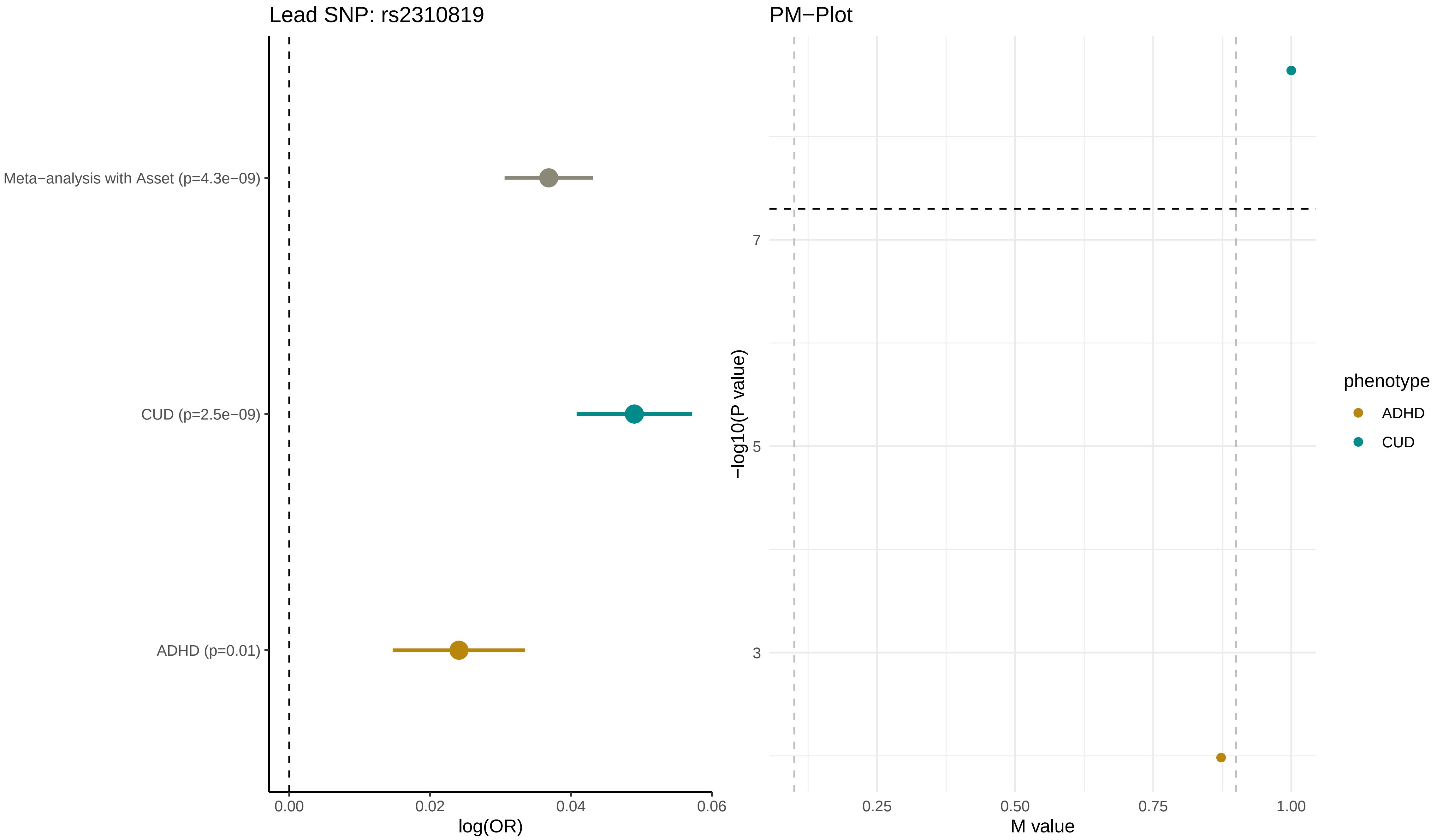
Lead SNP: rs2310819
Meta−analysis with Asset (p=4.3e−09)
CUD (p=2.5e−09)
ADHD (p=0.01)
0.00
0.02
0.04
0.06
log(OR)
PM−Plot
7
5
3
−log10(P value)
0.25
0.50
0.75
1.00
M value
phenotype
ADHD
CUD

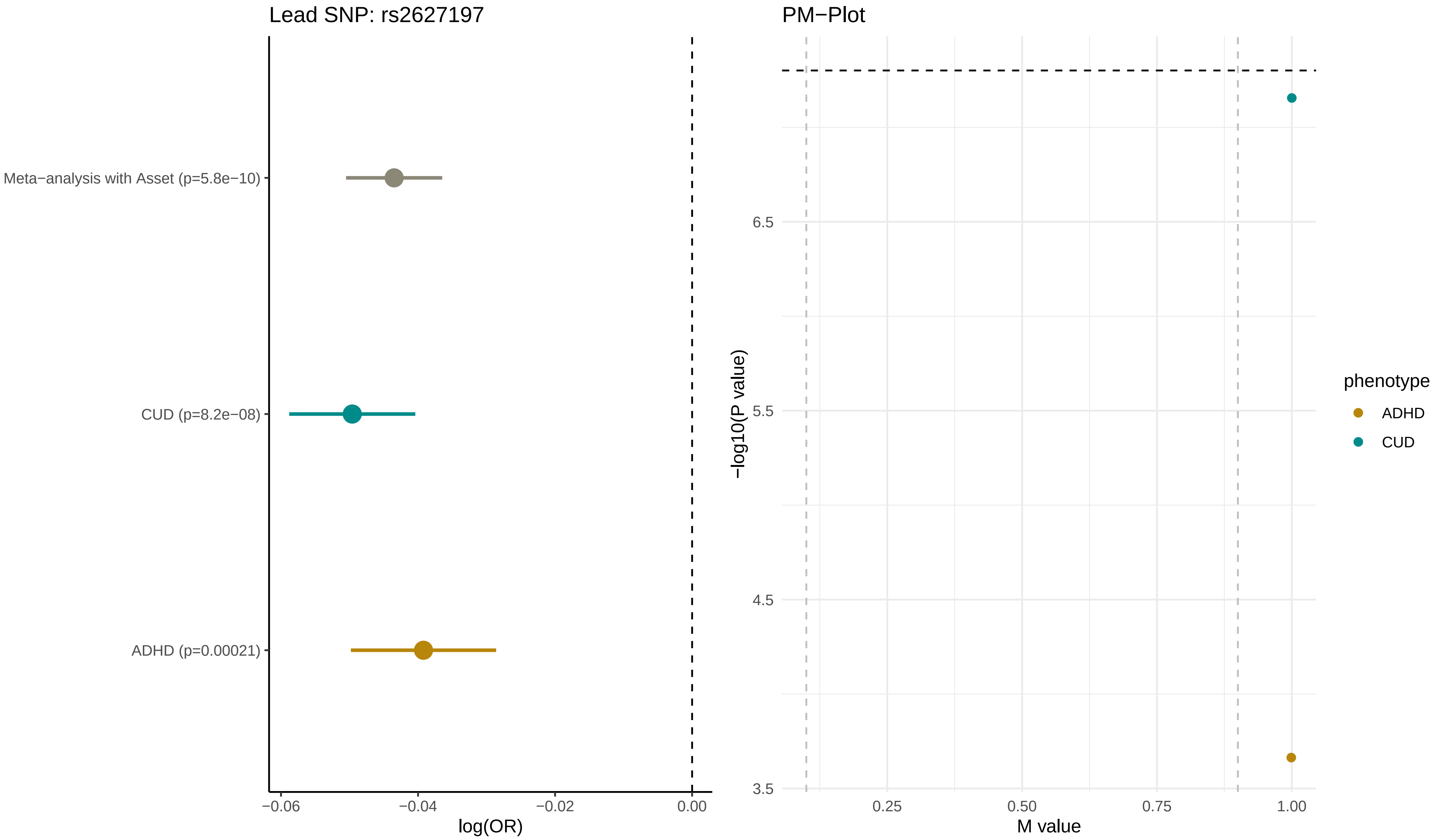

Lead SNP: rs2627197
Meta−analysis with Asset (p=5.8e−10)
CUD (p=8.2e−08)
ADHD (p=0.00021)
−0.06
−0.04
−0.02
0.00
log(OR)
PM−Plot
6.5
5.5
4.5
3.5
−log10(P value)
0.25
0.50
0.75
1.00
M value
phenotype
ADHD
CUD

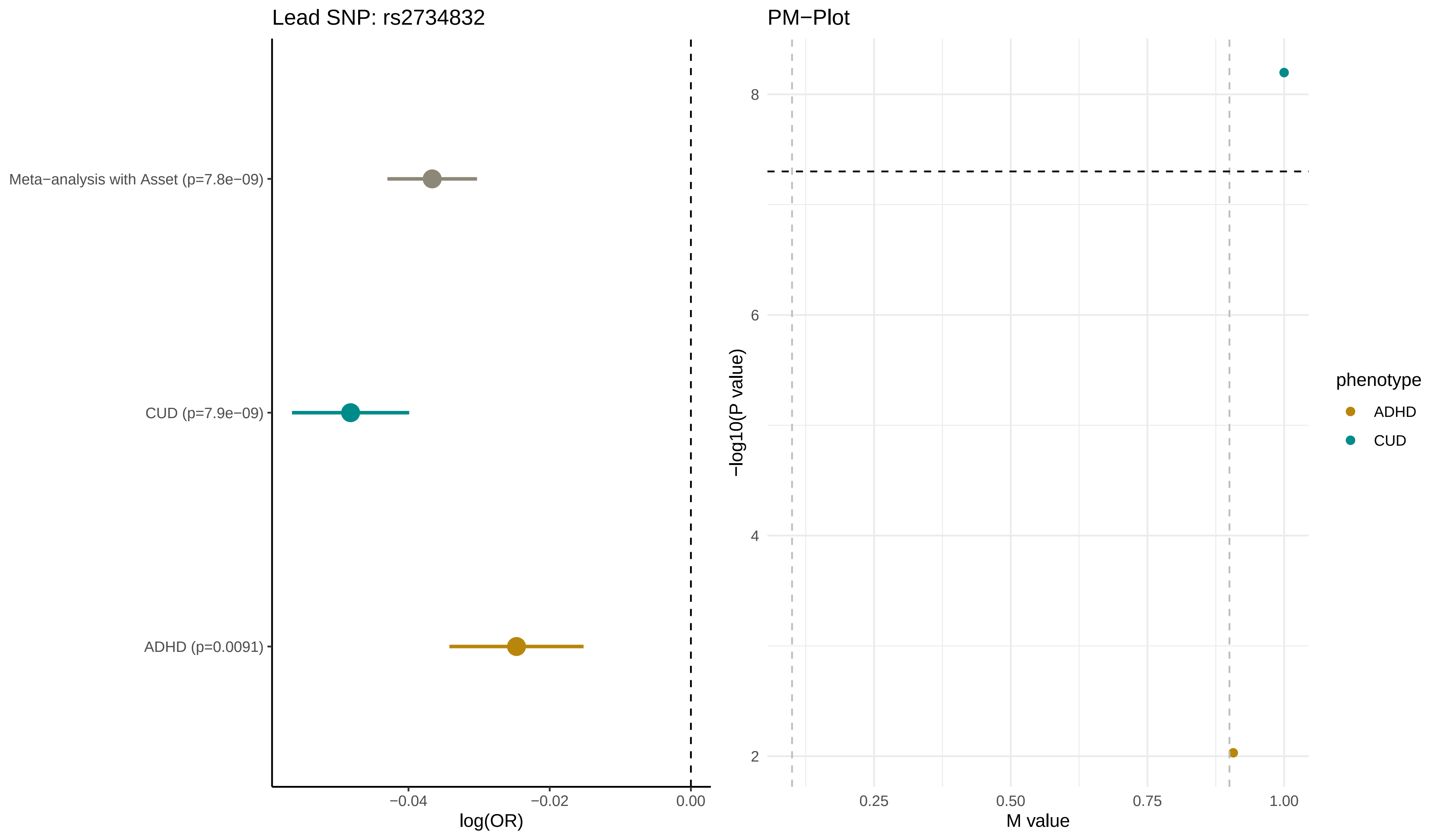

Lead SNP: rs2734832
Meta−analysis with Asset (p=7.8e−09)
CUD (p=7.9e−09)
ADHD (p=0.0091)
−0.04
−0.02
0.00
log(OR)
PM−Plot
8
6
4
2
−log10(P value)
0.25
0.50
0.75
1.00
M value
phenotype
ADHD
CUD

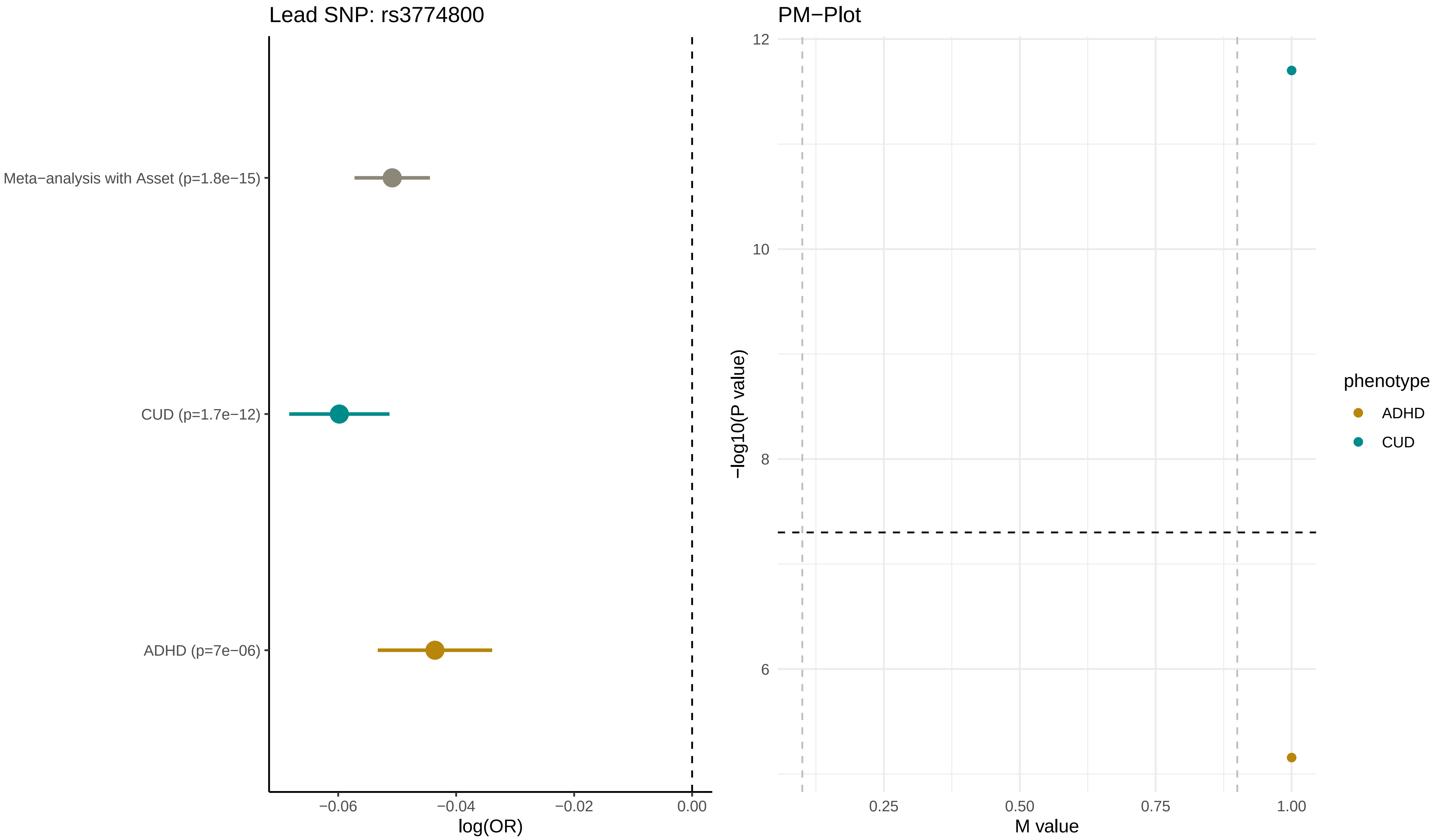

Lead SNP: rs3774800
Meta−analysis with Asset (p=1.8e−15)
CUD (p=1.7e−12)
ADHD (p=7e−06)
−0.06
−0.04
−0.02
0.00
log(OR)
PM−Plot
12
10
8
6
−log10(P value)
0.25
0.50
0.75
1.00
M value
phenotype
ADHD
CUD

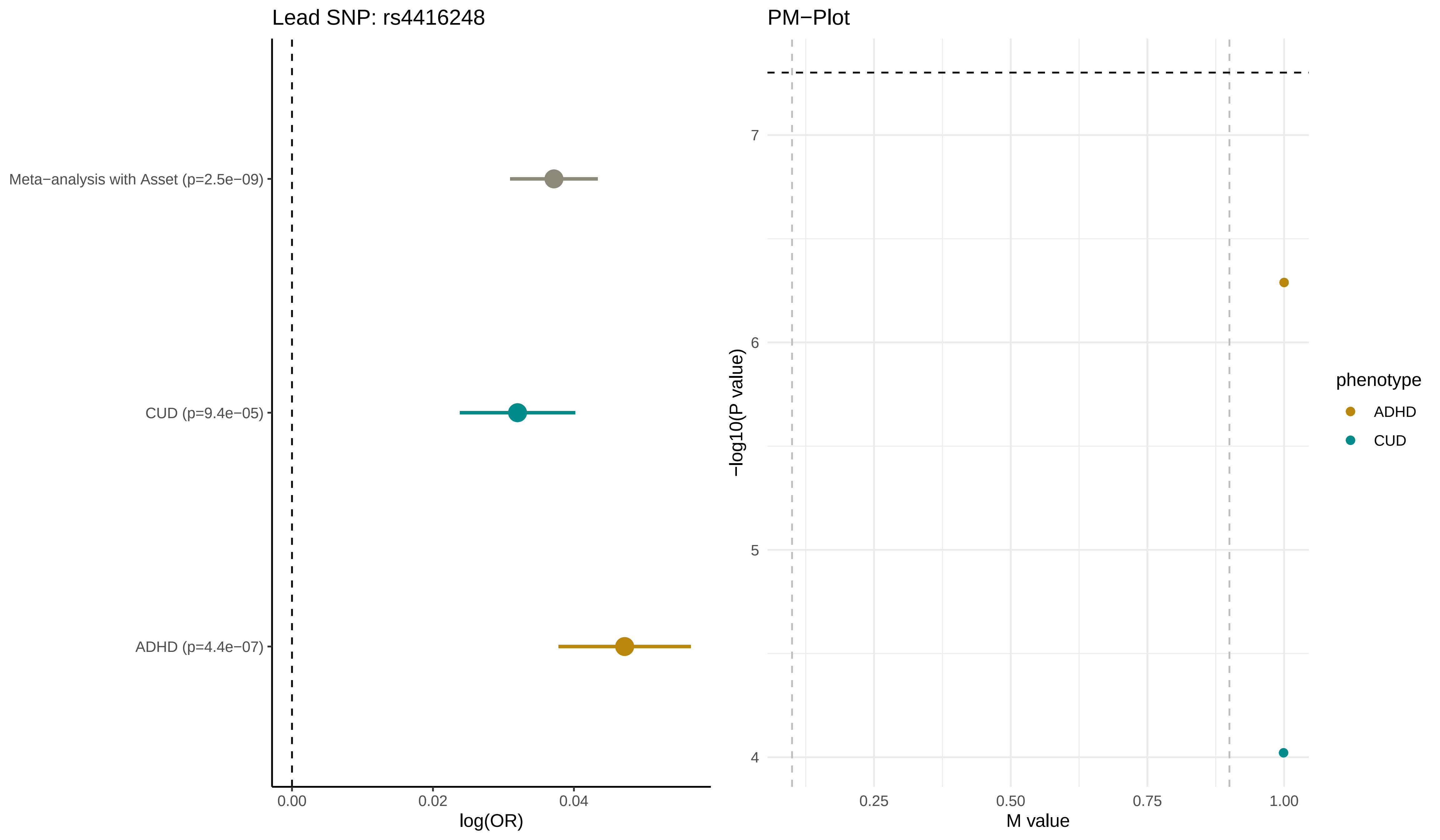
Lead SNP: rs4416248
Meta−analysis with Asset (p=2.5e−09)
CUD (p=9.4e−05)
ADHD (p=4.4e−07)
0.00
0.02
0.04
log(OR)
PM−Plot
7
6
5
4
−log10(P value)
0.25
0.50
0.75
1.00
M value
phenotype
ADHD
CUD

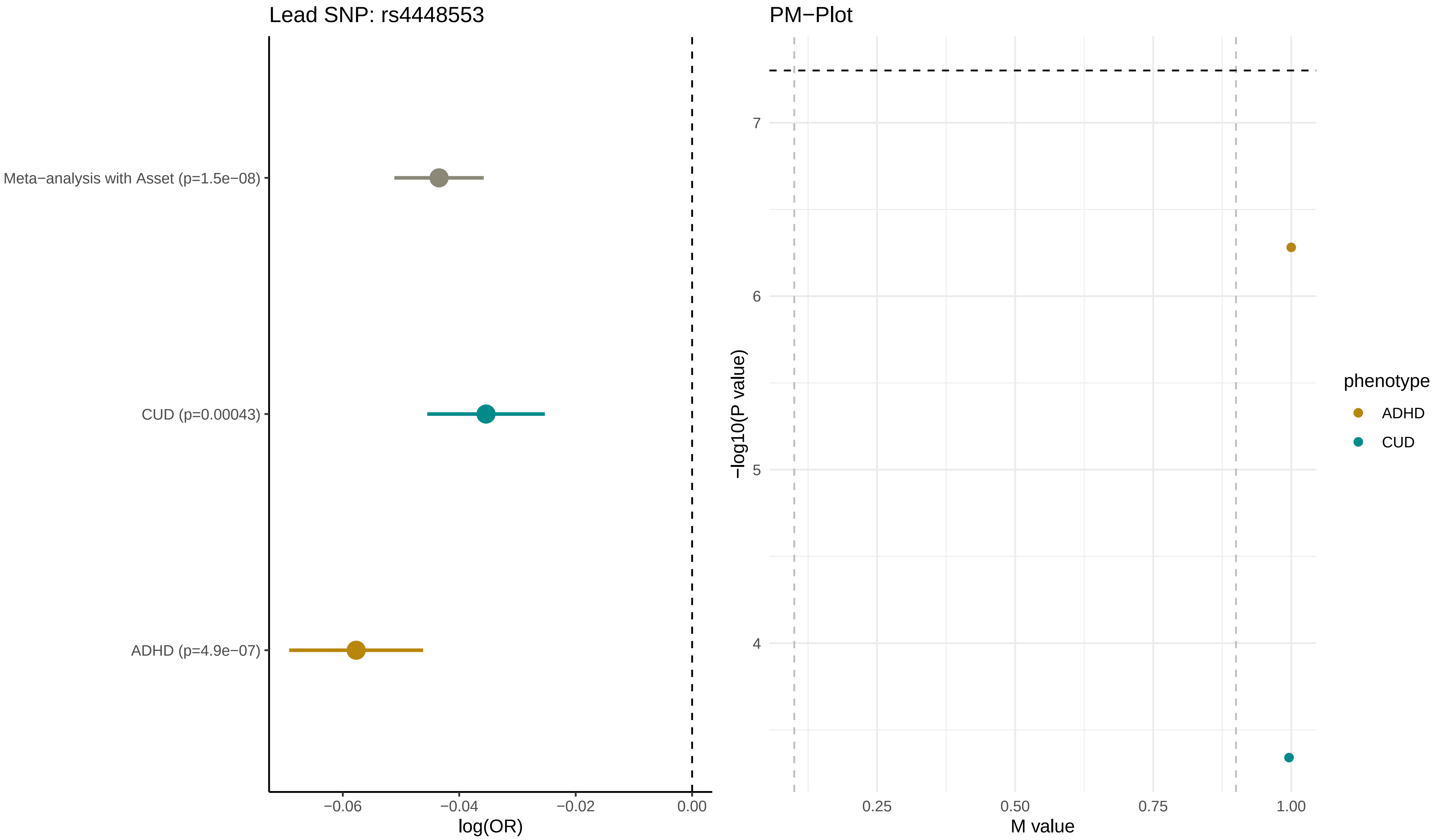
Lead SNP: rs4448553
Meta−analysis with Asset (p=1.5e−08)
CUD (p=0.00043)
ADHD (p=4.9e−07)
−0.06
−0.04
−0.02
0.00
log(OR)
PM−Plot
−log10(P value)
7
6
5
4
0.25
0.50
0.75
1.00
M value
phenotype
ADHD
CUD

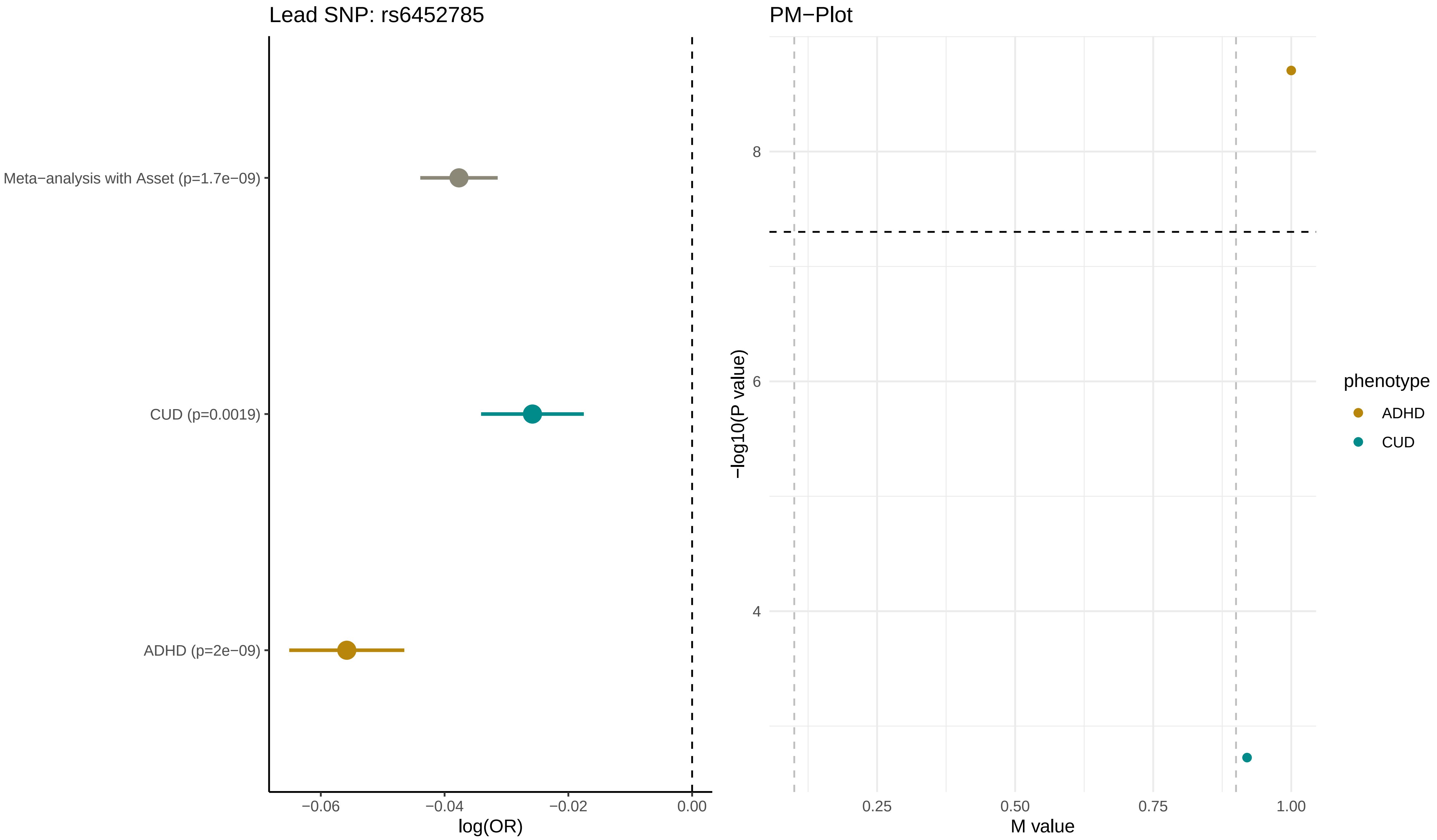

Lead SNP: rs6452785
Meta−analysis with Asset (p=1.7e−09)
CUD (p=0.0019)
ADHD (p=2e−09)
−0.06
−0.04
−0.02
0.00
log(OR)
PM−Plot
−log10(P value)
8
6
4
0.25
0.50
0.75
1.00
M value
phenotype
ADHD
CUD

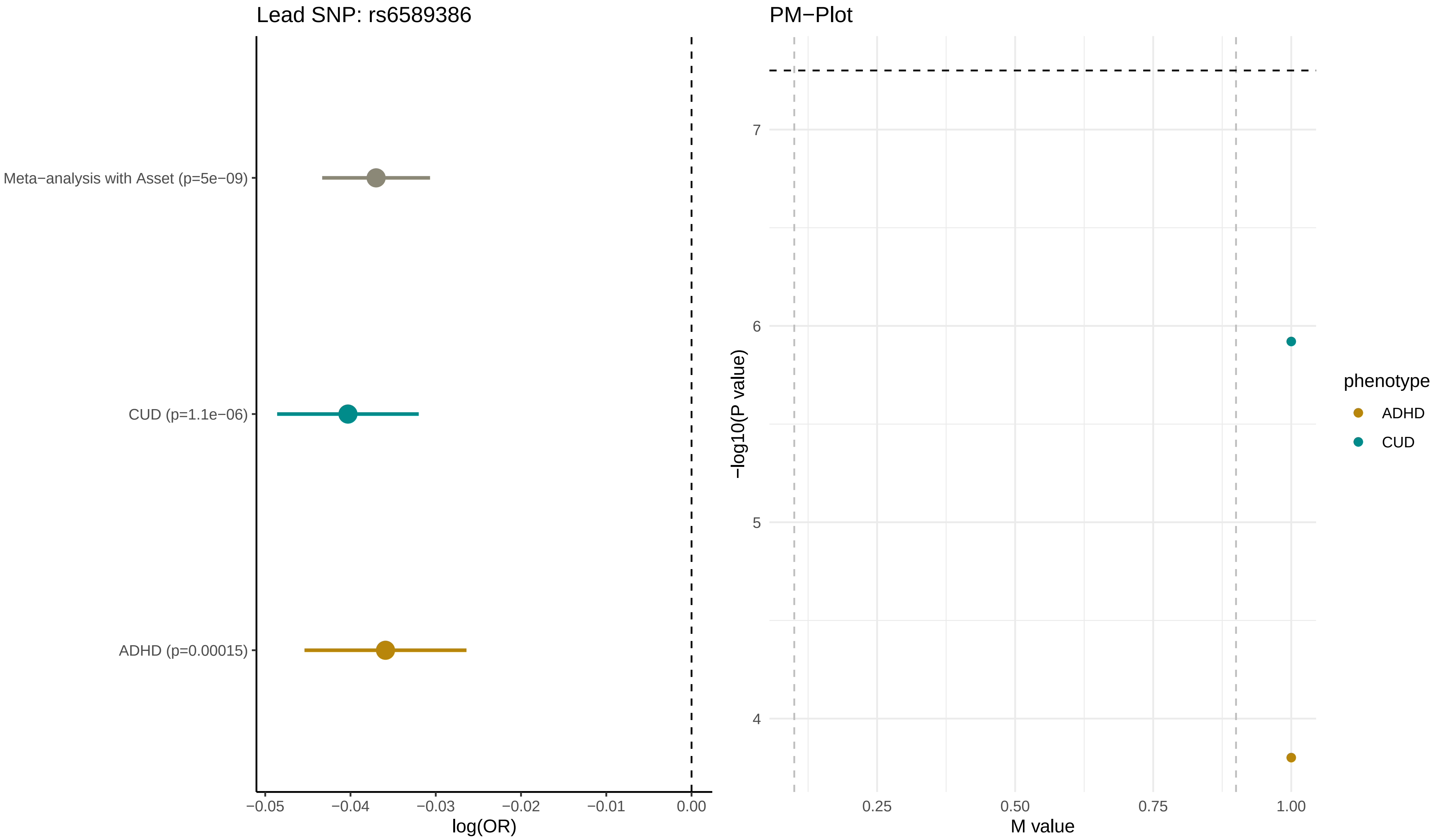
Lead SNP: rs6589386
Meta−analysis with Asset (p=5e−09)
CUD (p=1.1e−06)
ADHD (p=0.00015)
−0.05
−0.04
−0.03
−0.02
−0.01
0.00
log(OR)
PM−Plot
7
6
5
4
−log10(P value)
0.25
0.50
0.75
1.00
M value
phenotype
ADHD
CUD

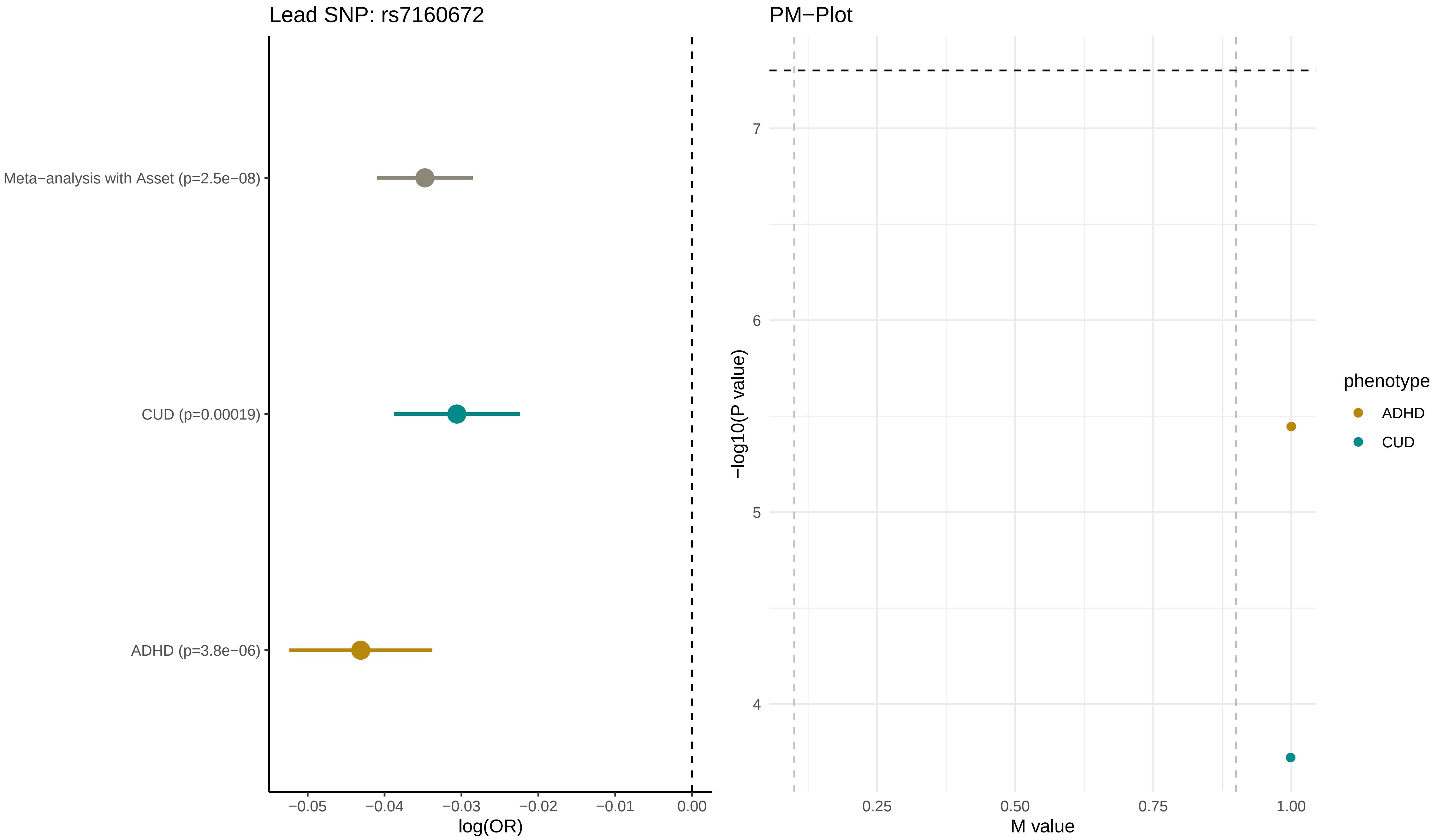

Lead SNP: rs7160672
Meta−analysis with Asset (p=2.5e−08)
CUD (p=0.00019)
ADHD (p=3.8e−06)
−0.05
−0.04
−0.03
−0.02
−0.01
0.00
log(OR)
PM−Plot
7
6
5
4
−log10(P value)
0.25
0.50
0.75
1.00
M value
phenotype
ADHD
CUD

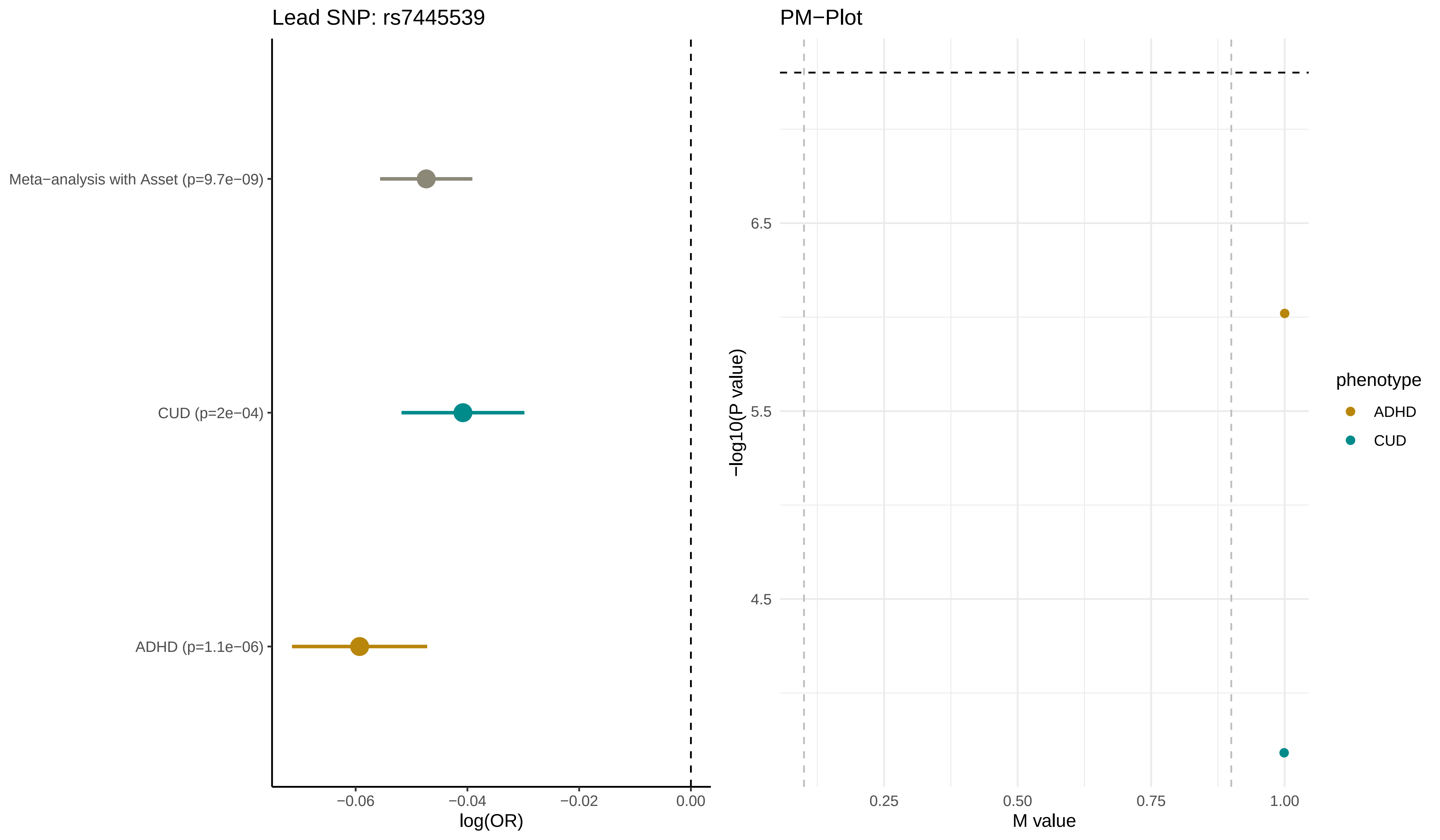

Lead SNP: rs7445539
Meta−analysis with Asset (p=9.7e−09)
CUD (p=2e−04)
ADHD (p=1.1e−06)
−0.06
−0.04
−0.02
0.00
log(OR)
PM−Plot
6.5
5.5
4.5
−log10(P value)
0.25
0.50
0.75
1.00
M value
phenotype
ADHD
CUD

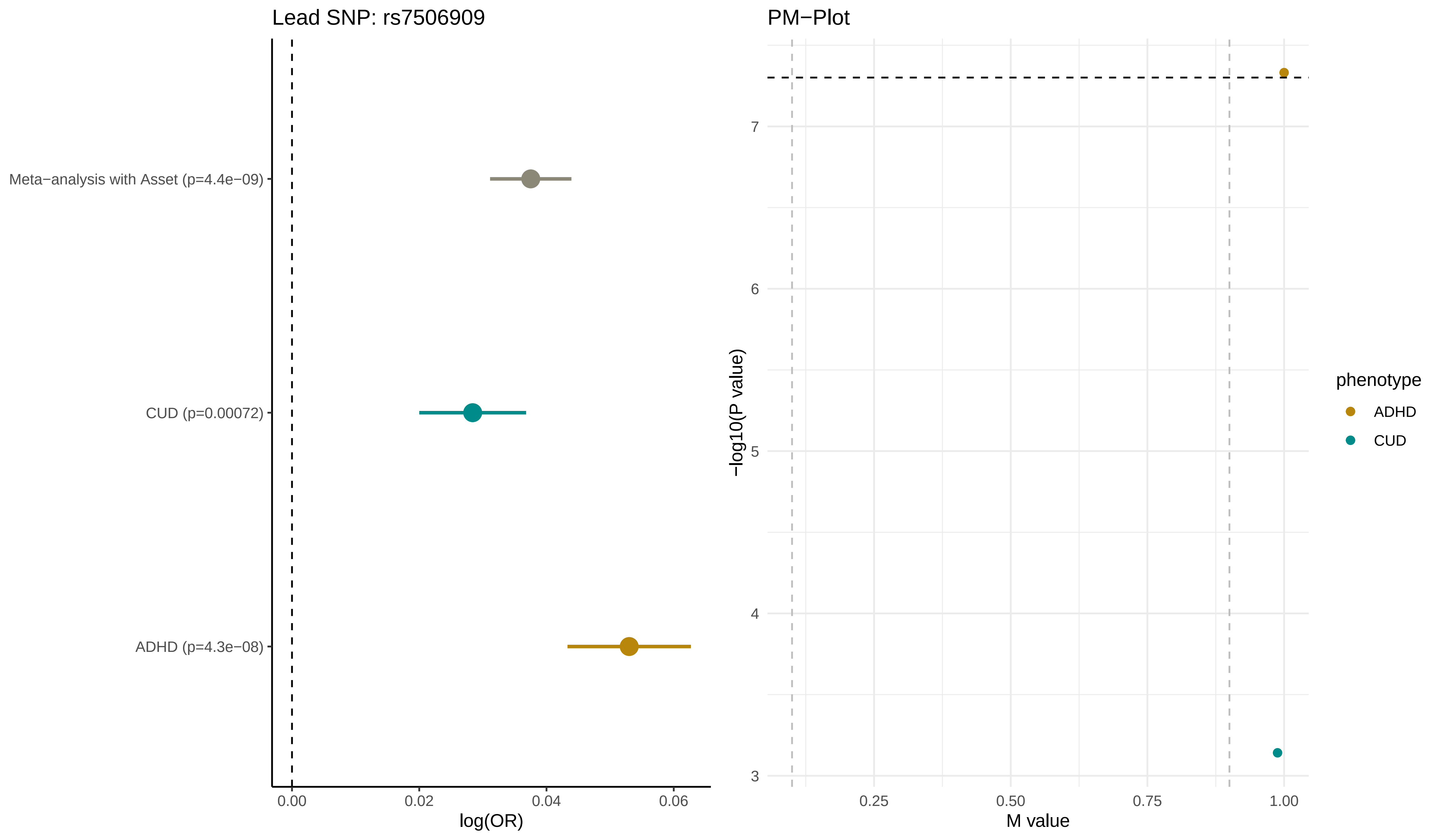
Lead SNP: rs7506909
Meta−analysis with Asset (p=4.4e−09)
CUD (p=0.00072)
ADHD (p=4.3e−08)
0.00
0.02
0.04
0.06
log(OR)
PM−Plot
−log10(P value)
3
4
5
6
7
0.25
0.50
0.75
1.00
M value
phenotype
ADHD
CUD

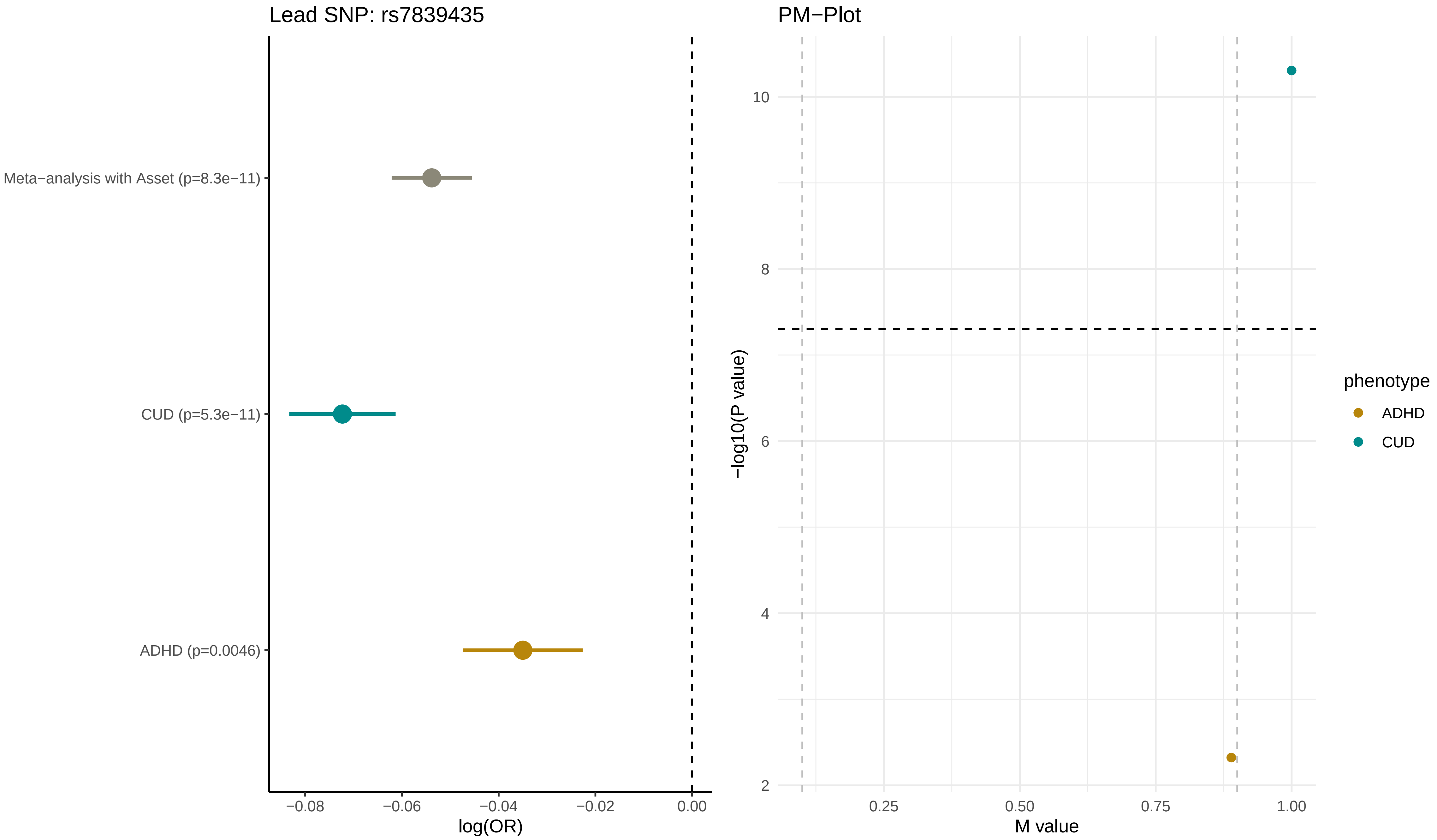

Lead SNP: rs7839435
Meta−analysis with Asset (p=8.3e−11)
CUD (p=5.3e−11)
ADHD (p=0.0046)
−0.08
−0.06
−0.04
−0.02
0.00
log(OR)
PM−Plot
10
8
6
4
2
−log10(P value)
0.25
0.50
0.75
1.00
M value
phenotype
ADHD
CUD

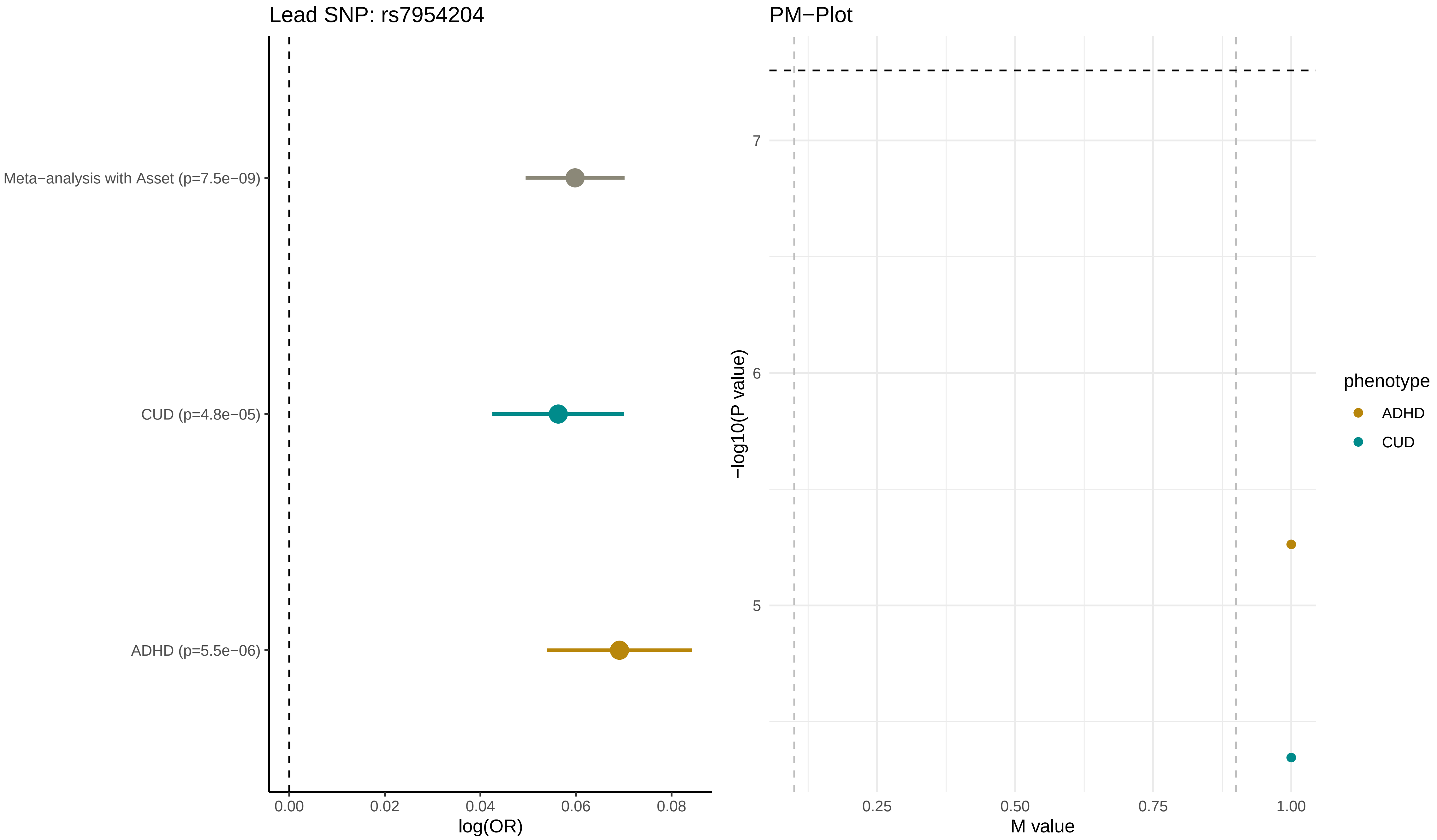
Lead SNP: rs7954204
Meta−analysis with Asset (p=7.5e−09)
CUD (p=4.8e−05)
ADHD (p=5.5e−06)
0.00
0.02
0.04
0.06
0.08
log(OR)
PM−Plot
7
6
5
−log10(P value)
0.25
0.50
0.75
1.00
M value
phenotype
ADHD
CUD

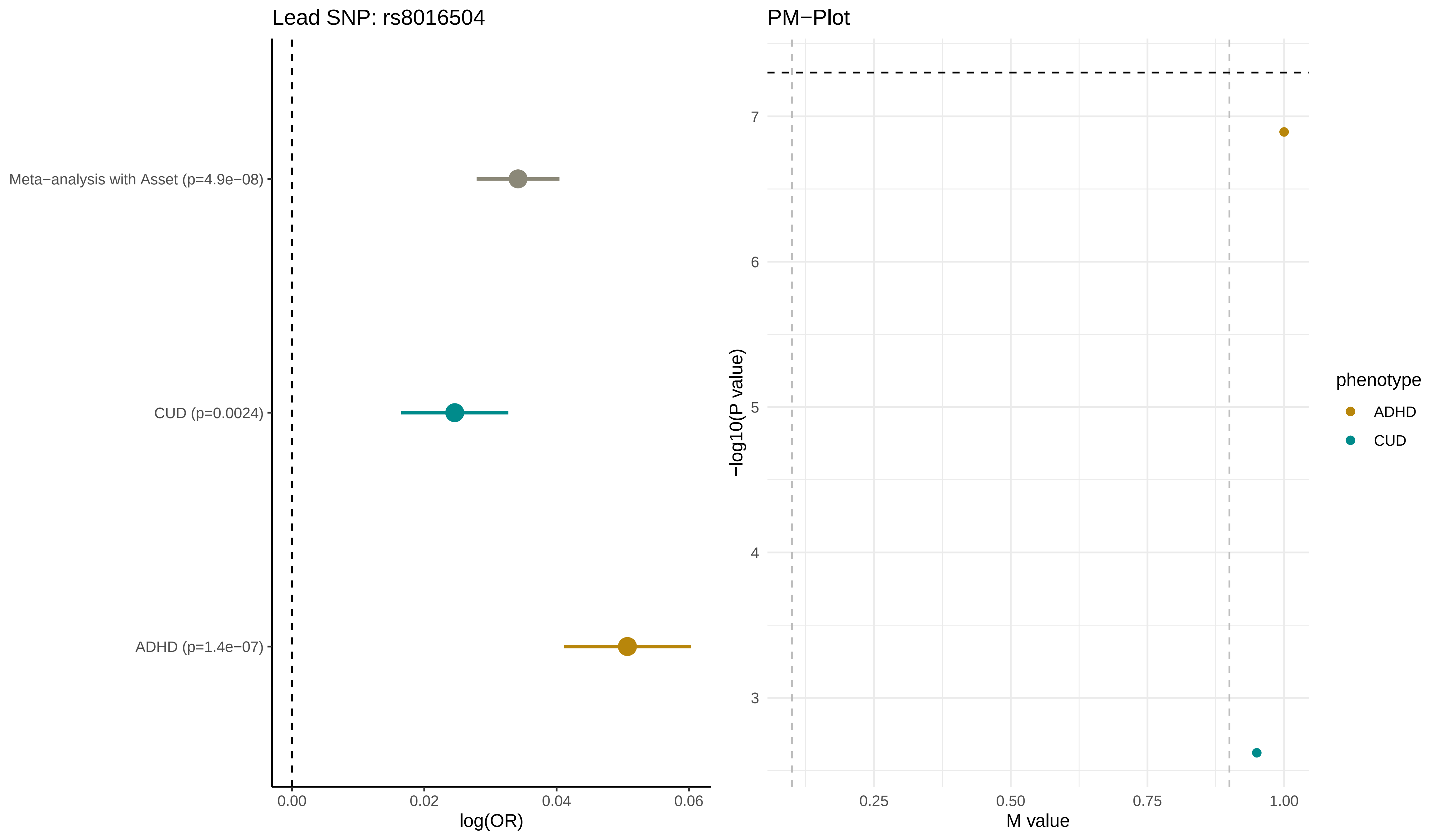

Lead SNP: rs8016504
Meta−analysis with Asset (p=4.9e−08)
CUD (p=0.0024)
ADHD (p=1.4e−07)
0.00
0.02
0.04
0.06
log(OR)
PM−Plot
7
6
5
4
3
−log10(P value)
0.25
0.50
0.75
1.00
M value
phenotype
ADHD
CUD

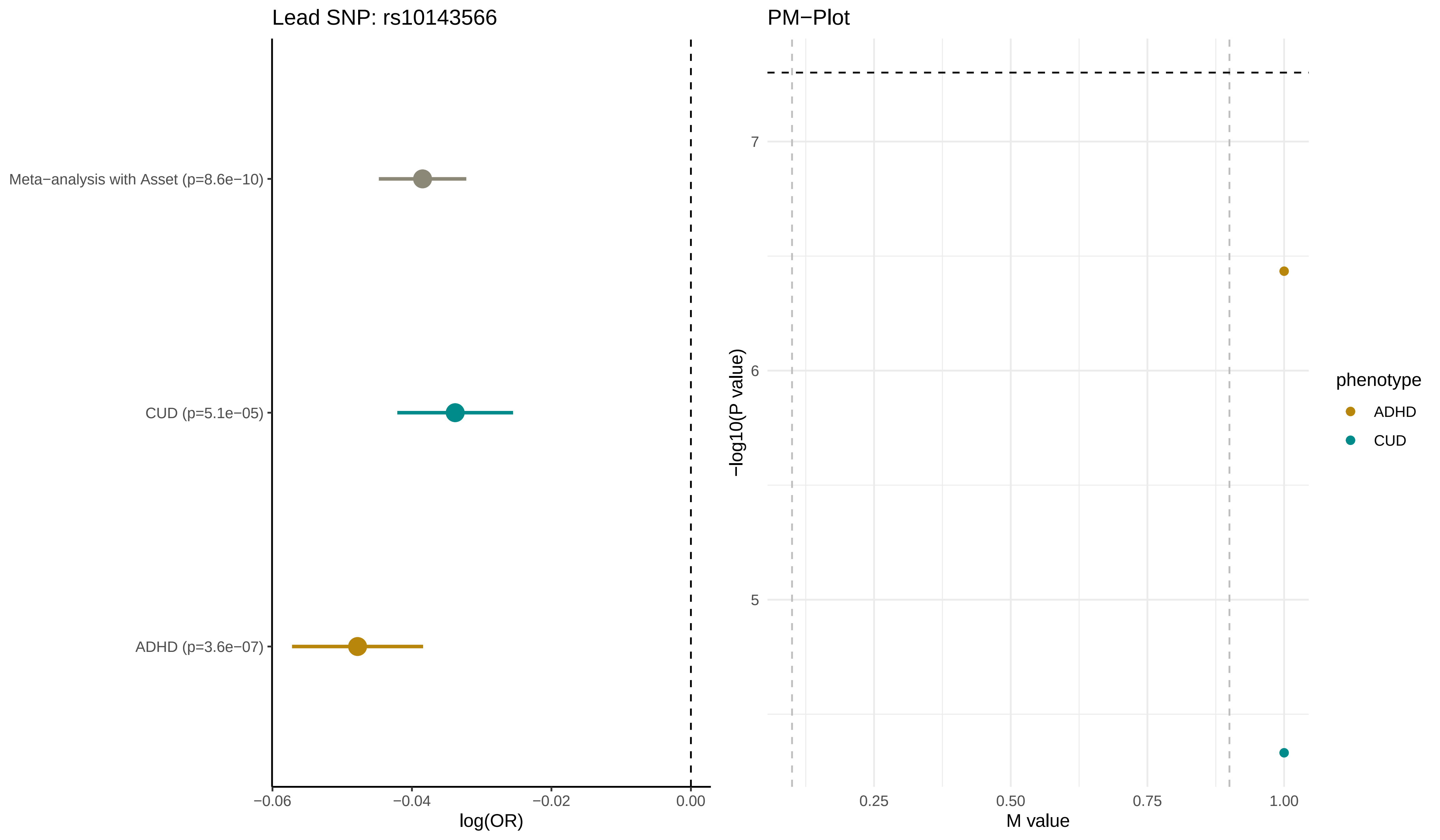
Lead SNP: rs10143566
Meta−analysis with Asset (p=8.6e−10)
CUD (p=5.1e−05)
ADHD (p=3.6e−07)
−0.06
−0.04
−0.02
0.00
log(OR)
PM−Plot
7
6
5
−log10(P value)
0.25
0.50
0.75
1.00
M value
phenotype
ADHD
CUD

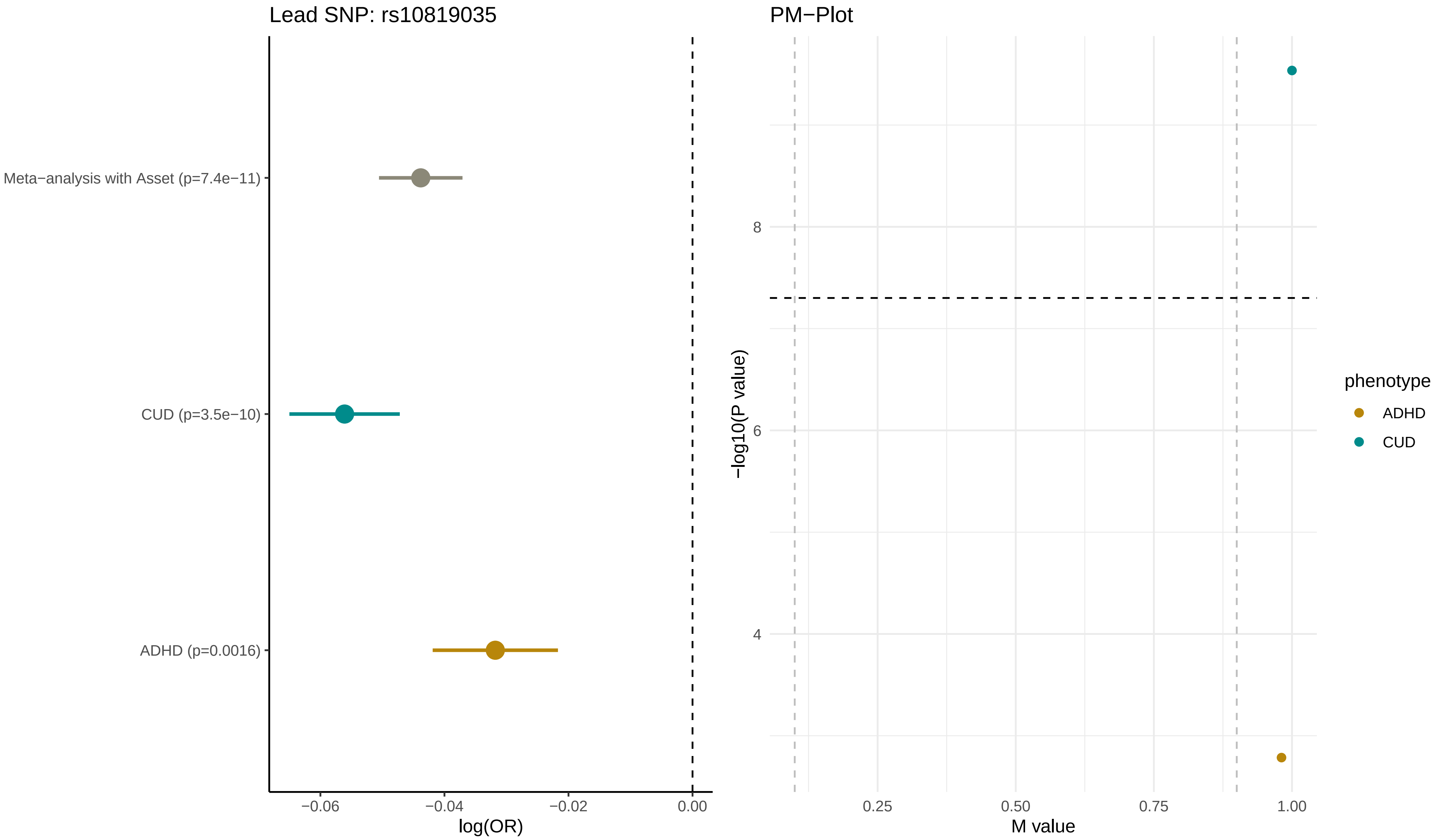

Lead SNP: rs10819035
Meta−analysis with Asset (p=7.4e−11)
CUD (p=3.5e−10)
ADHD (p=0.0016)
−0.06
−0.04
−0.02
0.00
log(OR)
PM−Plot
−log10(P value)
8
6
4
0.25
0.50
0.75
1.00
M value
phenotype
ADHD
CUD

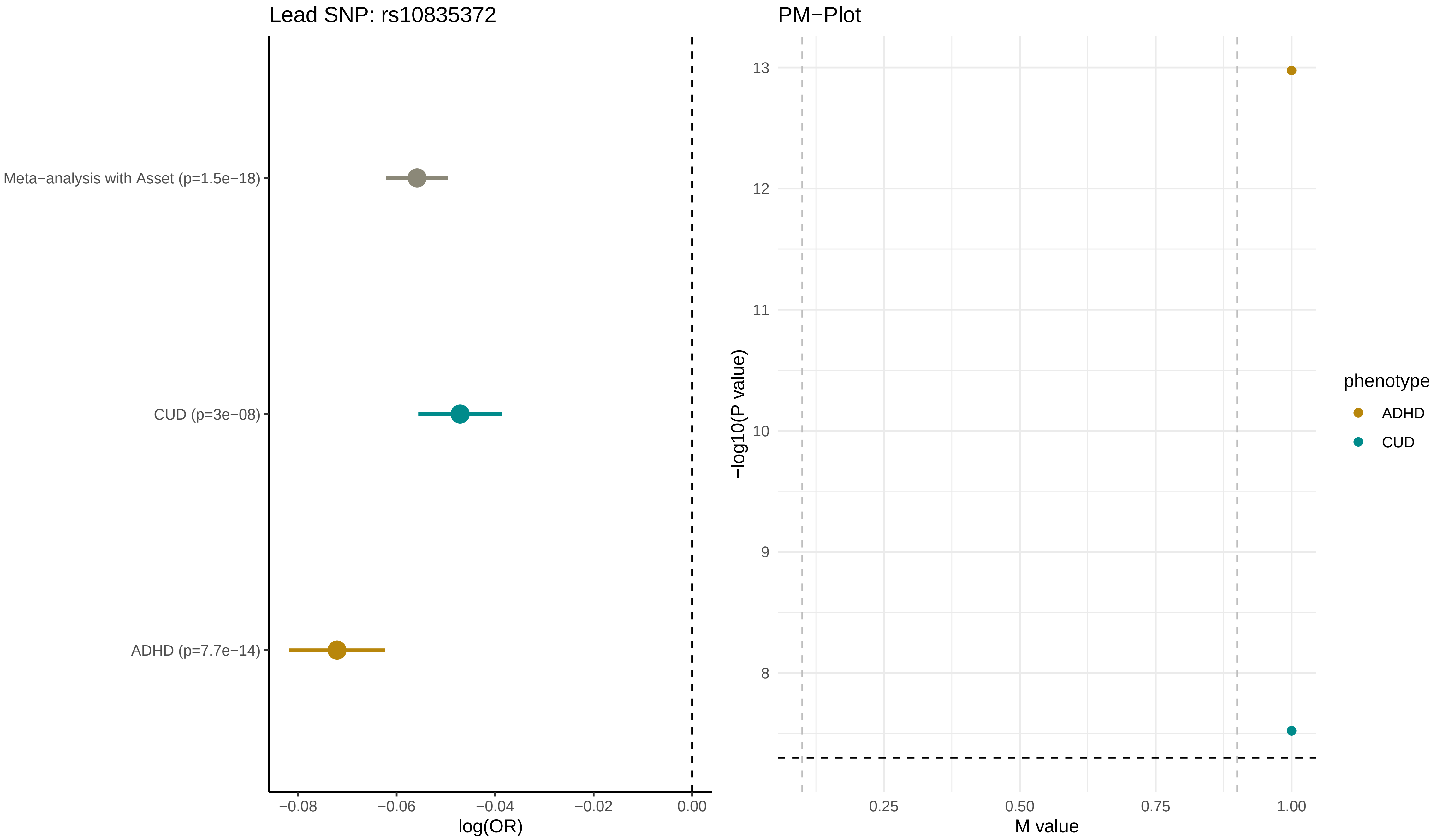

Lead SNP: rs10835372
Meta−analysis with Asset (p=1.5e−18)
CUD (p=3e−08)
ADHD (p=7.7e−14)
−0.08
−0.06
−0.04
−0.02
0.00
log(OR)
PM−Plot
13
12
11
10
9
8
−log10(P value)
0.25
0.50
0.75
1.00
M value
phenotype
ADHD
CUD

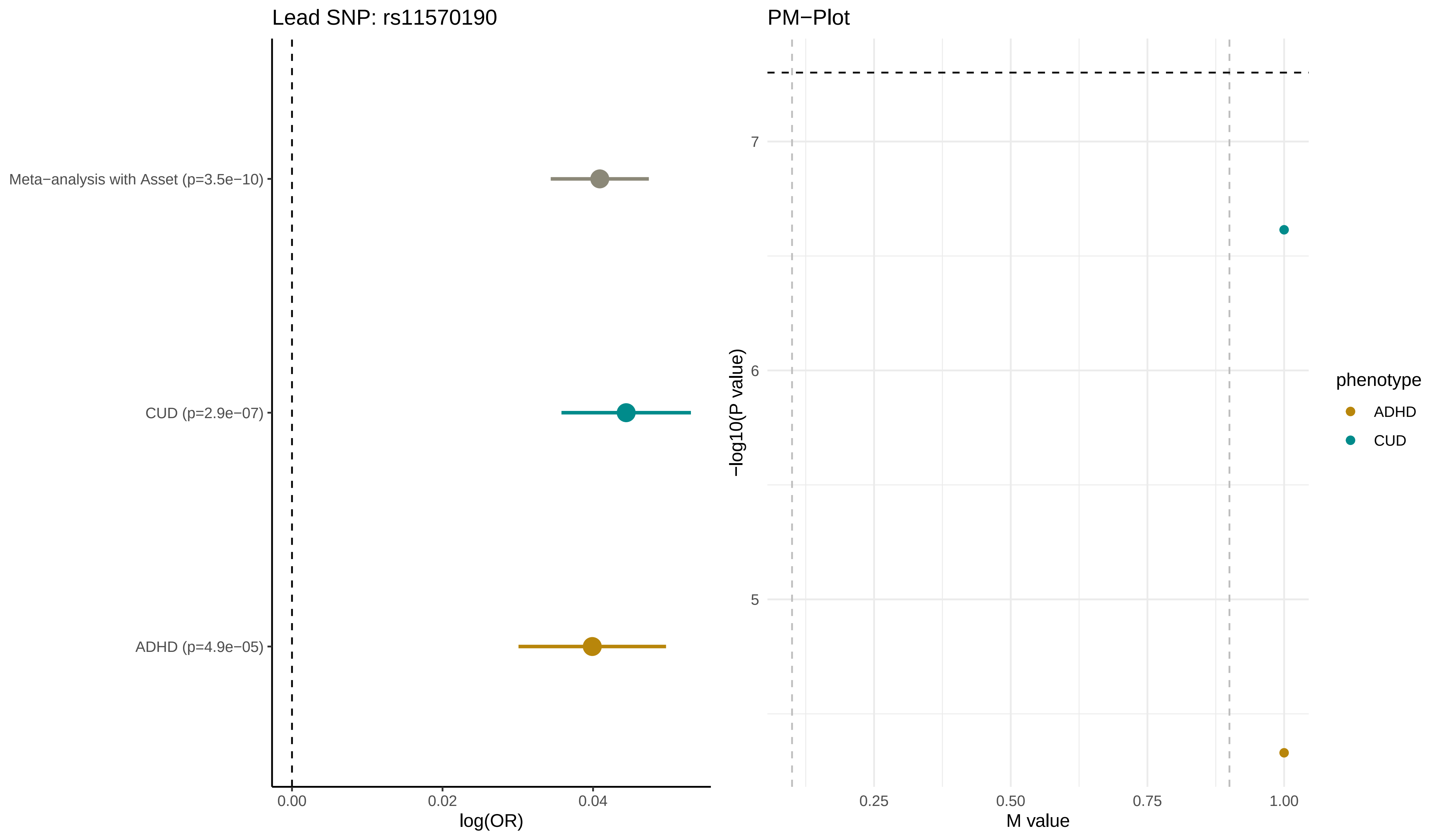

Lead SNP: rs11570190
Meta−analysis with Asset (p=3.5e−10)
CUD (p=2.9e−07)
ADHD (p=4.9e−05)
0.00
0.02
0.04
log(OR)
PM−Plot
7
6
5
−log10(P value)
0.25
0.50
0.75
1.00
M value
phenotype
ADHD
CUD

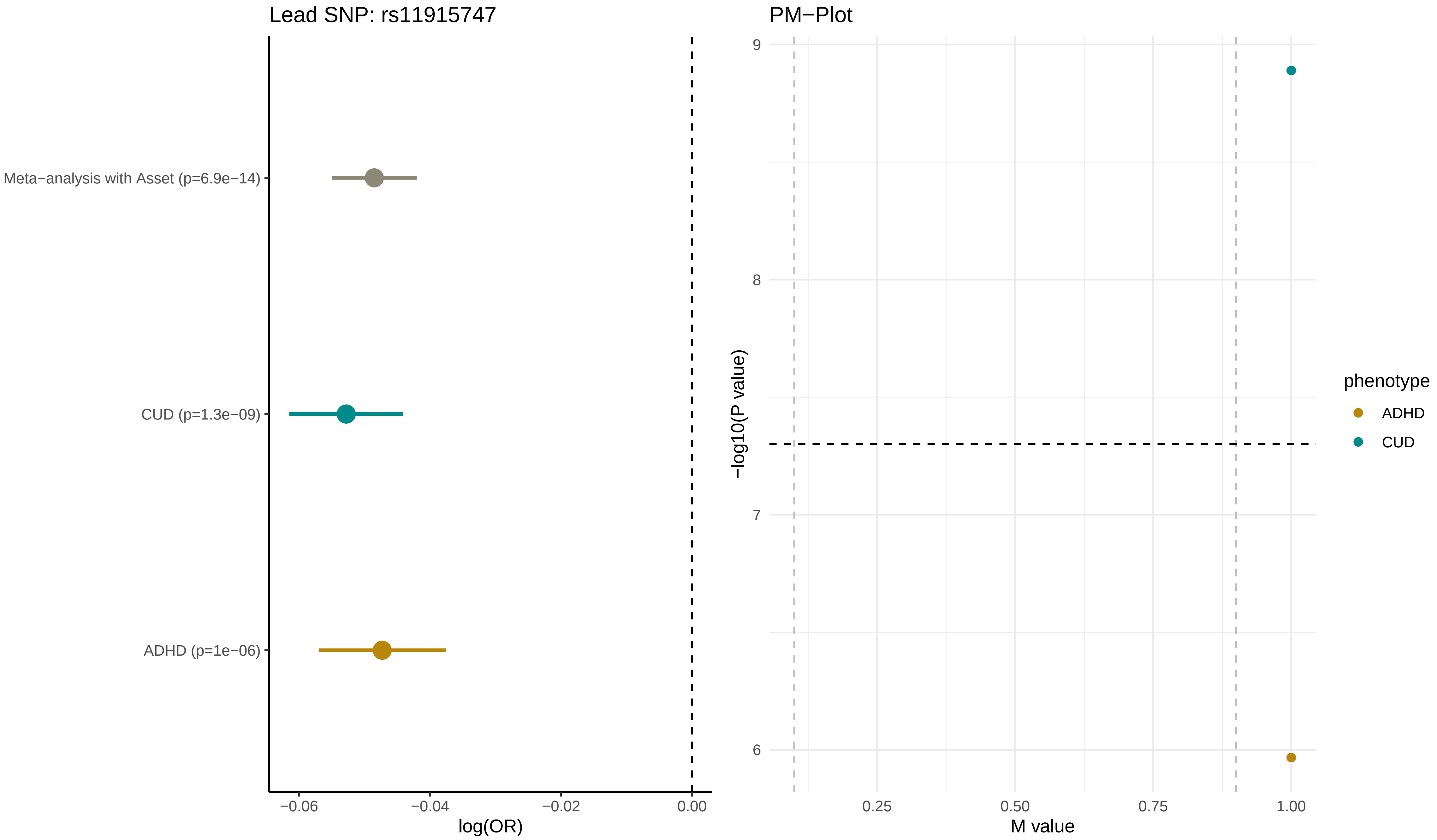
Lead SNP: rs11915747
Meta−analysis with Asset (p=6.9e−14)
CUD (p=1.3e−09)
ADHD (p=1e−06)
−0.06
−0.04
−0.02
0.00
log(OR)
PM−Plot
9
8
7
6
−log10(P value)
0.25
0.50
0.75
1.00
M value
phenotype
ADHD
CUD

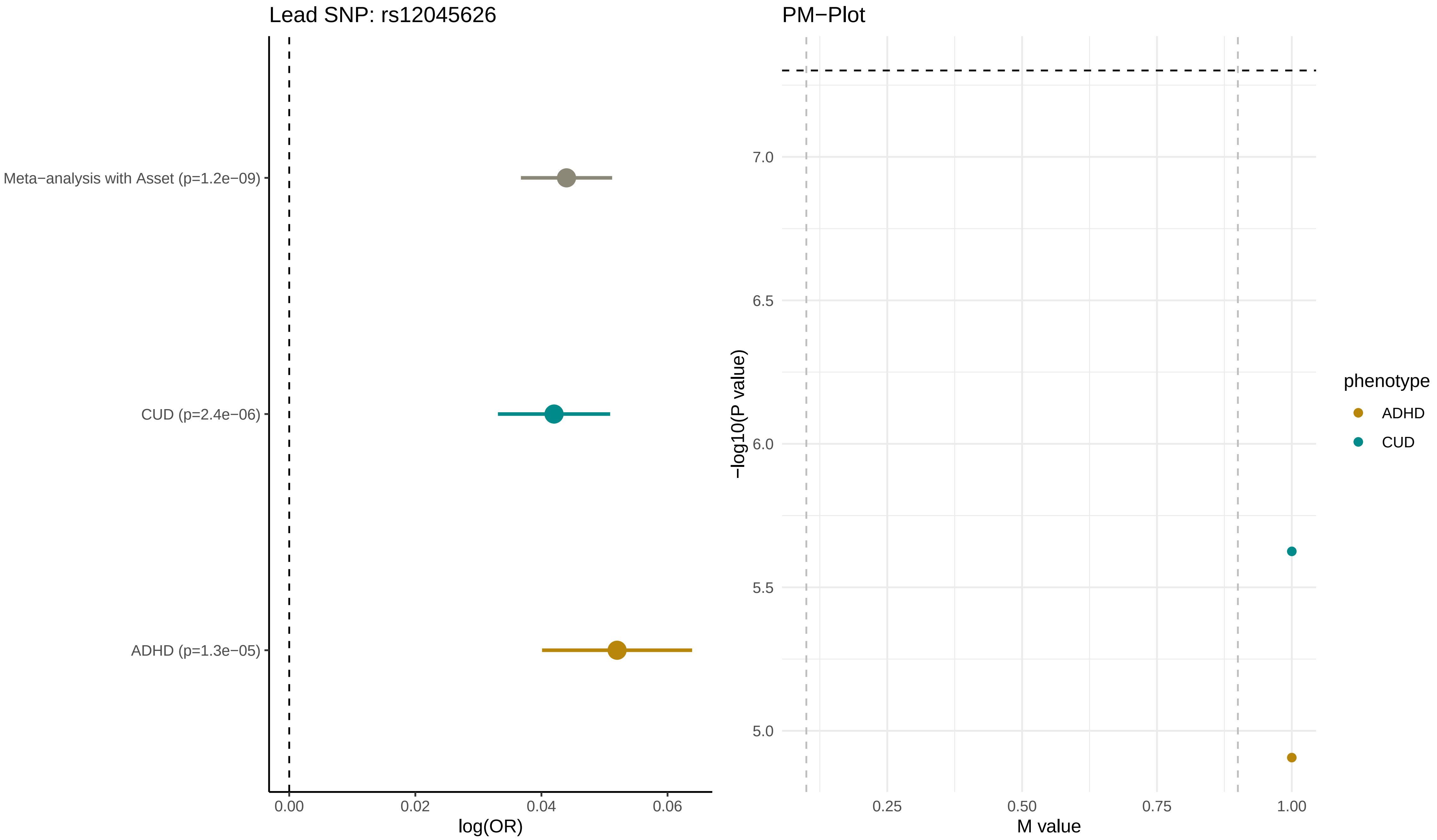
Lead SNP: rs12045626
Meta−analysis with Asset (p=1.2e−09)
CUD (p=2.4e−06)
ADHD (p=1.3e−05)
0.00
0.02
0.04
0.06
log(OR)
PM−Plot
7.0
6.5
6.0
5.5
5.0
−log10(P value)
0.25
0.50
0.75
1.00
M value
phenotype
ADHD
CUD

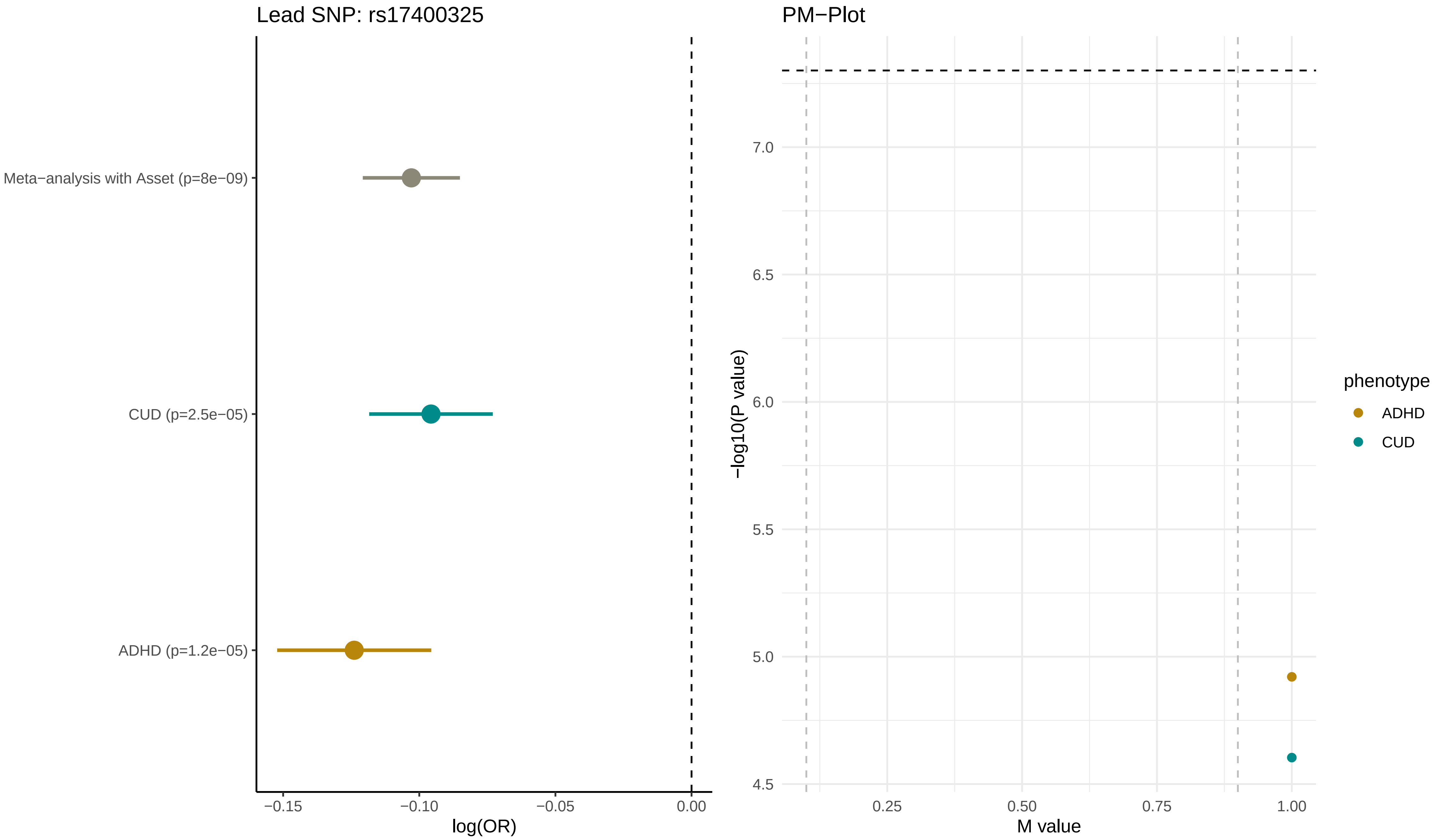
Lead SNP: rs17400325
Meta−analysis with Asset (p=8e−09)
CUD (p=2.5e−05)
ADHD (p=1.2e−05)
−0.15
−0.10
−0.05
0.00
log(OR)
PM−Plot
7.0
6.5
6.0
5.5
5.0
4.5
−log10(P value)
0.25
0.50
0.75
1.00
M value
phenotype
ADHD
CUD

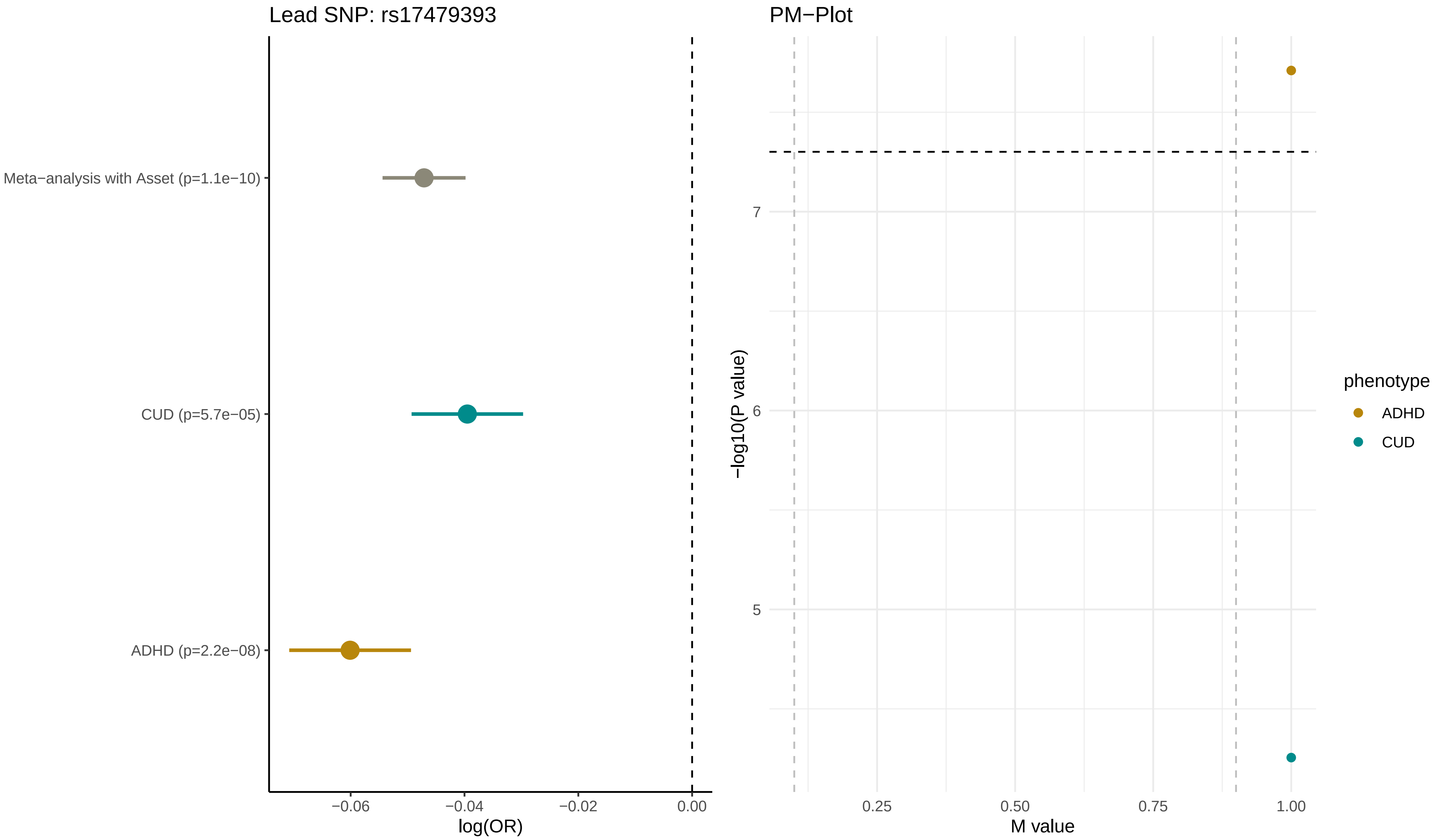

Lead SNP: rs17479393
Meta−analysis with Asset (p=1.1e−10)
CUD (p=5.7e−05)
ADHD (p=2.2e−08)
−0.06
−0.04
−0.02
0.00
log(OR)
PM−Plot
−log10(P value)
7
6
5
0.25
0.50
0.75
1.00
M value
phenotype
ADHD
CUD

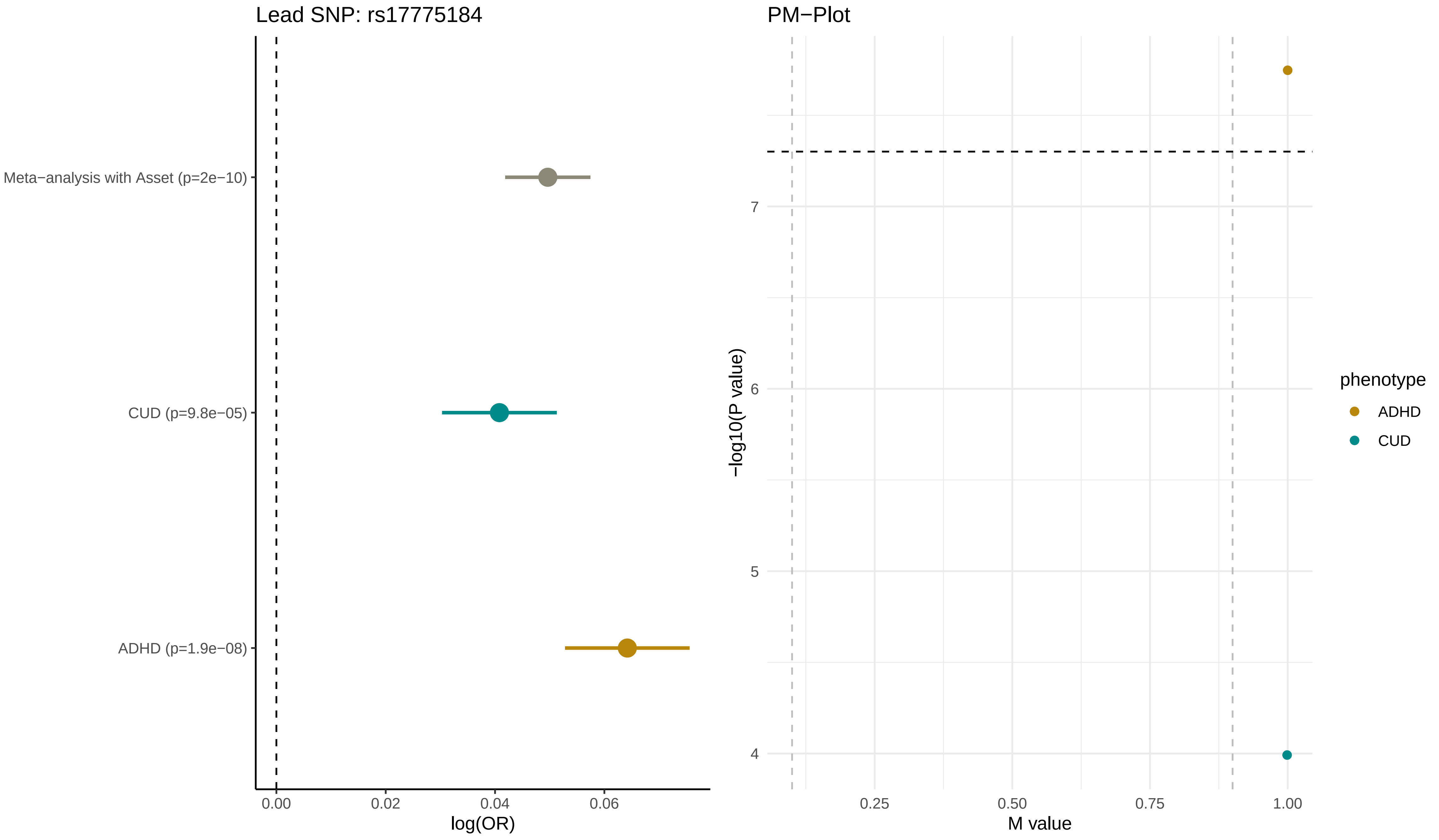

Lead SNP: rs17775184
Meta−analysis with Asset (p=2e−10)
CUD (p=9.8e−05)
ADHD (p=1.9e−08)
0.00
0.02
0.04
0.06
log(OR)
PM−Plot
−log10(P value)
7
6
5
4
0.25
0.50
0.75
1.00
M value
phenotype
ADHD
CUD

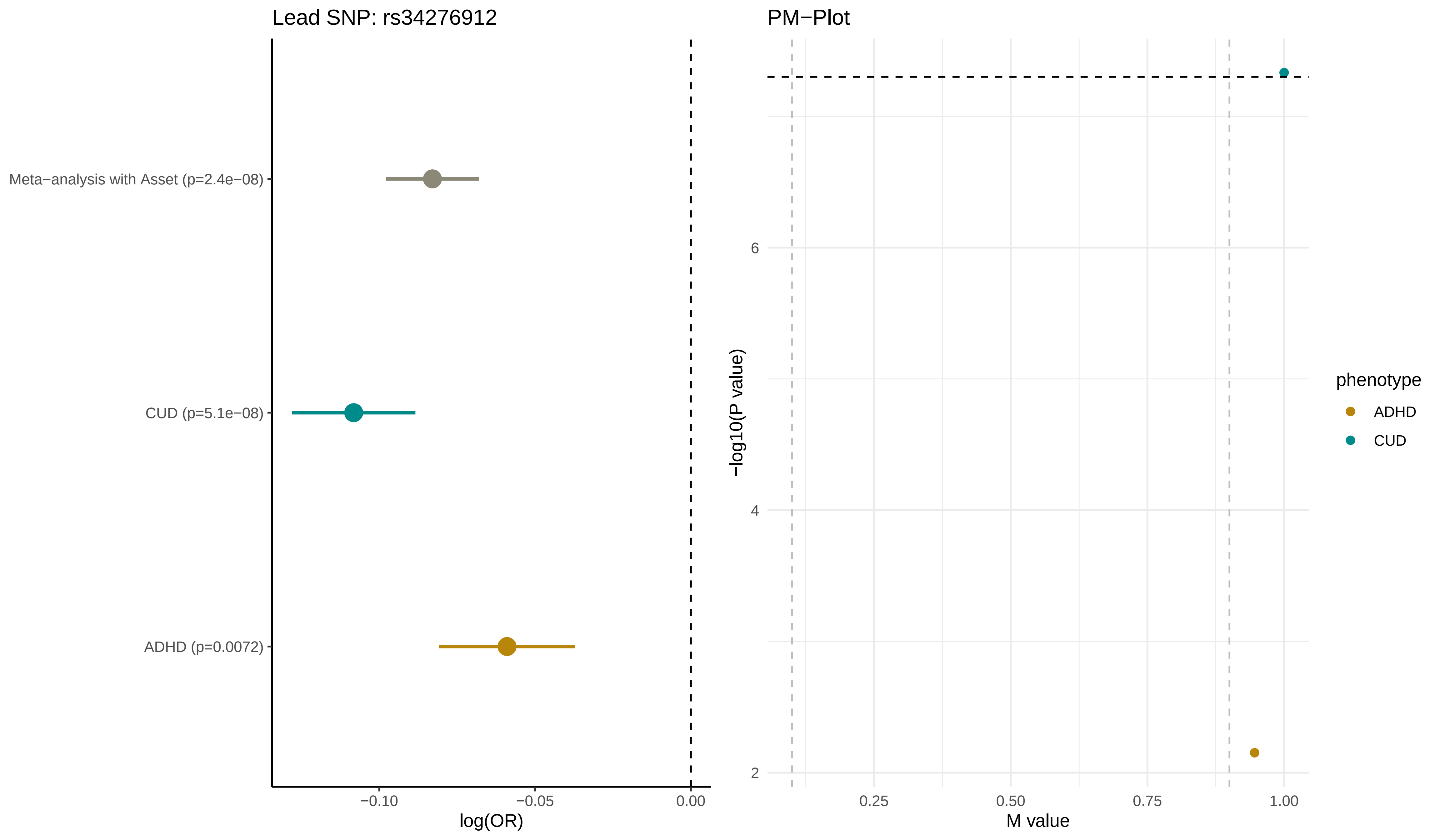

Lead SNP: rs34276912
Meta−analysis with Asset (p=2.4e−08)
CUD (p=5.1e−08)
ADHD (p=0.0072)
−0.10
−0.05
0.00
log(OR)
PM−Plot
−log10(P value)
6
4
2
0.25
0.50
0.75
1.00
M value
phenotype
ADHD
CUD

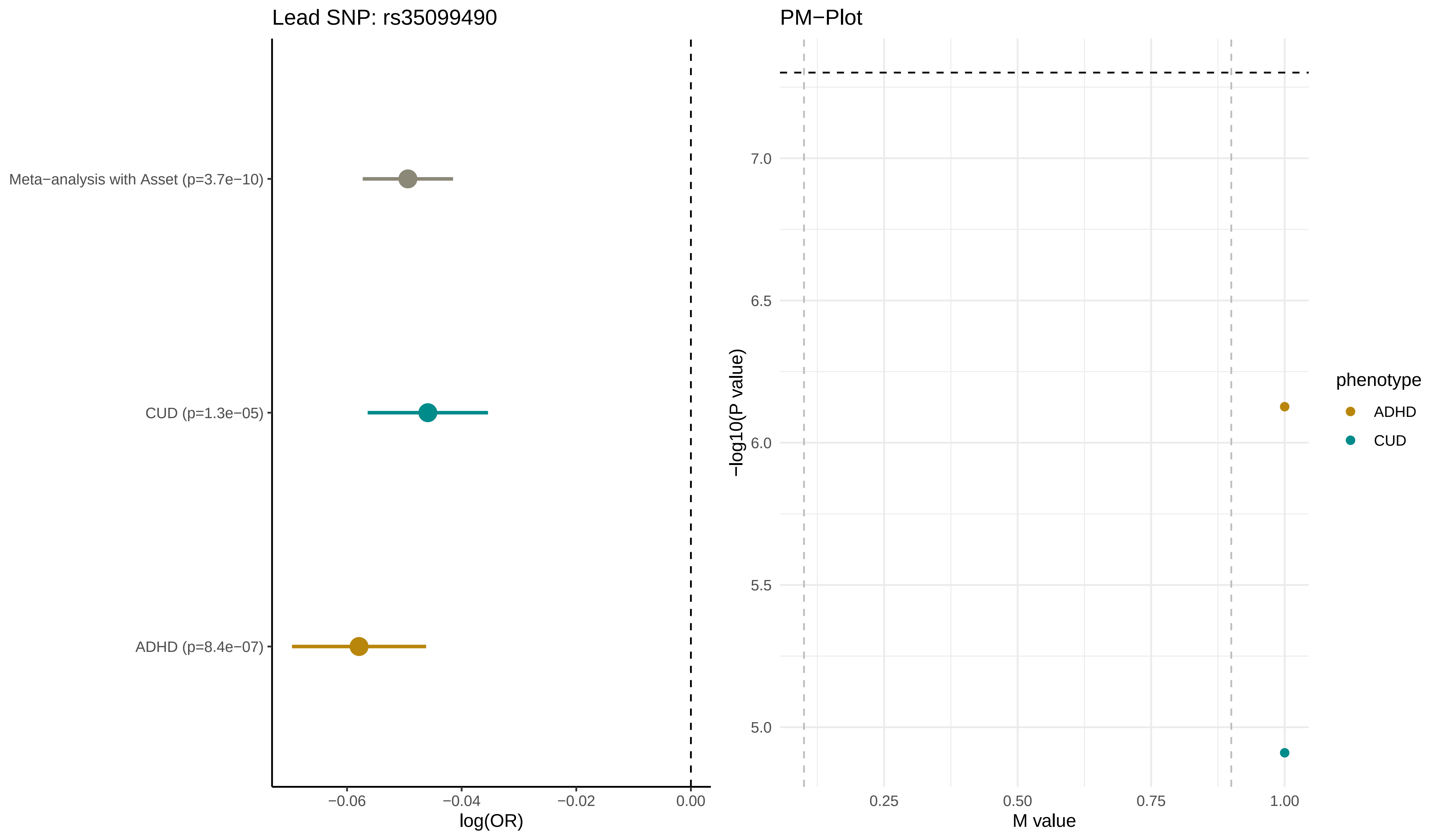

Lead SNP: rs35099490
Meta−analysis with Asset (p=3.7e−10)
CUD (p=1.3e−05)
ADHD (p=8.4e−07)
−0.06
−0.04
−0.02
0.00
log(OR)
PM−Plot
7.0
6.5
6.0
5.5
5.0
−log10(P value)
0.25
0.50
0.75
1.00
M value
phenotype
ADHD
CUD

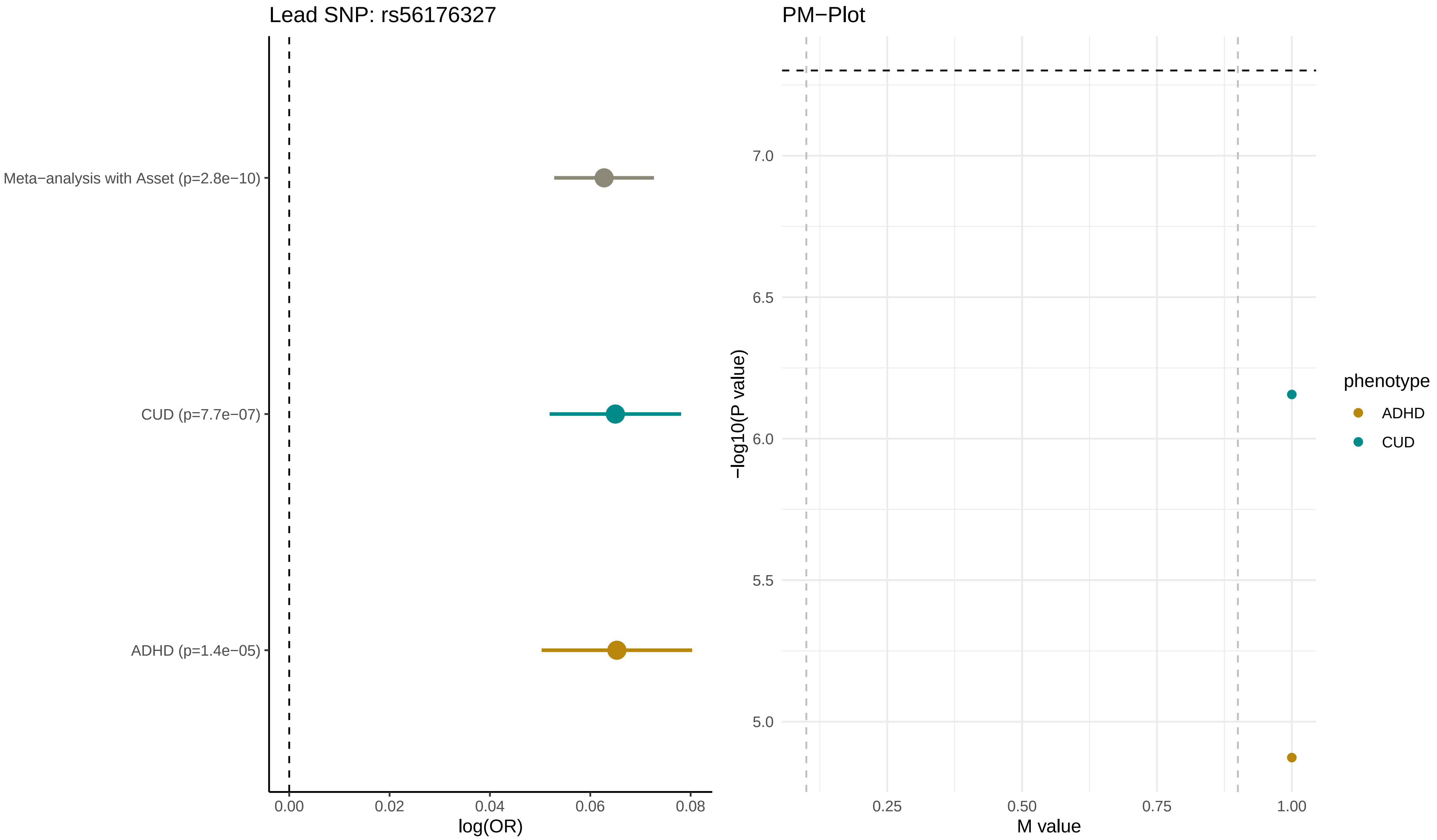

Lead SNP: rs56176327
Meta−analysis with Asset (p=2.8e−10)
CUD (p=7.7e−07)
ADHD (p=1.4e−05)
0.00
0.02
0.04
0.06
0.08
log(OR)
PM−Plot
−log10(P value)
7.0
6.5
6.0
5.5
5.0
0.25
0.50
0.75
1.00
M value
phenotype
ADHD
CUD

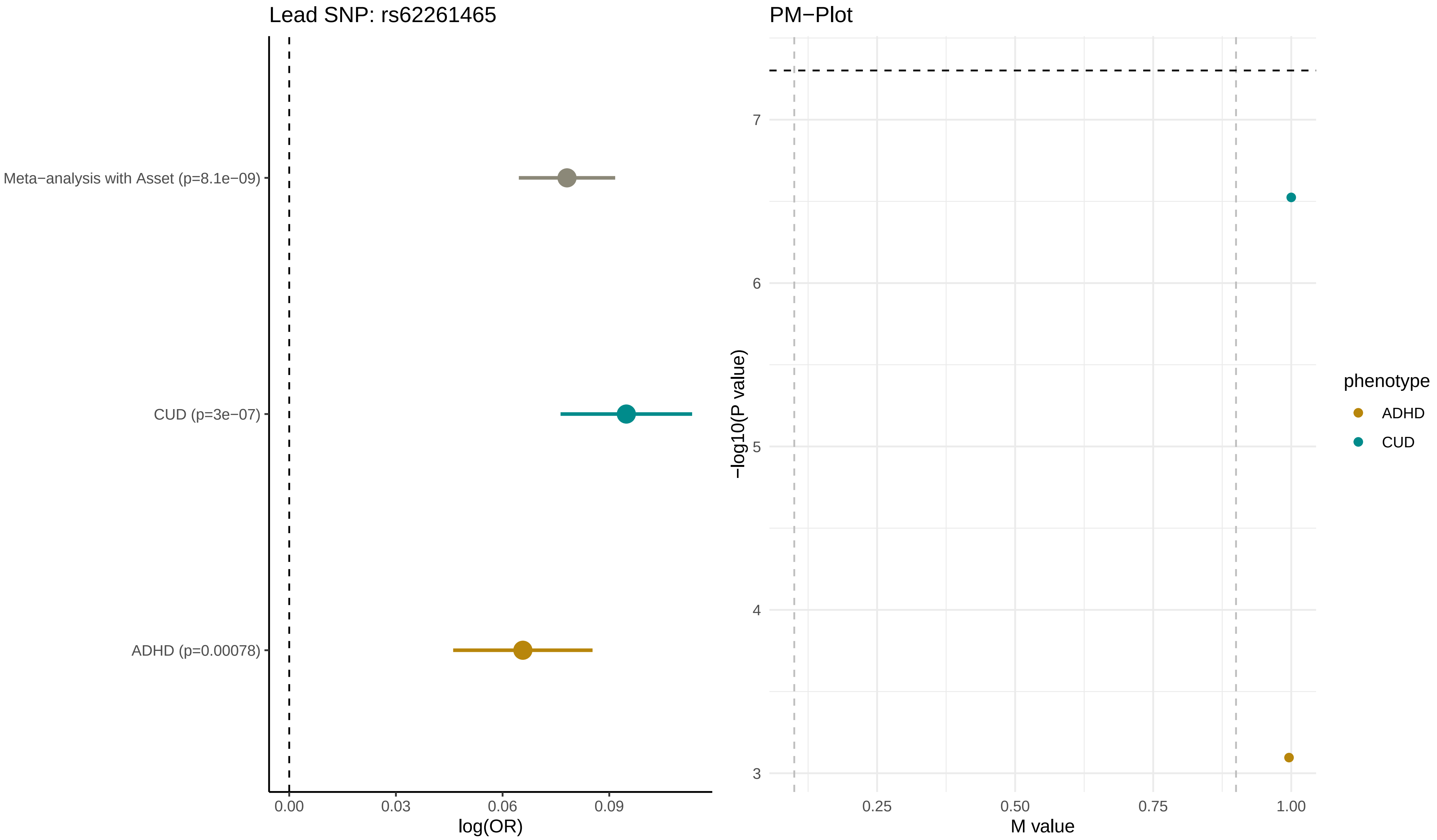

Lead SNP: rs62261465
Meta−analysis with Asset (p=8.1e−09)
CUD (p=3e−07)
ADHD (p=0.00078)
0.00
0.03
0.06
0.09
log(OR)
PM−Plot
7
6
5
4
3
−log10(P value)
0.25
0.50
0.75
1.00
M value
phenotype
ADHD
CUD

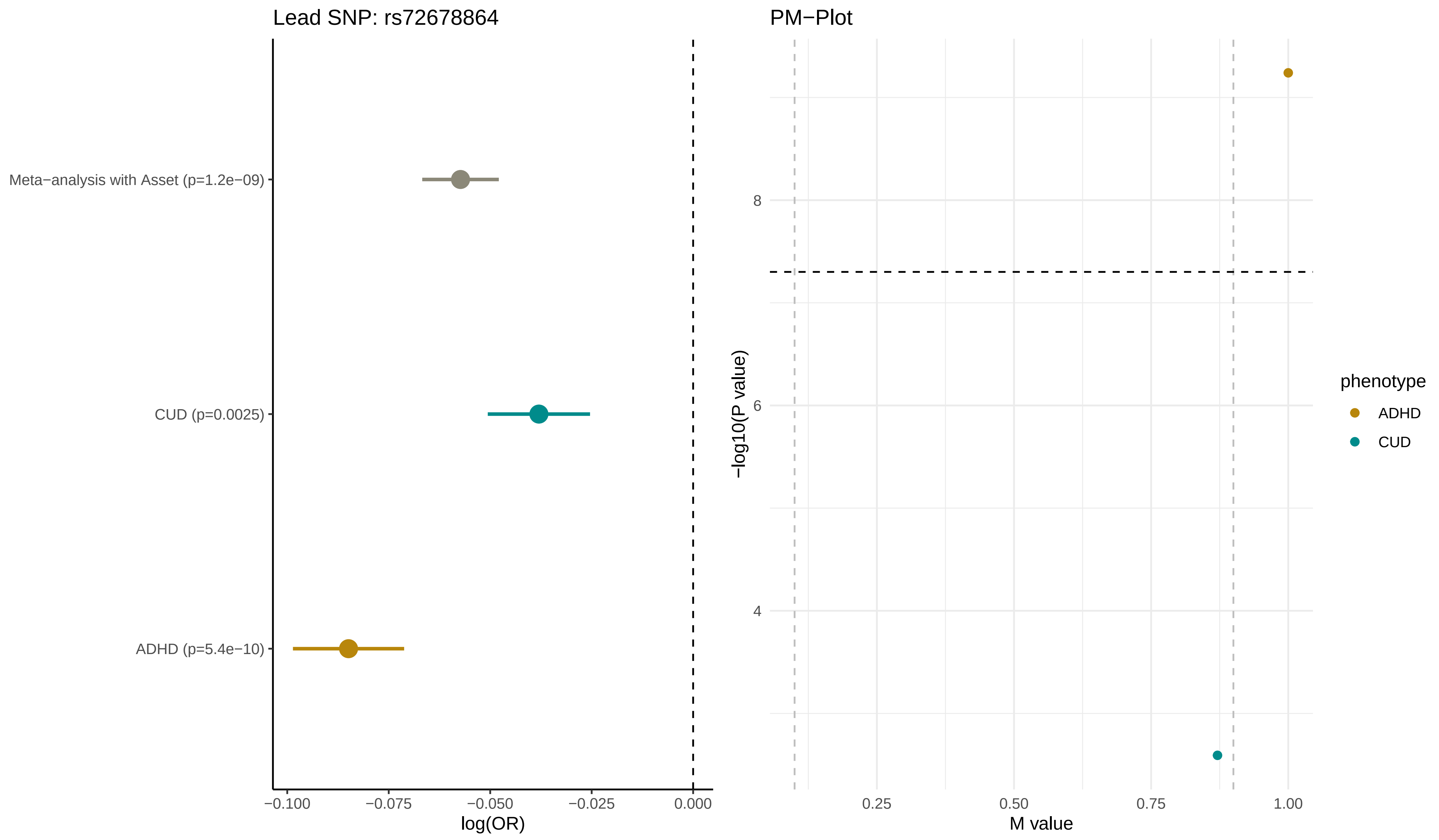

Lead SNP: rs72678864
Meta−analysis with Asset (p=1.2e−09)
CUD (p=0.0025)
ADHD (p=5.4e−10)
−0.100
−0.075
−0.050
−0.025
0.000
log(OR)
PM−Plot
8
6
4
−log10(P value)
0.25
0.50
0.75
1.00
M value
phenotype
ADHD
CUD

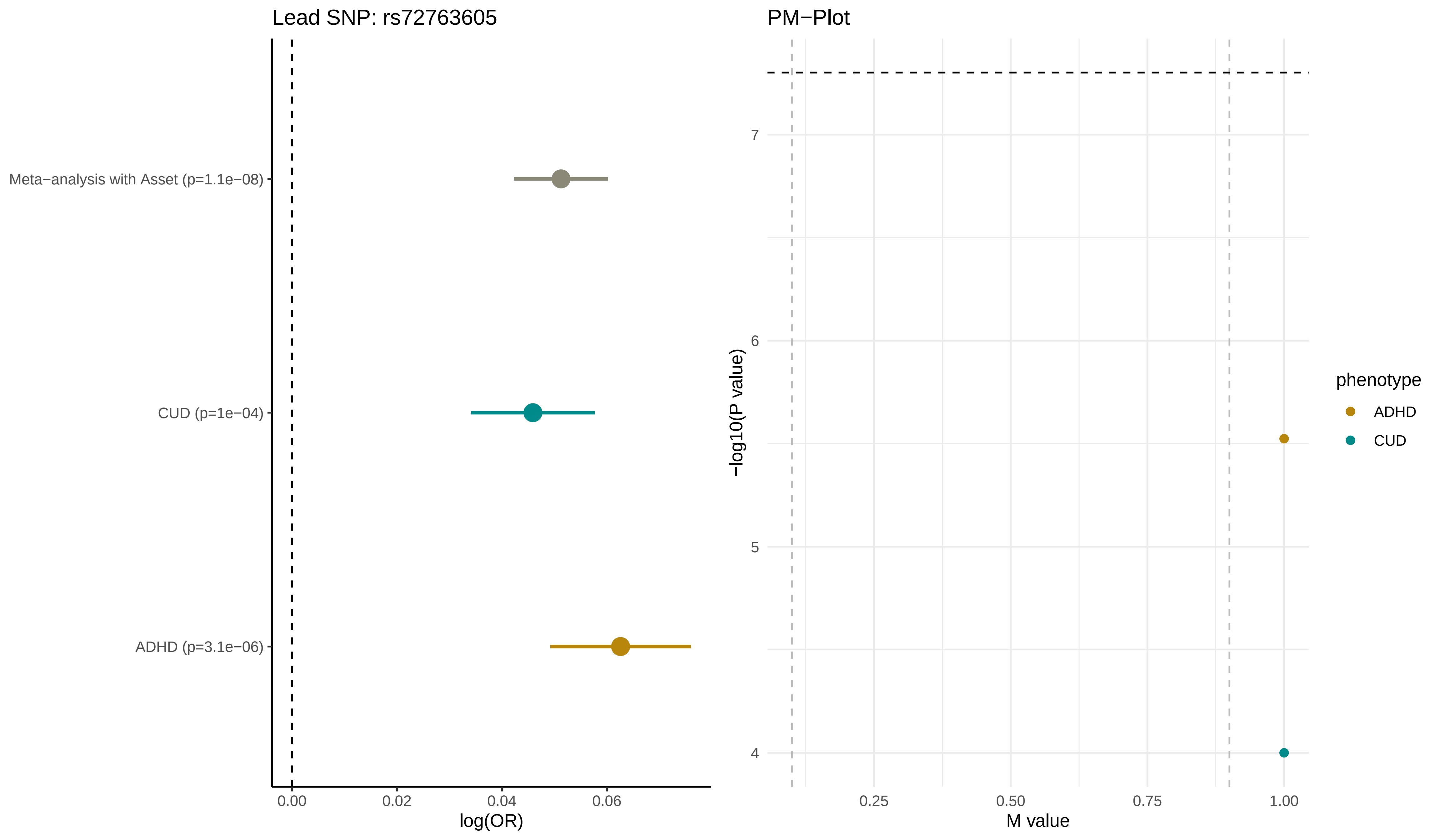

Lead SNP: rs72763605
Meta−analysis with Asset (p=1.1e−08)
CUD (p=1e−04)
ADHD (p=3.1e−06)
0.00
0.02
0.04
0.06
log(OR)
PM−Plot
7
6
5
4
−log10(P value)
0.25
0.50
0.75
1.00
M value
phenotype
ADHD
CUD

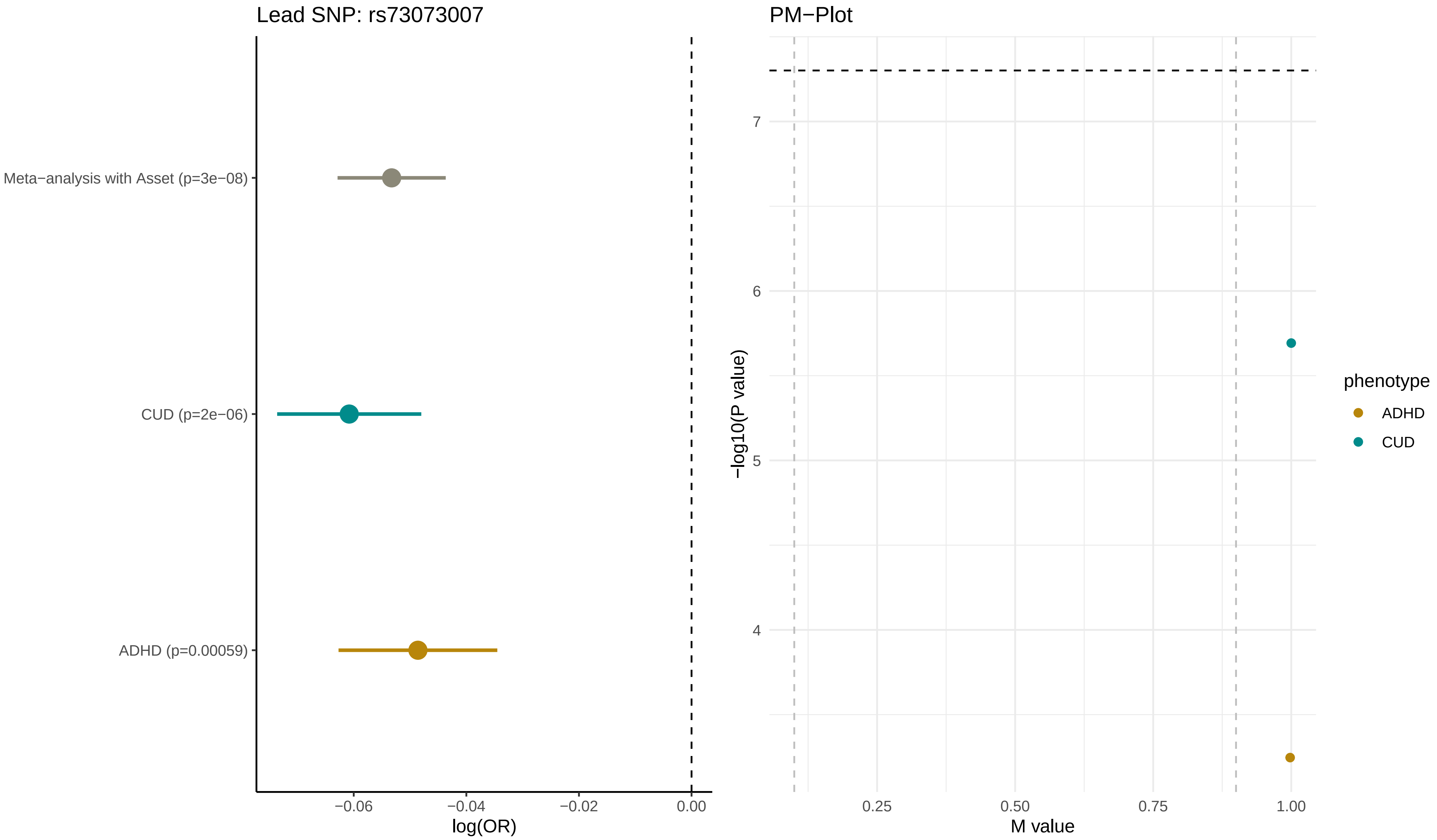

Lead SNP: rs73073007
Meta−analysis with Asset (p=3e−08)
CUD (p=2e−06)
ADHD (p=0.00059)
−0.06
−0.04
−0.02
0.00
log(OR)
PM−Plot
7
6
5
4
−log10(P value)
0.25
0.50
0.75
1.00
M value
phenotype
ADHD
CUD

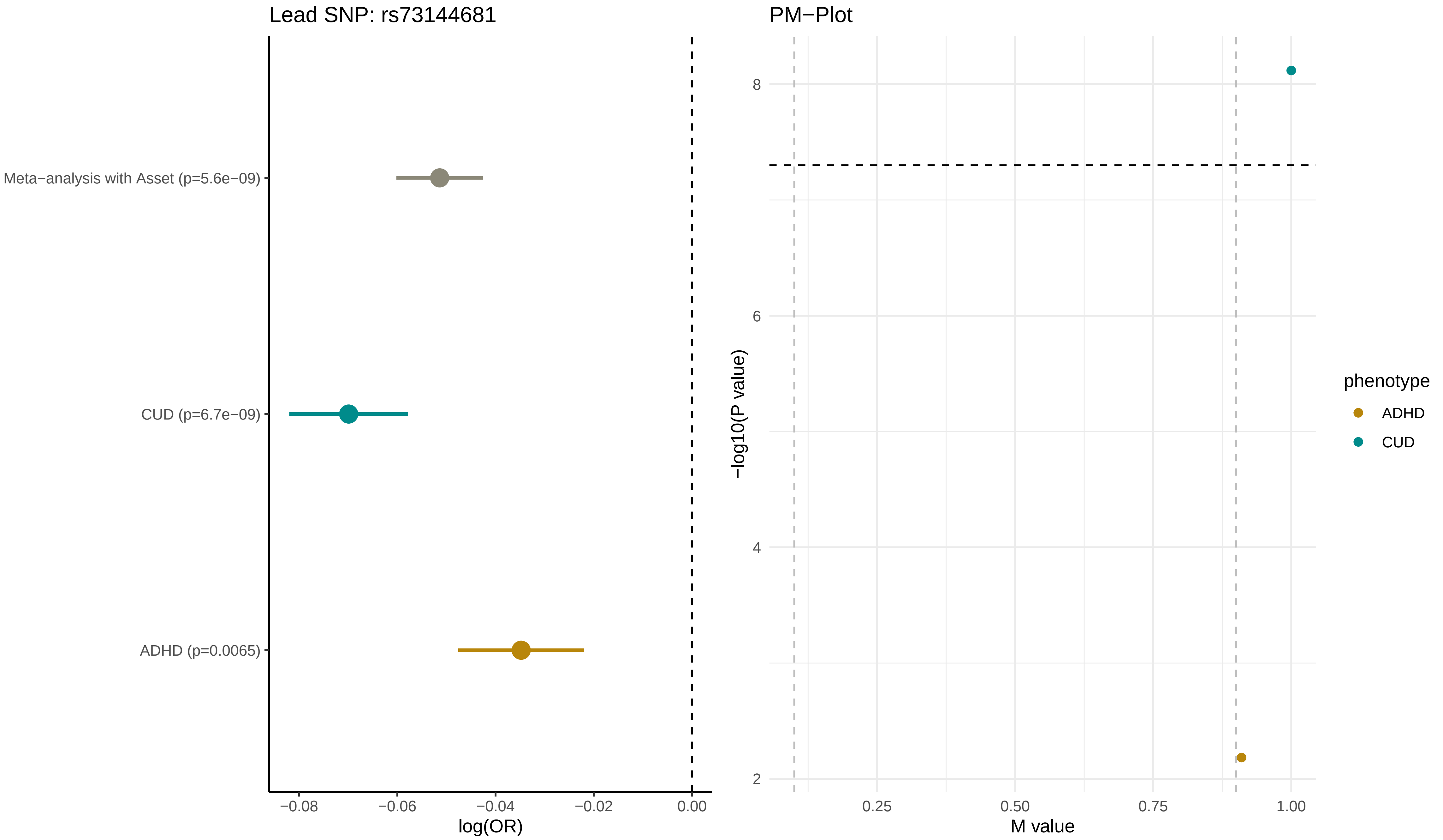

Lead SNP: rs73144681
Meta−analysis with Asset (p=5.6e−09)
CUD (p=6.7e−09)
ADHD (p=0.0065)
−0.08
−0.06
−0.04
−0.02
0.00
log(OR)
PM−Plot
8
6
4
2
−log10(P value)
0.25
0.50
0.75
1.00
M value
phenotype
ADHD
CUD

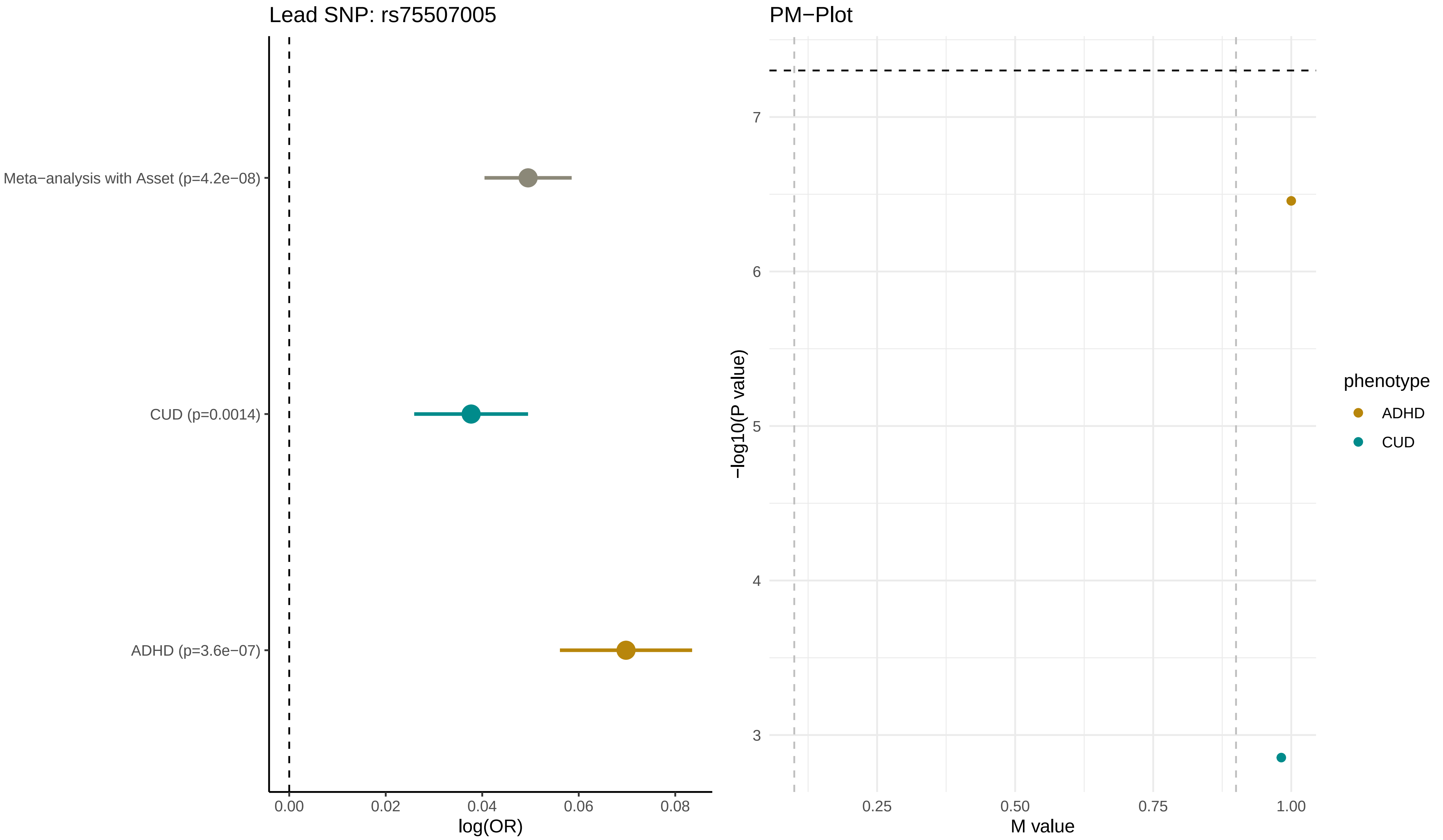

Lead SNP: rs75507005
Meta−analysis with Asset (p=4.2e−08)
CUD (p=0.0014)
ADHD (p=3.6e−07)
0.00
0.02
0.04
0.06
0.08
log(OR)
PM−Plot
7
6
5
4
3
−log10(P value)
0.25
0.50
0.75
1.00
M value
phenotype
ADHD
CUD
