## Supplementary _Information for "Disentangling the shared genetics of ADHD, cannabis use disorder and cannabis use and prediction of cannabis use disorder in ADHD"

**Supplementary Note**

Functional genomics datasets implemented in FUMA used to link ADHD-CUD and ADHD-CU variants to genes using the SNP2GENE function:

**posMapAnnoDs;**

PsychENCODE/enhancer.bed.gz, PsychENCODE/enhancer_high_conf.bed.gz, PsychENCODE/PFC_H3K27ac_peak.bed.gz, PsychENCODE/TC_H3K27ac_peak.bed.gz, PsychENCODE/CBC_H3K27ac_peak.bed.gz, PsychENCODE/TARs.bed.gz, BOCA/DLPFC_neuron.bed.gz, BOCA/DLPFC_glia.bed.gz, BOCA/OFC_neuron.bed.gz, BOCA/OFC_glia.bed.gz, BOCA/VLPFC_neuron.bed.gz, BOCA/VLPFC_glia.bed.gz, BOCA/ACC_neuron.bed.gz, BOCA/ACC_glia.bed.gz, BOCA/STC_neuron.bed.gz, BOCA/STC_glia.bed.gz, BOCA/ITC_neuron.bed.gz, BOCA/ITC_glia.bed.gz, BOCA/PMC_neuron.bed.gz, BOCA/PMC_glia.bed.gz, BOCA/INS_neuron.bed.gz, BOCA/INS_glia.bed.gz, BOCA/PVC_neuron.bed.gz, BOCA/PVC_glia.bed.gz, BOCA/AMY_neuron.bed.gz, BOCA/AMY_glia.bed.gz, BOCA/HIPP_neuron.bed.gz, BOCA/HIPP_glia.bed.gz, BOCA/MDT_neuron.bed.gz, BOCA/MDT_glia.bed.gz, BOCA/NAC_neuron.bed.gz, BOCA/NAC_glia.bed.gz, BOCA/PUT_neuron.bed.gz, BOCA/PUT_glia.bed.gz

**eqtlMaptss:**

PsychENCODE/PsychENCODE_eQTLs.txt.gz, CMC/CMC_SVA_cis.txt.gz, CMC/CMC_SVA_trans.txt.gz, CMC/CMC_NoSVA_cis.txt.gz, CMC/CMC_NoSVA_trans.txt.gz, BRAINEAC/CRBL.txt.gz, BRAINEAC/FCTX.txt.gz, BRAINEAC/HIPP.txt.gz, BRAINEAC/MEDU.txt.gz, BRAINEAC/OCTX.txt.gz, BRAINEAC/PUTM.txt.gz, BRAINEAC/SNIG.txt.gz, BRAINEAC/TCTX.txt.gz, BRAINEAC/THAL.txt.gz, BRAINEAC/WHMT.txt.gz, BRAINEAC/aveALL.txt.gz, GTEx/v8/Brain_Amygdala.txt.gz, GTEx/v8/Brain_Anterior_cingulate_cortex_BA24.txt.gz, GTEx/v8/Brain_Caudate_basal_ganglia.txt.gz, GTEx/v8/Brain_Cerebellar_Hemisphere.txt.gz, GTEx/v8/Brain_Cerebellum.txt.gz, GTEx/v8/Brain_Cortex.txt.gz, GTEx/v8/Brain_Frontal_Cortex_BA9.txt.gz, GTEx/v8/Brain_Hippocampus.txt.gz, GTEx/v8/Brain_Hypothalamus.txt.gz, GTEx/v8/Brain_Nucleus_accumbens_basal_ganglia.txt.gz, GTEx/v8/Brain_Putamen_basal_ganglia.txt.gz, GTEx/v8/Brain_Spinal_cord_cervical_c-1.txt.gz, GTEx/v8/Brain_Substantia_nigra.txt.gz

**eqtlMapAnnoDs:**

PsychENCODE/enhancer.bed.gz, PsychENCODE/enhancer_high_conf.bed.gz, PsychENCODE/PFC_H3K27ac_peak.bed.gz, PsychENCODE/TC_H3K27ac_peak.bed.gz, PsychENCODE/CBC_H3K27ac_peak.bed.gz, PsychENCODE/TARs.bed.gz, BOCA/DLPFC_neuron.bed.gz, BOCA/DLPFC_glia.bed.gz, BOCA/OFC_neuron.bed.gz, BOCA/OFC_glia.bed.gz, BOCA/VLPFC_neuron.bed.gz, BOCA/VLPFC_glia.bed.gz, BOCA/ACC_neuron.bed.gz, BOCA/ACC_glia.bed.gz, BOCA/STC_neuron.bed.gz, BOCA/STC_glia.bed.gz, BOCA/ITC_neuron.bed.gz, BOCA/ITC_glia.bed.gz, BOCA/PMC_neuron.bed.gz, BOCA/PMC_glia.bed.gz, BOCA/INS_neuron.bed.gz, BOCA/INS_glia.bed.gz, BOCA/PVC_neuron.bed.gz, BOCA/PVC_glia.bed.gz, BOCA/AMY_neuron.bed.gz, BOCA/AMY_glia.bed.gz, BOCA/HIPP_neuron.bed.gz, BOCA/HIPP_glia.bed.gz, BOCA/MDT_neuron.bed.gz, BOCA/MDT_glia.bed.gz, BOCA/NAC_neuron.bed.gz, BOCA/NAC_glia.bed.gz, BOCA/PUT_neuron.bed.gz, BOCA/PUT_glia.bed.gz

**ciMapBuiltin:**

EP/PsychENCODE/EP_links_oneway.txt.gz, HiC/PsychENCODE/Promoter_anchored_loops.txt.gz, HiC/Giusti-Rodriguez_et_al_2019/Adult_Cortex.txt.gz, HiC/Giusti-Rodriguez_et_al_2019/Fetal_Cortex.txt.gz, HiC/GSE87112/Dorsolateral_Prefrontal_Cortex.txt.gz, HiC/GSE87112/Hippocampus.txt.gz

**ciMapRoadmap:**

E053, E054, E067, E068, E069, E070, E071, E072, E073, E074, E081, E082

**ciMapAnnoDs;**

PsychENCODE/enhancer.bed.gz, PsychENCODE/enhancer_high_conf.bed.gz, PsychENCODE/PFC_H3K27ac_peak.bed.gz, PsychENCODE/TC_H3K27ac_peak.bed.gz, PsychENCODE/CBC_H3K27ac_peak.bed.gz, PsychENCODE/TARs.bed.gz, BOCA/DLPFC_neuron.bed.gz, BOCA/DLPFC_glia.bed.gz, BOCA/OFC_neuron.bed.gz, BOCA/OFC_glia.bed.gz, BOCA/VLPFC_neuron.bed.gz, BOCA/VLPFC_glia.bed.gz, BOCA/ACC_neuron.bed.gz, BOCA/ACC_glia.bed.gz, BOCA/STC_neuron.bed.gz, BOCA/STC_glia.bed.gz, BOCA/ITC_neuron.bed.gz, BOCA/ITC_glia.bed.gz, BOCA/PMC_neuron.bed.gz, BOCA/PMC_glia.bed.gz, BOCA/INS_neuron.bed.gz, BOCA/INS_glia.bed.gz, BOCA/PVC_neuron.bed.gz, BOCA/PVC_glia.bed.gz, BOCA/AMY_neuron.bed.gz, BOCA/AMY_glia.bed.gz, BOCA/HIPP_neuron.bed.gz, BOCA/HIPP_glia.bed.gz, BOCA/MDT_neuron.bed.gz, BOCA/MDT_glia.bed.gz, BOCA/NAC_neuron.bed.gz, BOCA/NAC_glia.bed.gz, BOCA/PUT_neuron.bed.gz, BOCA/PUT_glia.bed.gz

**Supplementary Figures**

**Supplementary Figure 1. Common factor model using Genomic SEM**

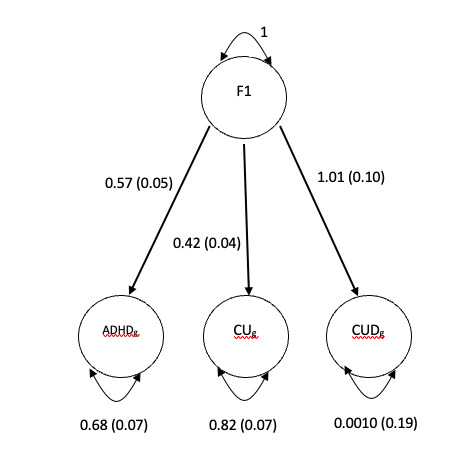

Standardized results from using Genomic SEM (with diagonally weighted least square estimation) to construct a genetically defined latent factor shared by ADHD (38,691 ADHD; 186,843 controls), CUD (42,281 CUD; 843,744 controls) and CU (162,082 individuals). Loadings for each phenotype is shown together with standard errors in parentheses next to the arrows. Residual variances are shown below each indicator phenotype.

**Supplementary Figure 2. Genetic mediation analysis using Genomic SEM**

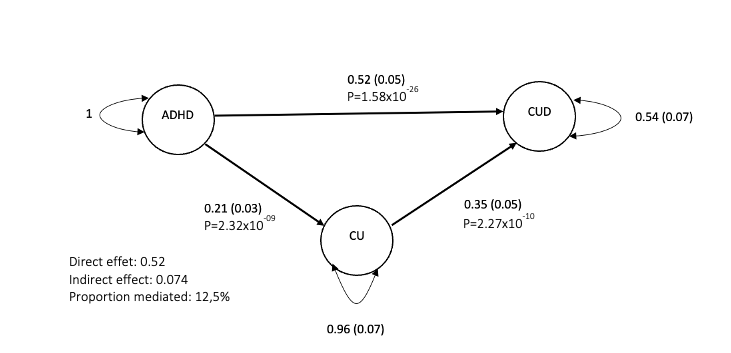

Estimation of the direct and CU-mediated effect of ADHD genetics on CUD. Regression of the genetically derived variance covariance matrixes of CUD on ADHD, CU and estimates (diagonally weighted least squares estimates) from subsequent model fit in Genomic SEM. Standard errors in parentheses and P-values indicating effect. Samples sizes are given in the legend of Supplementary Figure 1.

**Supplementary Figure 3. Manhattan plot GWAS ADHD-CUD and ADHD-CU - concordant variants**

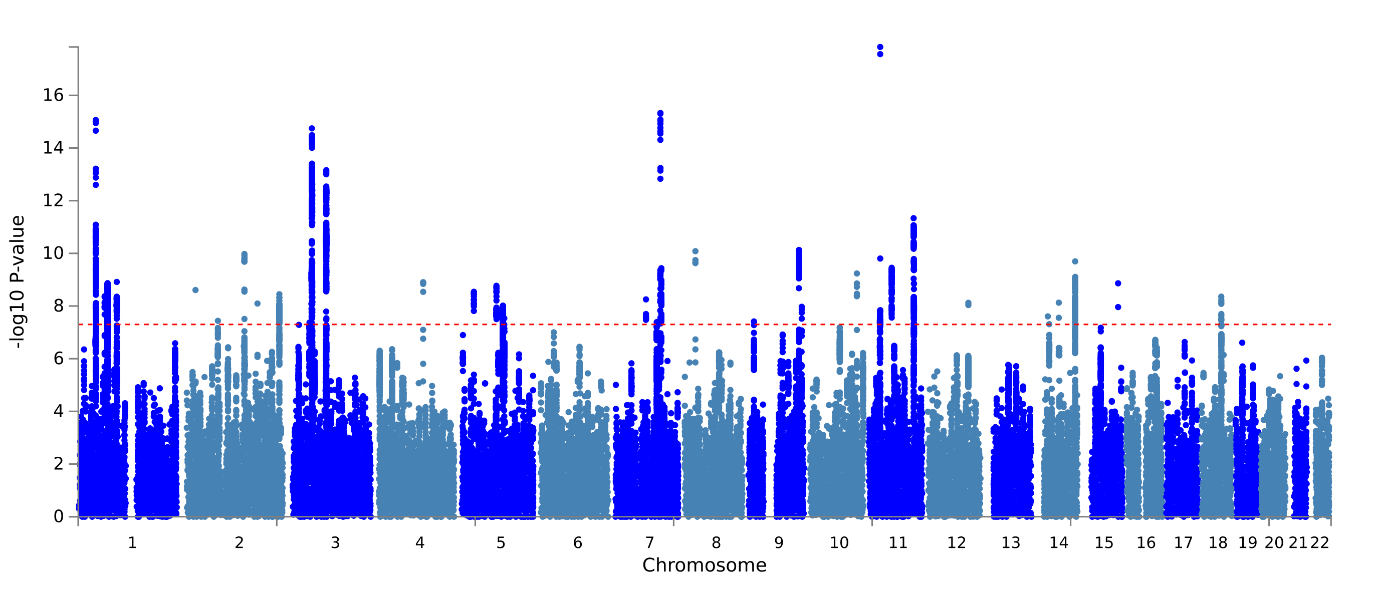

**a.**

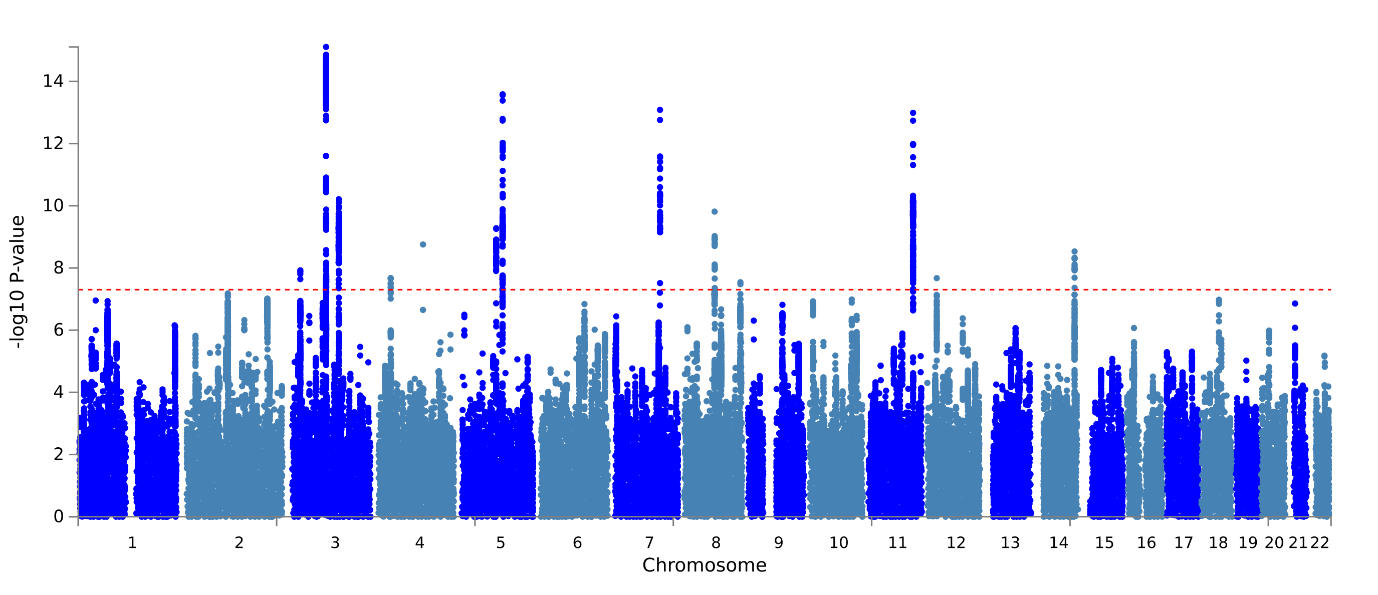

**b.**

Results from GWAS meta-analysis of **a.** ADHD-CUD (N=1,150,250) and ADHD-CU (N=387,616) **b.** for variants having alleles with concordant direction of effect on the analyzed phenotypes. On the x-axis two-sided P-values from the ASSET one-sided subset test. The red horizontal line represents the threshold for genome-wide significant association (P = 5x10^-8^).

**Supplementary Figure 4. Manhattan plots, GWAS of ADHD-CUD and ADHD-CU - discordant variants**

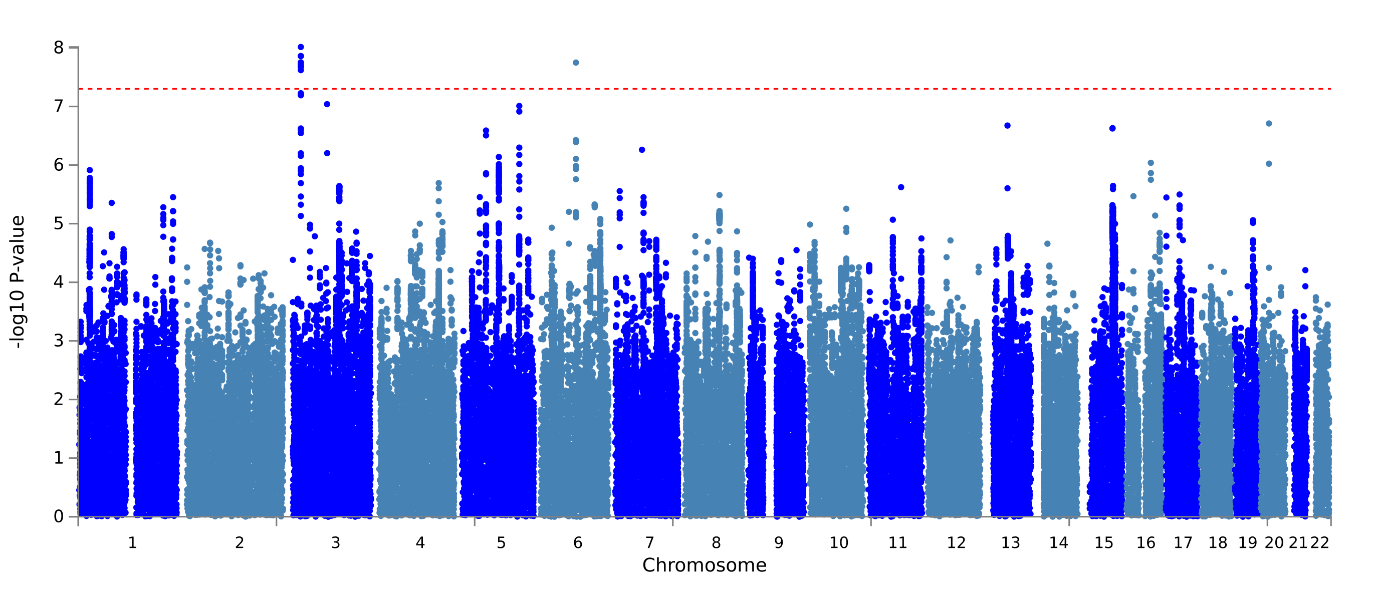

**a.**

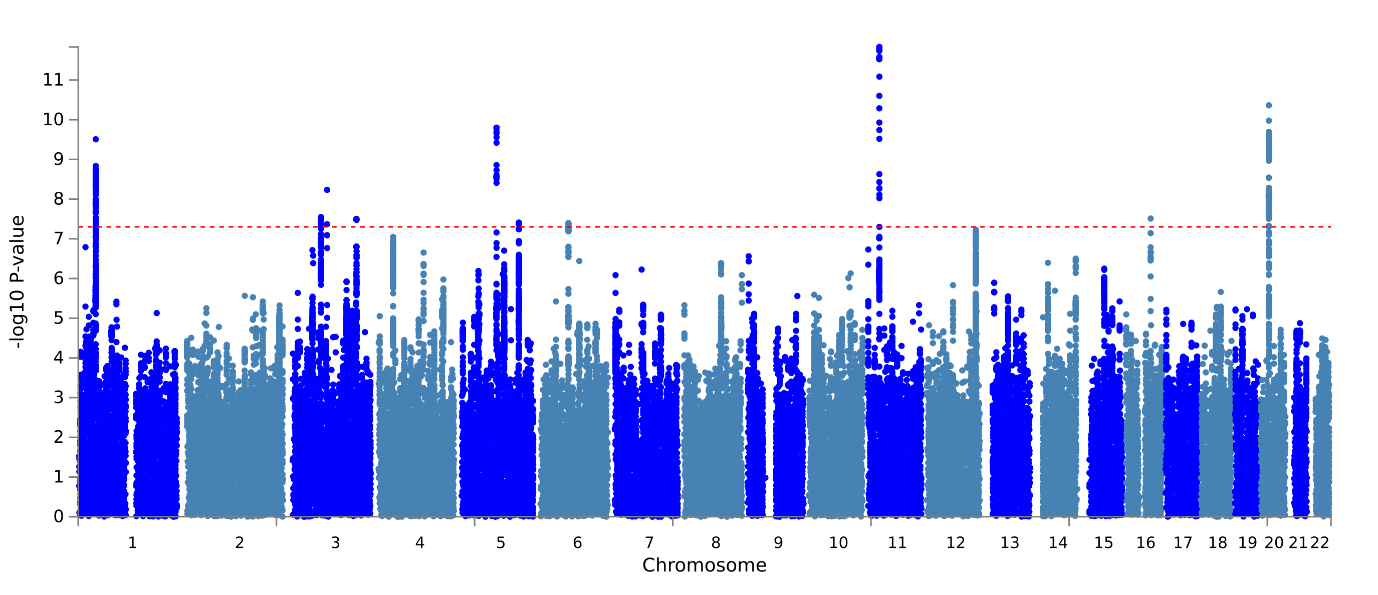

**b.**

Results from GWAS meta-analysis of **a.** ADHD-CUD (N=1,150,250) and ADHD-CU (N=387,616) **b.** for variants having alleles with discordant direction of effect on the analyzed phenotypes. On the x-axis two-sided P-values from the ASSET one-sided subset test. The red horizontal line represents the threshold for genome-wide significant association (P = 5x10^-8^).

**Supplementary Figure 5. Tissue specific gene expression of ADHD-CUD and ADHD-CU risk genes**

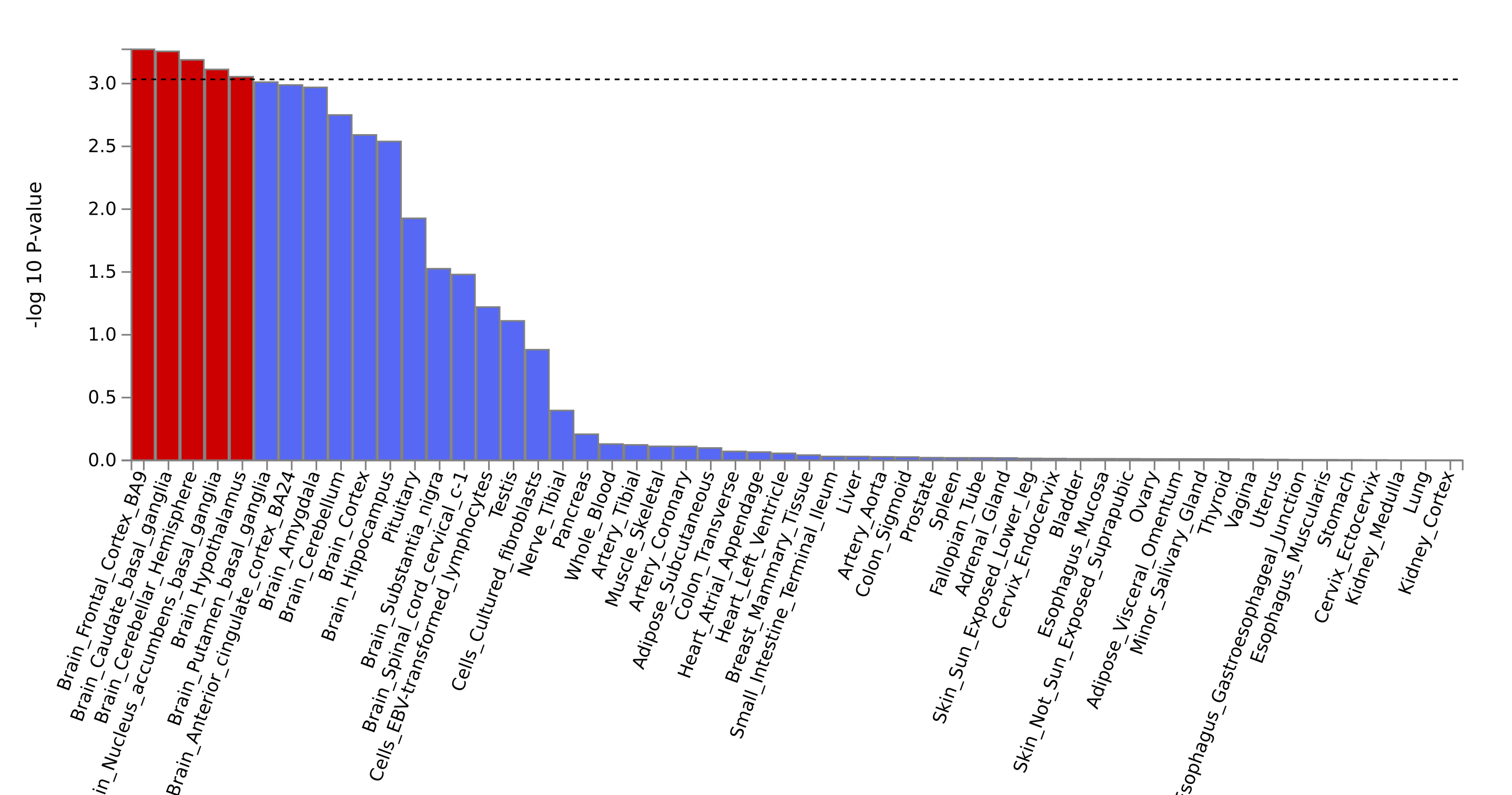

**a.**

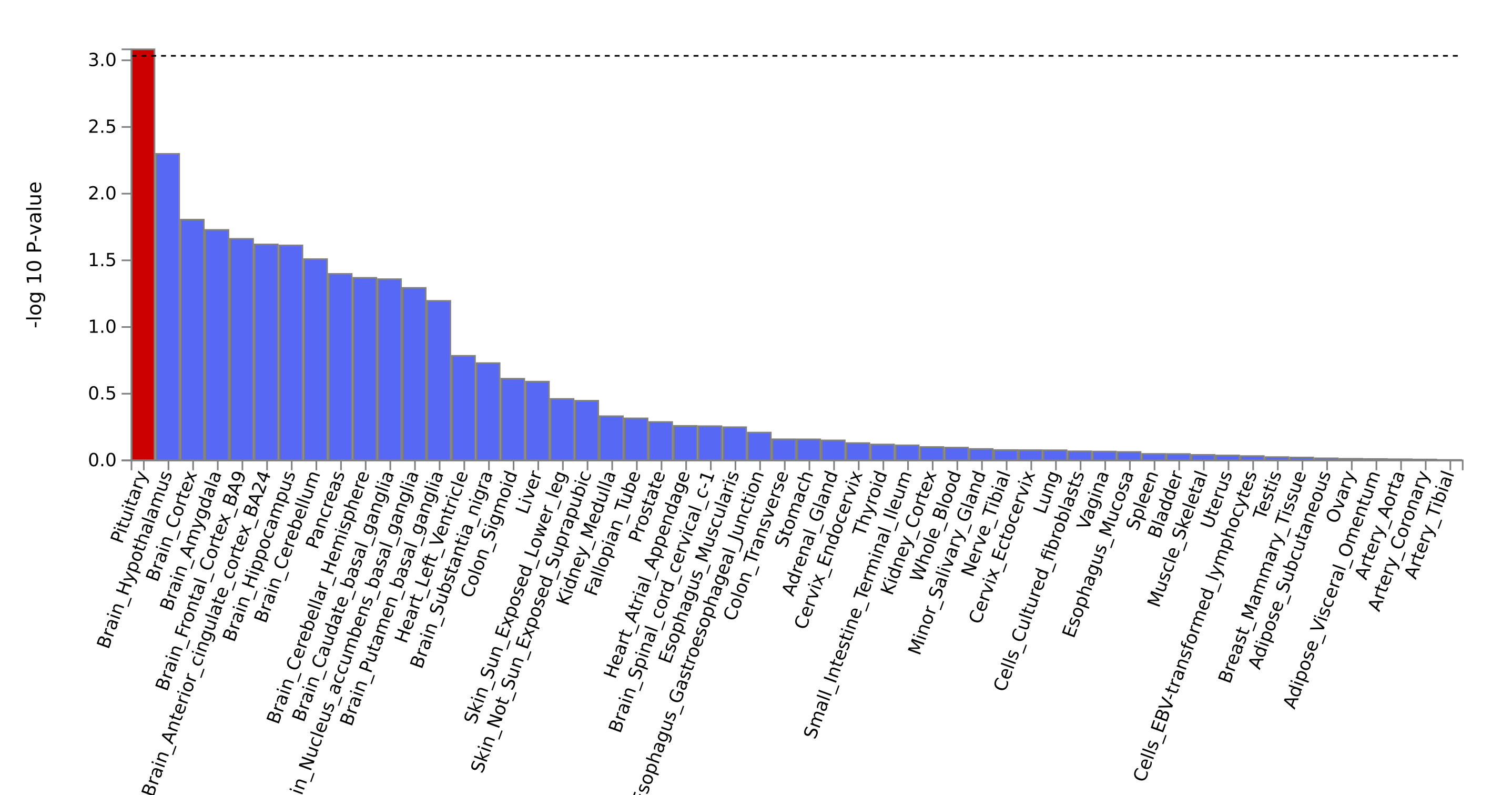

**b.**

Results (-log10 (one-sided P-value)) from MAGMA gene-property analysis of relationships between tissue specific gene expression profiles (GTEx v.7) and **a.** ADHD-CUD gene associations or **b.** ADHD-CU gene associations. The test was performed for average gene-expression per. tissue type conditioning on the average expression across all categories. Red bars indicate significant results after Bonferroni correction.

**Supplementary Figure 6. Test for difference in brain expressed genes after ADHD-CUD gene down-sampling**

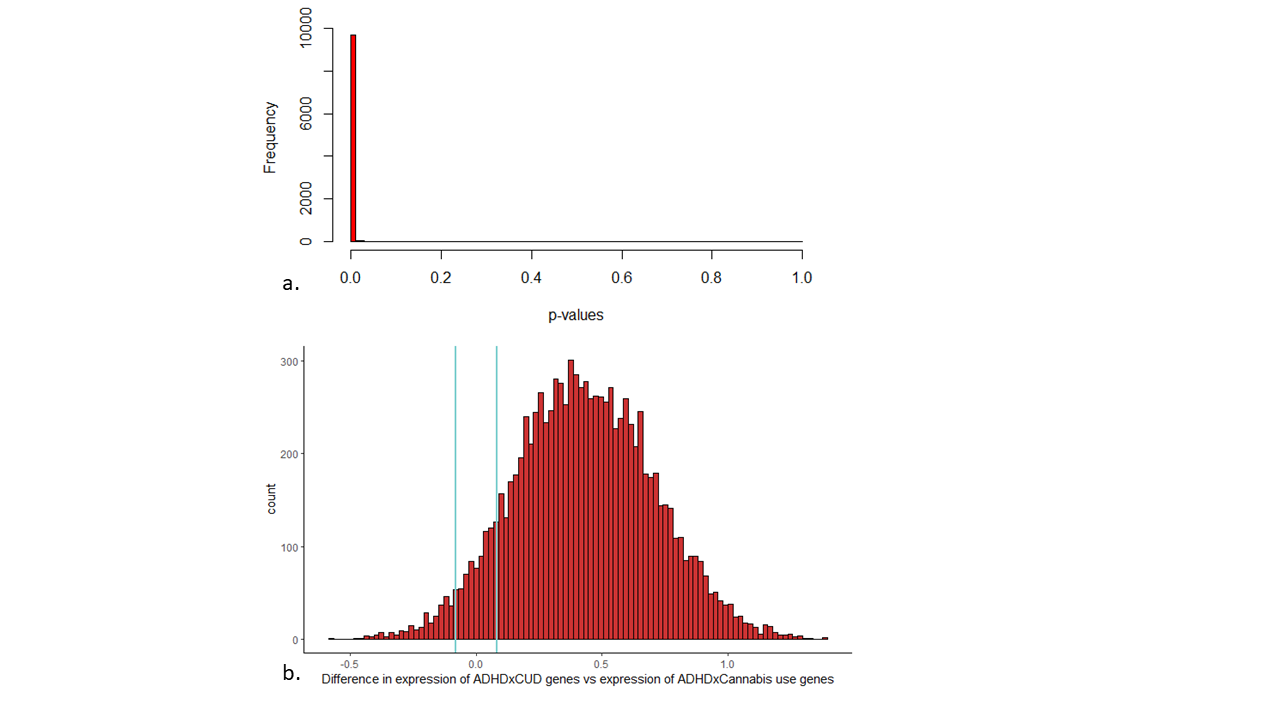

Results from simulation-test to evaluate if the significant difference in expression of ADHDxCUD genes and the ADHDxCU genes in BrianSpan data was due to power differences. The ADHD-CUD risk gene set was down-samples 10,000 times to 12 genes (similar size as the ADHD-CU risk gene set when excluding overlapping genes). A paired t-test was used to test for difference in mean expression of the down-sampled ADHD-CUD risk gene sets and the ADHD-CU gene set where each t-test was performed with 12 random selected ADHDxCUD genes against the 12 ADHDxCU genes. **a.** Distribution of the P-values from the t-test testing the 10,000 down-sampled gene-sets. **b.** counts (on the y-axis) of tests demonstrating decreased expression of ADHD-CUD genes compared to ADHD-CU genes (values below 0 on the x-axis) and tests demonstrating increased expression of ADHD-CUD genes compared to ADHD-CU genes (values above 0 on the x-axis). 361 (3.6%) of the tests demonstrated decreased expression of ADHD-CUD genes vs ADHD-CU genes; 9092 (90.9%) of the tests demonstrated increased expression of ADHD-CUD genes vs ADHD-CU genes; 547 (5.5%) of the tests demonstrated no significant difference (indicated by vertical blue lines).

**Supplementary Figure 7. Rare variant load in ADHD-CUD compared to ADHD without CUD and controls**

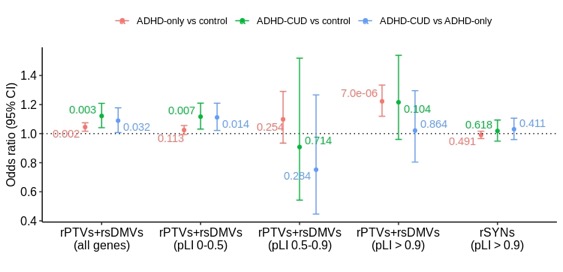

Odds ratio (and corresponding 95% confidence intervals represented by vertical lines) on the y-axis from logistic regression testing for the load of rare protein truncating variants (rPTVs) and rare severe damaging missense variants (rare SevereDMVs) in ADHD-CUD (N=333) compared to ADHD-only (N=3,483) in all genes. Values next to the vertical lines are P-values. Sub-analyses of the load of rPTVs+rare SevereDMVs in ADHD-CUD vs ADHD-only in genes stratified by their pLI score: (I) genes with high tolerance to loss of function mutations (0 < pLI < 0.5), (II) less constrained genes (0.5 < pLI < 0.9), (III) evolutionarily constrained genes (pLI > 0.9). For the three sub-analysis P < 0.0167 is considered significant.

For comparison results from testing for the load of rPTVs+rare SevereDMVs in ADHD-CUD vs controls (N=8,951) and in ADHD-only (N=3,483) vs controls (See also Supplementary Table 21).

**Supplementary Figure 8. Sex-stratified PGS analyses**

- ADHD
- ADHD+CUD

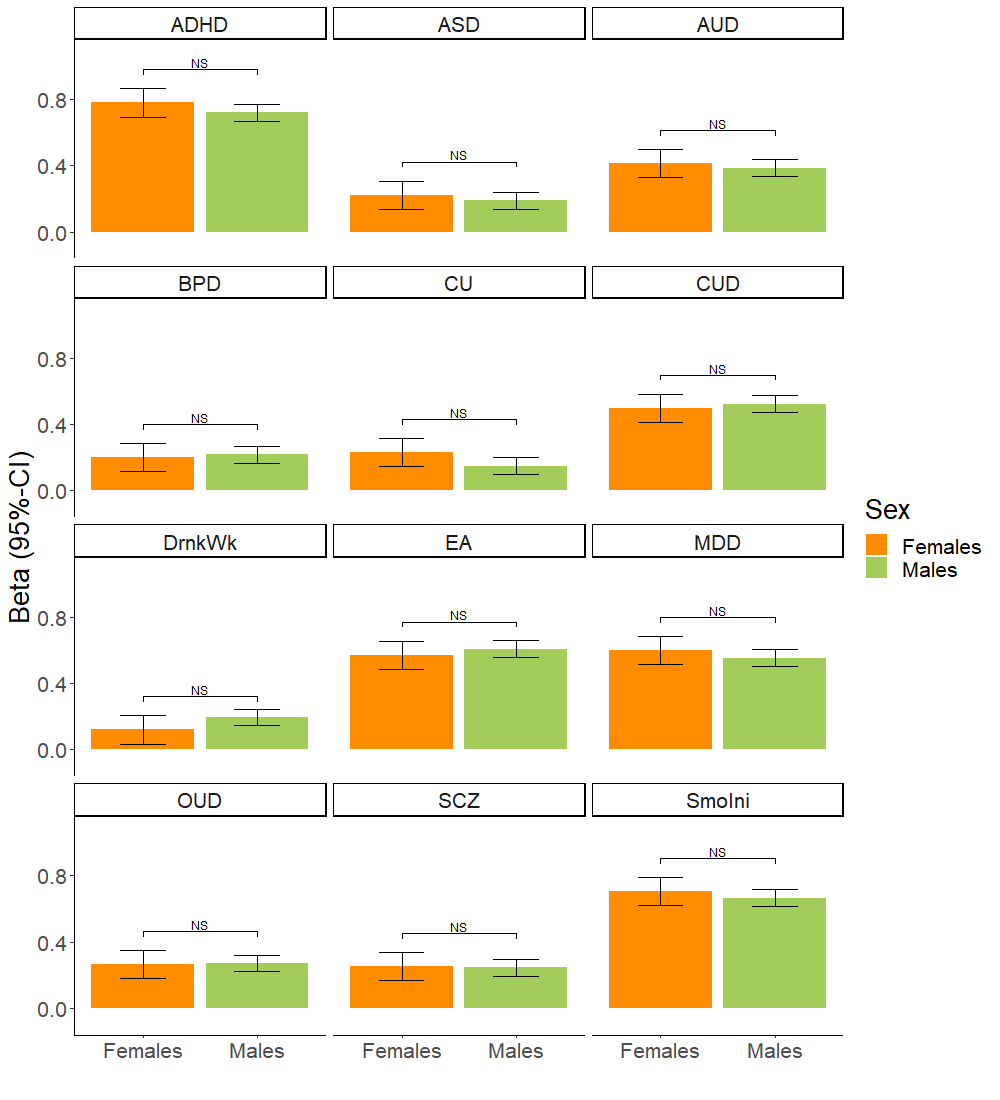

PGS analyses of females with ADHD-CUD (N = 534) vs males with ADHD-CUD (N = 1,545), on the x-axis bench marked against a control group of both males and females (N = 37,246). The slope (beta) of the linear regression (95% CI) is shown on the y-axis. Significant difference (P-value) between beta for females-ADHD-CUD and males-ADHD-CUD is indicated with horizontal line. NS indicate no significant difference in the Wald test of equal group effect (see also Supplementary Table 24).

**Supplementary Figure 9. Absolute risk of CUD among individuals with ADHD**

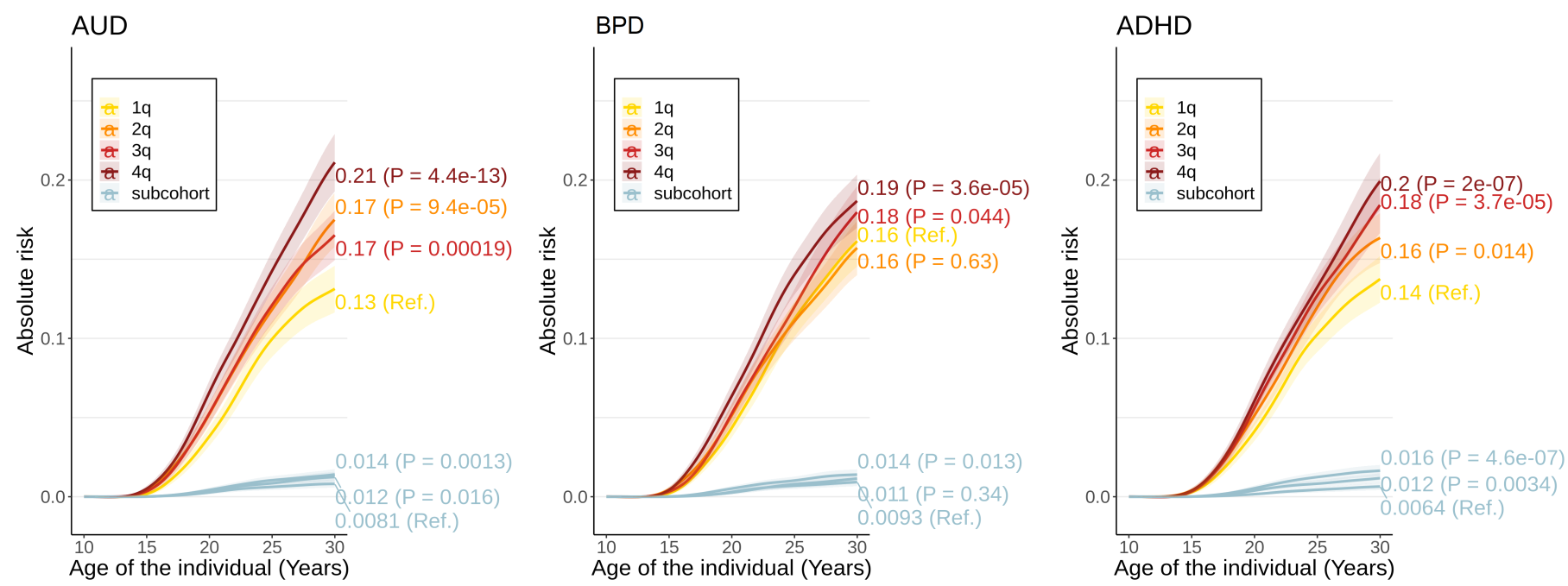

a. **b. c.**

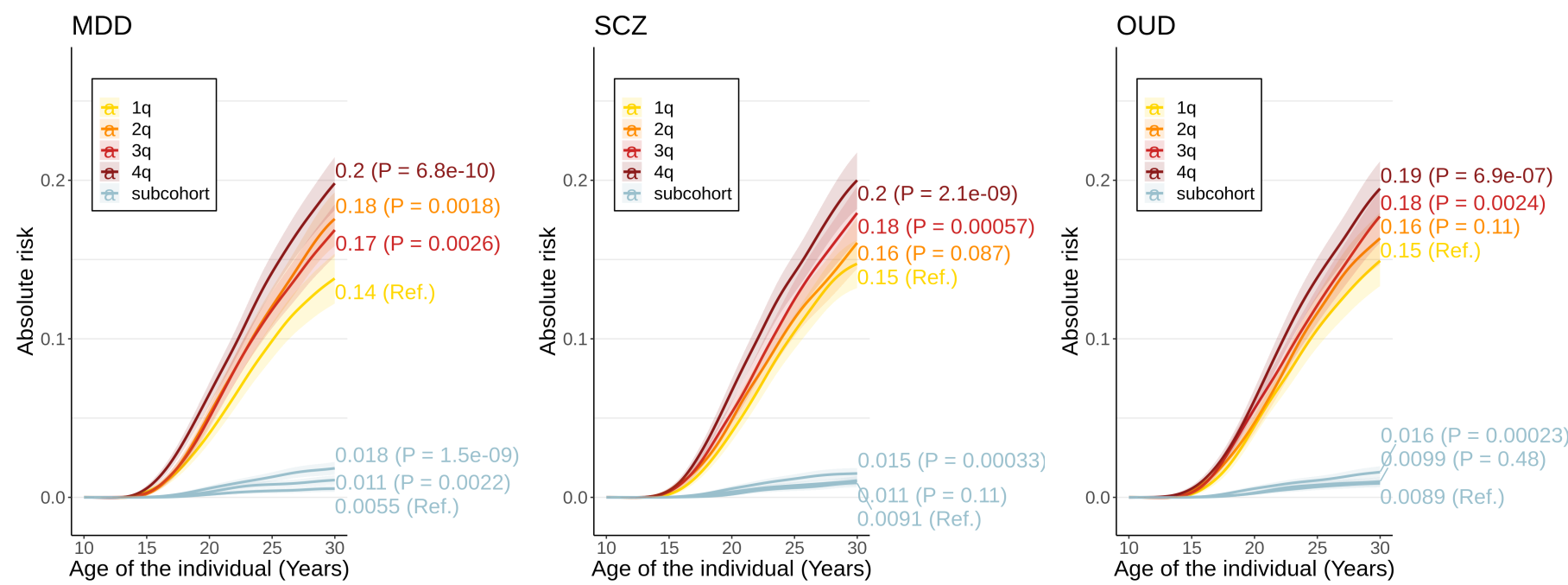

**d. e. f.**

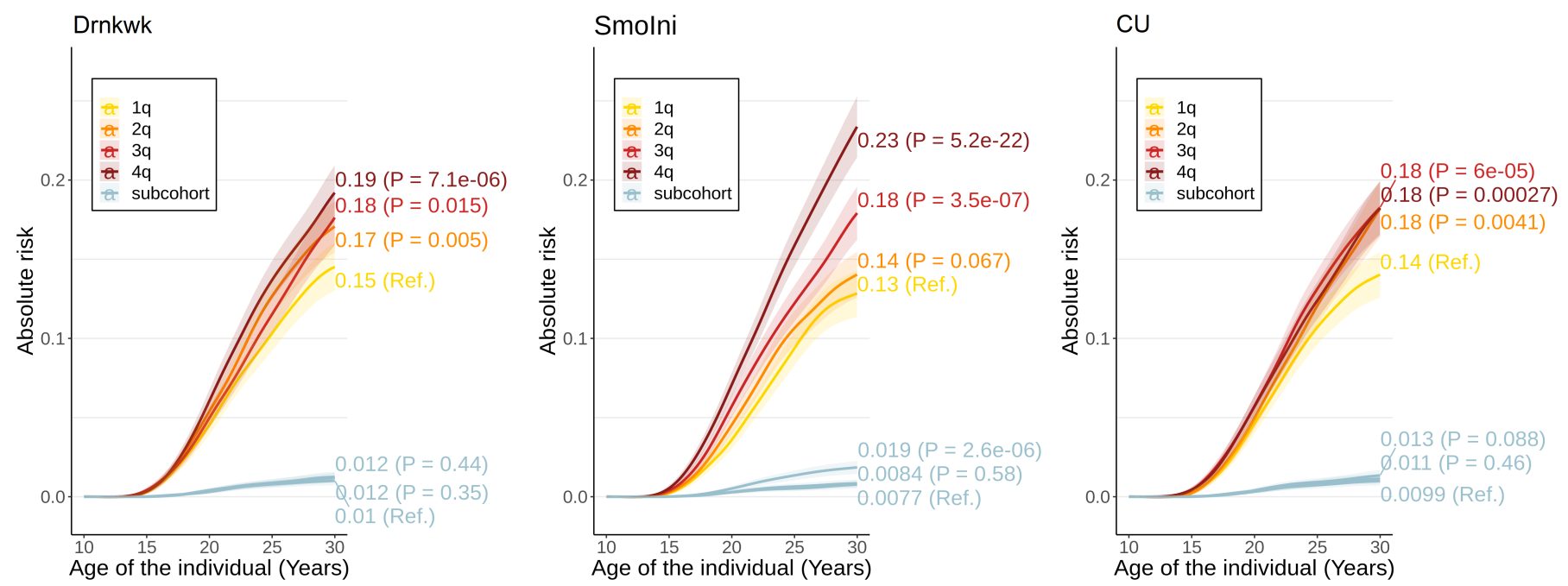

**g. h. i.**

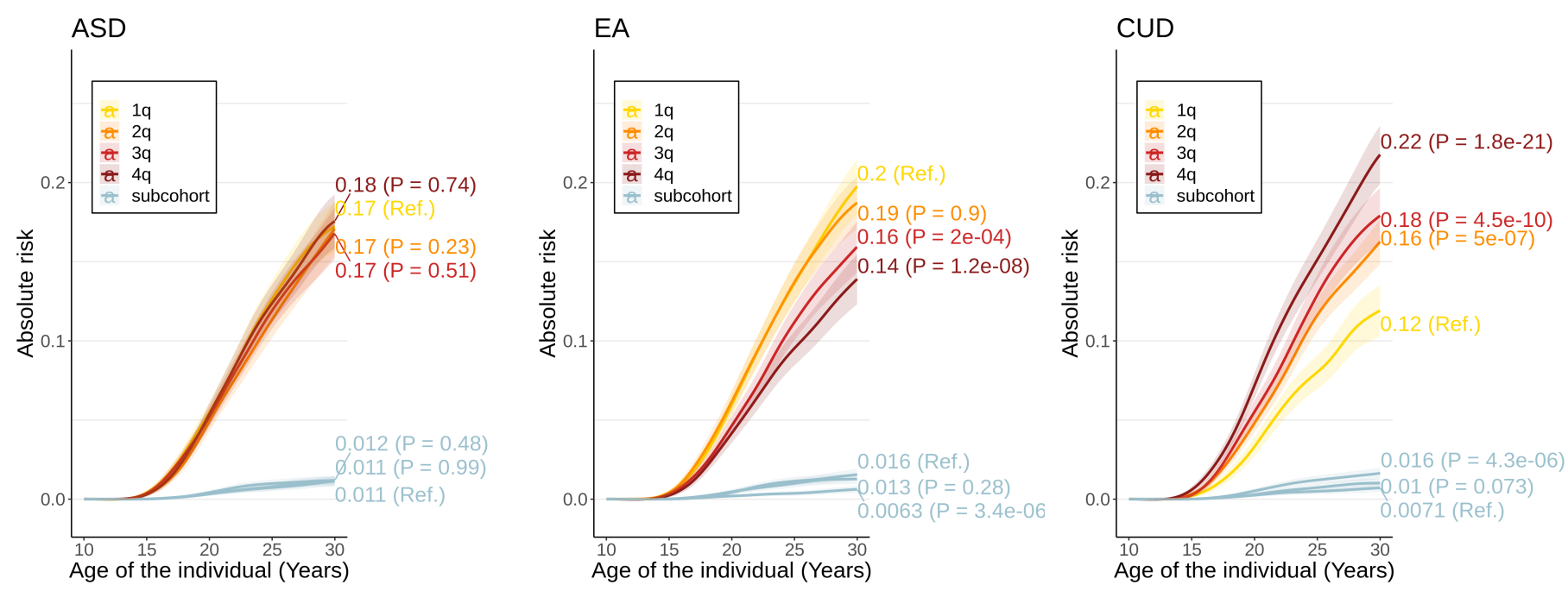

**j. k. l.**

Absolute risk (95% CI) (on the y-axis) over time (on the x-axis) of CUD among individuals with ADHD (N=25545) stratified into quartiles (1q-4q) based on their PGS for **(a)** alcohol use disorder (AUD) **(b)** bipolar disorder (BIP) **(c)** ADHD **(d)** major depressive disorder (MDD) **(e)** schizophrenia (SZ) **(f)** opioid use disorder (OUD) **(g)** drinks per week (Drink) **(h)** smoking initiation (SmokIni) **(i)** CU (CanUse) **(j)** autism spectrum disorder (ASD) **(k)** educational attainment **(l)** CUD. Absolute risk (95% CI) of CUD among individuals in the population-based sub-cohort (without ADHD; N = 37,246)) stratified into tertiles based on their PGS, is shown in light blue.

**Supplementary Figure 10. Absolute risk of CUD among individuals with ADHD who at least one parent diagnosed with a psychiatric disorder**

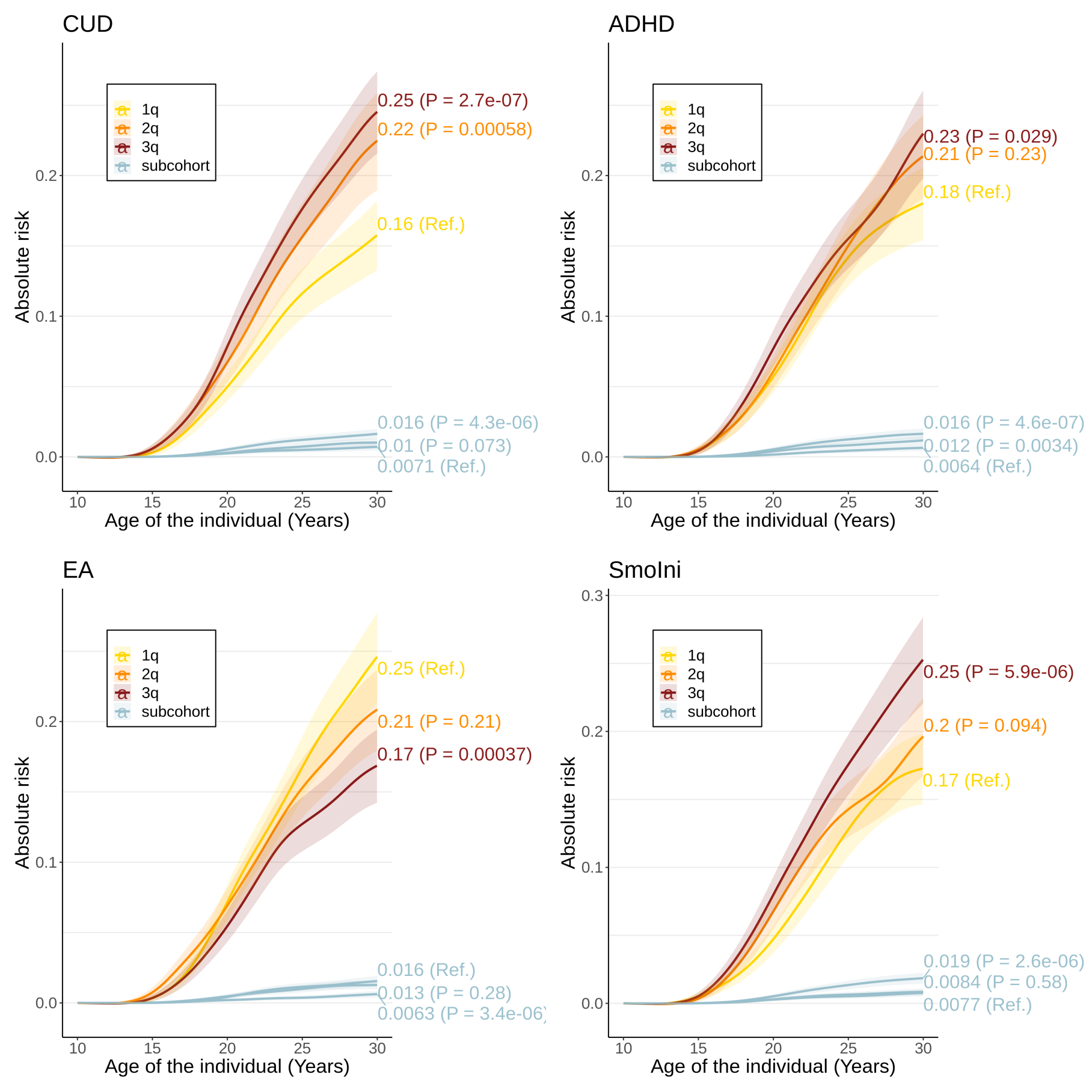

Absolute risk (95% CI) (on the y-axis) over time (on the x-axis) of comorbid CUD (N= 2,079) among individuals with ADHD having at least one parent diagnosed with psychiatric disorder (N=6,999) stratified into tertiles (1q-3q) based on their PGS for: CUD, ADHD, educational attainment (EA), and smoking initiation (SmoIni), PGS-phenotype is given in upper left corner. For comparison the absolute risk (95% CI) of CUD among individuals in the population-based sub-cohort (without ADHD; N = 37,246) stratified into tertiles based on their PGS, is shown in light blue.

**Supplementary Figure 11. Absolute risk of CUD among females with ADHD**

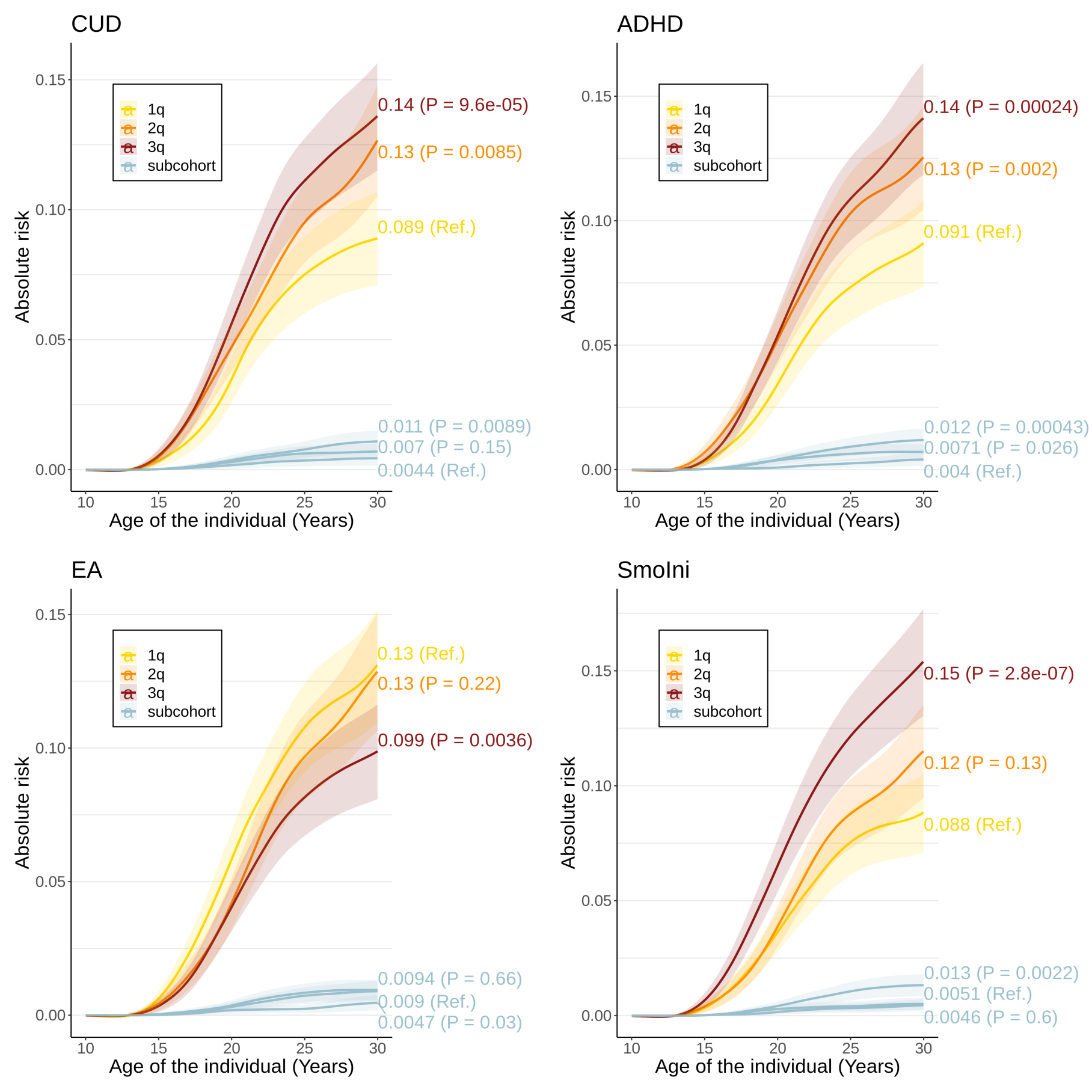

Absolute risk (95% CI) (on the y-axis) over time (on the x-axis) of comorbid CUD (N = 534) among females with ADHD (N = 7,863) stratified into tertiles (1q-3q) based on their PGS for: CUD, ADHD, educational attainment (EA), and smoking initiation (SmoIni), PGS-phenotype is given in upper left corner. For comparison the absolute risk (95% CI) of CUD among females in the population-based sub-cohort (without ADHD, N = 18,607) stratified into tertiles based on their PGS, is shown in light blue.

**Supplementary Figure 12. Absolute risk of CUD among males with ADHD**

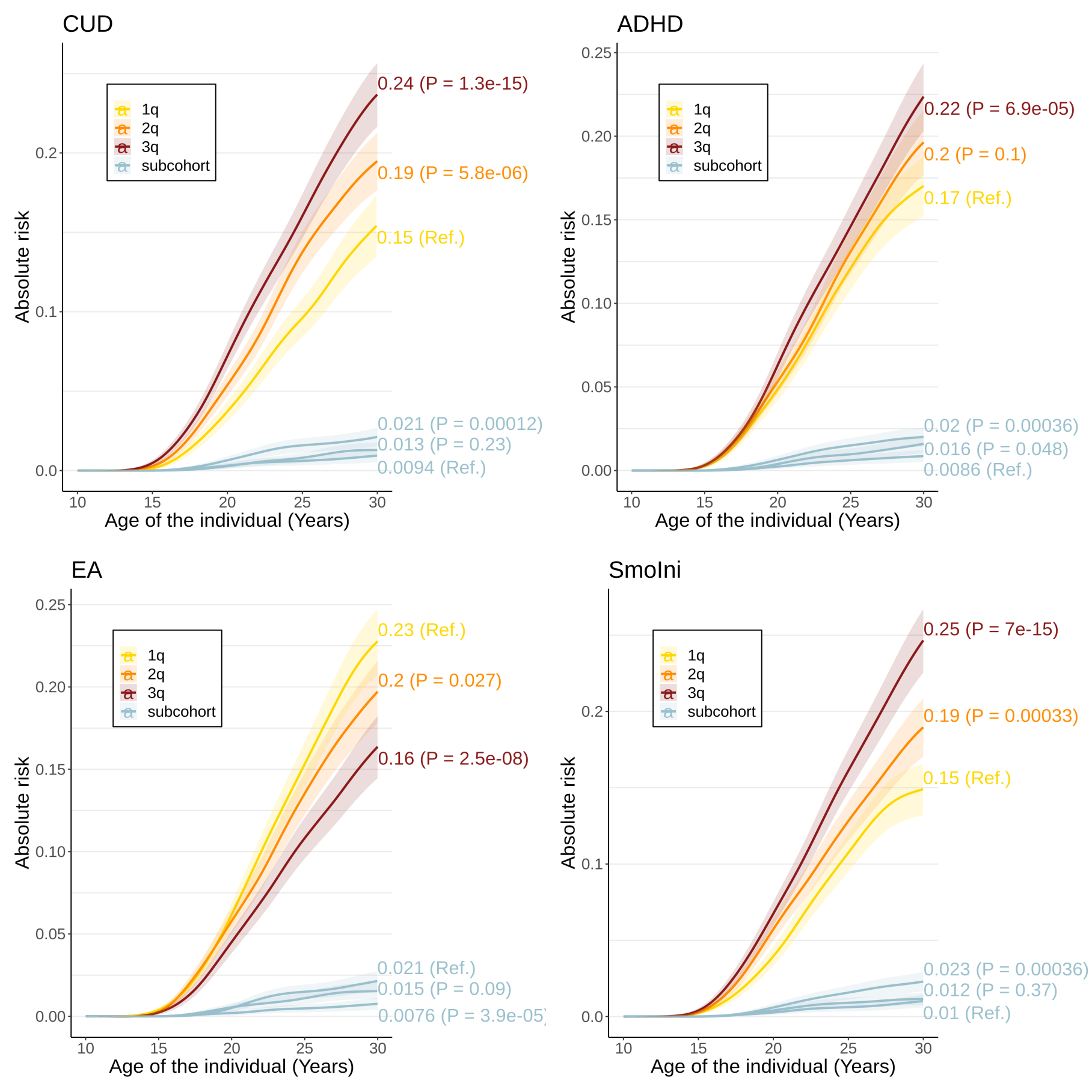

Absolute risk (95% CI) (on the y-axis) over time (on the x-axis) of comorbid CUD (N = 1,545) among males with ADHD (N = 17,682) stratified into tertiles (1q-3q) based on their PGS for: CUD, ADHD, educational attainment, and smoking initiation (SmoIni), PGS-phenotype given in upper left corner. For comparison the absolute risk (95% CI) of CUD among males in the population-based sub-cohort (without ADHD, N = 18,960) stratified into tertiles based on their PGS, is shown in light blue.

**Supplementary Figure 13. Workflow and overview of methods**

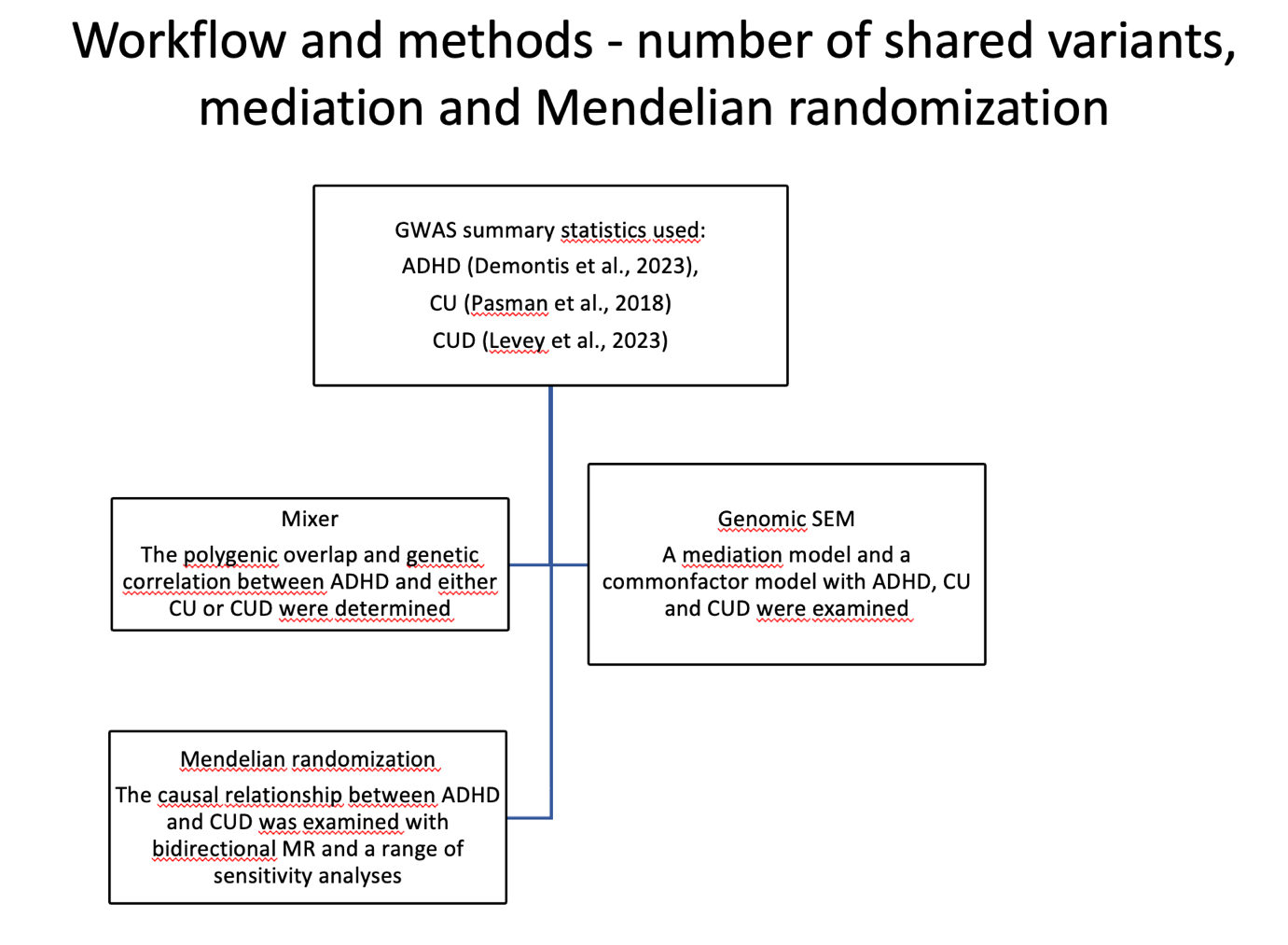

Overview of workflow and methods for analyses of number of shared variants, mediation and Mendelian randomization analyses.

**Supplementary Figure 14. Workflow and overview of methods**

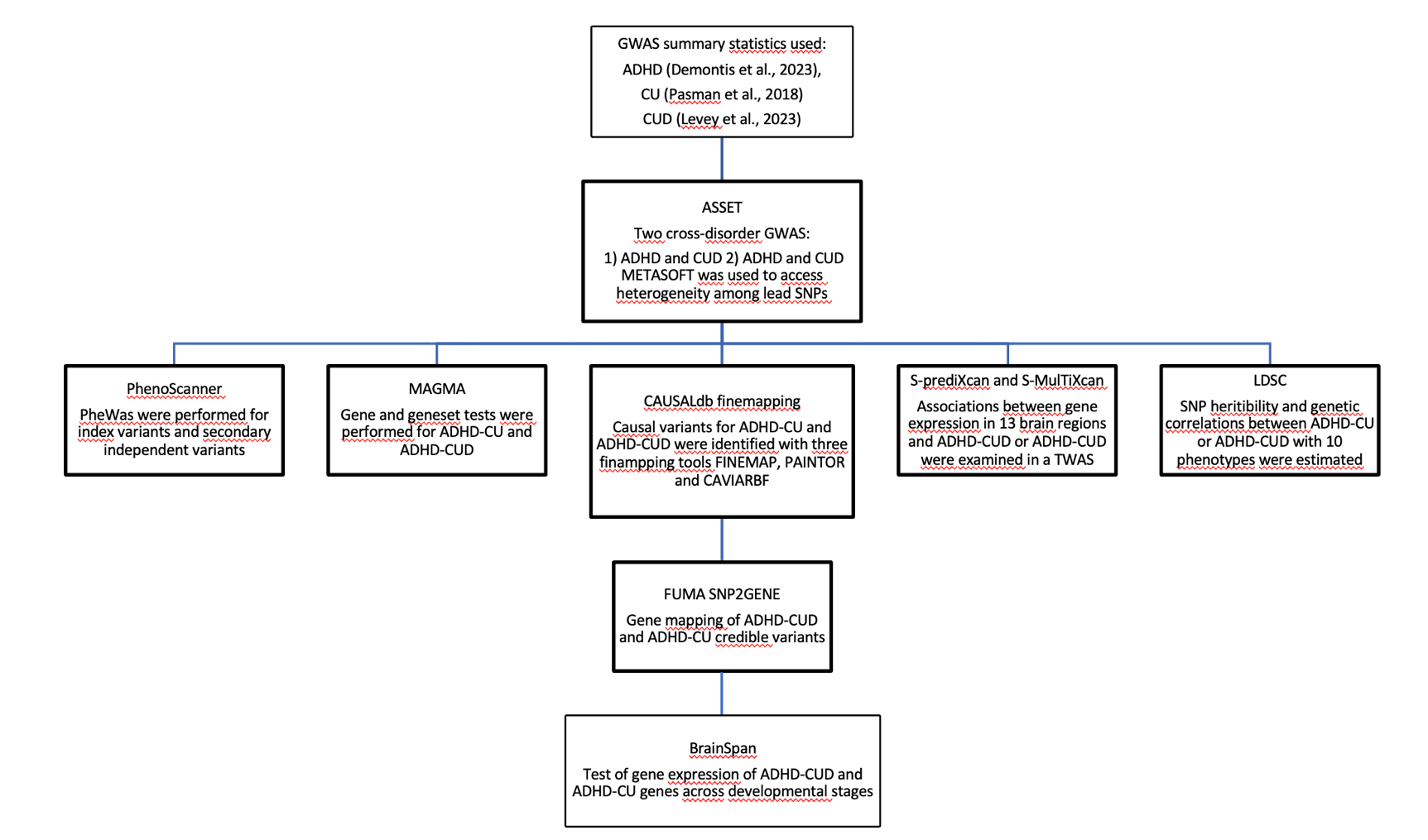

Overview of workflow and methods for cross-disorder GWAS meta-analyses, and secondary GWAS analyses.

**Supplementary Figure 15. Workflow and overview of methods**

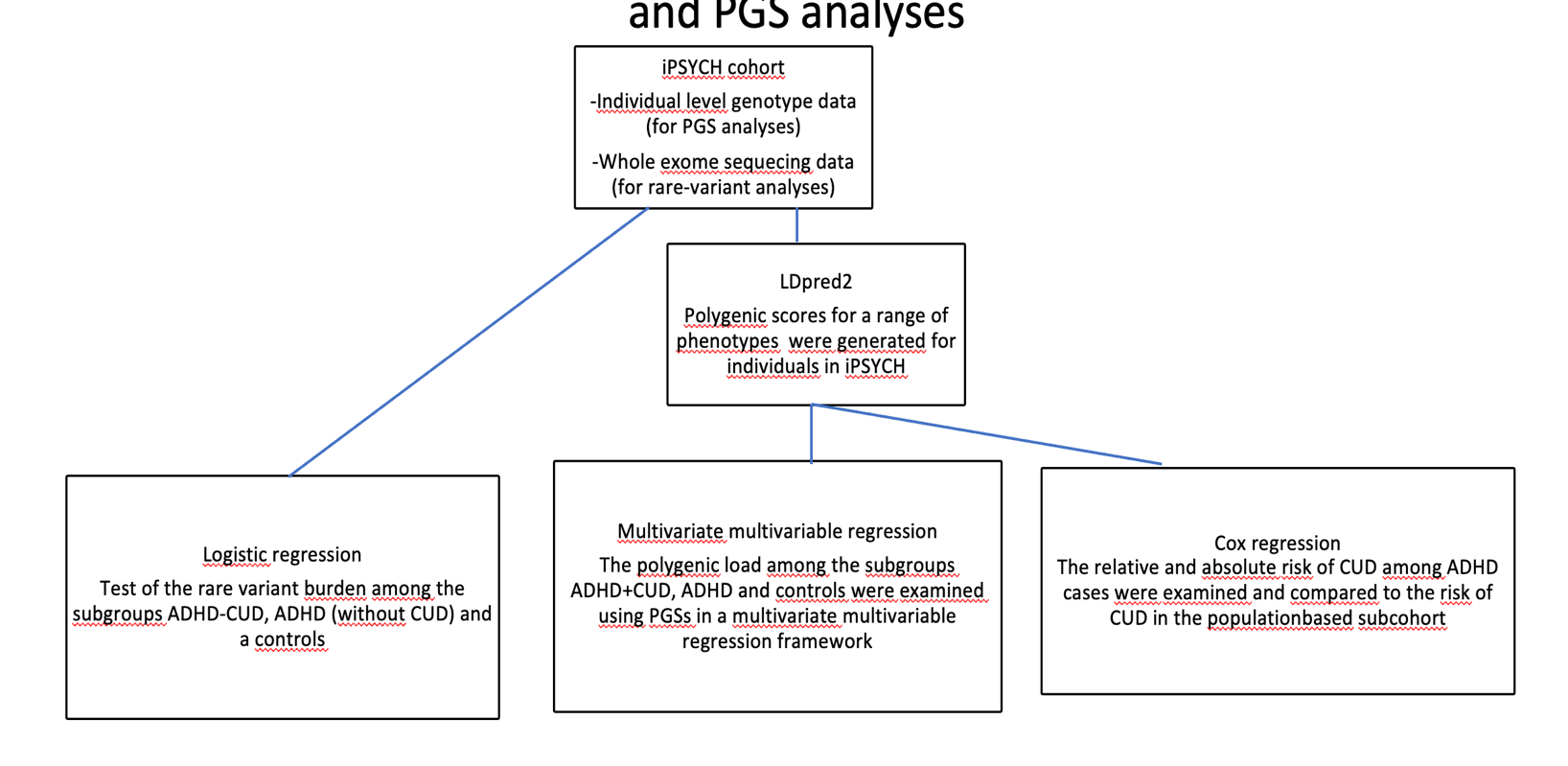

Overview of workflow and methods for rare variant and PGS analyses in the iPSYCH cohort.

**VA Million Veteran Program: Core Acknowledgement, February 2023**

**MVP Program Office**

- Sumitra Muralidhar, Ph.D., Program Director

US Department of Veterans Affairs, 810 Vermont Avenue NW, Washington, DC 20420

- Jennifer Moser, Ph.D., Associate Director, Scientific Programs

US Department of Veterans Affairs, 810 Vermont Avenue NW, Washington, DC 20420

- Jennifer E. Deen, B.S., Associate Director, Cohort & Public Relations

US Department of Veterans Affairs, 810 Vermont Avenue NW, Washington, DC 20420

**MVP Executive Committee**

- Co-Chair: Philip S. Tsao, Ph.D.

VA Palo Alto Health Care System, 3801 Miranda Avenue, Palo Alto, CA 94304

- Co-Chair: Sumitra Muralidhar, Ph.D.

US Department of Veterans Affairs, 810 Vermont Avenue NW, Washington, DC 20420

- J. Michael Gaziano, M.D., M.P.H.

VA Boston Healthcare System, 150 S. Huntington Avenue, Boston, MA 02130

- Elizabeth Hauser, Ph.D.

Durham VA Medical Center, 508 Fulton Street, Durham, NC 27705

- Amy Kilbourne, Ph.D., M.P.H.

VA HSR&D, 2215 Fuller Road, Ann Arbor, MI 48105

- Shiuh-Wen Luoh, M.D., Ph.D.

VA Portland Health Care System, 3710 SW US Veterans Hospital Rd, Portland, OR 97239

- Michael Matheny, M.D., M.S., M.P.H.

VA Tennessee Valley Healthcare System, 1310 24^th^ Ave. South, Nashville, TN 37212

- Dave Oslin, M.D.

Philadelphia VA Medical Center, 3900 Woodland Avenue, Philadelphia, PA 19104

**MVP Co-Principal Investigators**

- J. Michael Gaziano, M.D., M.P.H.

VA Boston Healthcare System, 150 S. Huntington Avenue, Boston, MA 02130

- Philip S. Tsao, Ph.D.

VA Palo Alto Health Care System, 3801 Miranda Avenue, Palo Alto, CA 94304

**MVP Core Operations**

- Lori Churby, B.S., Director, MVP Regulatory Affairs

VA Palo Alto Health Care System, 3801 Miranda Avenue, Palo Alto, CA 94304

- Stacey B. Whitbourne, Ph.D., Director, MVP Cohort Management

VA Boston Healthcare System, 150 S. Huntington Avenue, Boston, MA 02130

- Jessica V. Brewer, M.P.H., Director, MVP Recruitment & Enrollment

VA Boston Healthcare System, 150 S. Huntington Avenue, Boston, MA 02130

- Shahpoor (Alex) Shayan, M.S., Director, MVP Recruitment and Enrollment Informatics

VA Boston Healthcare System, 150 S. Huntington Avenue, Boston, MA 02130

- Luis E. Selva, Ph.D., Executive Director, MVP Biorepositories

VA Boston Healthcare System, 150 S. Huntington Avenue, Boston, MA 02130

- Saiju Pyarajan Ph.D., Director, Data and Computational Sciences

VA Boston Healthcare System, 150 S. Huntington Avenue, Boston, MA 02130

- Kelly Cho, M.P.H, Ph.D., Director, MVP Phenomics Data Core

VA Boston Healthcare System, 150 S. Huntington Avenue, Boston, MA 02130

- Scott L. DuVall, Ph.D., Director, VA Informatics and Computing Infrastructure (VINCI)

VA Salt Lake City Health Care System, 500 Foothill Drive, Salt Lake City, UT 84148

- Mary T. Brophy M.D., M.P.H., Director, VA Central Biorepository

VA Boston Healthcare System, 150 S. Huntington Avenue, Boston, MA 02130

- MVP Coordinating Centers
  - MVP Coordinating Center, Boston - J. Michael Gaziano, M.D., M.P.H.

VA Boston Healthcare System, 150 S. Huntington Avenue, Boston, MA 02130

- MVP Coordinating Center, Palo Alto – Philip S. Tsao, Ph.D.

VA Palo Alto Health Care System, 3801 Miranda Avenue, Palo Alto, CA 94304

- MVP Information Center, Canandaigua – Brady Stephens, M.S.

Canandaigua VA Medical Center, 400 Fort Hill Avenue, Canandaigua, NY 14424

- Cooperative Studies Program Clinical Research Pharmacy Coordinating Center, Albuquerque – Todd Connor, Pharm.D.; Dean P. Argyres, B.S., M.S.

New Mexico VA Health Care System, 1501 San Pedro Drive SE, Albuquerque, NM 87108

**MVP Publications and Presentations Committee**

- Co-Chair: Themistocles L. Assimes, M.D., Ph. D

VA Palo Alto Health Care System, 3801 Miranda Avenue, Palo Alto, CA 94304

- Co-Chair: Adriana Hung, M.D.; M.P.H

VA Tennessee Valley Healthcare System, 1310 24^th^ Ave. South, Nashville, TN 37212

- Co-Chair: Henry Kranzler, M.D.

Philadelphia VA Medical Center, 3900 Woodland Avenue, Philadelphia, PA 19104

**MVP Local Site Investigators**

- Samuel Aguayo, M.D., Phoenix VA Health Care System

650 E. Indian School Road, Phoenix, AZ 85012

- Sunil Ahuja, M.D., South Texas Veterans Health Care System

7400 Merton Minter Boulevard, San Antonio, TX 78229

- Kathrina Alexander, M.D., Veterans Health Care System of the Ozarks

1100 North College Avenue, Fayetteville, AR 72703

- Xiao M. Androulakis, M.D., Columbia VA Health Care System

6439 Garners Ferry Road, Columbia, SC 29209

- Prakash Balasubramanian, M.D., William S. Middleton Memorial Veterans Hospital

2500 Overlook Terrace, Madison, WI 53705

- Zuhair Ballas, M.D., Iowa City VA Health Care System

601 Highway 6 West, Iowa City, IA 52246-2208

- Jean Beckham, Ph.D., Durham VA Medical Center

508 Fulton Street, Durham, NC 27705

- Sujata Bhushan, M.D., VA North Texas Health Care System

4500 S. Lancaster Road, Dallas, TX 75216

- Edward Boyko, M.D., VA Puget Sound Health Care System

1660 S. Columbian Way, Seattle, WA 98108-1597

- David Cohen, M.D., Portland VA Medical Center

3710 SW U.S. Veterans Hospital Road, Portland, OR 97239

- Louis Dellitalia, M.D., Birmingham VA Medical Center

700 S. 19th Street, Birmingham AL 35233

- L. Christine Faulk, M.D., Robert J. Dole VA Medical Center

5500 East Kellogg Drive, Wichita, KS 67218-1607

- Joseph Fayad, M.D., VA Southern Nevada Healthcare System

6900 North Pecos Road, North Las Vegas, NV 89086

- Daryl Fujii, Ph.D., VA Pacific Islands Health Care System

459 Patterson Rd, Honolulu, HI 96819

- Saib Gappy, M.D., John D. Dingell VA Medical Center

4646 John R Street, Detroit, MI 48201

- Frank Gesek, Ph.D., White River Junction VA Medical Center

163 Veterans Drive, White River Junction, VT 05009

- Jennifer Greco, M.D., Sioux Falls VA Health Care System

2501 W 22nd Street, Sioux Falls, SD 57105

- Michael Godschalk, M.D., Richmond VA Medical Center

1201 Broad Rock Blvd., Richmond, VA 23249

- Todd W. Gress, M.D., Ph.D., Hershel “Woody” Williams VA Medical Center

1540 Spring Valley Drive, Huntington, WV 25704

- Samir Gupta, M.D., M.S.C.S., VA San Diego Healthcare System

3350 La Jolla Village Drive, San Diego, CA 92161

- Salvador Gutierrez, M.D., Edward Hines, Jr. VA Medical Center

5000 South 5th Avenue, Hines, IL 60141

- John Harley, M.D., Ph.D., Cincinnati VA Medical Center

3200 Vine Street, Cincinnati, OH 45220

- Kimberly Hammer, Ph.D., Fargo VA Health Care System

2101 N. Elm, Fargo, ND 58102

- Mark Hamner, M.D., Ralph H. Johnson VA Medical Center

109 Bee Street, Mental Health Research, Charleston, SC 29401

- Adriana Hung, M.D., M.P.H., VA Tennessee Valley Healthcare System

1310 24th Avenue, South Nashville, TN 37212

- Robin Hurley, M.D., W.G. (Bill) Hefner VA Medical Center

1601 Brenner Ave, Salisbury, NC 28144

- Pran Iruvanti, D.O., Ph.D., Hampton VA Medical Center

100 Emancipation Drive, Hampton, VA 23667

- Frank Jacono, M.D., VA Northeast Ohio Healthcare System

10701 East Boulevard, Cleveland, OH 44106

- Darshana Jhala, M.D., Philadelphia VA Medical Center

3900 Woodland Avenue, Philadelphia, PA 19104

- Scott Kinlay, M.B.B.S., Ph.D., VA Boston Healthcare System

150 S. Huntington Avenue, Boston, MA 02130

- Jon Klein, M.D., Ph.D., Louisville VA Medical Center

800 Zorn Avenue, Louisville, KY 40206

- Michael Landry, Ph.D., Southeast Louisiana Veterans Health Care System

2400 Canal Street, New Orleans, LA 70119

- Peter Liang, M.D., M.P.H., VA New York Harbor Healthcare System

423 East 23rd Street, New York, NY 10010

- Suthat Liangpunsakul, M.D., M.P.H., Richard Roudebush VA Medical Center

1481 West 10th Street, Indianapolis, IN 46202

- Jack Lichy, M.D., Ph.D., Washington DC VA Medical Center

50 Irving St, Washington, D. C. 20422

- C. Scott Mahan, M.D., Charles George VA Medical Center

1100 Tunnel Road, Asheville, NC 28805

- Ronnie Marrache, M.D., VA Maine Healthcare System

1 VA Center, Augusta, ME 04330

- Stephen Mastorides, M.D., James A. Haley Veterans’ Hospital

13000 Bruce B. Downs Blvd, Tampa, FL 33612

- Elisabeth Mates M.D., Ph.D., VA Sierra Nevada Health Care System

975 Kirman Avenue, Reno, NV 89502

- Kristin Mattocks, Ph.D., M.P.H., Central Western Massachusetts Healthcare System

421 North Main Street, Leeds, MA 01053

- Paul Meyer, M.D., Ph.D., Southern Arizona VA Health Care System

3601 S 6th Avenue, Tucson, AZ 85723

- Jonathan Moorman, M.D., Ph.D., James H. Quillen VA Medical Center

Corner of Lamont & Veterans Way, Mountain Home, TN 37684

- Timothy Morgan, M.D., VA Long Beach Healthcare System

5901 East 7th Street Long Beach, CA 90822

- Maureen Murdoch, M.D., M.P.H., Minneapolis VA Health Care System

One Veterans Drive, Minneapolis, MN 55417

- James Norton, Ph.D., VA Health Care Upstate New York

113 Holland Avenue, Albany, NY 12208

- Olaoluwa Okusaga, M.D., Michael E. DeBakey VA Medical Center

2002 Holcombe Blvd, Houston, TX 77030

- Kris Ann Oursler, M.D., Salem VA Medical Center

1970 Roanoke Blvd, Salem, VA 24153

- Ana Palacio, M.D., M.P.H., Miami VA Health Care System

1201 NW 16th Street, 11 GRC, Miami FL 33125

- Samuel Poon, M.D., Manchester VA Medical Center

718 Smyth Road, Manchester, NH 03104

- Emily Potter, Pharm.D., VA Eastern Kansas Health Care System

4101 S 4th Street Trafficway, Leavenworth, KS 66048

- Michael Rauchman, M.D., St. Louis VA Health Care System

915 North Grand Blvd, St. Louis, MO 63106

- Richard Servatius, Ph.D., Syracuse VA Medical Center

800 Irving Avenue, Syracuse, NY 13210

- Satish Sharma, M.D., Providence VA Medical Center

830 Chalkstone Avenue, Providence, RI 02908

- River Smith, Ph.D., Eastern Oklahoma VA Health Care System

1011 Honor Heights Drive, Muskogee, OK 74401

- Peruvemba Sriram, M.D., N. FL/S. GA Veterans Health System

1601 SW Archer Road, Gainesville, FL 32608

- Patrick Strollo, Jr., M.D., VA Pittsburgh Health Care System

University Drive, Pittsburgh, PA 15240

- Neeraj Tandon, M.D., Overton Brooks VA Medical Center

510 East Stoner Ave, Shreveport, LA 71101

- Philip Tsao, Ph.D., VA Palo Alto Health Care System

3801 Miranda Avenue, Palo Alto, CA 94304-1290

- Gerardo Villareal, M.D., New Mexico VA Health Care System

1501 San Pedro Drive, S.E. Albuquerque, NM 87108

- Agnes Wallbom, M.D., M.S., VA Greater Los Angeles Health Care System

11301 Wilshire Blvd, Los Angeles, CA 90073

- Jessica Walsh, M.D., VA Salt Lake City Health Care System

500 Foothill Drive, Salt Lake City, UT 84148

- John Wells, Ph.D., Edith Nourse Rogers Memorial Veterans Hospital

200 Springs Road, Bedford, MA 01730

- Jeffrey Whittle, M.D., M.P.H., Clement J. Zablocki VA Medical Center

5000 West National Avenue, Milwaukee, WI 53295

- Mary Whooley, M.D., San Francisco VA Health Care System

4150 Clement Street, San Francisco, CA 94121

- Allison E. Williams, N.D., Ph.D., R.N, Bay Pines VA Healthcare System

10,000 Bay Pines Blvd Bay Pines, FL 33744

- Peter Wilson, M.D., Atlanta VA Medical Center

1670 Clairmont Road, Decatur, GA 30033

- Junzhe Xu, M.D., VA Western New York Healthcare System

3495 Bailey Avenue, Buffalo, NY 14215-1199

- Shing Shing Yeh, Ph.D., M.D., Northport VA Medical Center

79 Middleville Road, Northport, NY 11768
